# Ticking Differently: Elucidating Sexual Dimorphism in Human Aging through Metabolomics, Proteomics and Genomics

**DOI:** 10.1101/2025.10.30.25339071

**Authors:** Sihao Xiao, Pallavi Kaushik, Gaoyu Du, Bowen Liu, Salman B. Hosawi, Benoit Hastoy, M. Austin Argentieri, Laura M. Winchester, Alejo J. Nevado-Holgado, Rima Kaddurah-Daouk, Najaf Amin, Cornelia van Duijn

## Abstract

That males and females age differently has been overlooked while developing aging clocks. Here, we developed a sex-specific metabolic aging clock in 390,941 individuals from the UK Biobank and integrated it with genetic, proteomic and epidemiological data to identify mechanisms accelerating/decelerating metabolic aging in males and females. Our findings reveal dysregulation of cholesterol metabolism, immune system, hemostasis, and cell growth, survival and apoptosis as common mechanisms accelerating metabolic aging in males and females, and upregulation of oxidative stress detoxification, cellular resilience and tissue integrity as common mechanisms decelerating metabolic aging. In females, a further dysregulation of carbohydrate/glucose metabolism, circadian rhythm and hormone metabolism accelerating metabolic aging is observed, while dysregulation of energy metabolism, cancer and longevity pathway is specifically observed in males. Among reproductive factors, late puberty and higher parity manifest as protective factors, decelerating metabolic aging in both sexes, and additionally childbirth at older age decelerating metabolic aging in women. Accelerated metabolic age strongly predicted morbidity and mortality in both sexes, except that the magnitude of association was several folds higher in males, and obesity explained most of the disease associations in females, suggesting that obesity influences metabolic aging and subsequent health outcomes differently in males and females. Consistent with the upregulation of molecular mechanisms involved in cancer in males, accelerated metabolic aging predicted several common cancers in males but not females.

Our study provides novel insights into the biological mechanisms underlying aging and disease susceptibility in males and females, underscoring the importance of considering sex differences in healthcare strategies and public health policies.

## Main Text

Females outlive males globally^1^ although females are frailer^2^ and have poorer physical functioning compared to males^3^. Both historical and recent data suggest that females live longer even during severe famines and epidemics^4-7^. These differences have been attributed to various factors, including hormonal influences such as estrogen^8,9^, genetics^10^, telomere length^11-13^, metabolic rate^10,14^, adiposity distribution^15,16^, and behavioral differences^17^ where males tend to engage in risky behaviors^17^ and often have poorer lifestyle habits, such as higher alcohol consumption, excessive smoking and poorer oral hygiene^18-20^. Such environmental insults are captured by epigenetic processes like DNA methylation, which shows a higher entropy in males with advancing age^10,21,22^.

Biological age predictors, often referred to as “aging clocks”, have been developed to estimate biological aging across different omics layers, demonstrating their substantial potential in aging research^23-31^. Most of these clocks, however, did not investigate sex-specific variations underlying aging and the association of biological age with disease and mortality outcome in each sex. A recent study of Reicher L et al.^32^ explored the differences in aging between sexes but in a relatively small sample of 10,000 individuals using environmental exposures, physiological parameters, and a narrow platform of molecular markers mainly concentrating on lipids. Further, all the previous studies focused on predicting chronological age and studying the impact on morbidity and mortality rather than understanding the mechanisms underlying accelerated/decelerated aging.

In this study, we develop metabolites-based sex-specific aging clocks leveraging the largest ever available data from 488,318 individuals in the UK Biobank with a focus on understanding common and sex-specific mechanisms underlying accelerated/decelerated metabolic aging. We first constructed a sex-specific metabolic age acceleration (metAgeGap) metric, which estimates the variation in individuals’ rate of metabolic aging compared to others with the same chronological age group. Next, we studied its association with sex-specific reproductive factors, future risks of age-related diseases and mortality, and predicted survival. Finally, to explore the common and divergent molecular regulators of metAgeGap in males and females, we performed a genome-wide and a proteome-wide association analysis of the sex-specific metAgeGap.

## Results

### Descriptive Statistics

The study population comprised 488,318 participants, including 223,396 males and 264,922 females. Participants taking lipids lowering drugs were excluded from the study, resulting in 168,460 males (mean age=55.86, sd=8.23) and 222,481 females (mean age=56.01, sd=8.00). Over the follow-up period, which averaged 12.1 years for males and 12.4 years for females, there were 2,343 recorded deaths in males and 1,602 in females (**Figure 1A)**.

**Figure 1:**
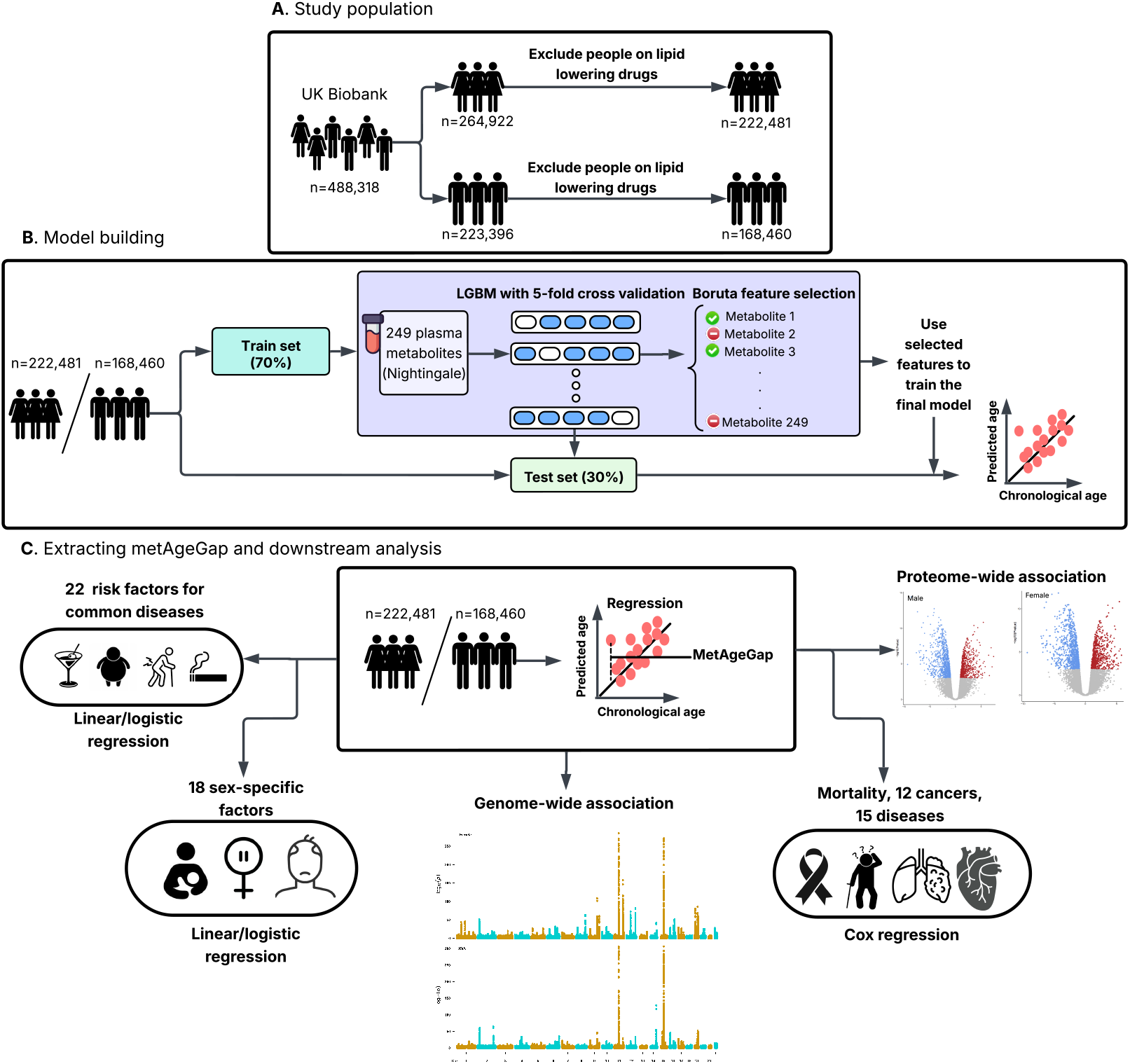
Overview of the study. **(A)** An overview of the study population - criteria used to select participants for the study. **(B)** A metabolome-based age clock was built for each sex using gradient boosting. The classification model was trained on 70% randomly selected males/females and tested on remaining data. Boruta feature selection algorithm was then used to select only relevant features for downstream analysis. **(C)** Extracting metAgeGap metric and using it for downstream analysis.

Most cancers, except lung cancer and non-Hodgkin lymphoma, were more prevalent in males compared to females. Females had a higher prevalence of most common diseases except cardiovascular diseases (ischemic heart disease, ischemic stroke, all stroke) and dementia, including vascular dementia **(Supplementary Table 1)**. Alcohol consumption and smoking were more prevalent among males. Females reported sleep difficulties and tiredness more often **(Supplementary Table 2)**.

### Metabolomic aging clock for males and females

The study design is illustrated in Figure 1. We used NMR-metabolomics data from the UK Biobank in a gradient boosting machine learning model to predict chronological age in males and females separately. The dataset was randomly split into 70% training and 30% test sets for each sex. The predictive performance of the model after five-fold cross validation (20% of the training set) was slightly better in females (*R*^2^ = 0.37) compared to males (*R*^2^ = 0.29) and showed a similar accuracy across testing set (females: Pearson r=0.60, *R*^2^ = 0.36, males: Pearson r=0.53, *R*^2^ = 0.29) **(Supplementary Figure 1A)**.

To quantify individual’s rate of age acceleration or deceleration relative to the population, we calculated metAgeGap as the difference between the predicted age of the individual and the mean predicted age of the corresponding age group. The distribution of the metAgeGap metric was similar across sexes **(Supplementary Figure 1B)**. The Boruta algorithm selected 93 metabolites in females and 76 in males **(Supplementary Figure 1C, Supplementary Figure 2)**. Among these, 61 were common to both sexes, while 32 and 15 metabolites uniquely predicted age in females and males, respectively. These include cholines, and phosphatidylcholines that were uniquely selected in females, and HDL size and triglyceride percentages in large HDL that uniquely predicted age in males **(Supplementary Figure 2)**. Relative importance of the top 20 features as assessed with Shapley Additive exPlanations (SHAP) shows that features contributed differently across sexes - for instance, glutamine (Gln) ranked higher in females (**Supplementary Figures 3 & 4**).

### Correlation of disease-related risk factors with metAgeGap

Next, we tested the association of metAgeGap with 22 common disease-related risk factors (**Figure 2**). Obesity (females: beta=0.94, p=2.25^*^10^-308^, males: beta=0.30, p=1.58^*^10^-40^), BMI (females: beta=0.54, p=2.25^*^10^-308^, males: beta=0.12, p=1.16^*^10^-37^), and blood pressure (systolic blood pressure - females: beta=0.26, p=1.72^*^10^-196^, males: beta=0.10, p=1.08^*^10^-24^; diastolic blood pressure - females: beta=0.42, p=2.25^*^10^-308^, males: beta=0.07, p=9.91^*^10^-12^) showed a stronger association with accelerated metabolic aging in females, whereas type-II diabetes (females: beta=0.31, p=2.05^*^10^-03^, males: beta=1.05, p=3.02^*^10^-35^) showed stronger association in males. Interestingly, smoking (ever smoked and pack years smoked) had a larger impact on metabolic aging in females compared to males (**Figure 2**).

**Figure 2:**
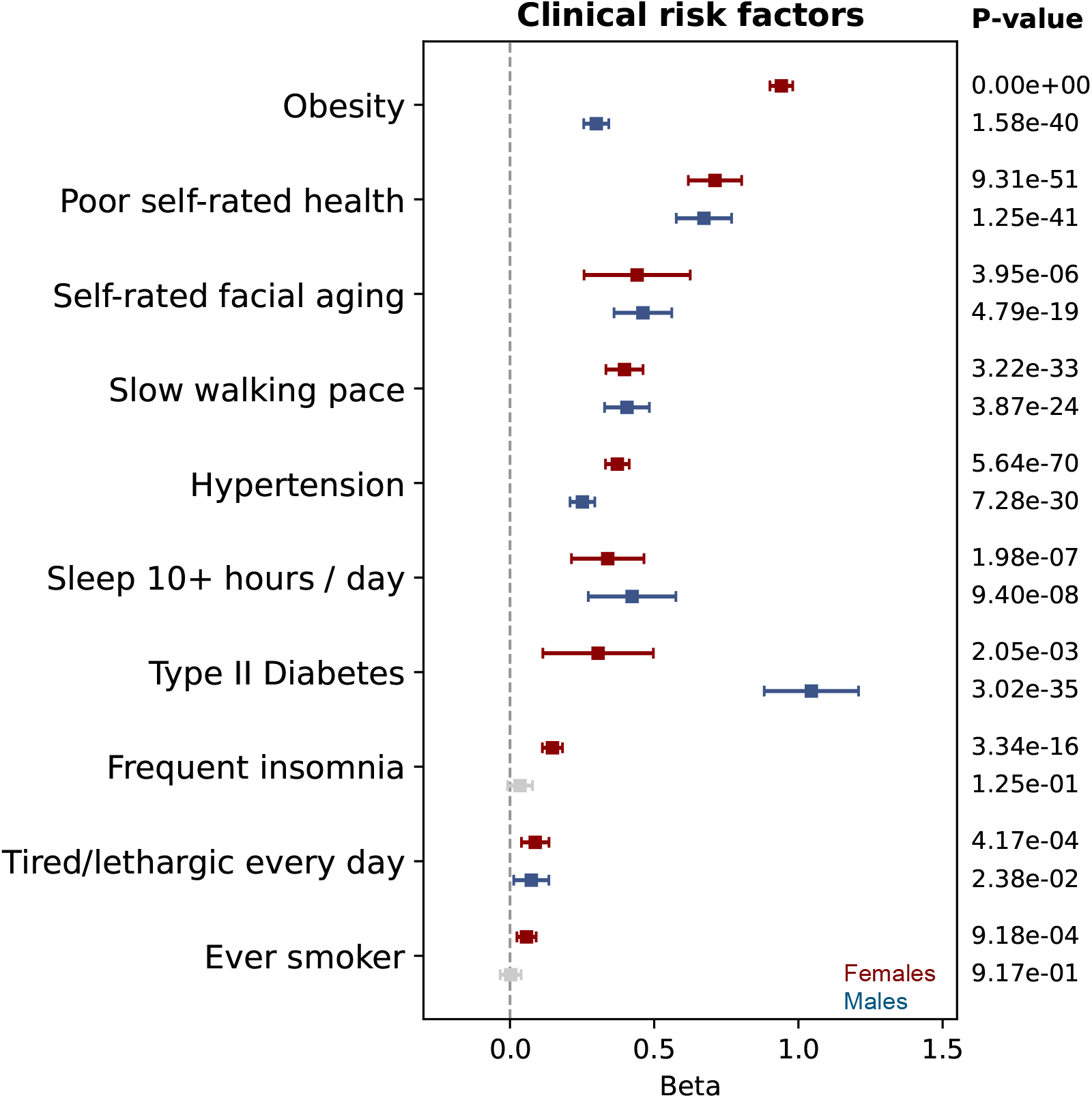
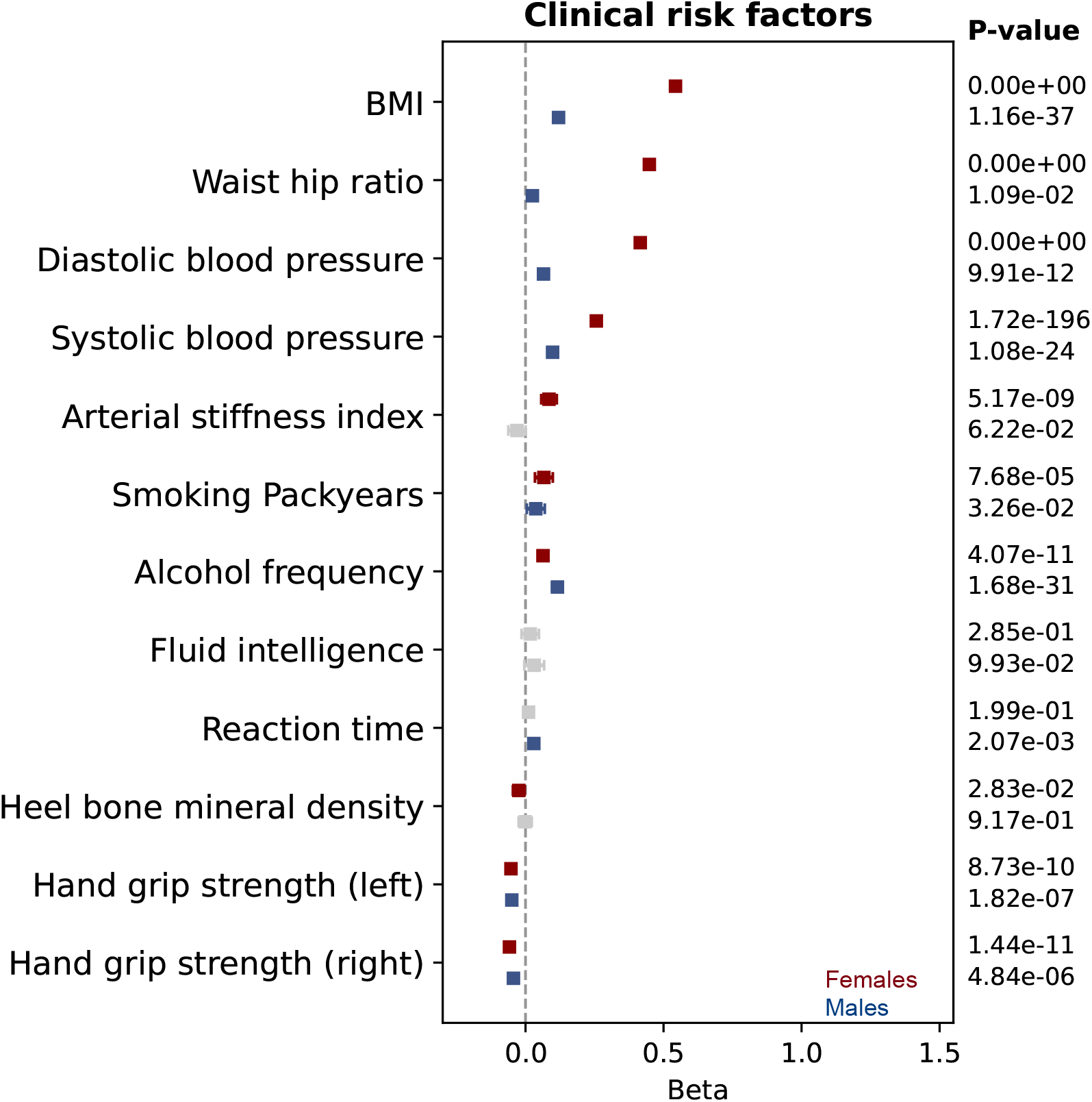
(A) Sex-specific association of age-related clinical risk factors (binary variables) with metAgeGap. (B) Sex-specific association of age-related clinical risk factors (continuous variables) with metAgeGap. Association in females is depicted in red and males in blue. Grey color showed if the association was not significant after FDR correction. The plots display per standard deviation change for easy comparison.

### Late puberty and higher parity are associated with younger metabolic age

Among reproductive factors, late puberty (beta=-0.19, p=3.77^*^10^-110^), the number of live births (beta=-0.16, p=1.39^*^10^-82^), higher age at the first (beta=-0.13, p=1.63^*^10^-32^) and last live births (beta=-0.18, p=1.81^*^10^-64^) and current use of estrogens/progestogens (beta=-1.36, p=7.42^*^10^-76^) were associated with younger metabolic age in females. Age at menopause was not associated with metabolic aging (**Supplementary Figure 5**). Similarly, factors related to puberty in males like higher age at first facial hair appearance (beta=-0.22, p=5.34^*^10^-16^) and voice break (beta= -0.23, p=2.99^*^10^-09^), and number of children (beta=-0.10, p=3.04^*^10^-25^) were associated with younger metabolic age (**Supplementary Figure 5**). Further balding was associated with older metabolic age in males (beta=0.05, p=3.05^*^10^-02^).

### Genome-wide association analysis (GWAS) of metAgeGap

To gain genetic insights into the etiology of metabolic age acceleration, we conducted a GWAS of metAgeGap (N: females=219,115, males=165,933). In females, we identified 23,756 genome-wide significant (p-value<5^*^10^-08^) polymorphisms associated with metAgeGap mapped to 147 independent genomic loci harboring 615 genes (**Supplementary Table 3**). In males, we identified 15,110 significantly associated polymorphisms with metAgeGap mapped to 120 independent genomic loci harboring 510 genes (**Supplementary Table 4**). The corresponding Manhattan and QQ plots and associations are provided (**Supplementary Figures 6 & 7**). 217 genes, including APOE, which has been consistently associated with longevity in GWAS^33^, overlapped between males and females (**Supplementary Figure 8**). A substantial number of loci appeared to be sex specific. For instance, in females we identified several genome-wide significant loci that were not associated in males, e.g., the locus on chromosome 1p13 including gene CELSR2; several independent loci on chromosome 2 harboring genes ABCG5, ABCG8, LCT, MCM6, GLS, USP37 and STK25; several loci on chromosome 3 harboring PPARG, DNAJC13, SLC2A2 and AHSG; locus 5q33 harboring virus receptors HAVCR1 & HAVCR2; locus 6p21-22 harboring CDKAL2 and VEGFA; loci on chromosome 7p harboring DNAH11 and TMED4 and 7q33 harboring STRA8; several independent loci on chromosome 8 harboring genes MFHAS1, XKR6, SLC7A2, NAT2, TRIB1, PTK2 and VPS28; locus 9q34 harboring SURF4 and SLC2A6; locus 10p15 harboring AKR1CL1 and 10q25 harboring TCF7L2 gene; locus 11q24 harboring ST3GAL4; several loci on chromosome 12 harboring genes TULP3, PHC1, SLC38A4 and TIMELESS; loci on chromosome 13 harboring N4BP2L2 and GAS6; chromosome 14q harboring MAP4K5 and ELMSAN1; loci on chromosome 17 harboring GLTPD2 and SLC25A10; loci on chromosome 19p13 Harboring LDLR and SUPG1 genes; several loci on chromosome 20 harboring RIN2, FAM182B, GDF5, PLCG1, HNF4A, and PLTP; and locus 22q13 that harbors CELSR1 and PPARA.

Similarly, there were several genomic regions that appeared associated in males only. These include several loci on chromosome 1 harboring MIER1, IL6R, LGR6 and SARG genes; loci on chromosome 2 harboring SH3YL1, GCKR and SPC25; chromosome 3p14 harboring FRMD4B; chromosome 5 including FBXO4 and SNCAIP; locus 7q21 harboring BRI3; loci on chromosome 8 harboring RBPMS and TMEM70; loci on chromosome 10 harboring FAM107B, MRC1L1, HKDC1, CYP26A1, PKD2L1 and NDUFB8; locus 11p15 harboring SBF2; locus 14q32 harboring SLC25A47; locus 15q15 harboring MAP1A; loci on chromosome 16 harboring BCAR1 and GCSH; chromosome 17 harboring SERPINF2, RAB5C and APOH; locus 18q21 harboring ATP8B1; locus 19q13.11 harboring HPN and locus 22q12 harboring APOL3.

Genes associated with metAgeGap in both males and females were enriched in cholesterol metabolism, PPAR signaling and regulation of Insulin-like Growth Factor (IGF) transport and uptake pathways among several others (**Supplementary Table 5**). Genes associated with metAgeGap in females only were enriched in cholesterol metabolism (**Supplementary Table 6**), while genes associated with metAgeGap in males only did not cluster in any specific pathway (**Supplementary Table 7**).

LD Score Regression (LDSC) analysis suggests significant genetic correlations of metAgeGap with Cystatin C, Hemoglobin A1c (HbA1c), estimated Glomerular Filtration Rate (eGFR), and albumin among others (**Supplementary Tables 8 & 9, Supplementary Figure 9**) in all sexes. The strength of correlation and significance for albumin was, however, stronger in males (r=-0.6, FDR= 2.24^*^10^-11^) compared to females (r=-0.24, FDR=3.48^*^10^-08^). In males only a significant genetic correlation of metAgeGap was observed with sex hormone binding globulin (SHBG), testosterone, edema, calcium, mouth ulcers and mean corpuscular hemoglobin concentration. In females, strongest significant genetic correlation of metAgeGap was observed with LDL (r=0.60, FDR=2.11^*^10^-54^) and apolipoprotein B (r=0.62, FDR=2.7^*^10^-53^), followed by obesity, various vascular pathologies, paternal illnesses and age at death, strenuous physical activity, eye problems, birth weight and age at menopause (**Supplementary Figure 9**).

### Proteins involved in detoxification of Reactive Oxygen Species decelerate metabolic aging

To identify proteins involved in accelerated/decelerated metabolic aging, we performed a proteome-wide association analysis of metAgeGap in the UK Biobank. In females, 1,739 out of 2,923 proteins were significantly associated (FDR < 0.05) with metAgeGap, of which 1,587 associated with older metabolic age and 152 with younger metabolic age (**Supplementary Table 10, Figure 3**). In males, 1,942 proteins were significantly associated with metAgeGap, of which 1,115 associated with older metabolic age and 827 with younger metabolic age. 906 proteins were consistently associated with older metabolic age and 52 proteins were consistently associated with younger metabolic age in both males and females. Proteins that associated with older metabolic age clustered in immune system regulation, coagulation, PI3K-Akt signaling, lysosome, apoptosis, intestinal IgA production, regulation of Insulin-like Growth Factor (IGF) transport/uptake, tumour necrosis factor (TNF), nuclear factor-κB (NF-κB) and MAP kinase (MAPK) signaling pathways among several others (**Supplementary Table 11**). Proteins that were associated with younger metabolic age clustered in cadherin binding and detoxification of reactive oxygen species (ROS) (**Supplementary Table 12**).

**Figure 3:**
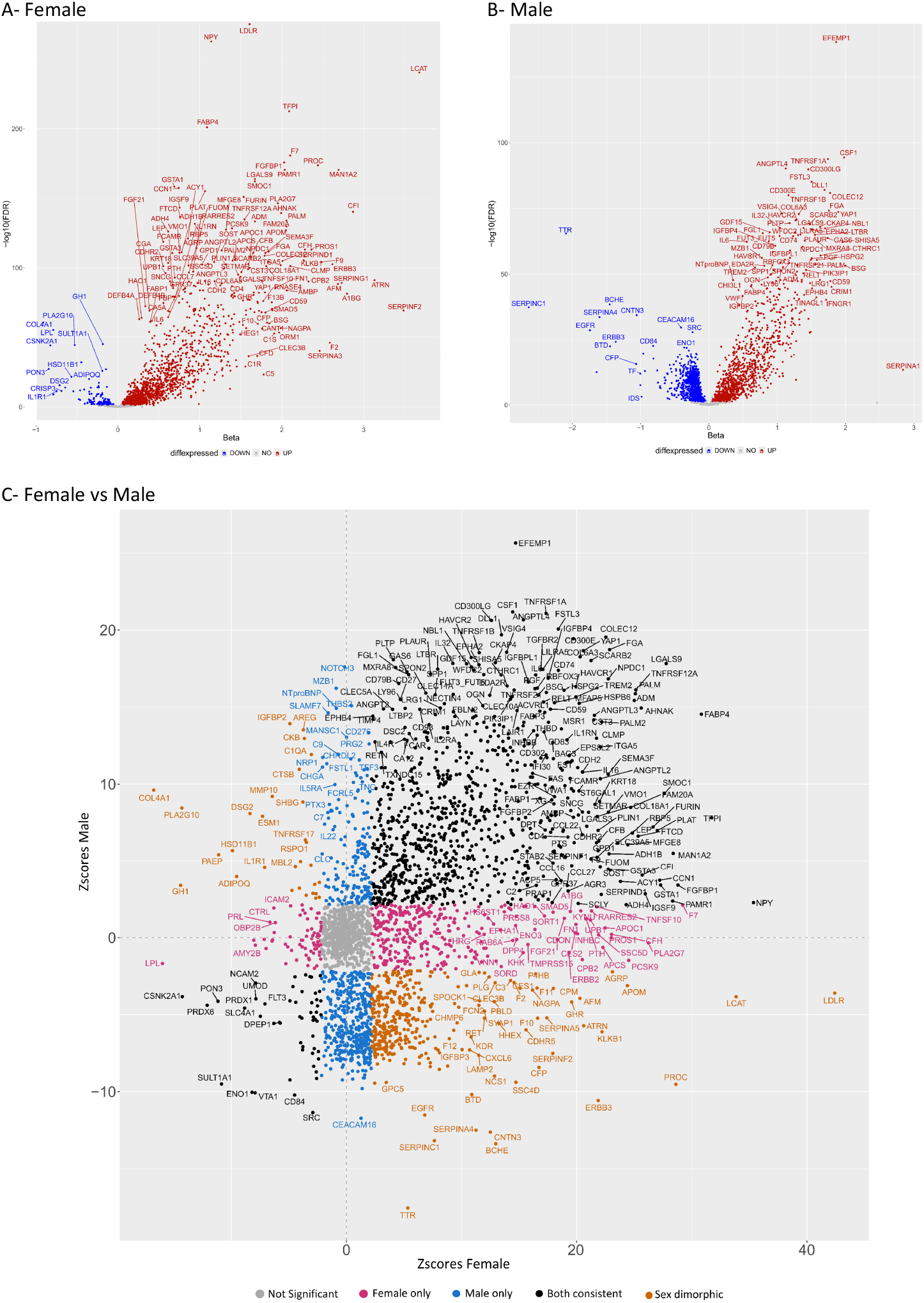
Proteome-wide association analysis of metAgeGap in males and females. **(A)** Volcano association plot of for females and **(B)** for males. Horizontal axis depicts the effect estimate and the vertical axis shows the strength of the association (-log10 of the FDR). **(C)** Comparison of associations (Z-scores) between males and females. Pink circles are protein associated in females only, blue in males only, black circles are proteins associated with metAgeGap in both sexes and orange are the proteins associated with metAgeGap in both sexes but with opposite effects (dimorphic).

When comparing males and females, 366 proteins were significantly associated with older metabolic age in females but younger metabolic age in males. These proteins include LDLR, which also showed a significant association in GWAS of metAgeGap in females but not in males, LCAT and APOM, all of which are cholesterol-related proteins. These 366 proteins clustered in mechanisms involved in hemostasis (**Supplementary Table 13**). 385 proteins showed association with metAgeGap in females only. These proteins were largely in hormone metabolism, immune system regulation and breast cancer (**Supplementary Table 14**). There were only 30 proteins that were significantly associated with older metabolic age in males but younger metabolic age in females. These proteins were involved in various bindings mechanisms and cytokine receptor activity (**Supplementary Table 15**). Further there were 588 proteins associated with metAgeGap in males only. These proteins were involved in apoptosis, cancers including colorectal, lung, pancreatic and prostate cancers, FOXO signaling in particular AKT-mediated inactivation of FOXO1A, AMPK signaling and various viral and bacterial infections (**Supplementary Table 16**).

### Genetics, lifestyle and sex hormones differentially influence aging associated sex dimorphic proteins

To gain insights into the top sex dimorphic aging proteins we evaluated their age distributions (**Figure 4**) and the impact of BMI, smoking, alcohol use, menopause and hormone replacement therapy (HRT) on the distribution of these proteins (**Supplementary Figures 10-15**). Since the top proteins associated with higher metabolic age in females are involved in lipid metabolism, we further evaluated the age distribution of density lipoprotein (LDL) (**Supplementary Figure 14**). BMI significantly increased expression of proteins involved in hemostasis (F7 & PROC) and lipid metabolism (LDLR, PCSK9 & LDL) in both males and females, however females in the normal weight and underweight categories also displayed high levels of these proteins and LDL over age, unlike normal and underweight males, suggesting that while BMI explains the differences within the females in metabolic aging, it does not explain the difference between males and females (**Supplementary Figure 10**). Current smoking significantly increased levels of blood clotting factors (F7, PROC and PLA2G7) and LDLR in females but not in males, which explains the observed association between smoking and metabolic aging in females. Smoking also significantly lowered the plasma levels of TTR and CEACAM16, both of which showed significant protective effects on metabolic aging in males (**Supplementary Figure 11**) but not in females. No noticeable differences were observed between males and females when the impact of alcohol use was evaluated (**Supplementary Figure 12**). Female-specific aging proteins (F7, PROC, LDLR and PCSK9) were influenced by menopause and to some extent HRT use (**Supplementary Figure 13**). No noteworthy difference was observed between females with low and high estradiol levels (**Supplementary Figure 14**). However, males with low testosterone levels showed significantly different levels of most of the aging associated sex dimorphic proteins (**Supplementary Figure 15**).

**Figure 4:**
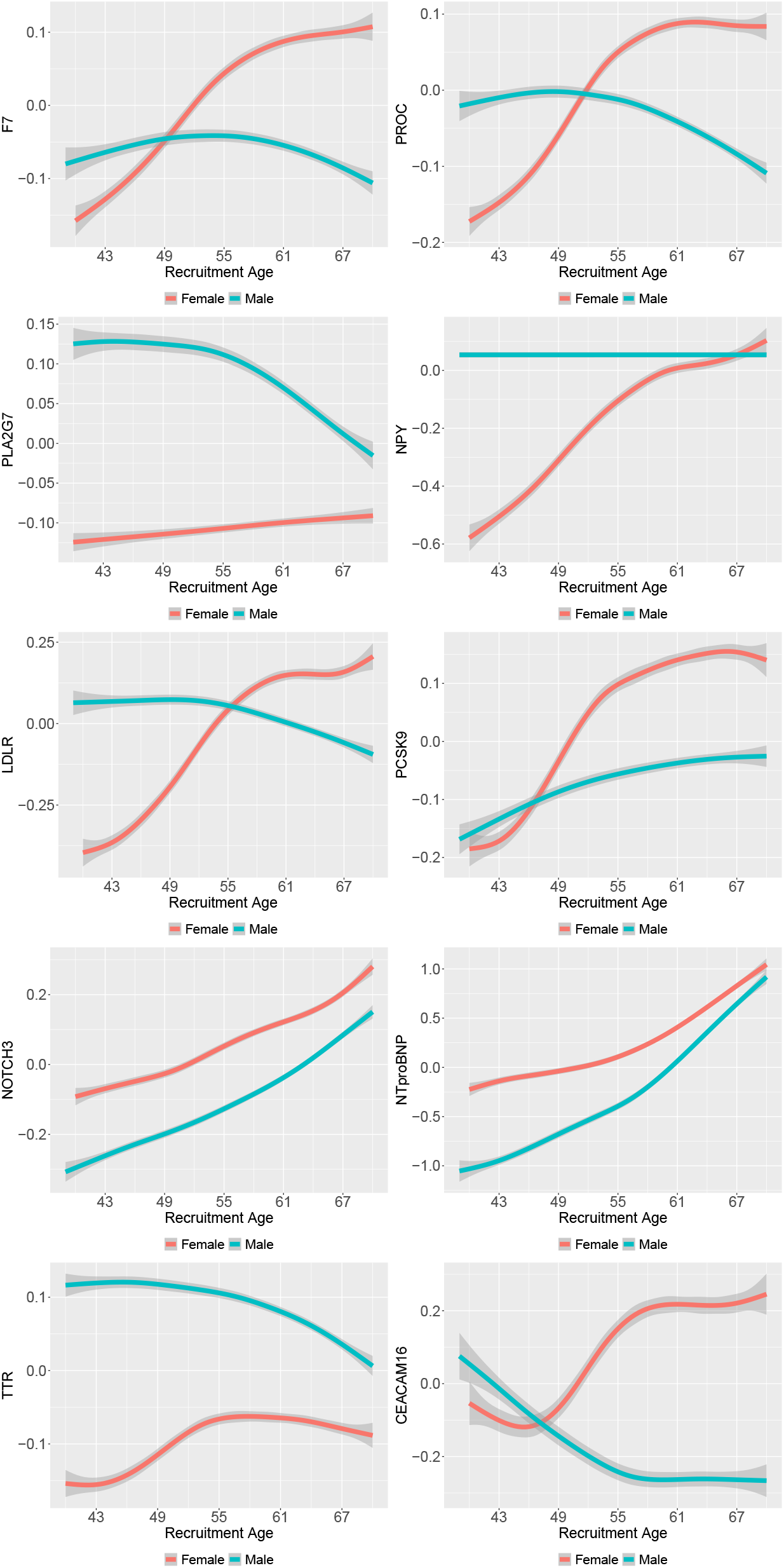
Sex specific age distribution of the top aging-related sex dimorphic proteins. Horizontal axis shows age at recruitment and vertical axis depicts the standardized levels of relative protein abundance.

To explore whether the protein quantitative loci (pQTLs; *cis & trans*) underlying these top proteins have differential influence on metabolic aging in males and females we compared the male and female specific summary statistics of all the pQTLs (**Supplementary Table 17**) from the GWAS of metAgeGap (**Supplementary Figure 16**). The most interesting finding was a particular missense variant (rs1260326_T) in *GCKR* gene, a locus that showed genome-wide significance in male specific GWAS only (**Supplementary Figure 6**), influenced the plasma levels of seven of these sex dimorphic proteins including F7, CEACAM16, LDLR, NOTCH3, PCSK9, TTR and NPY. Rs1260326_T was significantly negatively associated with metaAgeGap in males (beta=-0.23, p-value = 1.04^*^10^-40^) and showed suggestive positive association in females (beta=0.06, p-value=3.64^*^10^-06^) (**Supplementary Table 18**). The second most interesting gene was *MLX1PL*, whose multiple different variants influenced the levels of five proteins, including F7, LDLR, CEACAM16, NPY and TTR (**Supplementary Table 17**). All these variants showed inverse associations with metAgeGap in males and females (**Supplementary Table 18**). Among other pQTLs that showed differential effects in males and females include those in *PCSK9, CELSR2, SMARCA2/LDLR, BUD13-DT*, and *TIMD4/HAVCR1* that showed genome-wide significance in female-specific metAgeGap GWAS only and pQTLs in *CYP26A1, LPA* and *PRKCA* that showed genome-wide significance in males only. Two pQTLs in *APOE*, including rs7412_C and rs445925_G, also showed differential effects on metAgeGap in males and females. All these pQTLs influenced plasma levels of CEACAM16, F7, LDLR, NPY, PCSK9 and PLA2G7 (**Supplementary Table 17**).

### MetAgeGap predicts several common cancers in males but not in females

When comparing disease risks of the biologically youngest group (bottom 10% metAgeGap) with that of biologically oldest group (top 10% metAgeGap), youngest males had a higher cumulative risk for ischemic heart disease (IHD) than the oldest females, reflecting a higher baseline risk in males **(Figure 5)**. Further, the differences in cumulative risks of COPD and mortality between the top and bottom 10% were more pronounced in males than in females. The increase in cumulative risk for osteoarthritis was more gradual and linear in all sexes, with females being more at risk than males after the age of 50, suggesting a different risk trajectory in all sexes. For chronic kidney and liver diseases, no differences were observed between males and females, where biologically younger individuals showed significantly lower cumulative risks compared to biologically older individuals (**Figure 5, Supplementary Figure 17)**.

**Figure 5:**
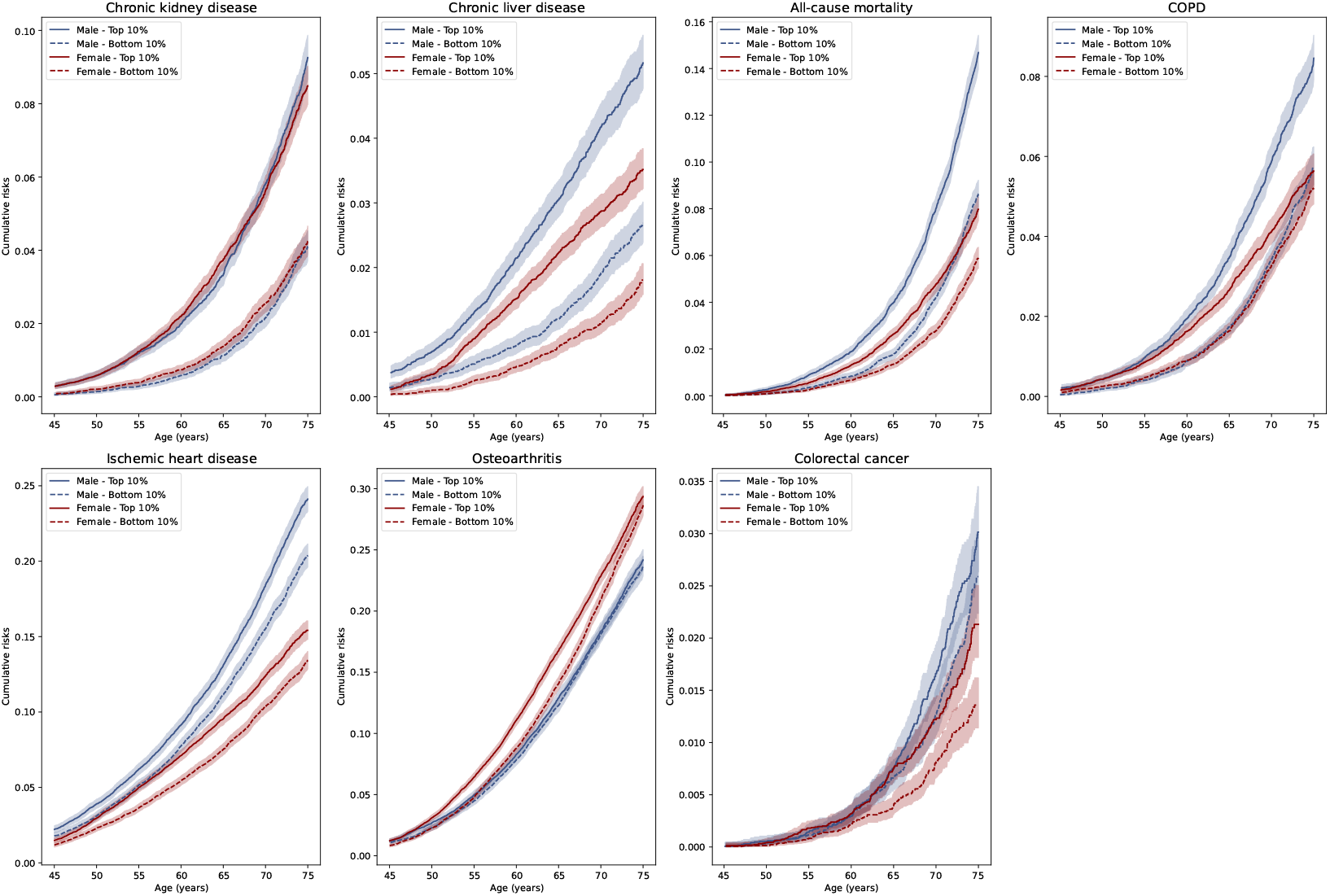
Cumulative incidence plot of top and bottom 10% of the metAgeGap in males and females for major incident diseases, and mortality. 95% confidence interval shown as lighter shading. X-axis denotes the chronological age and Y-axis denotes cumulative risk.

When studying the association of metabolic aging with 15 incident morbidities and mortality (**Figure 6**), stark differences were observed between males and females for cancers (**Supplementary Tables 19 & 20**). MetAgeGap was significantly associated with liver (HR=1.74, CI=1.52, 1.99), oesophageal (HR=1.30, CI=1.19, 1.42) and lung cancers (HR=1.10, CI=1.04, 1.17) only in males (**Figure 6A, 6B**). Adjusting for alcohol use and smoking did not attenuate these associations (**Supplementary Figures 18-21**). Colorectal cancer was significantly associated with metAgeGap in all sexes. In females after adjusting for BMI and physical activity, metAgeGap’s association with kidney (HR=1.11, CI=0.99, 1.24) and breast (HR=1.02, CI=1.01, 1.04) cancers attenuated.

Among non-cancer illnesses, except for neurodegenerative disorders (Alzheimer’s and Parkinson’s), metAgeGap was significantly associated with all diseases in males (**Figure 6C, 6D**). MetAgeGap was significantly associated with dementia (HR=1.07, CI=1.02, 1.12), ischemic stroke (HR=1.14, CI=1.10, 1.18), macular degeneration (HR=1.05, CI=1.01, 1.08), all strokes (HR=1.11, CI=1.08, 1.15), rheumatoid arthritis (HR=1.08, CI=1.04, 1.12), and osteoporosis (HR=1.11, CI=1.07, 1.16) in males but not in females (**Supplementary Tables 21 & 22**). The impact of metAgeGap on mortality and the risk of heart disease was far more pronounced in males. Adjusting for BMI led to a higher dilution of effects in females but not in males, especially for kidney, liver, and heart diseases, suggesting that BMI can have a different impact on aging in males and females (**Figure 6C, 6D**).

**Figure 6:**
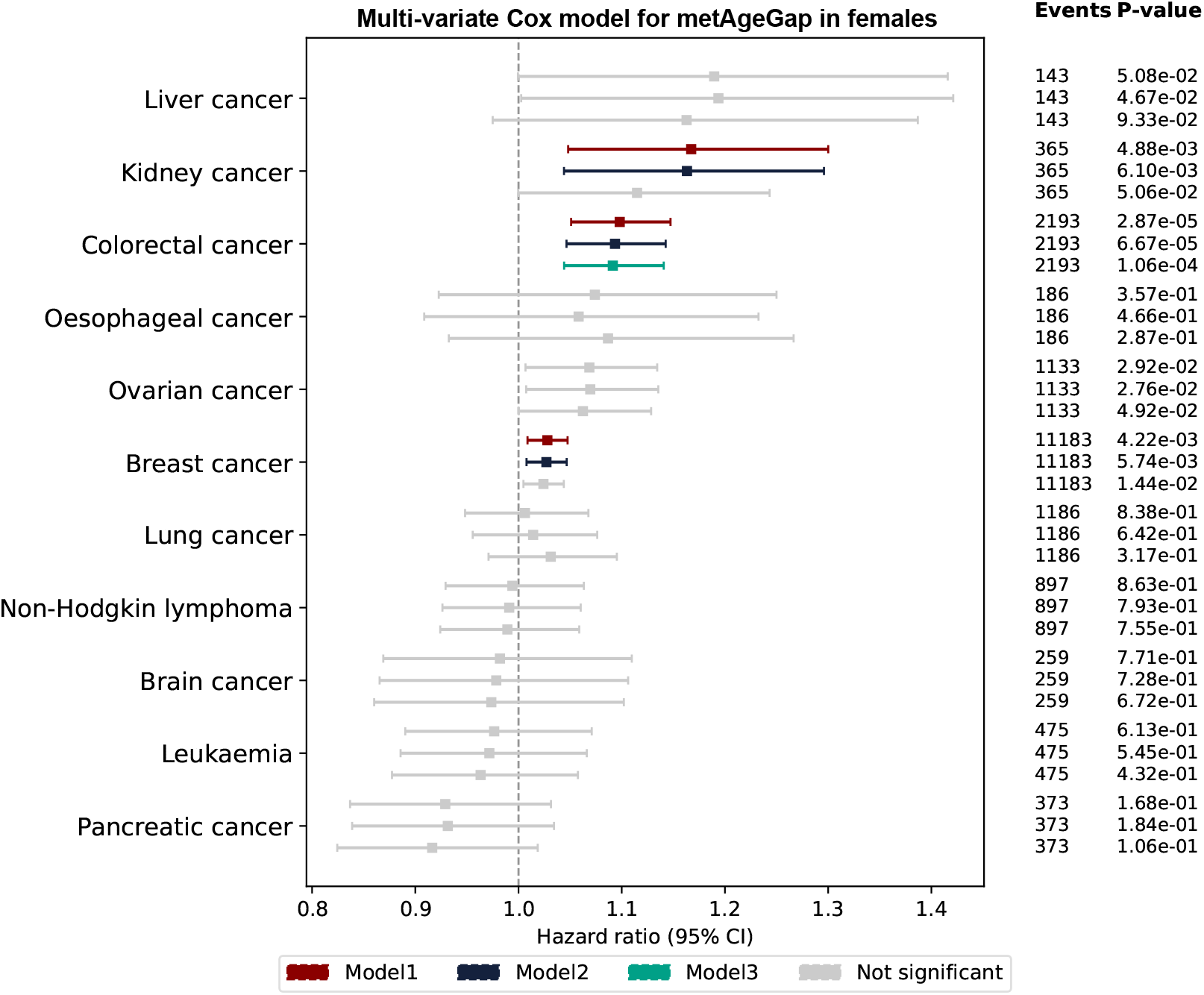
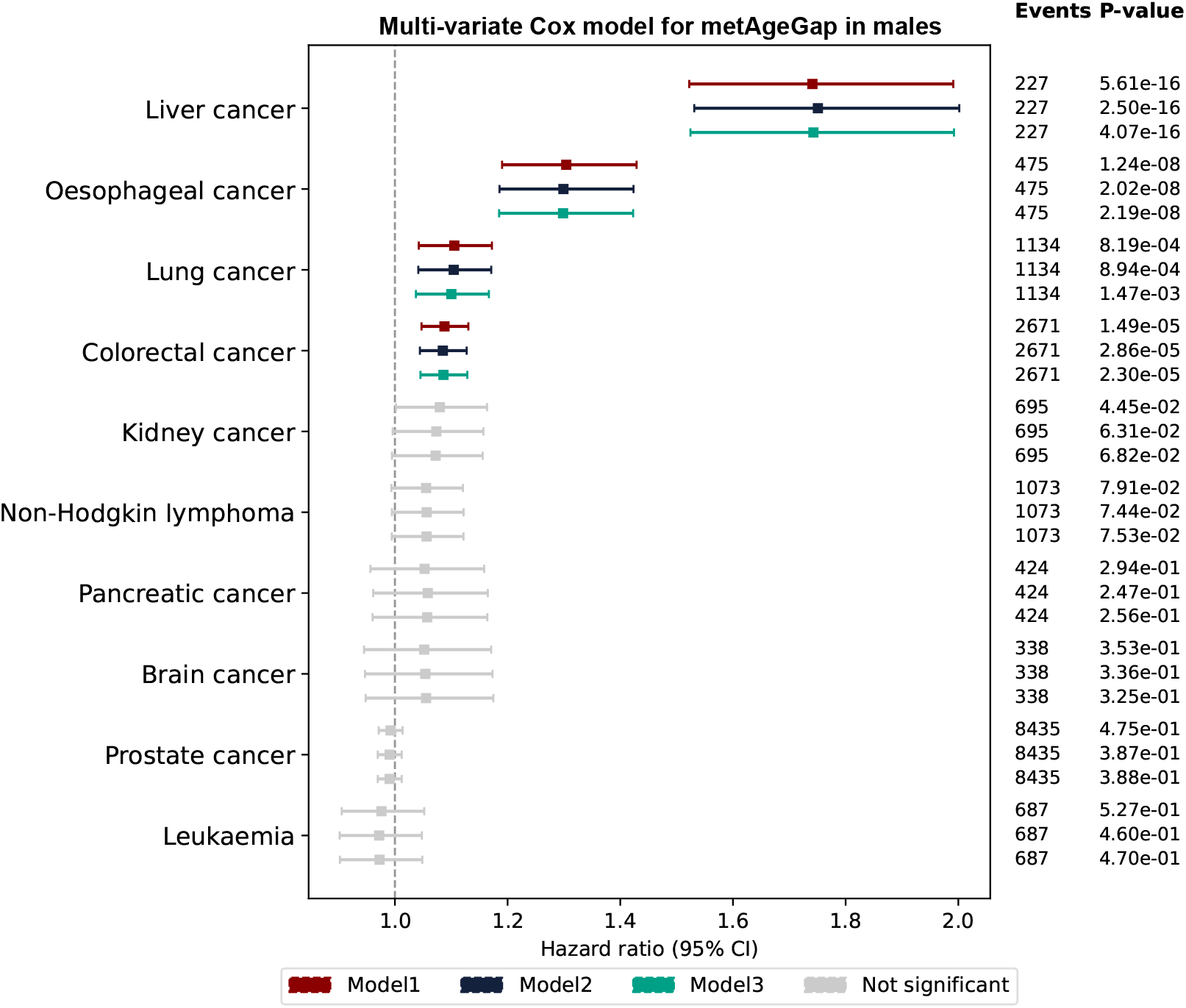
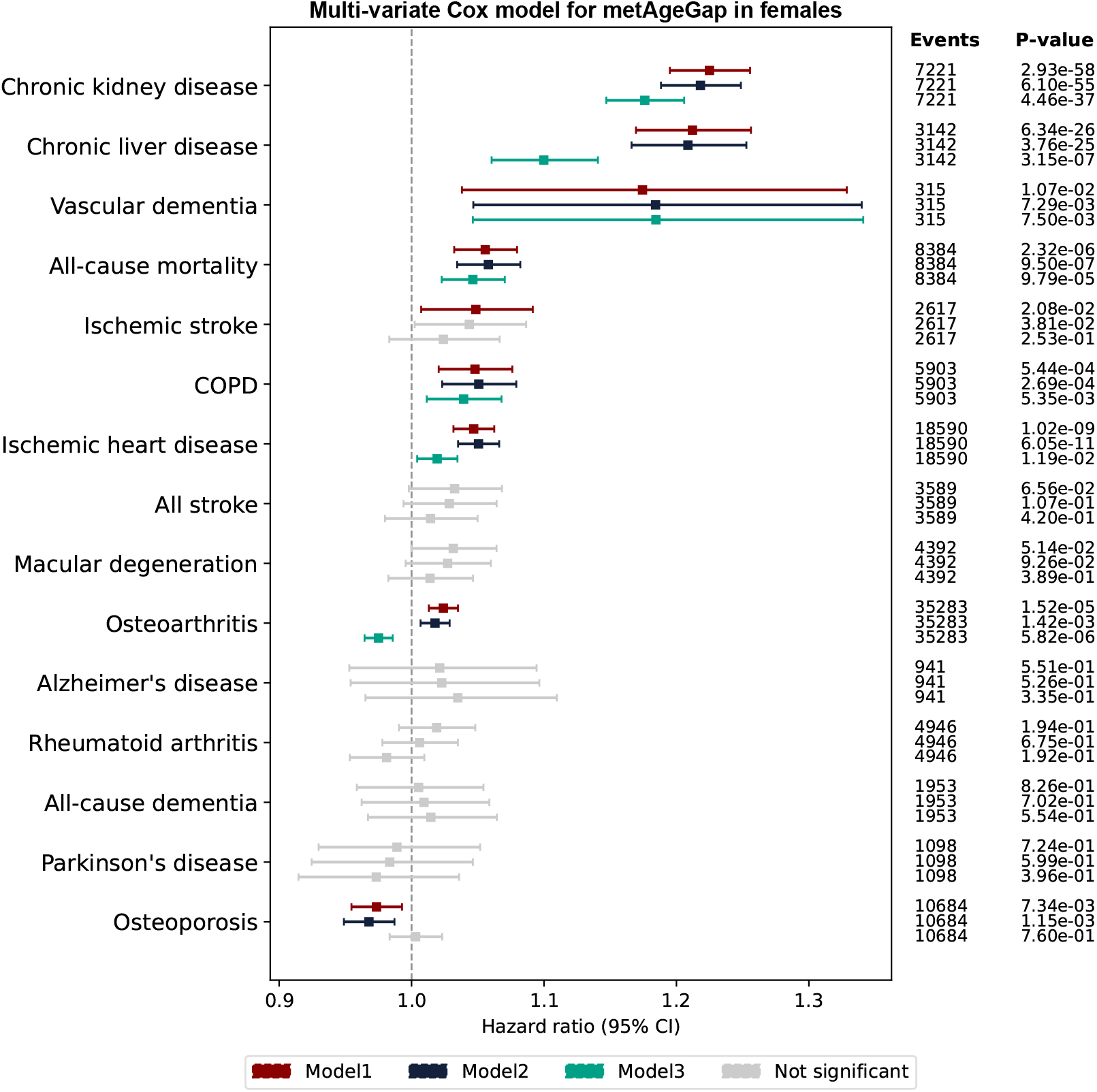
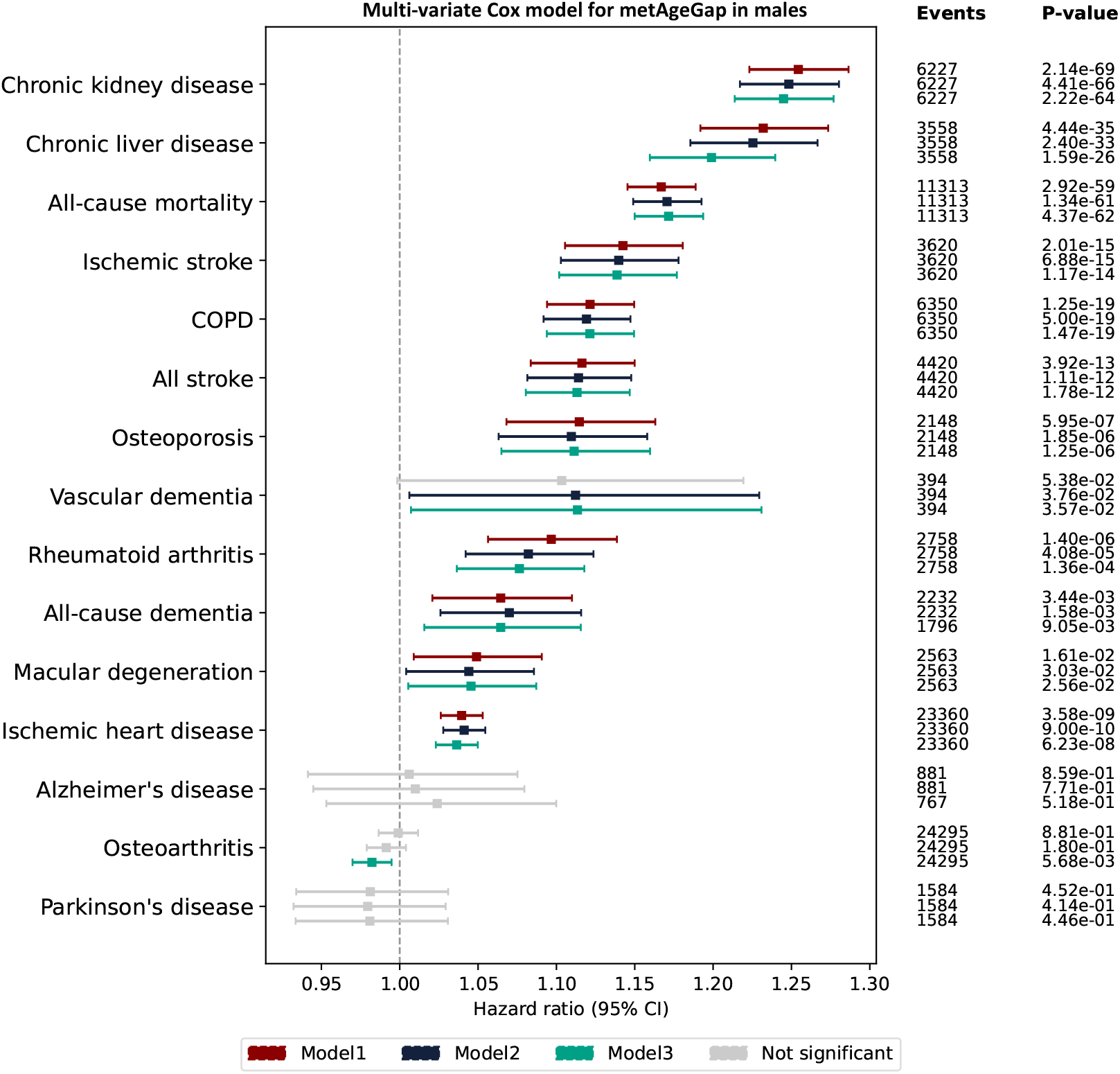
Forest plot of the association of metAgeGap with chronic diseases and mortality. **(A)** shows association between metAgeGap and various cancers in females using multi-variate cox proportional hazard model. In model 1, the exposure was metAgeGap with adjustment of chronological age. Model 2 was further adjusted for recruitment centre, Townsend deprivation index, and, ethnicity. Model 3 further adjusted for physical activity and BMI. P-values (p-val) were corrected for FDR multiple testing and non-significant associations after corrections were shown as grey color. **(B)** shows association between metAgeGap and various cancers in males using multi-variate cox proportional hazard model. **(C)** shows association between metAgeGap and health outcomes in females using multi-variate cox proportional hazard model. **(D)** shows association between metAgeGap and health outcomes in males using multi-variate cox proportional hazard model.

## Discussion

In this study, we developed and validated a sex-specific metabolic aging clock using NMR-based metabolome data from the UK Biobank, enabling us to investigate biological aging through a metabolic lens in a population of 488,318 individuals. Our findings show that metabolic aging, as captured by the metAgeGap metric, is a complex and sex-differentiated process with distinct biochemical, phenotypic, proteomic and genetic correlates.

First, that our models could predict chronological age with moderate accuracy (R^2^ = 0.29 for males, 0.37 for females) suggests that the metabolome holds reliable, albeit incomplete, clues about the biological clock. Interestingly, females consistently showed stronger prediction performance—a possible reflection of the tighter regulation or narrower variability of metabolic processes in women. While 61 features were common to both sexes—suggesting conserved core pathways—the presence of 32 female-specific and 15 male-specific features points to divergent metabolic processes driving aging. In females, unique contributions from cholines and phosphatidylcholines align with known roles in membrane biology and lipid signaling^34,35^, whereas males exhibited distinct associations with HDL particle size and triglyceride composition^36,37^, implicating differences in lipid transport and cardiovascular risk.

The metAgeGap metric showed strong and consistent associations with cardiometabolic risk factors, but the magnitude and pattern of these associations were sex-dependent. Obesity, BMI, smoking and blood pressure exhibited stronger associations in females, while type 2 diabetes showed the largest effect in males. These differences underscore that while the pathways to aging may be parallel, they are not identical and therefore sex should be considered when examining the metabolic determinants of biological age and their relevance to disease risk.

Perhaps the most intriguing finding of the current study is the association of puberty and hormonal exposures with metabolic aging. In women, later puberty, higher parity, later age at first and last live births and hormone use paint a portrait of youth preserved, with significantly younger metabolic ages. Meanwhile, in men, delayed signs of puberty — like voice breaking or facial hair — and number of children fathered were similarly linked to a slower metabolic clock. These findings suggest that earlier exposure to sex hormones accelerates biological aging in both sexes, consistent with hypotheses proposing a trade-off between early reproductive maturation and long-term somatic maintenance. These findings align with previous evidence suggesting that hormonal and physiological factors related to reproduction can modulate long-term metabolic health and aging trajectories^38,39^. However, our findings of protective effects of parity in both females and males contradict the evolutionary theories of aging, which hypothesize a trade-off between reproduction and lifespan^40-43^. Literature shows conflicting results about parity and longevity in women^40-43^ but somewhat consistent in males^44^. Our findings appear more plausible with the hypothesized effects of fetal michrochimerism on maternal health and longevity^45^. Further later age at birth has also been found to associate with longevity in females^44^.

Crucially, metAgeGap was significantly associated with incident morbidity and mortality outcomes, again revealing pronounced sex differences. Males with higher metabolic age showed elevated risks across nearly all disease categories, including cardiovascular, renal, hepatic, and oncological outcomes. Notably, liver, esophageal, and lung cancers exhibited strong associations in males, independent of lifestyle factors such as alcohol use and smoking. In contrast, females displayed fewer and weaker associations, although osteoarthritis risk increased significantly after midlife, and some disease risks (e.g., breast and kidney cancer) were attenuated after adjustment for BMI and physical activity. The attenuating effect of BMI was consistently more pronounced in females, suggesting that the interplay between adiposity and metabolic aging may differ by sex. These findings are consistent with earlier studies, which showed a stronger association between BMI and mortality in females than in males^46^. The sex difference can be explained by the fact that females tend to accumulate more subcutaneous fat, while males store more visceral fat^47-49^, which is more closely linked to metabolic risks, highlighting the importance of developing sex-specific clocks. The female body seemed to carry a certain metabolic resilience, especially for cancers and neurodegenerative diseases.

In the GWAS, 217 genes overlapped between males and females, indicating a shared genetic basis of metabolic aging, bringing out cholesterol metabolism and PPAR signaling as common pathways of metabolic aging. These included the *APOE* gene, which is one of the two genes consistently been associated with longevity in large-scale GWAS^33,50^ and has also shown sex dimorphic effects in earlier studies on plasma protein levels^51,52^. Of note is that a large number of genomic loci (147 vs 120) were identified in females, suggesting potentially higher polygenic complexity. Despite a larger gene set, no unique pathway was observed for female-specific genes beyond those shared. There were, however, some interesting genes including those involved in glucose transport, e.g., SLC2A2 and SLC2A6, lactose intolerance and gut microbiome composition LCT, circadian rhythm TIMELESS, virus receptors HAVCR1 and HAVCR2, and genes involved in metabolic and brain health CELSR2 and CELSR1 that showed no association with metabolic aging in males. In contrast to females, male-specific loci included genes such as IL6R, GCKR, SNCAIP, HKDC1 and APOH, some of which are implicated in inflammation and metabolism. Female-specific metAgeGap genetic correlations were observed with iron-deficiency anemia, obesity and cardio-vascular pathology, suggesting that metabolic aging in females may be more closely tied to cardiometabolic health, while male-specific metAgeGap genetic correlations with sex hormones, mouth ulcers and atrial fibrillation suggest a distinct health profile associated with male metabolic aging.

A key observation from this study is the enrichment of proteins related to immune regulation, coagulation, and several canonical signaling pathways (e.g., PI3K-Akt, NF-κB, MAPK, TNF, IGF) among those associated with older metabolic age. These pathways are well-established players in aging and age-related diseases. For instance, chronic low-grade inflammation (“inflammaging”)^53^ is increasingly recognized as a hallmark of aging and has been implicated in metabolic dysfunction, cardiovascular disease, and neurodegeneration^54^. The prominence of immune and inflammatory pathways in our analysis suggests that immunosenescence and dysregulated immune signaling may be central to the biological aging of metabolism.

Conversely, proteins linked to younger metabolic age were enriched in pathways involved in cell adhesion (cadherin binding) and oxidative stress detoxification. These processes are associated with cellular resilience and tissue integrity—features that are typically diminished with aging. The detoxification of reactive oxygen species (ROS) is critical for maintaining mitochondrial function and preventing oxidative damage, a major contributor to age-related decline^55^. The two most important antioxidant enzymes include superoxide dismutase (SOD) and glutathione peroxidase (GPx)^55^. In our study, we found low levels of SOD1 and peroxiredoxin 6 (PRDX6), which is a bifunctional enzyme with GPx activity, both in males and females with older metabolic age. The inverse association of these proteins with metAgeGap suggests potential protective mechanisms that could be targeted to slow down metabolic aging. Of note is that these redox mechanisms were not captured by the proteomic aging clock that we developed earlier although it showed much higher accuracy (R^2^ = 0.88) in predicting chronological age^56^.

While there was considerable overlap in proteins associated with metAgeGap across sexes, we observed marked divergence in both the number and functional roles of proteins that were uniquely or differentially associated. For example, 366 proteins were associated with older metabolic age in females but with younger metabolic age in males, and many of these were involved in lipid metabolism and hemostasis. Since we removed individuals who were on lipid lowering medication, these findings point towards sex disparities in lipid regulation, which is affected by menopause^57^ incidence, presentation and in particular treatment of cardiovascular diseases that remain underdiagnosed and undertreated in females compared to males thus affecting longevity in females^58^. Our study further shows that lifestyle factors such as smoking can worsen lipid metabolism and hemostasis in females but not in males across all ages.

The sex-specific proteomic associations further revealed pathways linked to cancer, hormone metabolism, and infectious disease susceptibility. In males, proteins uniquely associated with older metabolic age were enriched in apoptosis, FOXO and AMPK signaling—pathways with established roles in tumor suppression and longevity regulation^59,60^. The differential involvement of FOXO signaling, particularly via AKT-mediated inactivation of FOXO1A, aligns with evidence that metabolic stress and insulin resistance modulate aging-related pathways differently in males and females.

While this study is the largest of its kind to-date, there are, however, some limitations. First, the findings may not be generalizable to populations of non-European ancestries as the UK Biobank primarily includes individuals of European descent. Therefore, the impact of metabolic aging on disease risks in other ethnic groups remains to be explored. Second, the Nightingale metabolomics platform predominantly captures lipid-related metabolites, which explains the low predictive power in our study. This also led us to remove over 100k participants that were on lipid lowering medications, further impacting the statistical power of the study, particularly in studying the association of metAgeGap with rarer cancers. More metabolically diverse platforms may provide better resolution and insights into aging. Finally, the age range of the participants in the UK Biobank is relatively narrow, a wider age range may have provided a better fit. Nevertheless, with metAgeGap we have provided novel insights into the intrinsic and extrinsic factors leading to differences in metabolic aging in males and females.

In conclusion, although aging affects everyone, its pace and impact differ between males and females due to genetic, hormonal and environmental factors. The need to understand these differences extends beyond scientific curiosity, impacting real-world healthcare practices and outcomes. For example, cardiovascular disease is often underdiagnosed or misdiagnosed in females because they frequently experience atypical symptoms, such as nausea or fatigue, rather than the more well-known chest pain seen in males^61,62^. Our study demonstrated that the sex-specific metabolic aging clock is a powerful tool for measuring biological age and capturing aging signatures that are linked to common age-related diseases in both males and females. Our findings highlight the potential of this clock to identify the biological mechanisms underlying accelerated and decelerated aging and emphasize the importance of sex differences in aging processes. These clocks can shape our knowledge of sex-specific disease susceptibility, rates of physiological decline, and overall longevity, paving the path for more personalized prevention, treatment strategies and refined public health policies.

## Methods

### Study cohort

The study was performed in the UK Biobank (UKB). The UKB is a prospective cohort study including 502,505 participants recruited between 2006 and 2010^63^. Information on sociodemographic factors, lifestyle, early life, family history, psychosocial factors, health and medical history was collected through touchscreen questionnaires at baseline.

Linked hospital inpatient data, primary care data and cancer register data were accessed from the UKB data portal in August 2024, with a censoring date of November 30, 2023, December 31, 2023 and November 30, 2023 for participants recruited in England, Scotland, and Wales respectively. The follow-up time is between 8 and 16 years. Mortality data and cause of death information were accessed from the UKB data portal in August 2024, with a censoring date of November 30, 2022. The follow-up time is between 12 and 16 years. Methods and ICD diagnosis codes used to identify prevalent and incident chronic disease in UKB are shown in Supplementary Tables 23 & 24.

### Assessment of Metabolites

NMR metabolic biomarkers were generated by Nightingale Health, measuring 249 metabolic biomarkers for EDTA plasma samples from 488,318 UK Biobank participants^64^. Currently, data from ∼280,000 individuals is publicly available; this study had early access to the full dataset of ∼490,000 individuals, to be released in late 2025. Metabolic biomarkers were measured from randomly selected EDTA plasma samples (aliquot 3) using a high-throughput NMR-based metabolic biomarker profiling platform developed by Nightingale Health Ltd. The measurements took place between June 2019 and April 2020 (Phase 1) and between April 2020 and June 2022 (Phase 2) using eight spectrometers at Nightingale Health, based in Finland. The biomarkers span multiple metabolic pathways, including lipoprotein lipids in 14 subclasses, fatty acids and fatty acid compositions, as well as various low-molecular weight metabolites, such as amino acids, ketone bodies, and glycolysis metabolites quantified in molar concentration units.

### Assessment of Proteins

Proteomic profiling of 54,219 participants from the UK Biobank was carried out for protein analytes measured via the Olink Explore platform that links four Olink panels (Cardiometabolic, Inflammation, Neurology, and Oncology). UK Biobank Olink data are provided as Normalized Protein eXpression (NPX) values on a log2 scale. Details on sample selection, processing, and quality control are provided elsewhere^65^.

### Genotyping

Genotyping was conducted by Affymetrix using a bespoke BiLEVE Axiom array for ∼50K participants and the remaining ∼450K on the Affymetrix UK Biobank Axiom array. As the two arrays are broadly comparable with over 95% overlap in assessed gene variants, they were combined. Genetic data was phased prior to imputation with SHAPEIT3 followed by imputation using IMPUTE2. Details on genetic imputations are provided elsewhere^66^. The *APOE gene (allel*es *APOE ε2, APOE ε3, APOE ε4) was dir*ectly genotyped and defined by 2 single-nucleotide polymorphisms (SNPs), rs429358 and rs7412. Detailed information about the genotyping process and technical methods is available online.

### Statistical analysis

Descriptive analysis of population characteristics was performed using R package CBCgrps^67^. The study design and analysis pipeline are illustrated in Figure 1.

### Prediction of biological age

Biological age was predicted using 249 metabolites in males and females separately in a gradient boosting model. Samples were randomly split into a 70% training and 30% testing dataset. The model was first hyperparameter tuned using a Tree-structured Parzen Estimator (TPE) based method provided by the Optuna^68^ package in Python. Hyperparameters within a pre-set range were searched and optimized across 200 trials to maximize the 5-fold cross-validated Receiver Operating Characteristic (ROC) Area Under the Curve (AUC) score. After hyperparameter tuning, the performance of the best parameter in the training dataset with 5-fold cross-validation and in the 30% left out testing dataset was assessed.

### Feature interpretation and selection

To characterize the feature importance, SHapley Additive exPlanation (SHAP), a local tree explaining method based on game theory, was used^69^. SHAP calculates the contribution of each feature to the outcome in each individual and extends these local explanations to capture interactions between features directly. Compared to traditionally used permutation feature importance, SHAP plots can display the magnitude, prevalence, and direction of a feature’s effect. We then used a SHAP-based Boruta selection method provided by the shap-hypetune package^70^ to select all relevant features contributing to smoking status prediction. By constructing randomly permuted shadow features, Boruta compares the mean absolute SHAP values between input features and shadow features and only keeps features if they perform better than the best randomized features. In our study, we performed 200 iterations of the algorithm, and the features within the tail 5% were rejected. The model with Boruta selected features was hyperparameter tuned again before further analysis.

### Metabolic Age difference (metAgeGap)

MetAgeGap for the full UKB sample (n=488,318) was predicted using a robust method to mitigate the risk of overfitting. This process involved employing 5-fold cross-validation to ensure the reliability of the results. After identifying the best hyperparameters and selecting the proteins using the Boruta method, a gradient boosting model was trained within each fold. Subsequently, the predicted biological age for the corresponding test set was generated. MetAgeGap was calculated as the difference between the predicted metabolic age of an individual and the mean predicted metabolic age of the population belonging to the same chronological age group. This approach allowed for a robust estimation of the biological age across the UK Biobank cohort.

### Association of lifestyle, clinical biomarkers and risk factors with metAgeGap

To test the association of self-reported lifestyle factors, blood biochemistry biomarkers, and clinical risk factors with metAgeGap, generalized linear models from statsmodel v.0.14.0 package^71^ were used. For continuous exposure variables, standardization was applied before inclusion in the models. For questionnaire data, responses including ‘Do not know’ or ‘Prefer not to say’, were set to missing. Associations were adjusted for recruitment center, ethnicity, education years and Townsend deprivation index. FDR was applied to correct for multiple testing.

### Association metAgeGap with future health-related outcomes

To test the association between metAgeGap and incident health outcomes, all prevalent cases were removed beforehand. The multi-variate cox proportional hazard model provided by the lifeline v.0.27.8 package^72^ was used with a pre-set step size of 0.1. Survival outcomes were defined using follow-up time to event and the binary incident event indicator. For all incident outcomes in the whole UKB population, three successive models were tested with an increasing number of covariates: model 1 adjusted for chronological age; model 2 was adjusted for recruitment centre, Townsend deprivation index, and ethnicity; model 3 was further adjusted for physical activity and BMI; model 4 was further adjusted for smoking status, and alcohol frequency. P-values of the hazard ratio were corrected for FDR multiple testing. Forest plots were generated with a minimum sample size threshold of 80 to ensure adequate statistical power and reliable interpretation.

Cumulative incidence plots were generated utilizing the KaplanMeierFitter function from the lifelines package. Due to limitations in case numbers or at-risk numbers at both ends, the x-axis of the plot was constrained to the age range of 45 to 75. This adjustment ensured a more focused visualization of the cumulative incidence curve within a clinically relevant age range. P-values between cumulative incidence curves were calculated using log-rank test with adjustment for FDR multiple testing.

### GWAS of metAgeGap

Sex-specific GWAS were conducted using SAIGE^73^ software version 1.09. For constructing a genetic relationship matrix (GRM) in step 1, we used the pruned genotype dataset. Genotype pruning was conducted in PLINK^74^ software using the ‘indep-pairwise’ option with r^2^ of 0.5, a window size of 1000 markers and a step size of 100 markers. We further used ‘LOCO= TRUE’ option to construct the GRM. The GWAS analyses were adjusted for age, ethnicity, batch effects, and 40 genetic principal components identified within UKB genotyping data^66^. Identification of independent genome-wide significant loci (r2) and annotation was performed using SNP2GENE function in FUMA^75^. Gene enrichment analysis was performed for the genes in the independent loci using STRINGS database. LD score regression was performed using online tools Complex-Traits Genetics Visual Labs^76^ (CTG-VL) where summary statistics of GWAS against metAgeGap were correlated to publicly available GWAS result of 1,461 traits in the database. Significant correlations were identified if FDR adjusted p-value was smaller than 0.05.

### Proteome-wide association analysis of metAgeGap

MetAgeGap was tested for association with each of the 2,922 proteins measured on the OLINK platform in a linear regression model in R software (version 4.3.2) for males and females separately. The regression analysis was adjusted for technical confounders like plate and batch effects. FDR was applied to correct for multiple testing. Protein pathway analysis was performed using STRING database. Since we were interested in identifying differences between males and females, we applied an agnostic approach using all human genes available in the database as the background.

## Data Availability

UKB data are available through a procedure described at https://www.
ukbiobank.ac.uk/enable-your-research.

## Acknowledgements

All UK Biobank data was accessed under UK Biobank Application number 30418. This work was supported by the Centre of Artificial Intelligence in Precision Medicines (CAIPM), King Abdulaziz University, Jeddah, Saudi Arabia. We thank Nightingale Health Plc for providing early access to the UK Biobank dataset.

## Funding and conflict of interests

The computational aspects of this research were supported by the Wellcome Trust Core Award Grant Number 203141/Z/16/Z and the Oxford NIHR BRC. The views expressed are those of the author(s) and not necessarily those of the NHS, the NIHR or the Department of Health. S.X and S.B.H and B.H are funded by CAIPM, King Abdulaziz University, Jeddah, Saudi Arabia. P.K is funded by the US National Institute on Aging (NIH). R.K.D is an inventor on a series of patents on use of metabolomics for the diagnosis and treatment of CNS diseases and holds equity in Metabolon Inc., Chymia LLC and Metabosensor. This project was enabled in part by the Alzheimer’s Gut Microbiome Project (AGMP), supported by the National Institute on Aging grants: 1U19AG063744 and 3U19AG063744-04S1, awarded to R.K.D at Duke University in partnership with multiple academic institutions. As such, the investigators within the AGMP not listed in this publication’s authors’ list, provided analysis-ready data, but did not participate in designing the study, conducting the analyses or writing of this manuscript. A listing of AGMP investigators can be found at https://alzheimergut.org/meet-the-team/. A complete listing of the AD Metabolomics Consortium (ADMC) investigators can be found at: https://sites.duke.edu/adnimetab/team/. In addition, this work was supported by the Alzheimer Disease Metabolomics Consortium which is a part of NIA’s national initiatives AMP-AD (3U01AG061359, 3U01 AG024904-09S4). Najaf Amin is funded by NIH and Oxford-GSK Institute of Molecular and Computational Medicine (IMCM). Cornelia M van Duijn is supported by the NIH, NovoNordisk, the IMCM, CAIPM of the University of Oxford and King Abdul Aziz University, Alzheimer Research UK (ARUK), UK National Institute for Health and Care Research (NIHR) Oxford Research Center (BRC), ZonMW (Delta Dementie) and Alzheimer Nederland. Cornelia M van Duijn is currently the Research Director Brain Health of the Health Data Research UK (HDR UK) and the UK Dementia Research Institute (UK DRI), working in partnership with Dementias Platform UK (DPUK). M Austin Argentieri was funded by NIH grant number 5U01AG061359-05.

## Author contributions

Conceptualization: S.X, B.L, R.K.D, N.A, C.vD

Methodology: S.X, A.A, B.L, G.D

Analysis and visualization: N.A, P.K, S.X

Data extraction, preparation, and management in UKB: P.K

Supervision: A.J.NH, R.K.D, N.A, C.vD

Writing-original draft: P.K, S.X, N.A

Writing-review & editing: All authors

## Ethics approval

UK Biobank data use (Project Application Number 30418) was approved by the UK Biobank according to their established access procedures. UK Biobank has approval from the North West Multi-centre Research Ethics Committee (MREC) as a Research Tissue Bank (RTB), and as such researchers using UK Biobank data do not require separate ethical clearance and can operate under the RTB approval. Ethical approvals were granted and have been maintained by the relevant institutional ethical research committees in the UK.

### Data Access Statement

UK Biobank data are available through a procedure described at: https://www.ukbiobank.ac.uk/enable-your-research.

## List of Supplementary Materials

### Supplementary Information

Figures 1 – 21

Tables 1 – 24

### Supplementary File

This single Excel file contains Supplementary Tables 1 – 24 (each table is a sheet in the document).

Supplementary document has Supplementary figures.

## Notes

### Competing Interest Statement

The authors have declared no competing interest.

## References

1. Dattani, S. & Rodés-Guirao, L. Why do women live longer than men? OurWorldinData.org. (2023).

2. Gordon, E.H. et al. Sex differences in frailty: a systematic review and meta-analysis. Experimental gerontology 89, 30–40 (2017).

3. Peiffer, J.J. et al. Strength and functional characteristics of men and women 65 years and older. Rejuvenation Research 13, 75–82 (2010).

4. Nasiri, M.J. et al. COVID-19 clinical characteristics, and sex-specific risk of mortality: systematic review and meta-analysis. Frontiers in medicine 7, 459 (2020).

5. Pradhan, A. & Olsson, P.-E. Sex differences in severity and mortality from COVID-19: are males more vulnerable? Biology of sex Differences 11, 53 (2020).

6. Scully, E.P., Haverfield, J., Ursin, R.L., Tannenbaum, C. & Klein, S.L. xConsidering how biological sex impacts immune responses and COVID-19 outcomes. Nature Reviews Immunology 20, 442–447 (2020).

7. Zarulli, V. et al. Women live longer than men even during severe famines and epidemics. Proceedings of the National Academy of Sciences 115, E832–E840 (2018).

8. Gillies, G.E. & McArthur, S. Estrogen actions in the brain and the basis for differential action in men and women: a case for sex-specific medicines. Pharmacological reviews 62, 155–198 (2010).

9. Palmisano, B.T., Zhu, L., Eckel, R.H. & Stafford, J.M. Sex differences in lipid and lipoprotein metabolism. Molecular metabolism 15, 45–55 (2018).

10. Hägg, S. & Jylhävä, J. Sex differences in biological aging with a focus on human studies. Elife 10, e63425 (2021).

11. Brooks, R.C. & Garratt, M.G. Life history evolution, reproduction, and the origins of sex-dependent aging and longevity. Annals of the New York Academy of Sciences 1389, 92–107 (2017).

12. Gardner, M. et al. Gender and telomere length: systematic review and meta-analysis. Experimental gerontology 51, 15–27 (2014).

13. Lulkiewicz, M., Bajsert, J., Kopczynski, P., Barczak, W. & Rubis, B. Telomere length: how the length makes a difference. Molecular Biology Reports 47, 7181–7188 (2020).

14. Horstman, A.M., Dillon, E.L., Urban, R.J. & Sheffield-Moore, M. The role of androgens and estrogens on healthy aging and longevity. Journals of Gerontology Series A: Biomedical Sciences and Medical Sciences 67, 1140–1152 (2012).

15. Chang, E., Varghese, M. & Singer, K. Gender and Sex Differences in Adipose Tissue. Curr Diab Rep 18, 69 (2018).

16. Muscogiuri, G. et al. Obesity: a gender-view. J Endocrinol Invest 47, 299–306 (2024).

17. Harris, C.R. & Jenkins, M. Gender Differences in Risk Assessment: Why do Women Take Fewer Risksthan Men? Judgment and Decision making 1, 48–63 (2006).

18. Griswold, M.G. et al. Alcohol use and burden for 195 countries and territories, 1990–2016: a systematic analysis for the Global Burden of Disease Study 2016. The Lancet 392, 1015–1035 (2018).

19. Lipsky, M.S., Su, S., Crespo, C.J. & Hung, M. Men and oral health: a review of sex and gender differences. American journal of men’s health 15, 15579883211016361 (2021).

20. Villiers-Tuthill, A., Copley, A., McGee, H. & Morgan, K. The relationship of tobacco and alcohol use with ageing self-perceptions in older people in Ireland. BMC public health 16, 1–10 (2016).

21. Johnson, A.A. et al. The role of DNA methylation in aging, rejuvenation, and age-related disease. Rejuvenation research 15, 483–494 (2012).

22. Yusipov, I. et al. Age-related DNA methylation changes are sex-specific: a comprehensive assessment. Aging (Albany NY) 12, 24057 (2020).

23. Hertel, J. et al. Measuring biological age via metabonomics: the metabolic age score. Journal of proteome research 15, 400–410 (2016).

24. Horvath, S. DNA methylation age of human tissues and cell types. Genome biology 14, 1–20 (2013).

25. Lehallier, B. et al. Undulating changes in human plasma proteome profiles across the lifespan. Nature medicine 25, 1843–1850 (2019).

26. Lu, A.T. et al. DNA methylation GrimAge strongly predicts lifespan and healthspan. Aging (albany NY) 11, 303 (2019).

27. Menni, C. et al. Metabolomic markers reveal novel pathways of ageing and early development in human populations. International journal of epidemiology 42, 1111–1119 (2013).

28. Robinson, O. et al. Determinants of accelerated metabolomic and epigenetic aging in a UK cohort. Aging cell 19, e13149 (2020).

29. Rutledge, J., Oh, H. & Wyss-Coray, T. Measuring biological age using omics data. Nature Reviews Genetics 23, 715–727 (2022).

30. Tanaka, T. et al. Plasma proteomic biomarker signature of age predicts health and life span. Elife 9, e61073 (2020).

31. Van Den Akker, E.B. et al. Metabolic age based on the BBMRI-NL 1H-NMR metabolomics repository as biomarker of age-related disease. Circulation: Genomic and Precision Medicine 13, 541–547 (2020).

32. Reicher, L. et al. Phenome-wide associations of human aging uncover sex-specific dynamics. Nature Aging 4, 1643–1655 (2024).

33. Deelen, J. et al. A meta-analysis of genome-wide association studies identifies multiple longevity genes. Nature Communications 10, 3669 (2019).

34. Ridgway, N.D. The role of phosphatidylcholine and choline metabolites to cell proliferation and survival. Critical Reviews in Biochemistry and Molecular Biology 48, 20–38 (2013).

35. Sivanesan, S., Taylor, A., Zhang, J. & Bakovic, M. Betaine and Choline Improve Lipid Homeostasis in Obesity by Participation in Mitochondrial Oxidative Demethylation. Frontiers in Nutrition 5 (2018).

36. El Harchaoui, K. et al. High-density lipoprotein particle size and concentration and coronary risk. Annals of Internal Medicine 150, 84–93 (2009).

37. Miller, M. et al. Triglycerides and Cardiovascular Disease. Circulation 123, 2292–2333 (2011).

38. Fan, G. et al. Reproductive factors and biological aging: the association with all-cause and cause-specific premature mortality. Human Reproduction (Oxford, England) 40, 148–156 (2025).

39. Traub, M.L. & Santoro, N. Reproductive aging and its consequences for general health. Annals of the New York Academy of Sciences 1204, 179–187 (2010).

40. Kirkwood, T.B.L. Evolution of ageing. Nature 270, 301–304 (1977).

41. Kirkwood, T.B.L. & Rose, M.R. Evolution of senescence: late survival sacrificed for reproduction. Philosophical Transactions of the Royal Society of London. Series B: Biological Sciences 332, 15–24 (1997).

42. Kuningas, M. et al. The relationship between fertility and lifespan in humans. AGE 33, 615–622 (2011).

43. Williams, G.C. Pleiotropy, Natural Selection, and the Evolution of Senescence. Science of Aging Knowledge Environment 2001, cp13–cp13 (2001).

44. McArdle, P.F. et al. Does Having Children Extend Life Span? A Genealogical Study of Parity and Longevity in the Amish. The Journals of Gerontology: Series A 61, 190–195 (2006).

45. O’Donoghue, K. Fetal microchimerism and maternal health during and after pregnancy. Obstet Med 1, 56–64 (2008).

46. Tauqeer, Z., Gomez, G. & Stanford, F.C. Obesity in women: insights for the clinician. Journal of Women’s Health 27, 444–457 (2018).

47. Agrawal, S. et al. Inherited basis of visceral, abdominal subcutaneous and gluteofemoral fat depots. Nature Communications 13, 3771 (2022).

48. Chaston, T.B. & Dixon, J.B. Factors associated with percent change in visceral versus subcutaneous abdominal fat during weight loss: findings from a systematic review. International Journal of Obesity 32, 619–628 (2008).

49. Kuk, J.L. et al. Visceral fat is an independent predictor of all-cause mortality in men. Obesity (Silver Spring, Md.) 14, 336–341 (2006).

50. Timmers, P.R. et al. Genomics of 1 million parent lifespans implicates novel pathways and common diseases and distinguishes survival chances. eLife 8, e39856 (2019).

51. Bhak, Y. et al. Identification and replication of sex-dimorphic protein quantitative trait loci across multiple ancestries and their associations with diseases. Sci Rep 15, 31721 (2025).

52. Koprulu, M. et al. Sex differences in the genetic regulation of the human plasma proteome. Nat Commun 16, 4001 (2025).

53. Fulop, T. et al. Immunology of Aging: the Birth of Inflammaging. Clinical Reviews in Allergy & Immunology 64, 109–122 (2023).

54. Ferrucci, L. & Fabbri, E. Inflammageing: chronic inflammation in ageing, cardiovascular disease, and frailty. Nature Reviews Cardiology 15, 505–522 (2018).

55. Jomova, K. et al. Reactive oxygen species, toxicity, oxidative stress, and antioxidants: chronic diseases and aging. Archives of Toxicology 97, 2499–2574 (2023).

56. Argentieri, M.A. et al. Proteomic aging clock predicts mortality and risk of common age-related diseases in diverse populations. Nature Medicine 30, 2450–2460 (2024).

57. Ko, S.-H. & Kim, H.-S. Menopause-Associated Lipid Metabolic Disorders and Foods Beneficial for Postmenopausal Women. Nutrients 12, 202 (2020).

58. Vogel, B. et al. The Lancet women and cardiovascular disease Commission: reducing the global burden by 2030. The Lancet 397, 2385–2438 (2021).

59. Stancu, A.L. AMPK activation can delay aging. Discoveries 3, e53.

60. Yadav, R.K., Chauhan, A.S., Zhuang, L. & Gan, B. FoxO transcription factors in cancer metabolism. Seminars in Cancer Biology 50, 65–76 (2018).

61. Canto, J.G. et al. Symptom Presentation of Women With Acute Coronary Syndromes: Myth vs Reality. Archives of Internal Medicine 167, 2405–2413 (2007).

62. Joseph, N.M., Ramamoorthy, L. & Satheesh, S. Atypical Manifestations of Women Presenting with Myocardial Infarction at Tertiary Health Care Center: An Analytical Study. Journal of Mid-Life Health 12, 219–224 (2021).

63. Palmer, L.J. UK Biobank: bank on it. The Lancet 369, 1980–1982 (2007).

64. Nightingale Health Biobank Collaborative, G. Metabolomic and genomic prediction of common diseases in 700,217 participants in three national biobanks. Nat Commun 15, 10092 (2024).

65. Sun, B.B. et al. Plasma proteomic associations with genetics and health in the UK Biobank. Nature 622, 329–338 (2023).

66. Bycroft, C. et al. The UK Biobank resource with deep phenotyping and genomic data. Nature 562, 203–209 (2018).

67. Zhang, Z., Gayle, A.A., Wang, J., Zhang, H. & Cardinal-Fernandez, P. Comparing baseline characteristics between groups: an introduction to the CBCgrps package. Annals of Translational Medicine 5 (2017).

68. Akiba, T., Sano, S., Yanase, T., Ohta, T. & Koyama, M. Optuna: A Next-generation Hyperparameter Optimization Framework. 2623–2631 (Association for Computing Machinery, 2019).

69. Lundberg, S.M. et al. From local explanations to global understanding with explainable AI for trees. Nature machine intelligence 2, 56–67 (2020).

70. Cerliani, M. Shap-hypetune, (PyPi, 2022).

71. Seabold, S. & Perktold, J. Statsmodels: econometric and statistical modeling with python. SciPy 7 (2010).

72. Davidson-Pilon, C. lifelines: survival analysis in Python. Journal of Open Source Software 4, 1317 (2019).

73. Zhou, W. et al. SAIGE-GENE+ improves the efficiency and accuracy of set-based rare variant association tests. Nature Genetics 54, 1466–1469 (2022).

74. Purcell, S. et al. PLINK: A Tool Set for Whole-Genome Association and Population-Based Linkage Analyses. The American Journal of Human Genetics 81, 559–575 (2007).

75. Watanabe, K., Taskesen, E., van Bochoven, A. & Posthuma, D. Functional mapping and annotation of genetic associations with FUMA. Nature Communications 8, 1826 (2017).

76. Cuéllar-Partida, G. et al. Complex-Traits Genetics Virtual Lab: A community-driven web platform for post-GWAS analyses. (2019).

