## Supplementary figures for "Ticking Differently: Elucidating Sexual Dimorphism in Human Aging through Metabolomics, Proteomics and Genomics"

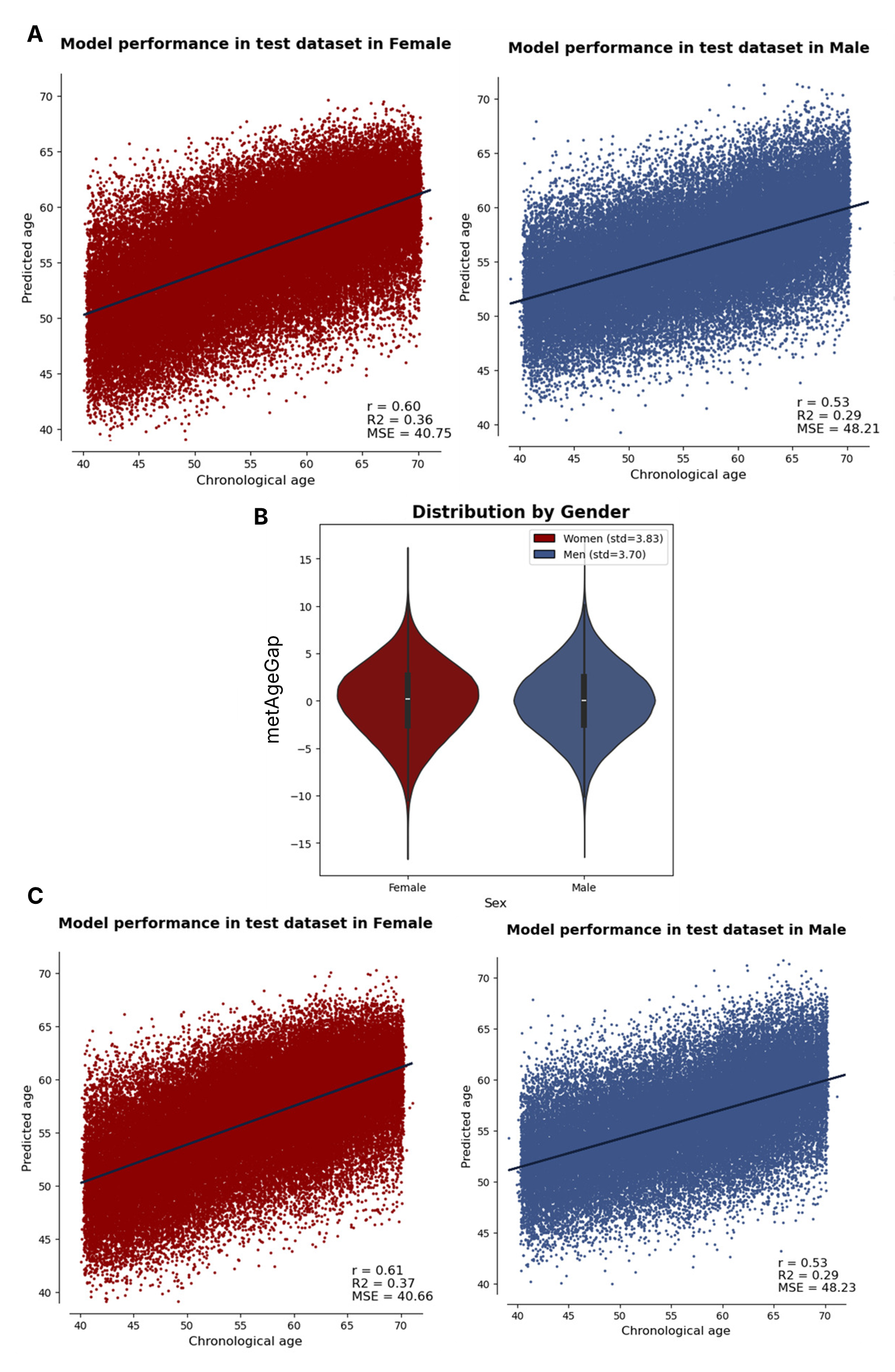

**Supplementary Figure 1: (A)** Model performance in predicting age of participants in the test set in the model trained using all metabolites in females and males respectively. **(B)** Distribution of residual between chronological age and predicted metabolic age - metabolic age gap (metAgeGap) in males and females. **(C)** Model performance in predicting age of participants in the test set in the model trained using 93, and 76 metabolites selected by Boruta in females and males respectively.

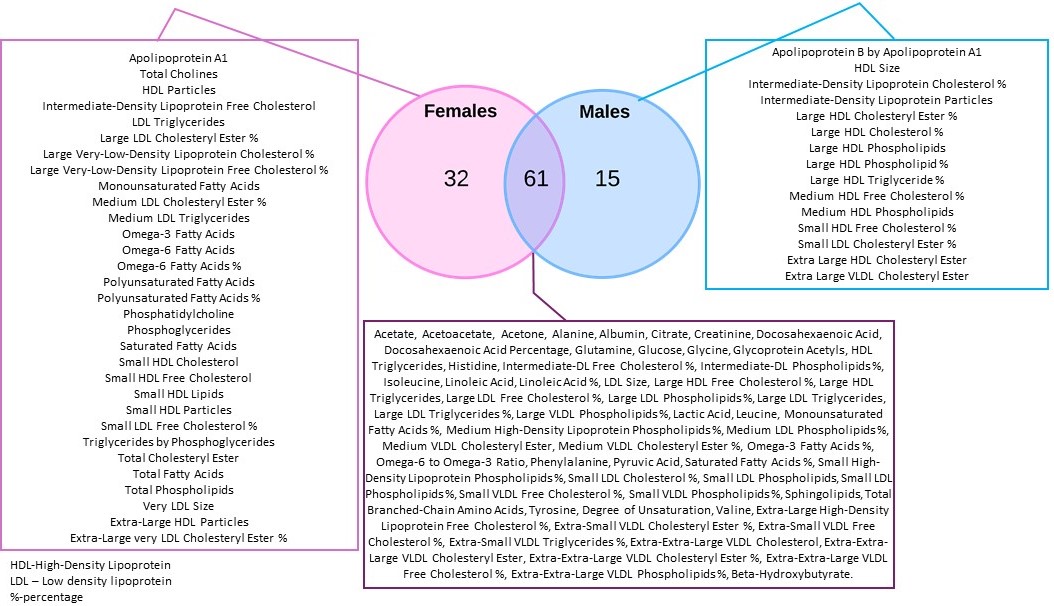

**Supplementary Figure 2:** An overview of the metabolites selected by the Boruta algorithm. 61 metabolites are common between males and females, while 32 and 15 are uniquely selected in females and males respectively.

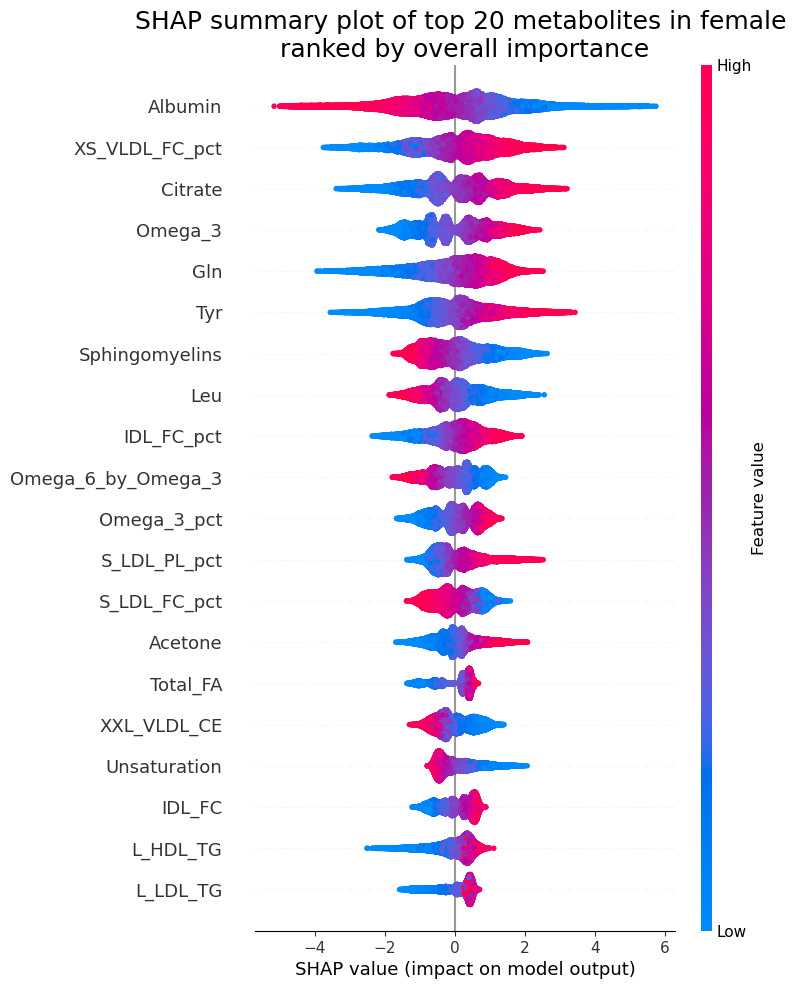

**Supplementary Figure 3.** SHAP values of the top 20 selected metabolites in females. Each dot denotes a participant, color of the dots denotes the metabolome expression level and X-axis denotes its contribution to the model decision.

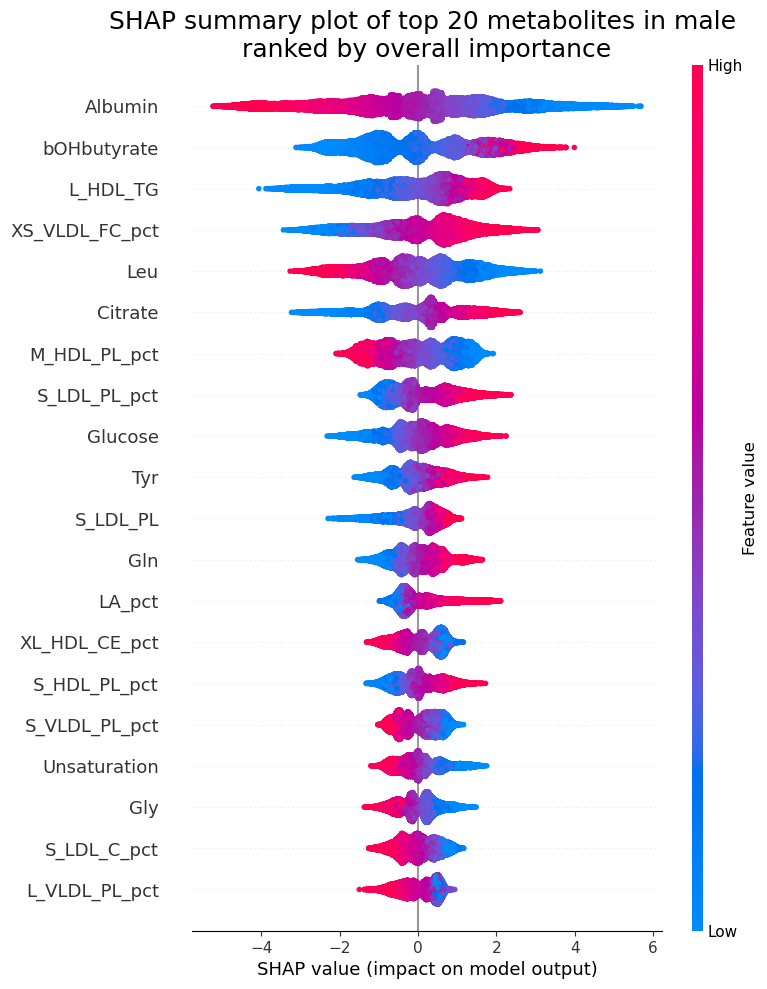

**Supplementary Figure 4:** SHAP values of the top 20 selected metabolites in males. Each dot denotes a participant, color of the dots denotes the metabolome expression level and X-axis denotes its contribution to the model decision.

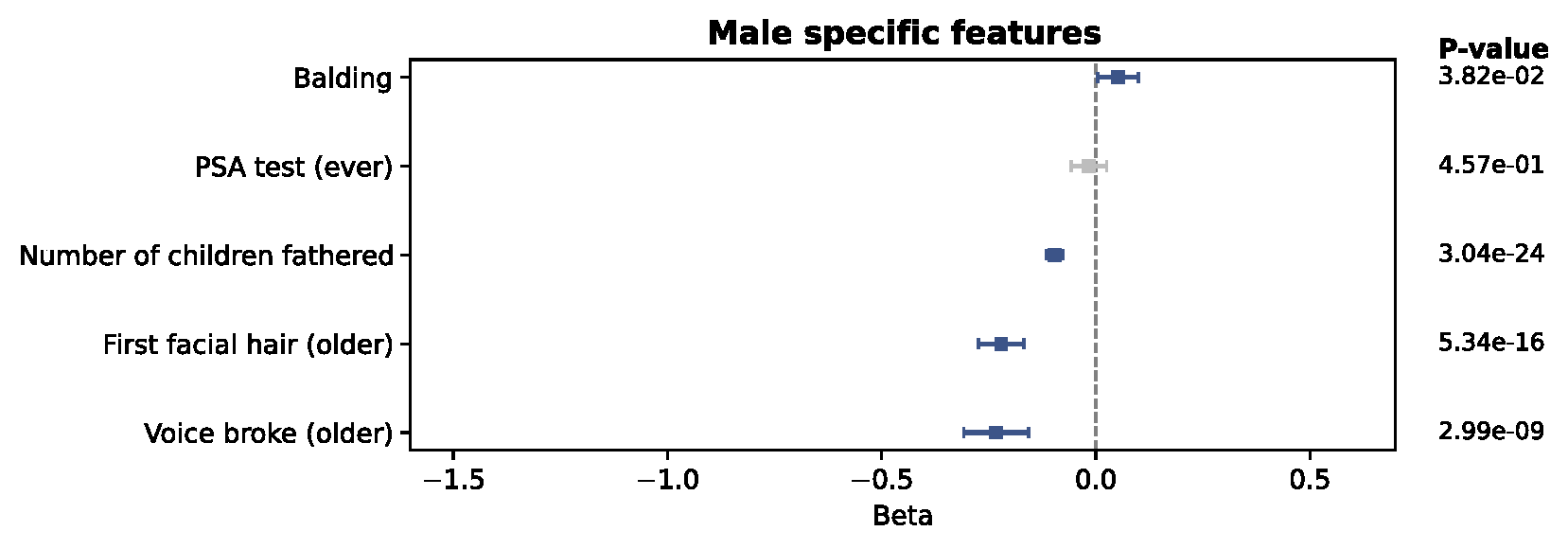

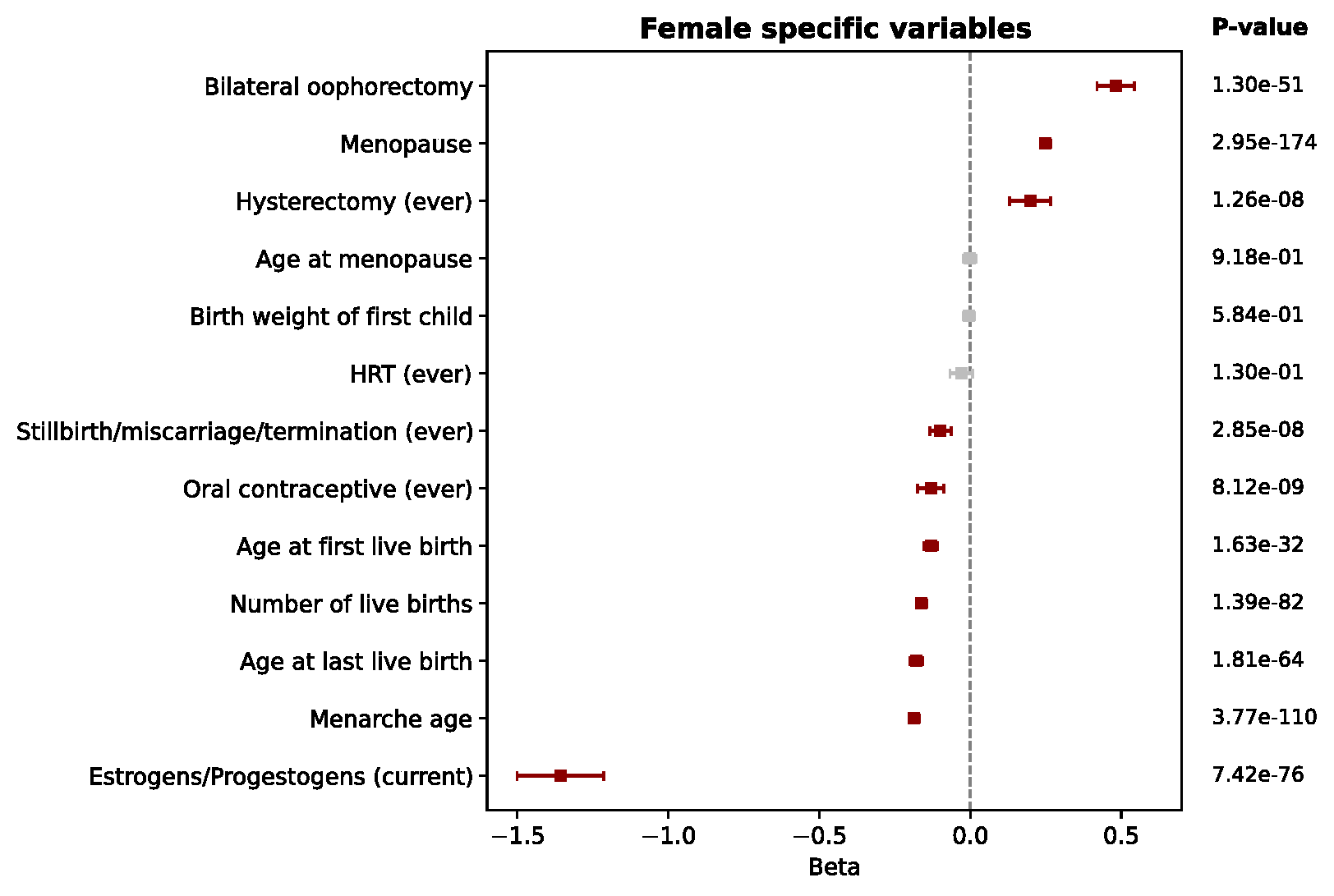

**Supplementary Figure 5:** Association of puberty and reproductive factors with metAgeGap**.** Linear regressions were performed between each exposure and metAgeGap adjusting for recruitment centre, ethnicity, education years, and Townsend deprivation index. Values were standardized if quantitative. Grey color showed if the association was not significant after FDR correction. Balding in males and menopause in females is associated with higher metAgeGap.

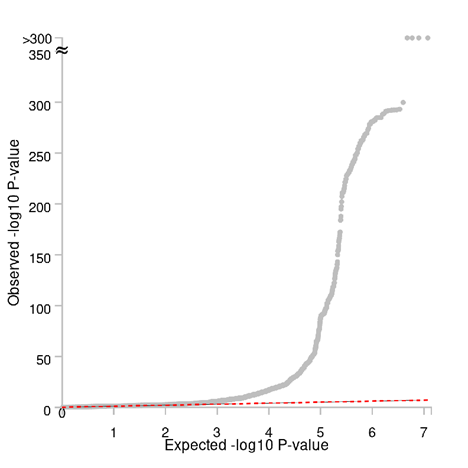

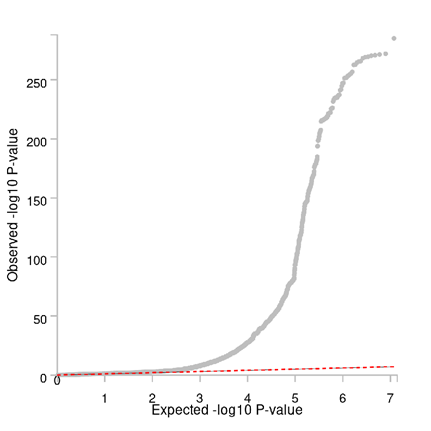

**Supplementary Figure 6**: Quantile-Quantile (QQ) plot for females (left) and males (right).

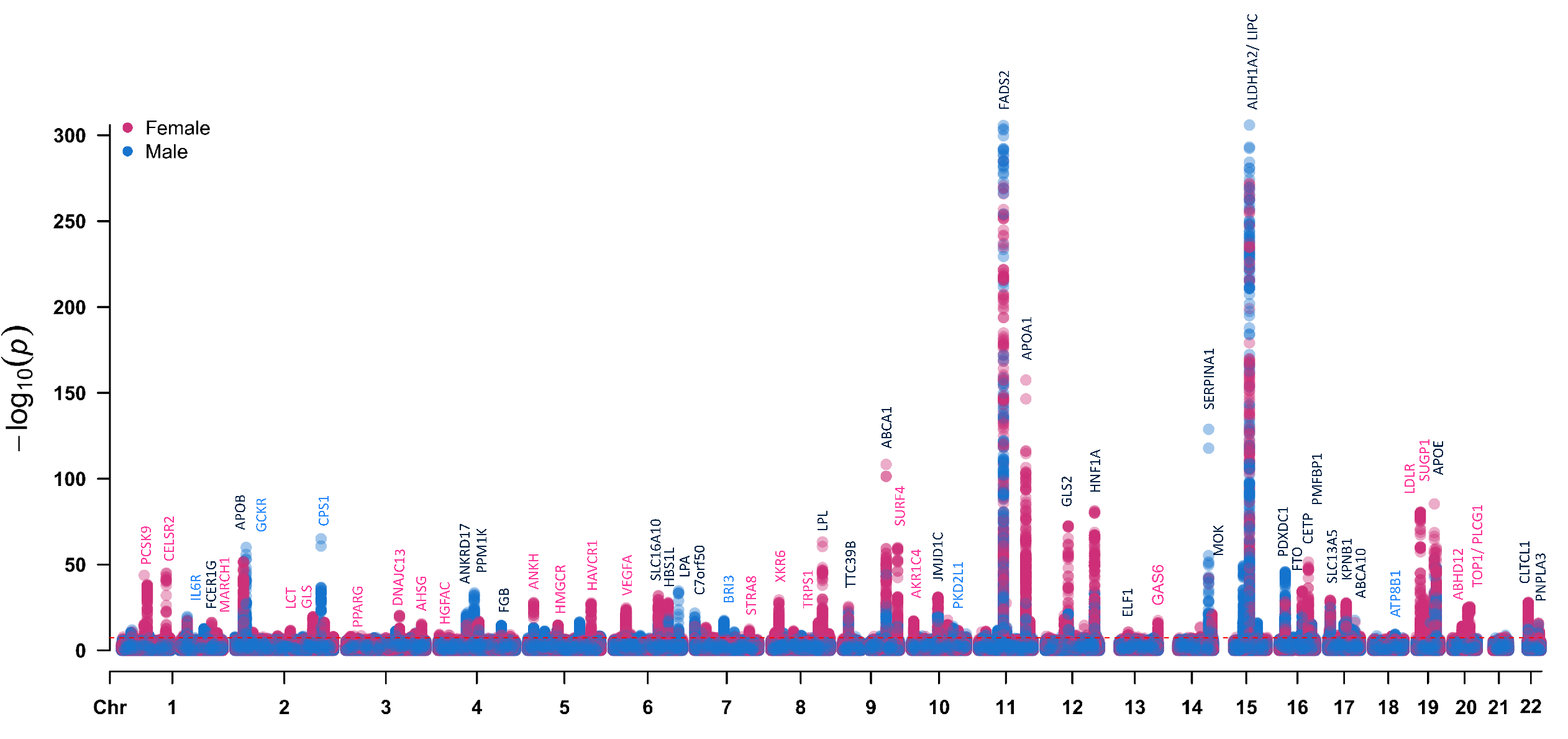

**Supplementary Figure 7**: Genome-wide association plot showing top genes associated with metAgeGap for both the sexes. Black genes mark loci that were significantly associated with metAgeGap in both males and females. Pink genes mark loci that were significant only in females and blue genes mark loci that were significant in males only.

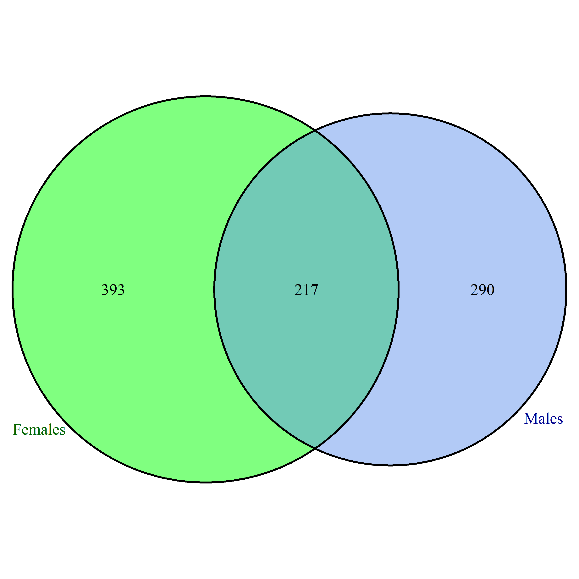

AARS, ABCA8, ABCA9, AC011530.4, AC018512.1, AC074091.13, AC092291.2, AC109829.1, AC110084.1, AC126614.1, ACP2, ADAL, ADAMTS4, AHNAK, AL590714.1, AL591806.1, ANKRD2, APOH, APOL3, ARHGAP30, ARHGAP9, ARHGEF25, ASB2, ATMIN, ATP1B2, ATP8B1, B4GALNT1, B4GALT3, BAIAP2L1, BAZ1B, BCAR1, BCL7B, BFAR, BLOC1S2, BRE, BRI3, C16orf46, C1orf116, C2orf16, C5orf51, CAPN3, CASC4, CATSPER2, CCDC121, CCNB2, CDAN1, CGREF1, CHRNB1, CHUK, CKMT1A, CKMT1B, CLEC18C, COBLL1, COL3A1, CPN1, CPS1, CTD-2132N18.3, CTDSPL2, CTRB1, CTRB2, CYP26A1, DCTN2, DDB2, DDIT3, DDX19A, DDX19B, DDX24, DHX58, DMPK, DMWD, DNAH2, DNAJC5G, DNMBP, DPAGT1, DPYSL5, DTX3, EIF2B4, EIF3J, EIF4A1, ELL3, EPB42, ERLIN1, EXOSC6, F12, FAM107B, FBXO4, FBXO46, FCHO2, FGG, FNDC4, FRMD4B, FRMD5, FUK, FXR2, FZD9, G6PC2, GANC, GCKR, GCSH, GIPR, GLI1, GPN1, GRK6, GSC2, GTF2E2, GTF3C2, H2AFX, HIF1AN, HKDC1, HMBS, HMG20A, HPN, HSPB9, HYOU1, HYPK, IFI27, IFI27L1, IFT172, IL6R, INHBC, INHBE, INPP5A, IQGAP2, IRF1, ITLN2, KAT2A, KCNH4, KHK, KIF5A, KLHDC9, KRTCAP3, LCMT2, LDHAL6B, LGR6, LRAT, LY96, MAD1L1, MADD, MAP1A, MAPRE3, MARS, MBD6, MEPE, MFAP1, MIER1, MPV17, MRC1L1, MRPL33, MYBPC3, MYEOV, MYO1E, NDUFA4L2, NDUFB8, NIT1, NLRC5, NOMO1, NPIPA1, NPIPA2, NPIPA5, NR1H3, NRBP1, OBFC1, OS9, OTUB2, OXCT1, PDIA3, PDPR, PEAK1, PEPD, PFDN2, PFKFB2, PFN3, PKD2, PKD2L1, PLA2G10, PLA2G4F, PLG, PPIP5K1, PPM1G, PPOX, PPP1R32, PRKCA, PVRL4, QPCTL, R3HDM2, RAB5C, RASGEF1C, RBKS, RBPMS, RNF130, RNFT1, RORA, RP11-113D6.10, RP11-123K3.4, RP11-178C3.1, RP11-286N22.8, RP11-296A16.1, RP11-432B6.3, RP11-529K1.3, RP11-544M22.13, RPS6KB1, RSPH6A, RUNX3, SAA2, SAA4, SAT2, SBF2, SCARB1, SCD, SCGB1A1, SCN1B, SEC31B, SENP3, SERF2, SERINC4, SERPINA11, SERPINA12, SERPINA6, SERPINA9, SERPINF2, SF3B3, SH3YL1, SHBG, SLC12A2, SLC22A2, SLC25A47, SLC26A10, SLC30A3, SLC35D1, SLC4A1AP, SLTM, SNCAIP, SNRPD2, SNX17, SPC25, SPG11, SPI1, ST3GAL2, STAC3, STAG1, STARD9, STRC, SUPT7L, SUPV3L1, TBL2, TCEB1, TCF23, TECPR1, TGM5, TGM7, TMEM171, TMEM62, TMEM70, TMPRSS11E, TNFSF12, TNFSF12-TNFSF13, TNFSF13, TP53, TP53BP1, TRIM54, TSPAN3, TSTD1, TTBK2, TUBGCP4, UBE2F, UBE2F-SCLY, UBE2T, UBR1, UCN, UFC1, UGT2B17, UNC119B, USF1, USP21, VMP1, VPS11, VPS39, WARS, WDR25, WDR76, WDR78, WDR81, WNT8B, YOD1, ZBTB4, ZMYND8, ZNF106, ZNF512, ZNF513, ZSCAN29

ABCG5, ABCG8, ABHD12, AC010547.9, AC011475.1, AC022431.2, AC083862.1, AC140061.12, ACAD11, ACKR4, ACMSD, ACOT8, ACPP, ADAMTS13, ADH6, AFM, AGBL3, AHSG, AKR1C3, AKR1C4, AKR1CL1, AL035252.1, AL121963.1, AMDHD1, ANGPTL3, ANKDD1B, ANKRD52, AP1G1, AP1M2, AP3B2, APOC2, APOF, ARID1A, ARMC6, ASAP3, ATF1, ATG4C, ATG4D, ATP13A1, ATP5B, ATXN1L, AZGP1, BACE1, BCAM, BCAS3, BCL3, BCS1L, BLMH, BLOC1S3, BSND, C15orf27, C19orf38, C19orf52, C19orf80, C1orf172, C1orf192, C1QBP, C1QTNF4, C20orf173, C2orf43, C7orf43, C7orf49, C9orf96, CACFD1, CALB2, CALD1, CAMTA2, CARM1, CBLC, CCDC155, CCNT2, CDKAL1, CDKN2D, CDPF1, CEACAM16, CEACAM19, CELSR1, CELSR2, CEP164, CEP250, CILP2, CINP, CLASRP, CLDN11, CMTR2, CNPY2, COL10A1, COL4A3BP, COQ10A, COX18, CPD, CPEB1, CPNE4, CPSF1, CPT1A, CS, CTB-129P6.11, CTC-260F20.3, CTDSP1, CTSA, CYP27A1, DARS, DCPS, DDX56, DERL2, DIP2B, DNAH11, DNAJB11, DNAJC13, DNAJC14, DNM2, DNTTIP1, DOCK6, DOCK7, DOK7, DYNC1H1, EIF5A2, ELMSAN1, EMILIN3, ENTPD6, ERGIC3, ETFA, EXOC3L2, FAM105A, FAM105B, FAM180B, FAM182B, FAM186A, FAM25A, FAM46B, FAM46C, FARP2, FBXO22, FCGR2B, FCGR3A, FCGR3B, FCRLA, FETUB, FOXA3, GAL3ST4, GAS6, GATAD2A, GDF5, GDNF, GINS1, GLS, GLTPD2, GMIP, GOSR1, GP1BA, GPC2, GTSE1, HAL, HAPLN4, HAVCR1, HAVCR2, HGFAC, HGS, HK3, HMGCR, HNF4A, HSPA6, HYDIN, IFT80, IGHMBP2, IL23A, ILF3, IRF2BP1, ISG20L2, ISL2, IST1, KANK2, KBTBD4, KIAA1324, KMT2A, KRI1, LAMTOR4, LARP4, LCT, LDHA, LDHAL6A, LDHC, LDLR, LIMA1, LITAF, LMAN2, LPAR2, LPIN3, MAR1, MAP3K19, MAP4K5, MARK4, MARVELD3, MAU2, MCM6, MED22, MEF2B, MEF2BNB, MEF2BNB-MEF2B, MFHAS1, MINK1, MIS12, MMP19, MMP9, MRPL12, MRPL21, MTCH2, MYBPHL, MYPOP, N4BP2L2, NAB1, NABP2, NACA, NANP, NAT2, NCAN, NCOR2, NDUFA13, NDUFS3, NEURL2, NFAT5, NINL, NKPD1, NPHP3, NQO1, NRG4, NUCB1, NUDC, NUP160, NUP88, OBP2B, P2RX7, PACSIN2, PAN2, PARVB, PBX4, PCIF1, PCSK9, PHC1, PHLPP2, PINX1, PKDREJ, PLCD4, PLCG1, PLTP, POLK, POLR2A, PPARA, PPARG, PPP1R15A, PPP1R37, PPP6R3, PRMT6, PROX1, PSMB6, PSRC1, PTGES3, PTK2, PTPMT1, PTPN9, PUM2, PYGB, QTRT1, R3HDM1, R3HDML, RAB3GAP1, RABEP1, RALGDS, RASSF6, RCN2, RELB, REXO4, RFXANK, RGS12, RHCE, RIN2, RNF25, RNF41, RORC, RP11-762I7.5, RP11-977G19.10, RP1L1, RPAIN, RPL22L1, RPL7A, RQCD1, S1PR5, SARNP, SARS, SCAPER, SCIMP, SDC1, SDHC, SFN, SGCD, SLC11A1, SLC11A2, SLC12A5, SLC25A10, SLC2A2, SLC2A6, SLC38A4, SLC39A4, SLC39A5, SLC44A2, SLC6A16, SLC7A2, SMARCA4, SMC4, SNX21, SOX7, SP4, SPAG7, SPATA25, SPC24, SPINT4, SPRN, SPTY2D1, ST3GAL4, STAG3, STAT1, STAT2, STK25, STK36, STRA8, SUGP1, SUGP2, SURF1, SURF2, SURF4, SURF6, SYCE1, TAOK1, TAT, TCEA3, TCF7L2, TIMD4, TIMELESS, TM4SF5, TM6SF2, TMED1, TMED4, TMEM140, TMEM163, TMEM255B, TMEM57, TMEM61, TNNC2, TOP1, TRAPPC6A, TRIB1, TRMU, TSG101, TSKS, TSSK6, TTC38, TTLL4, TULP2, TULP3, UBA5, UBE2C, UBE2Q2, UBXN4, UIMC1, USP1, USP24, USP37, VAC14, VEGFA, VIL1, VMO1, VPS28, WBP4, WDR91, WFDC3, XKR6, YIPF2, YJEFN3, ZDHHC18, ZFHX3, ZFP3, ZHX3, ZNF101, ZNF142, ZNF23, ZNF335, ZNF337, ZNF346, ZNF821, ZNF839, ZRANB3, ZSWIM1, ZSWIM3

ABCA1, ABCA10, ABCA5, ABCA6, AC079602.1, ACADS, ADAM10, ADAP1, ADCY5, ADH4, ADH5, ADM5, AFP, AGBL2, ALB, ALDH16A1, ALDH1A2, ANKH, ANKRD17, AP003733.1, APOA1, APOA2, APOA4, APOA5, APOB, APOC1, APOC3, APOC4, APOC4-APOC2, APOE, AQP9, BCAT2, BCL2L12, BEST1, BMPR1A, BUD13, C10orf62, C12orf43, C16orf45, C5orf56, C7orf50, CA11, CABP1, CDK2, CELF1, CETP, CHST4, CLTCL1, COX19, CPT1C, CTD-3148I10.9, CYP2W1, DAGLA, DBP, DHODH, DHX38, EFCAB13, EIF4E, ELF1, ERBB3, ESYT1, F11R, FADS1, FADS2, FADS3, FAM35A, FAM63B, FAM83E, FBXO39, FCER1G, FCGR2A, FCGRT, FEN1, FGA, FGB, FGF21, FLT3LG, FNBP4, FRK, FTH1, FTO, FUT1, FUT2, GLS2, GLUD1, GPER1, GPR146, HBS1L, HERPUD1, HNF1A, HOGA1, HP, HPR, hsa-mir-150, HSD17B14, HSP90AA1, IKZF4, INCENP, IRF3, ITGB3, IZUMO1, JAZF1, JMJD1C, KIAA1919, KPNB1, LIPC, LPA, LPL, MAMSTR, MAP2K6, METAP1, MIP, MLEC, MLXIPL, MMRN2, MOK, MPV17L, MYB, MYH11, MYRF, NDUFS2, NOSIP, NPEPPS, NR1I3, NRBF2, NT5DC1, NTAN1, NTN5, NUDT1, OASL, PA2G4, PAFAH1B2, PARN, PCP4L1, PCSK7, PDXDC1, PI4K2A, PIH1D1, PLEKHA4, PLRG1, PMEL, PMFBP1, PNPLA3, PPM1K, PPP4R4, PRMT1, PRR12, PRRG2, PSMC3, PVRL2, RAB3IL1, RAB5B, RAPSN, RASIP1, RBMS2, RCN3, REEP3, REV3L, RGS14, RNF111, RNF214, RP11-1021N1.1, RP11-603J24.9, RPL13A, RPL18, RPL41, RPS11, RPS26, RRAS, RRN3, SAMM50, SCAF1, SCGB1D1, SCGB1D2, SCGB2A1, SERPINA1, SERPINA10, SH2D7, SIDT2, SIK3, SLC13A5, SLC16A10, SLC22A3, SLC25A1, SLC34A1, SLC39A13, SMARCC2, SNX13, SNX8, SPACA4, SPHK2, SPPL3, SPRYD4, SULT2B1, SYT7, TAGLN, TBC1D2B, TBKBP1, TDRD15, TEKT1, TMEM258, TNPO1, TOMM40, TOMM40L, TRAF3IP2, TRIM59, TRPS1, TTC39B, TUBD1, TXNL4B, UGT2B15, WDR20, XAF1, ZC3H10, ZFAND2A, ZNF19, ZNF259

**Supplementary Figure 8**: Genes that were found associated with metAgeGap in males and females in GWAS.

**Supplementary Figure 9:** Scatter plot of male and female genetic correlations identified in with DSC. Black is significant in both males and females, red and blue are traits significantly genetically correlated in females and males respectively.

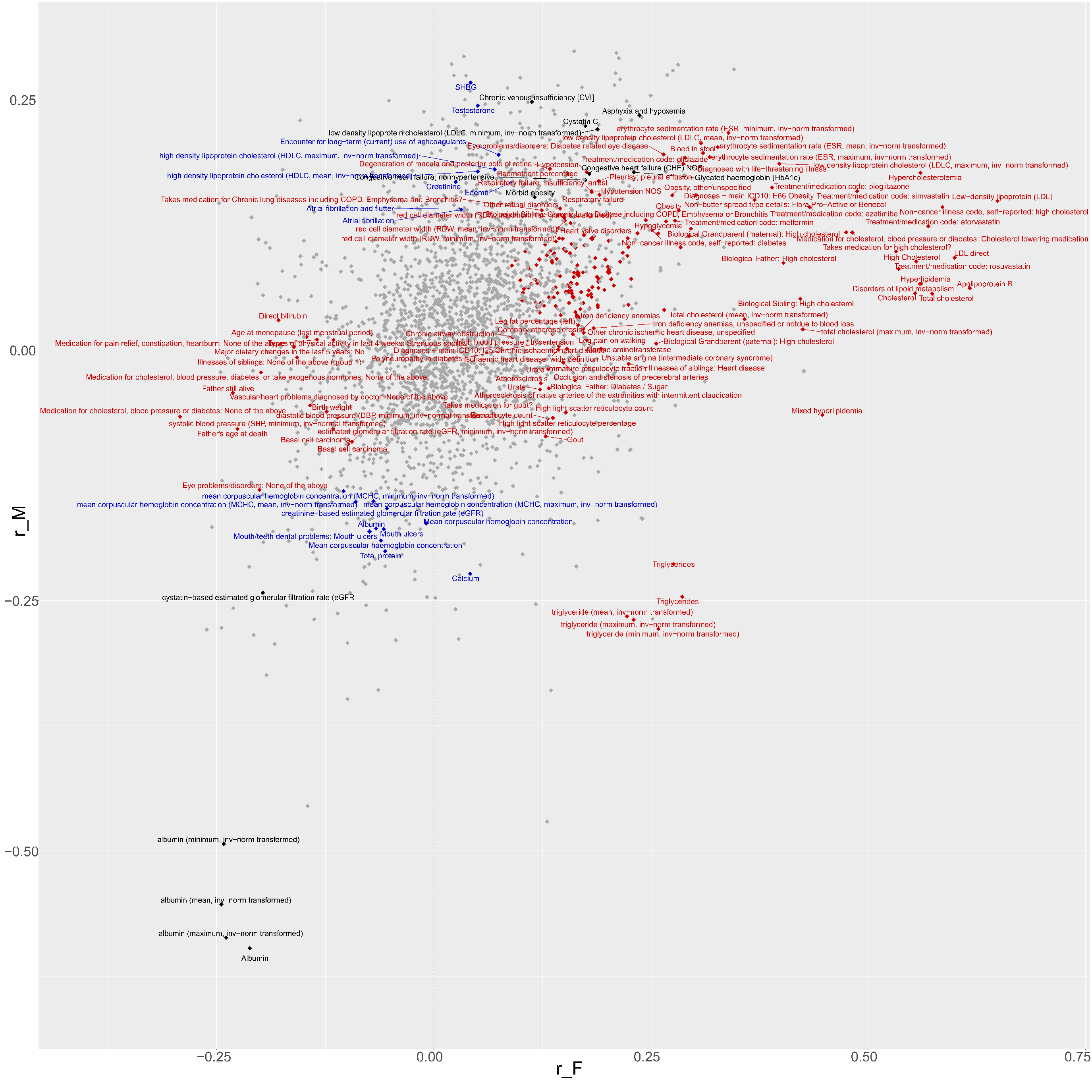

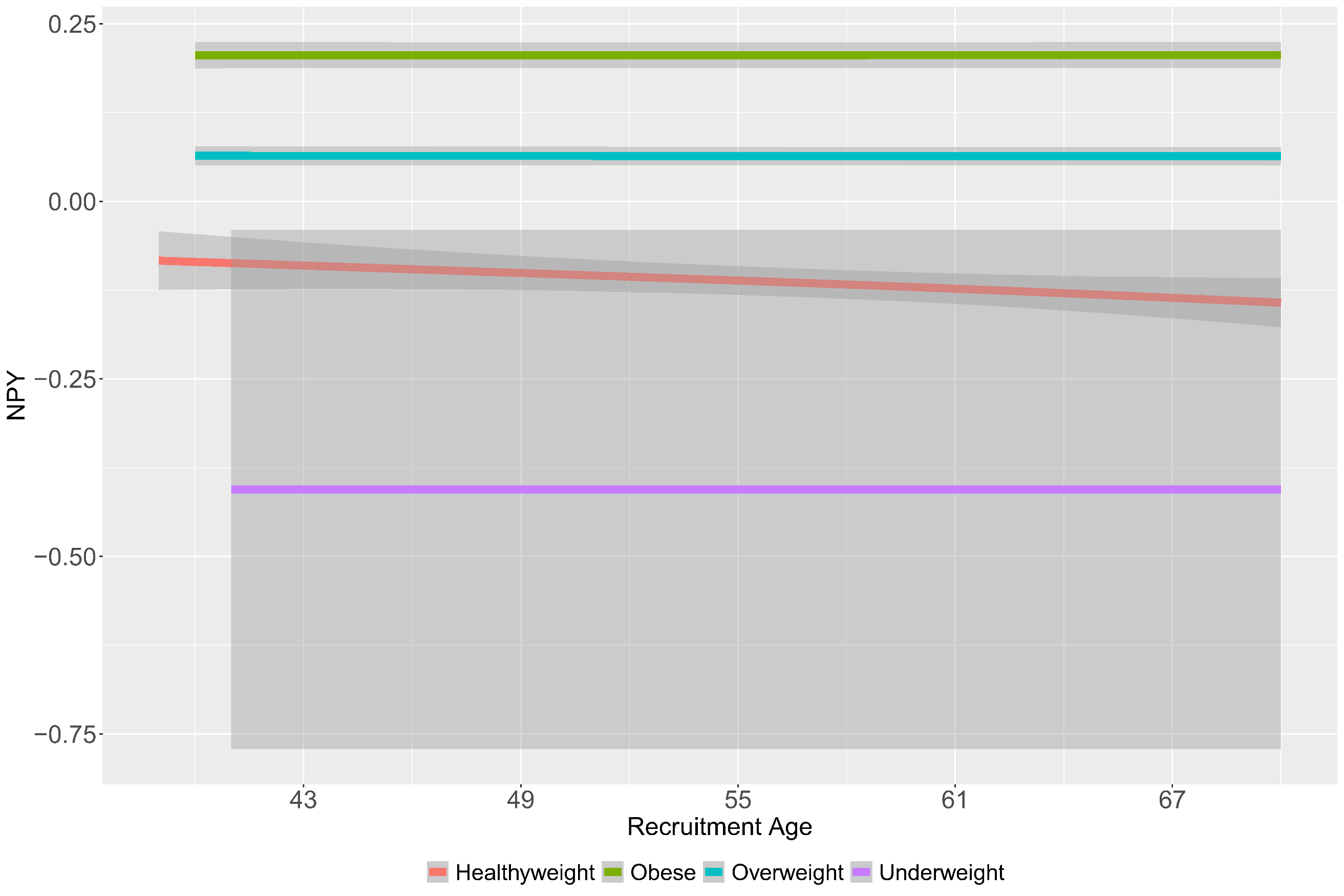

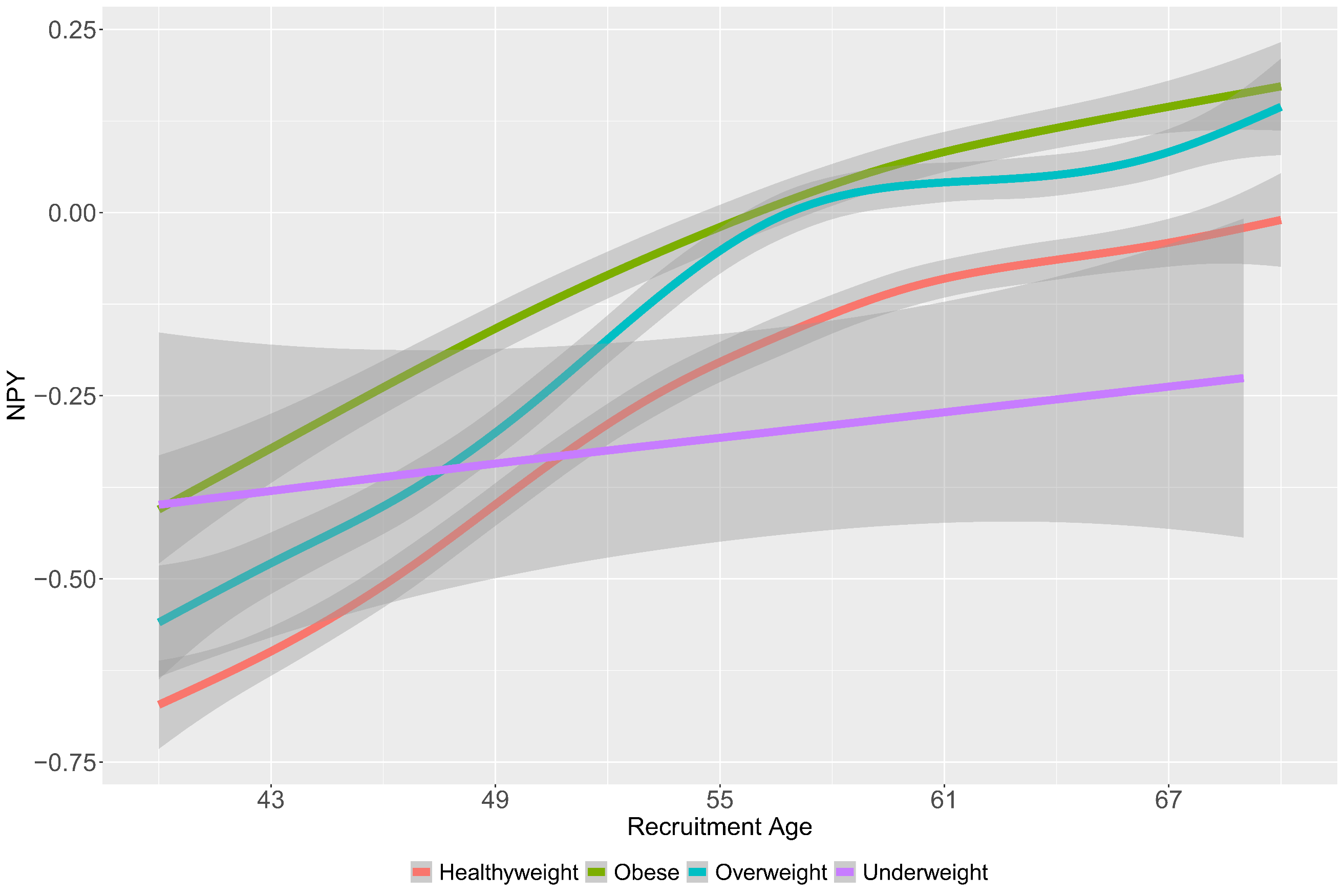

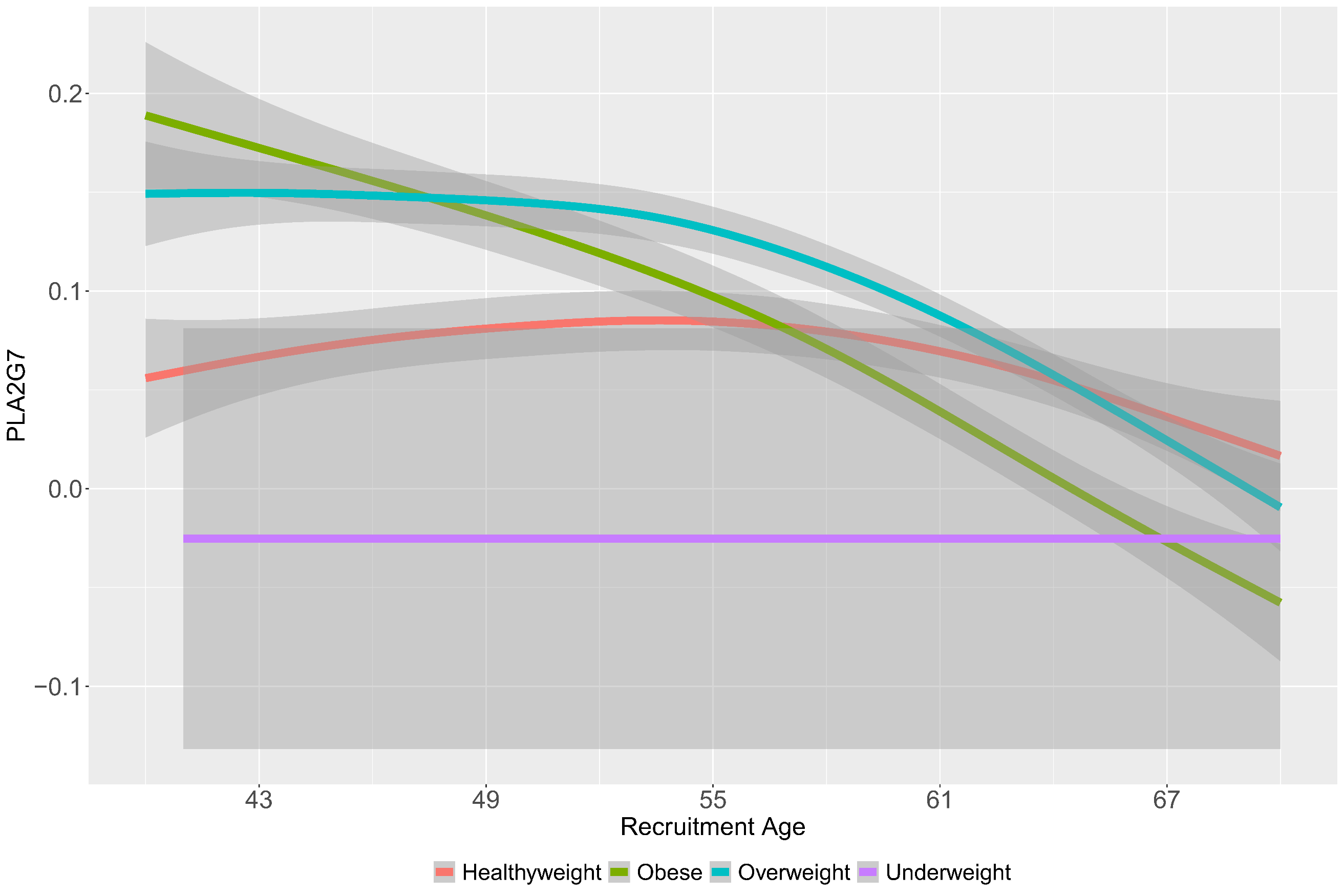

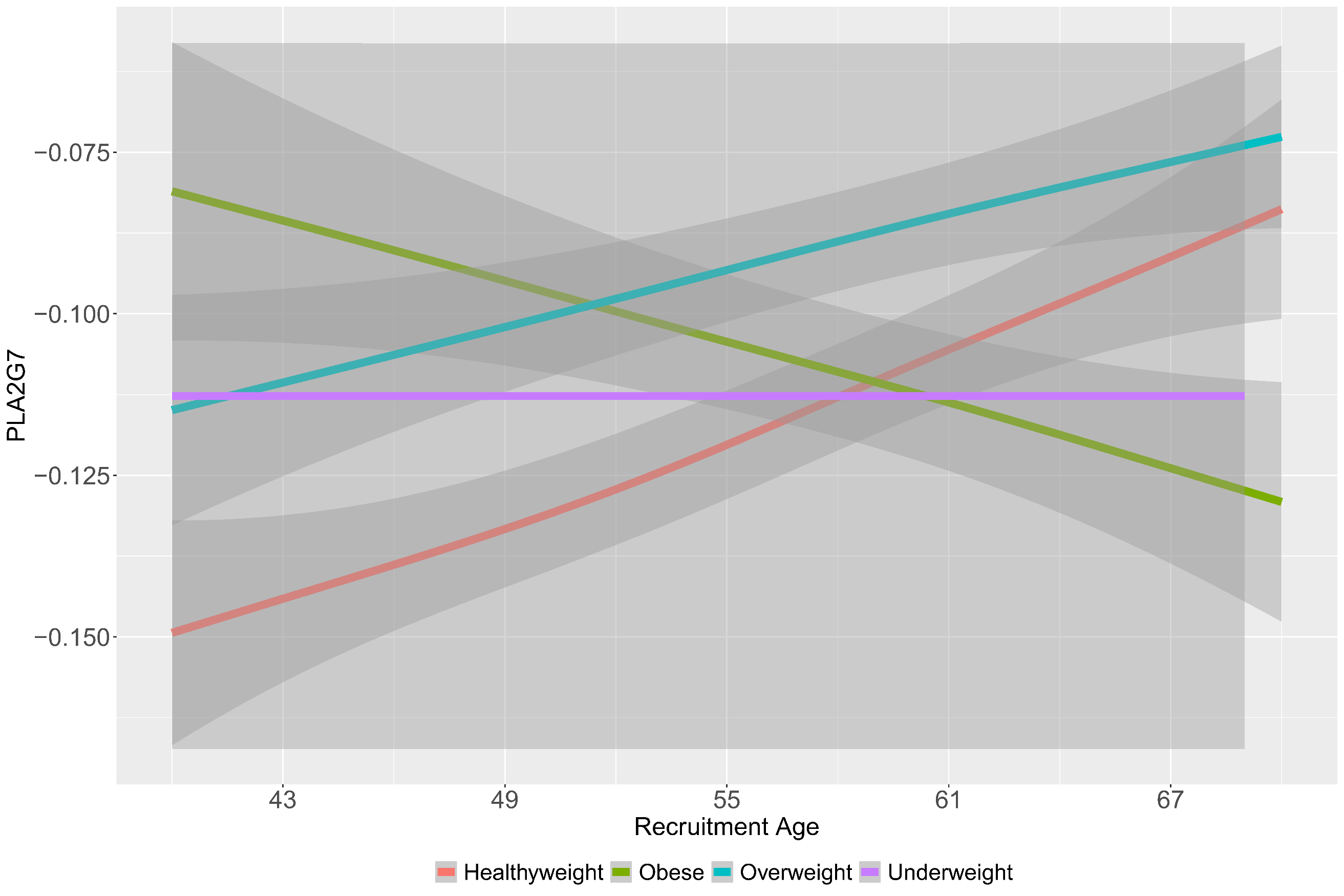

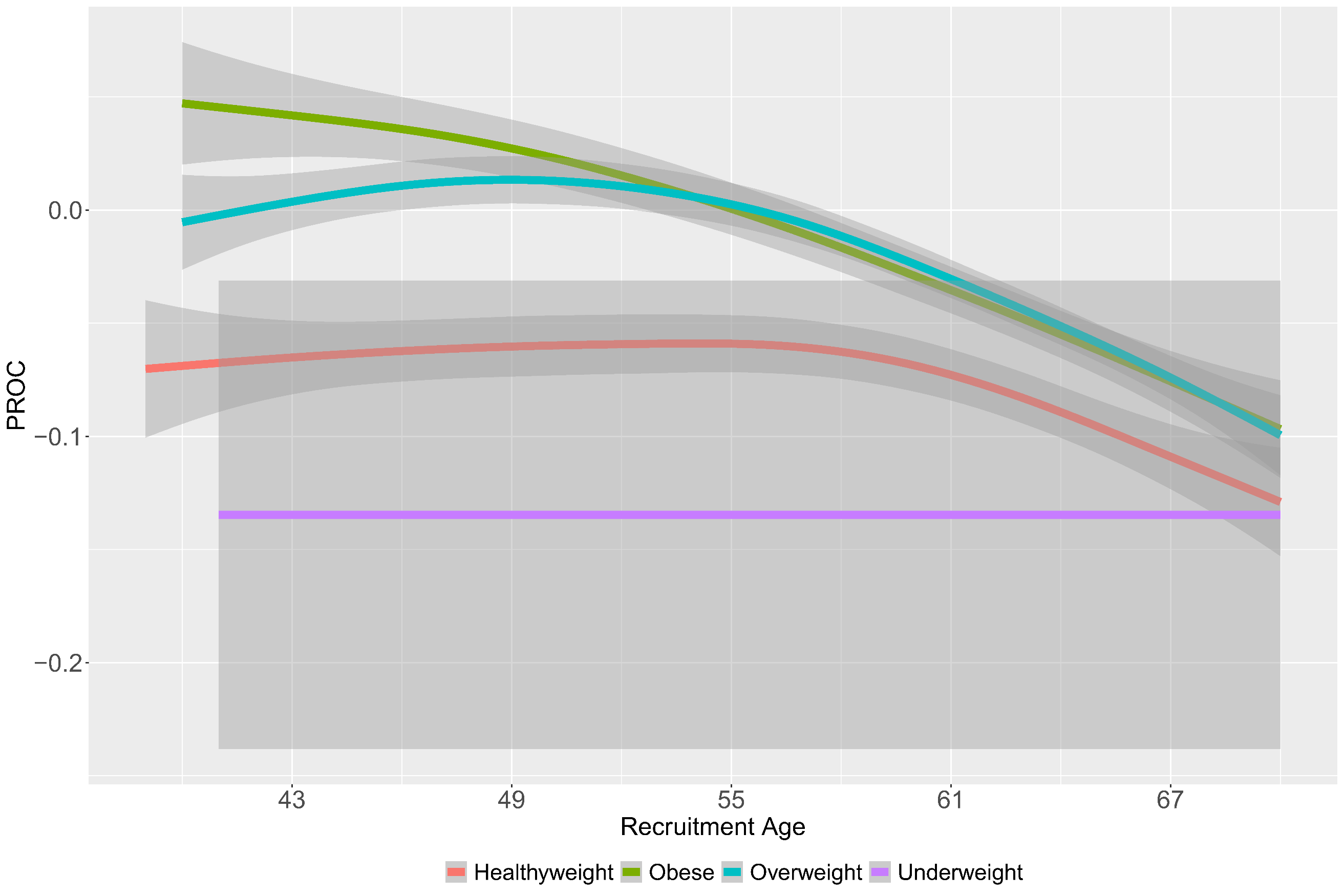

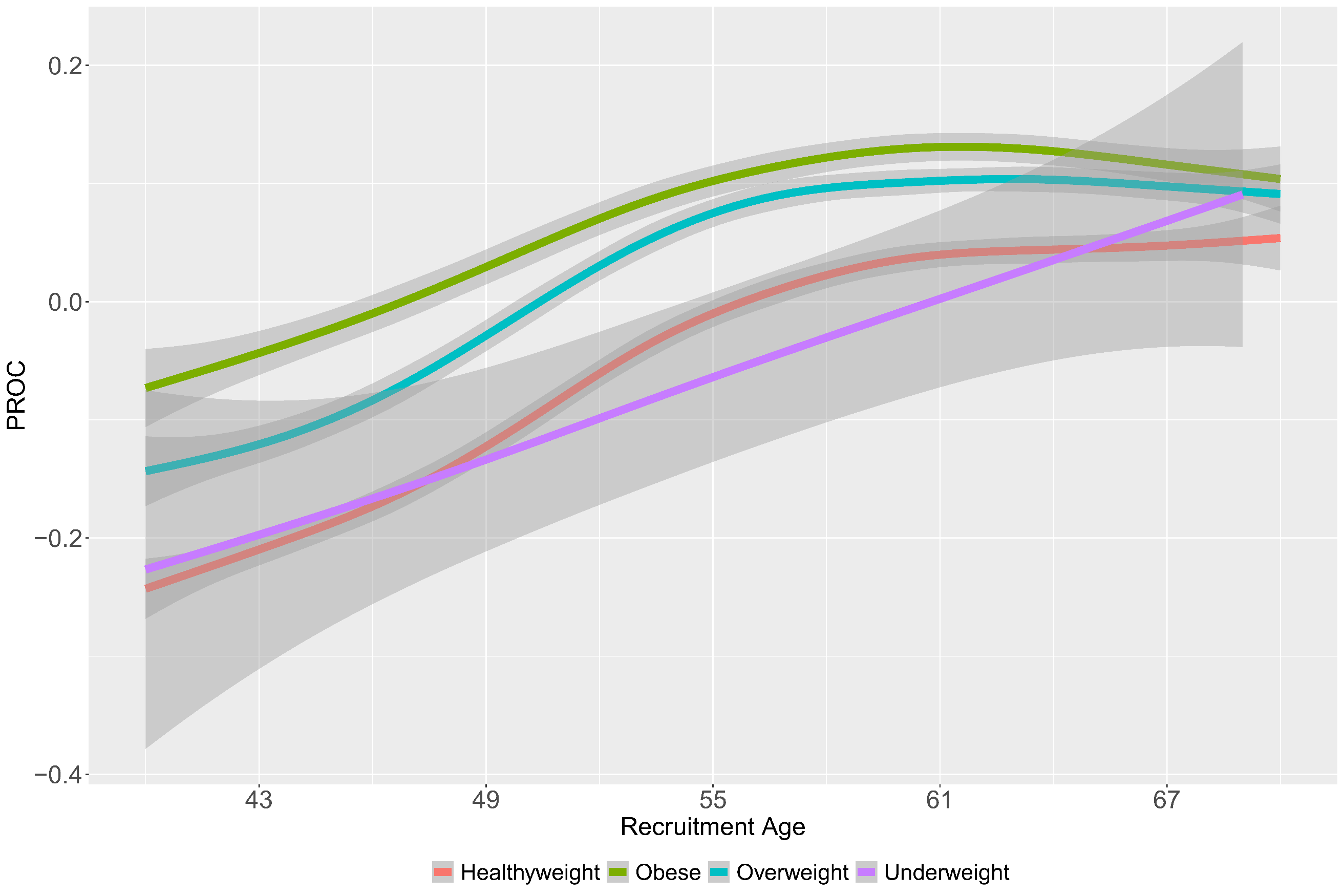

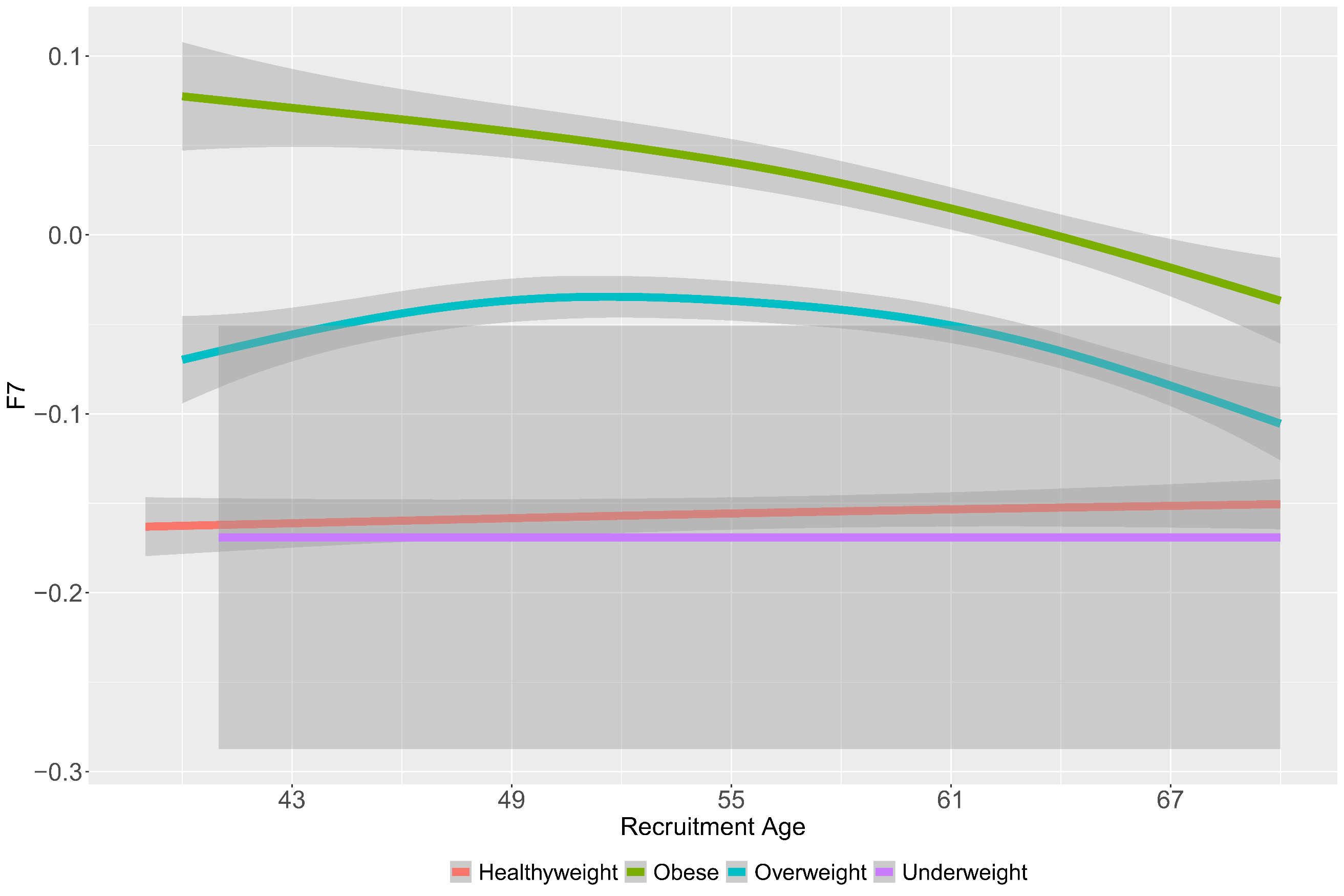

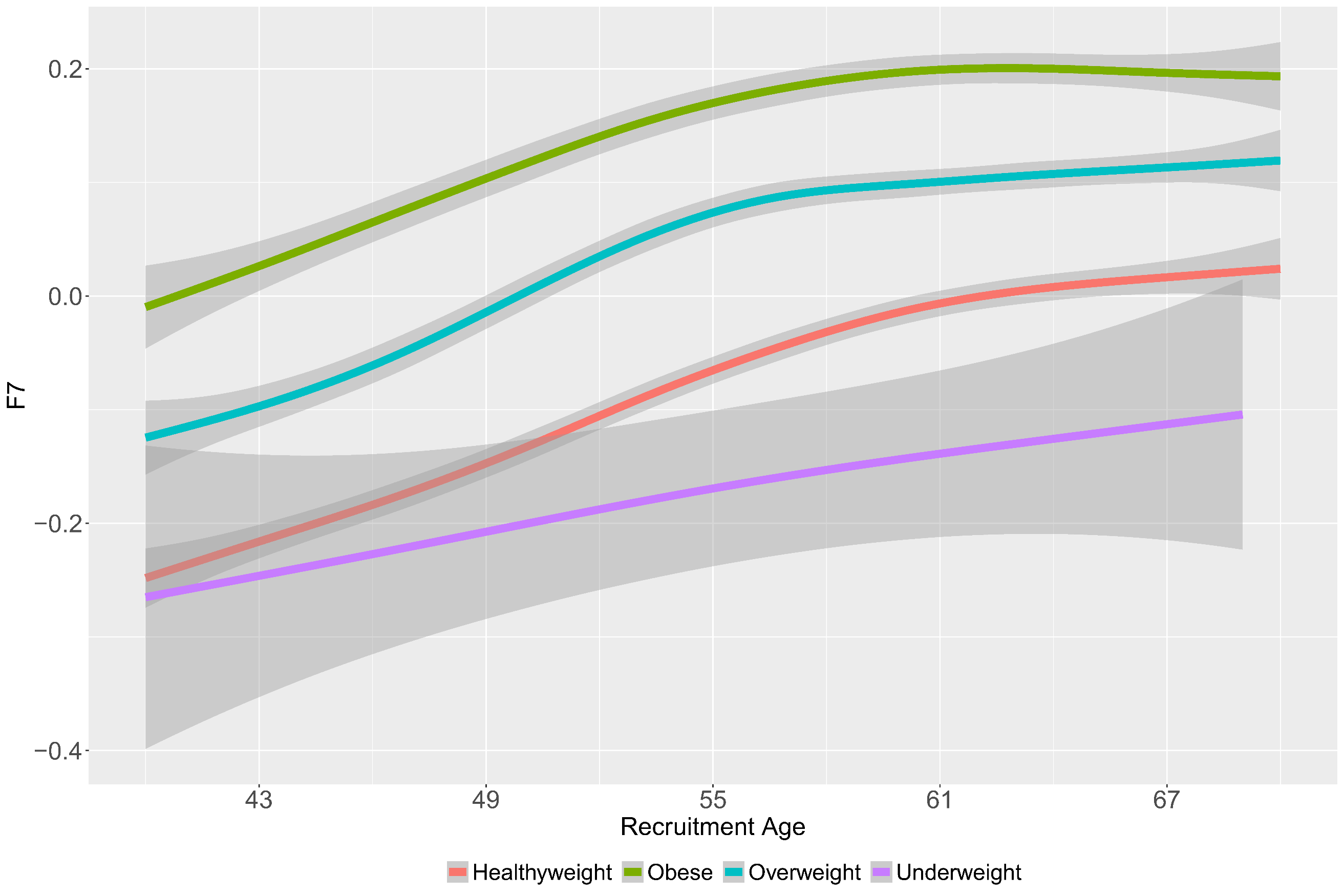

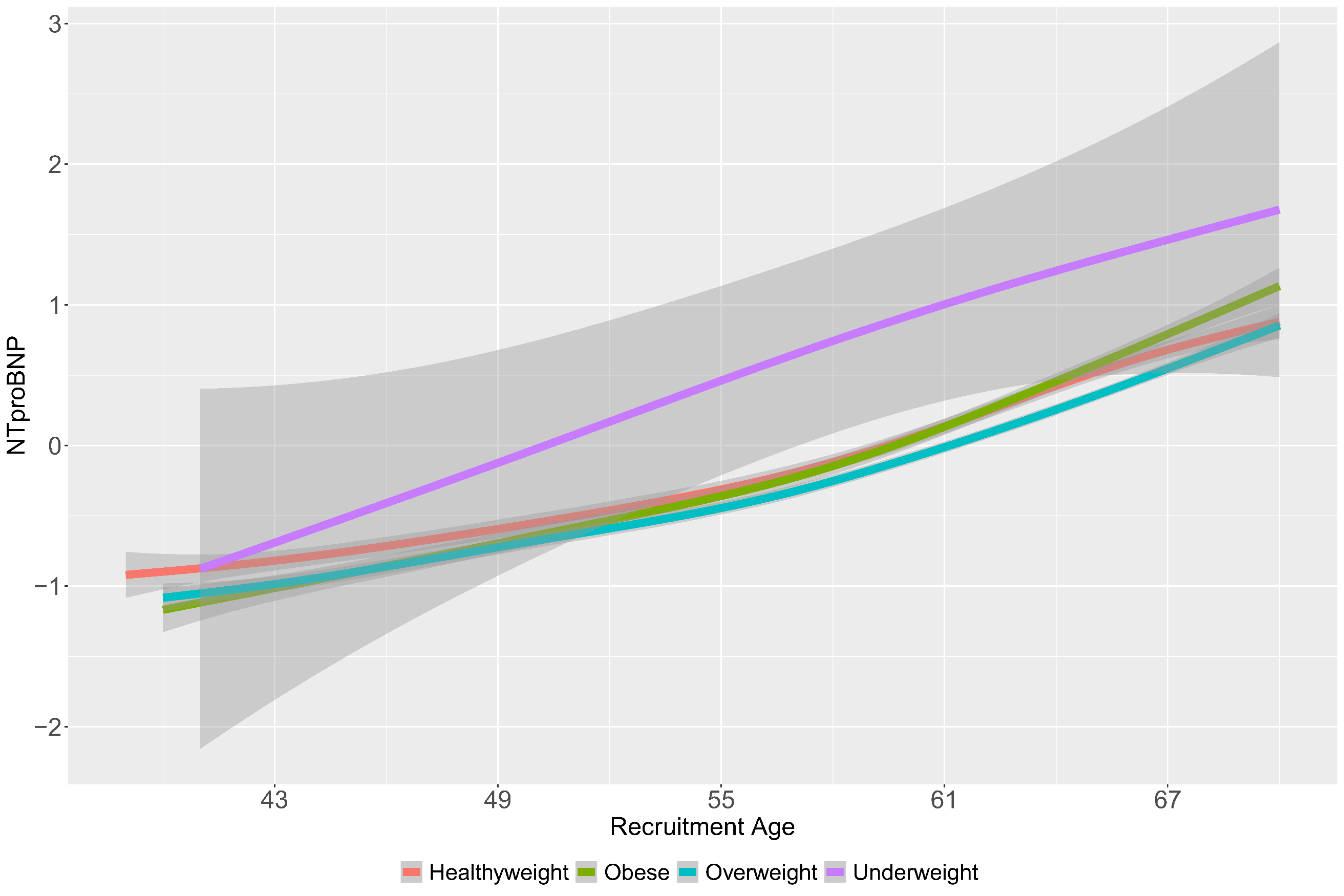

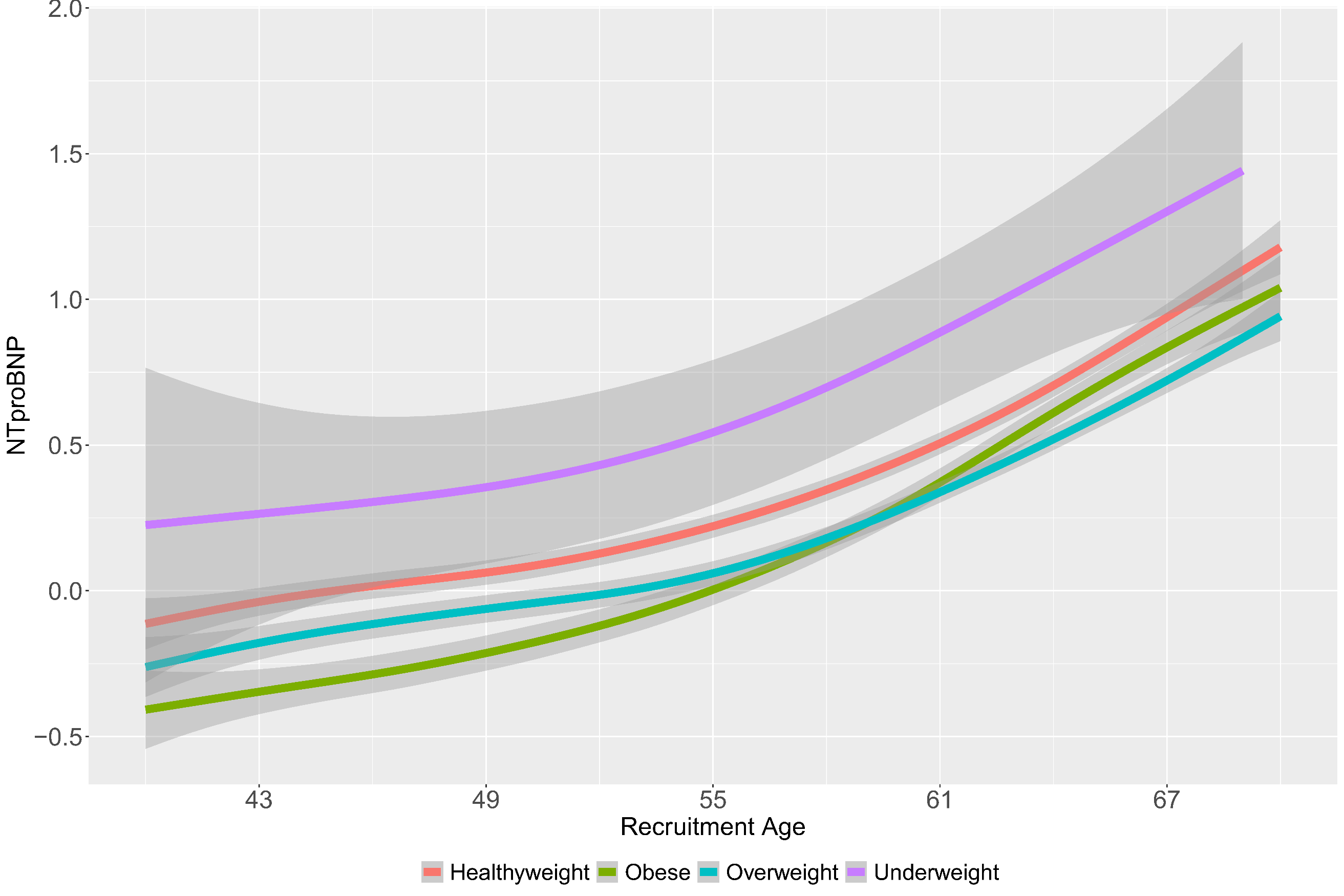

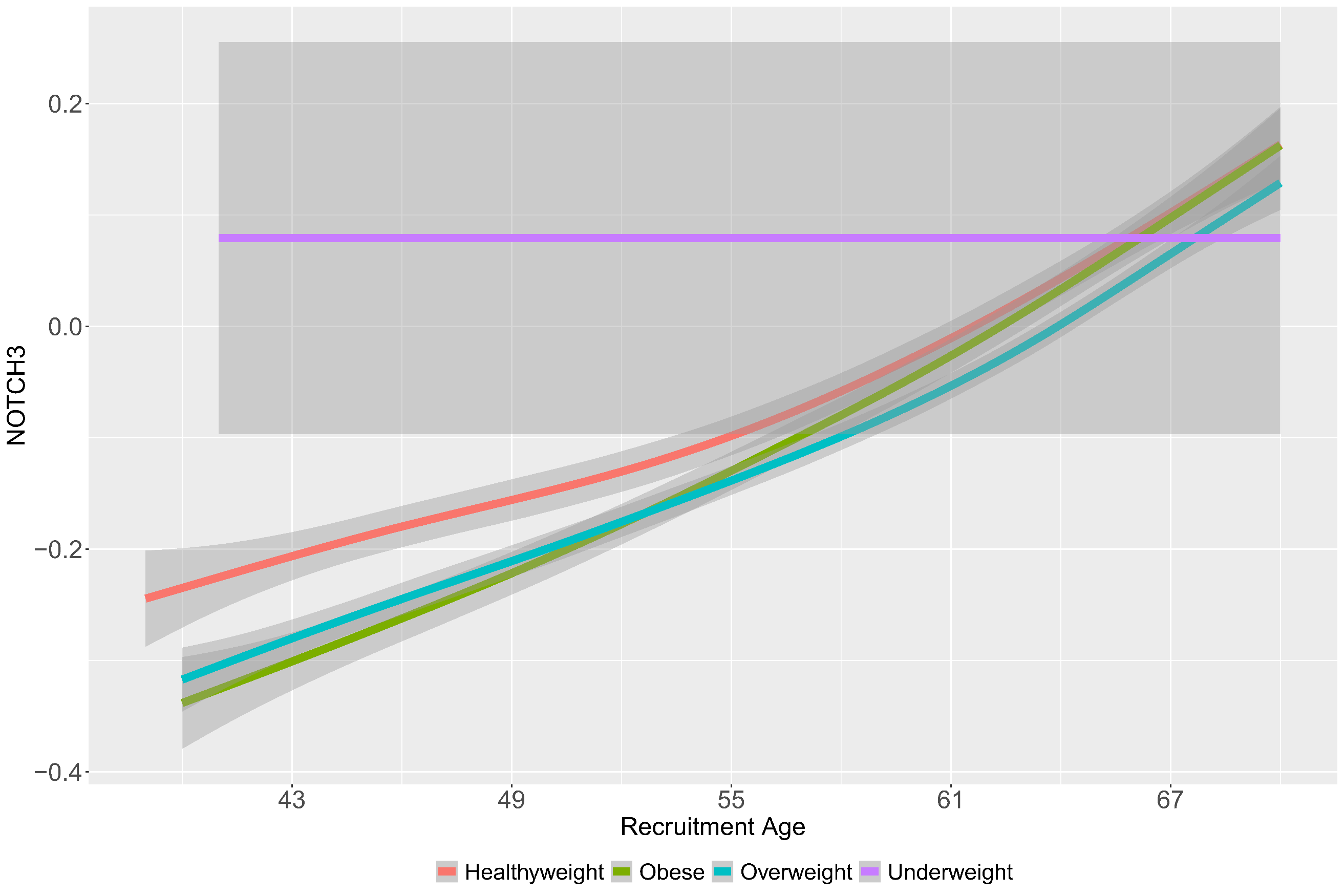

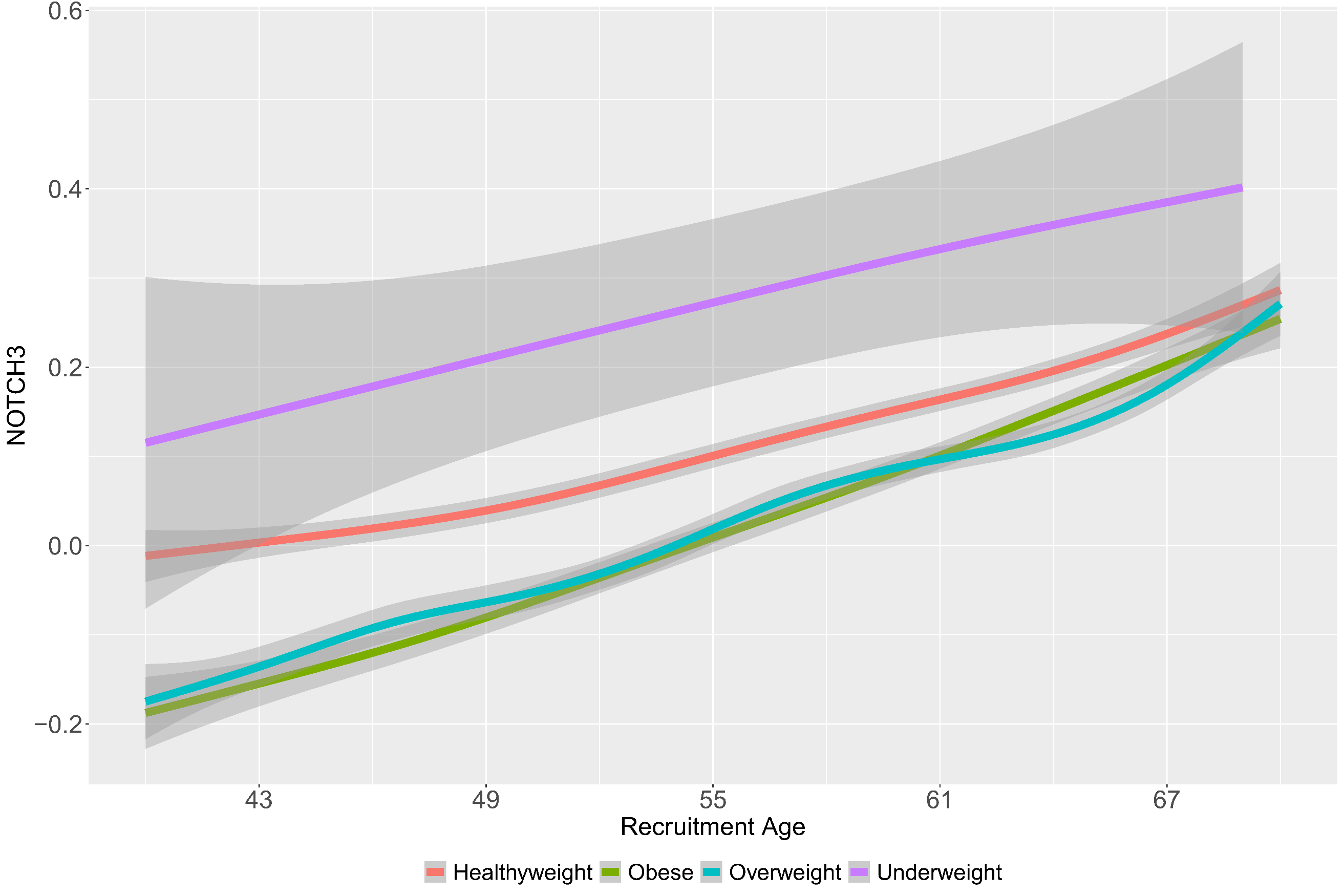

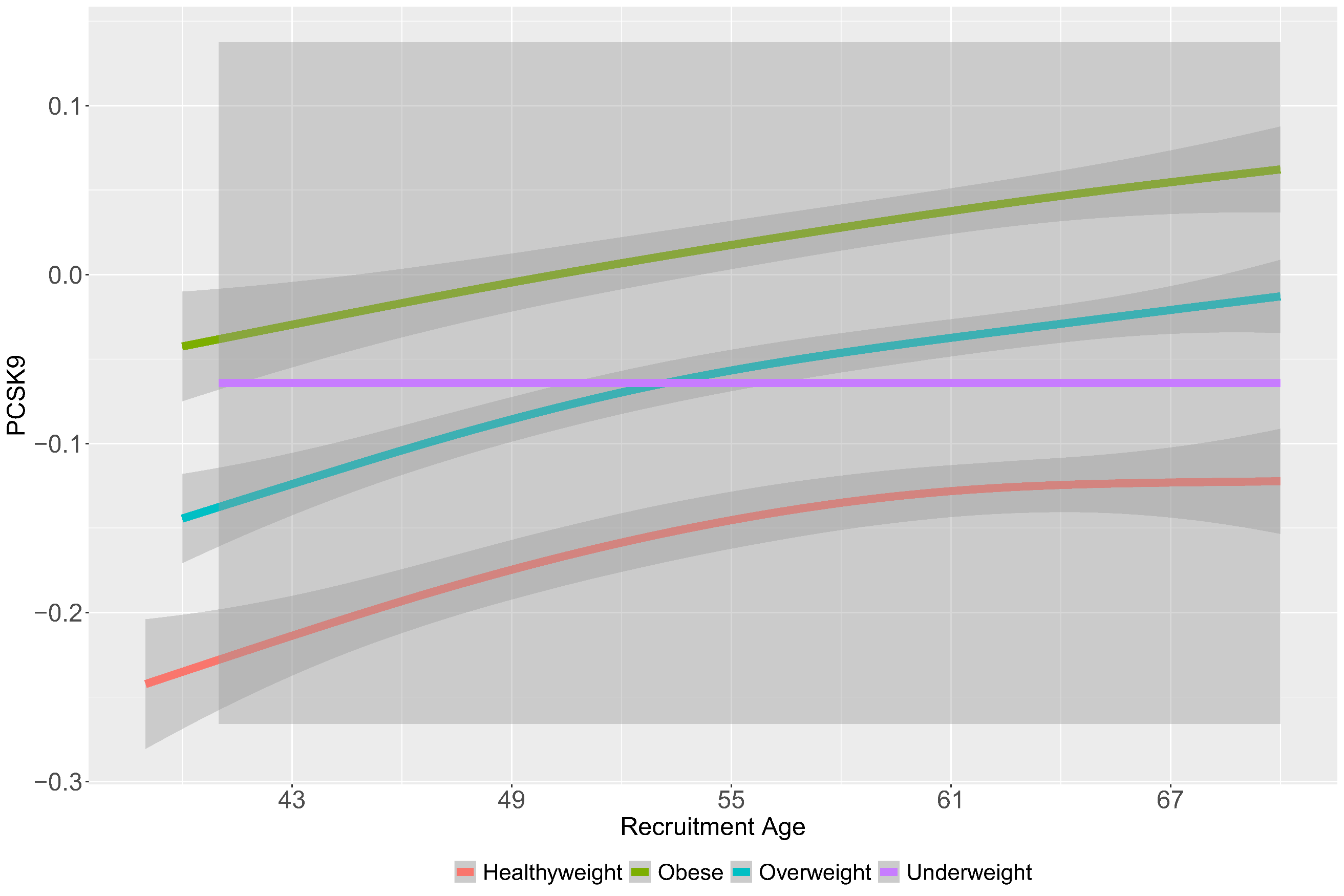

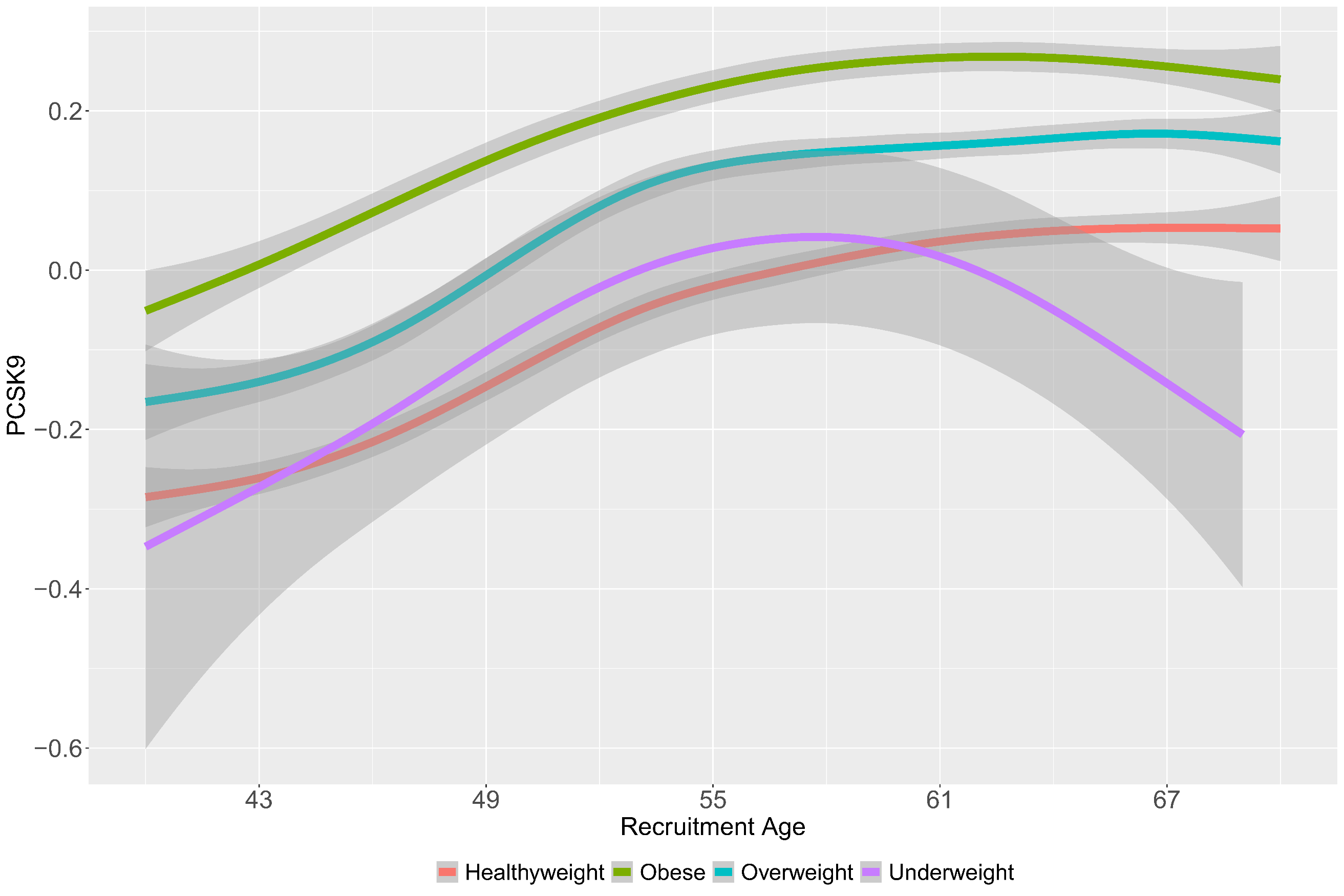

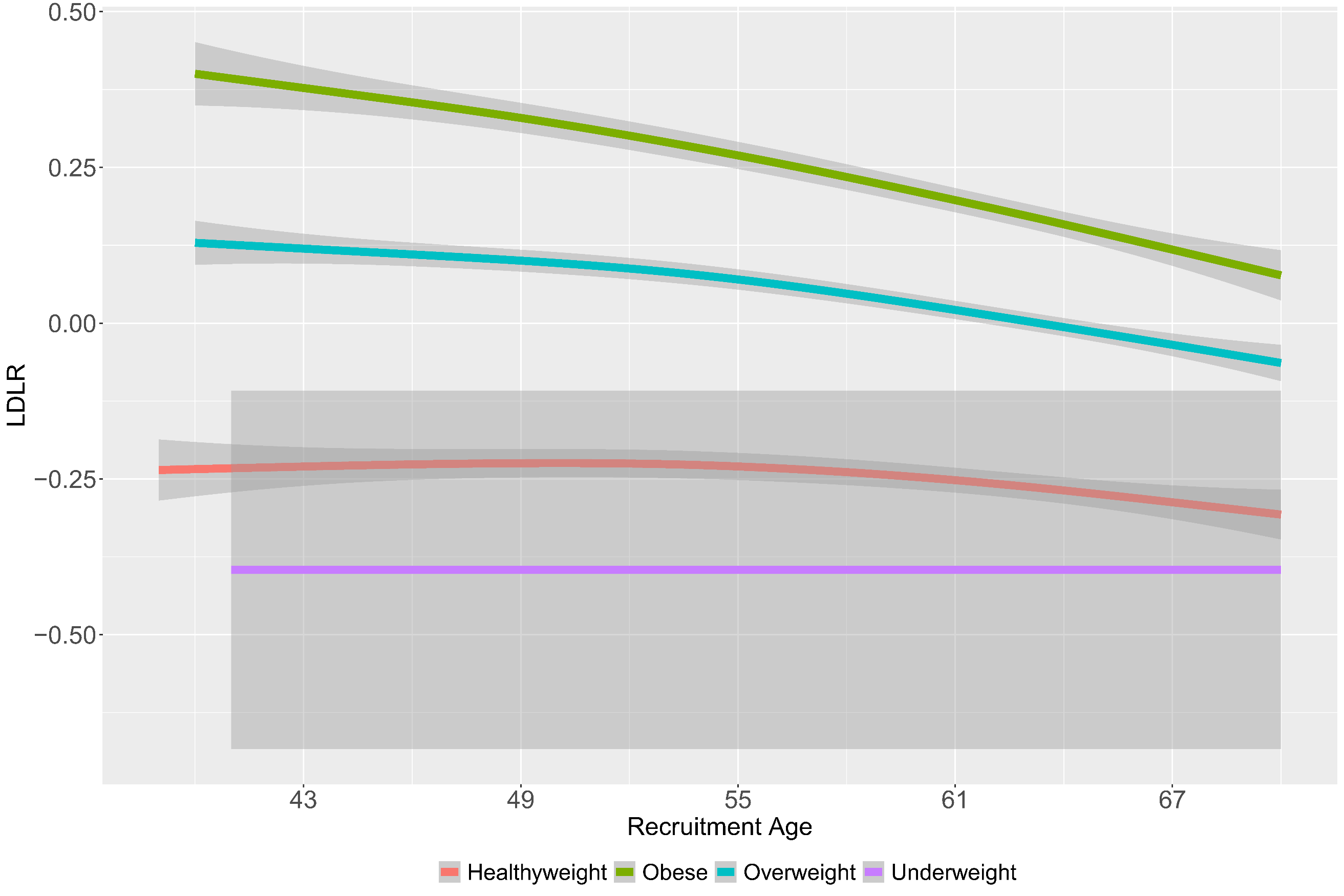

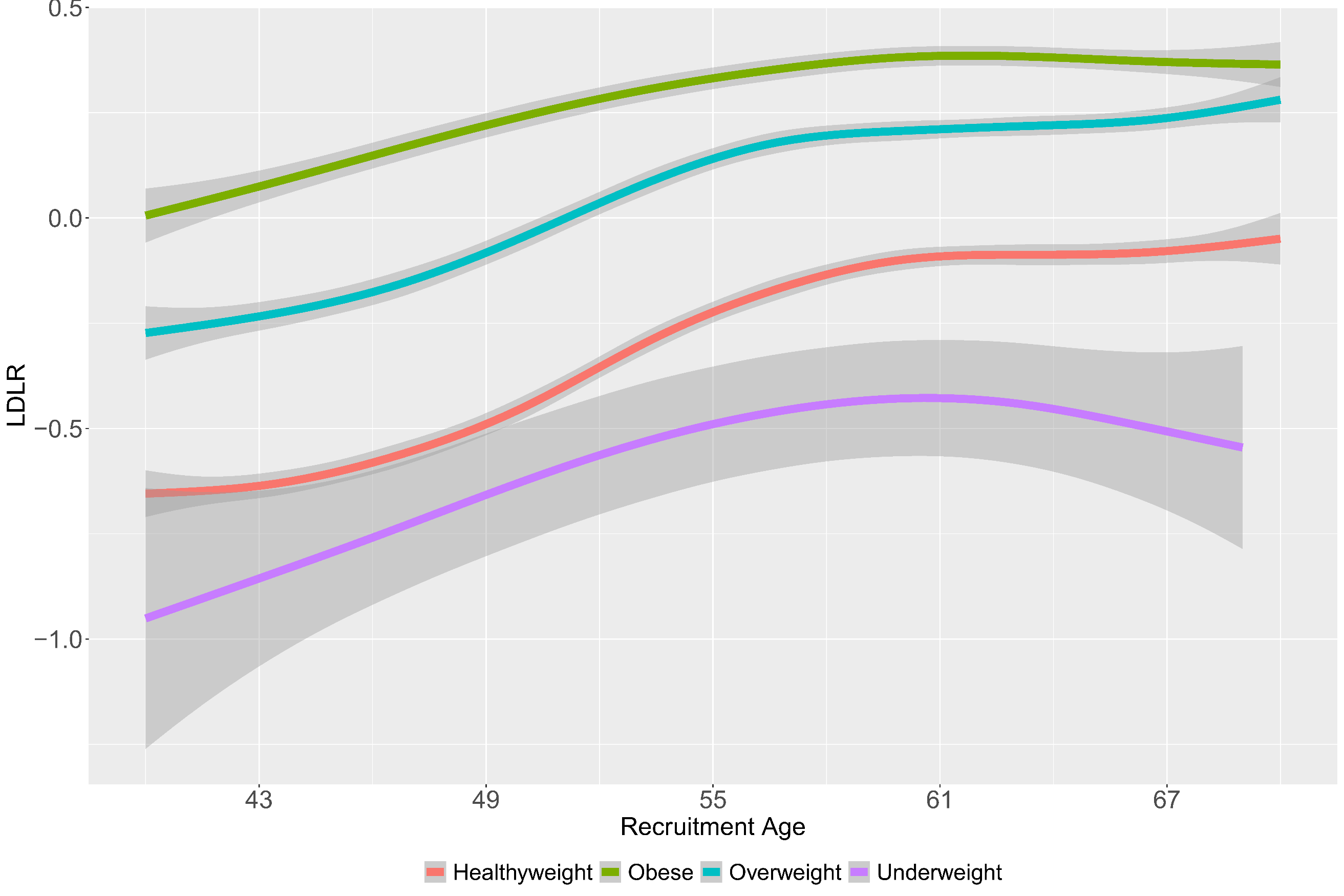

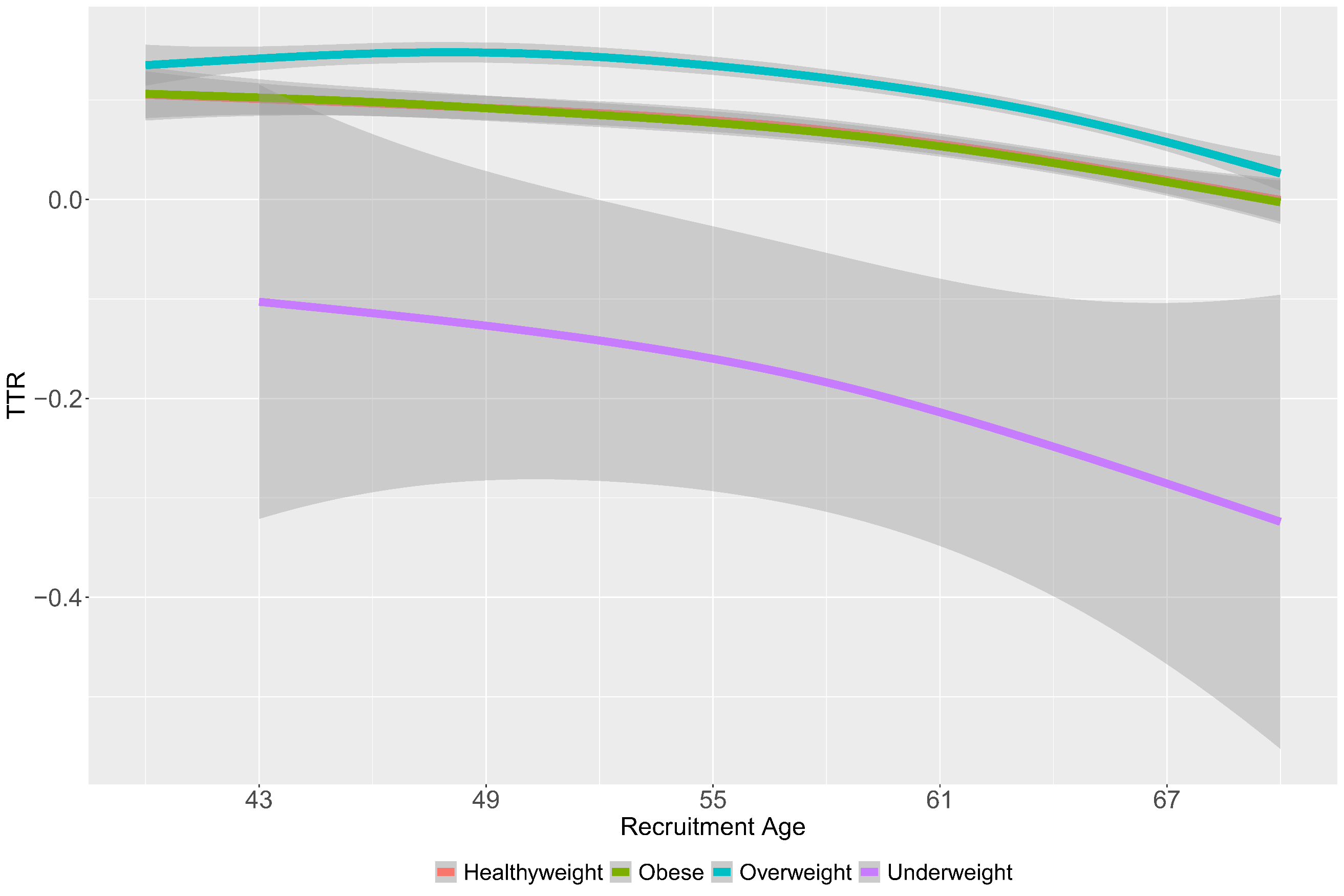

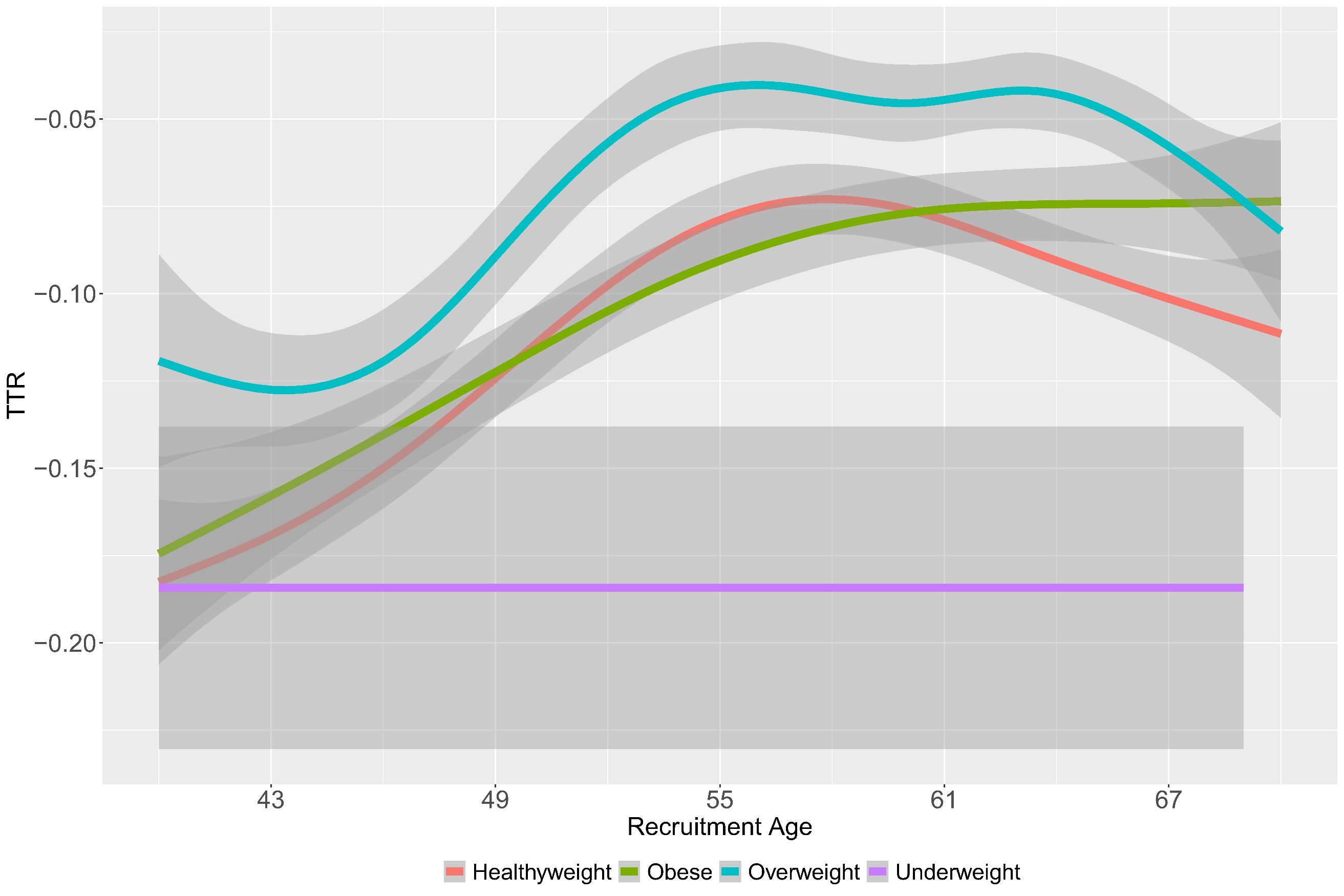

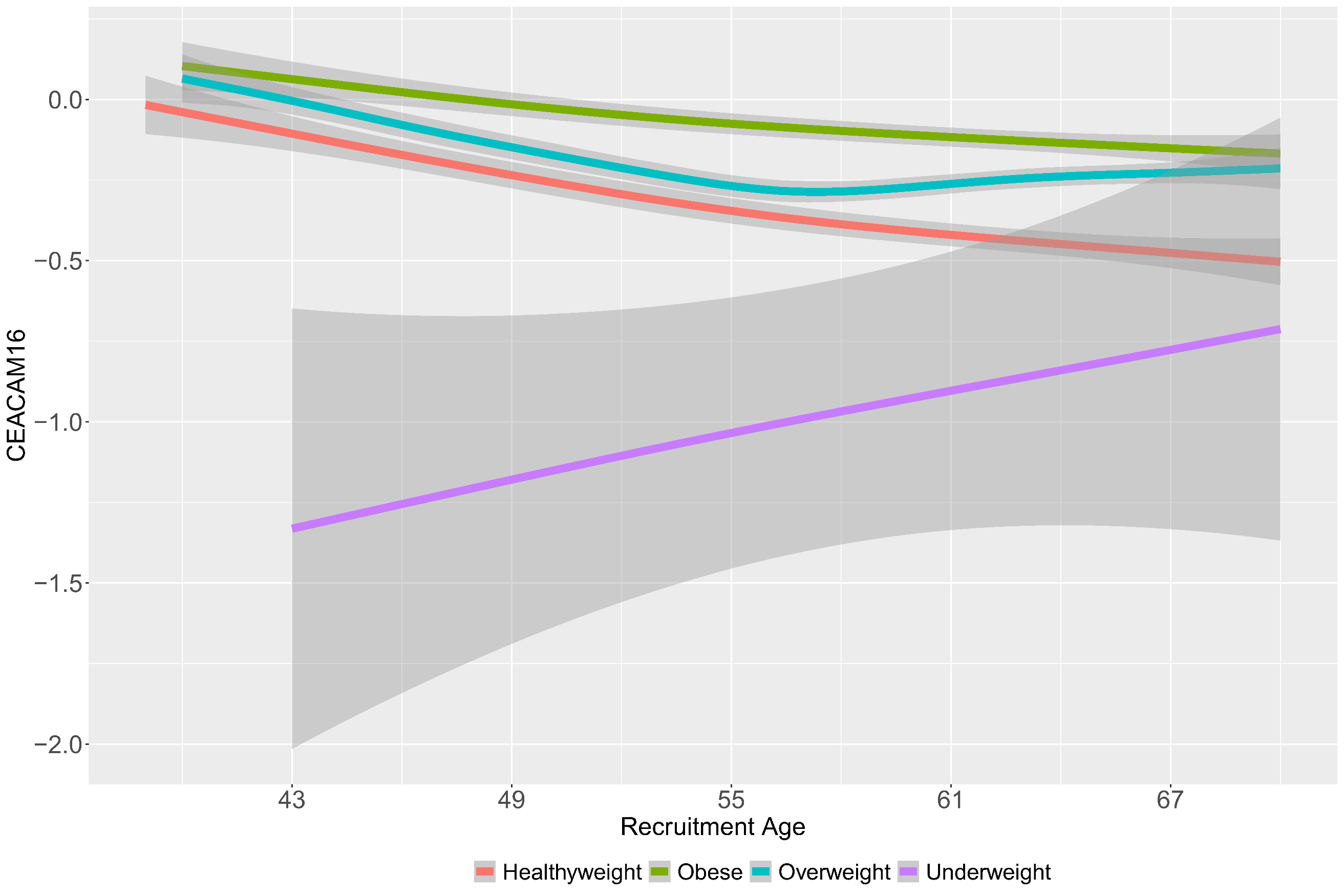

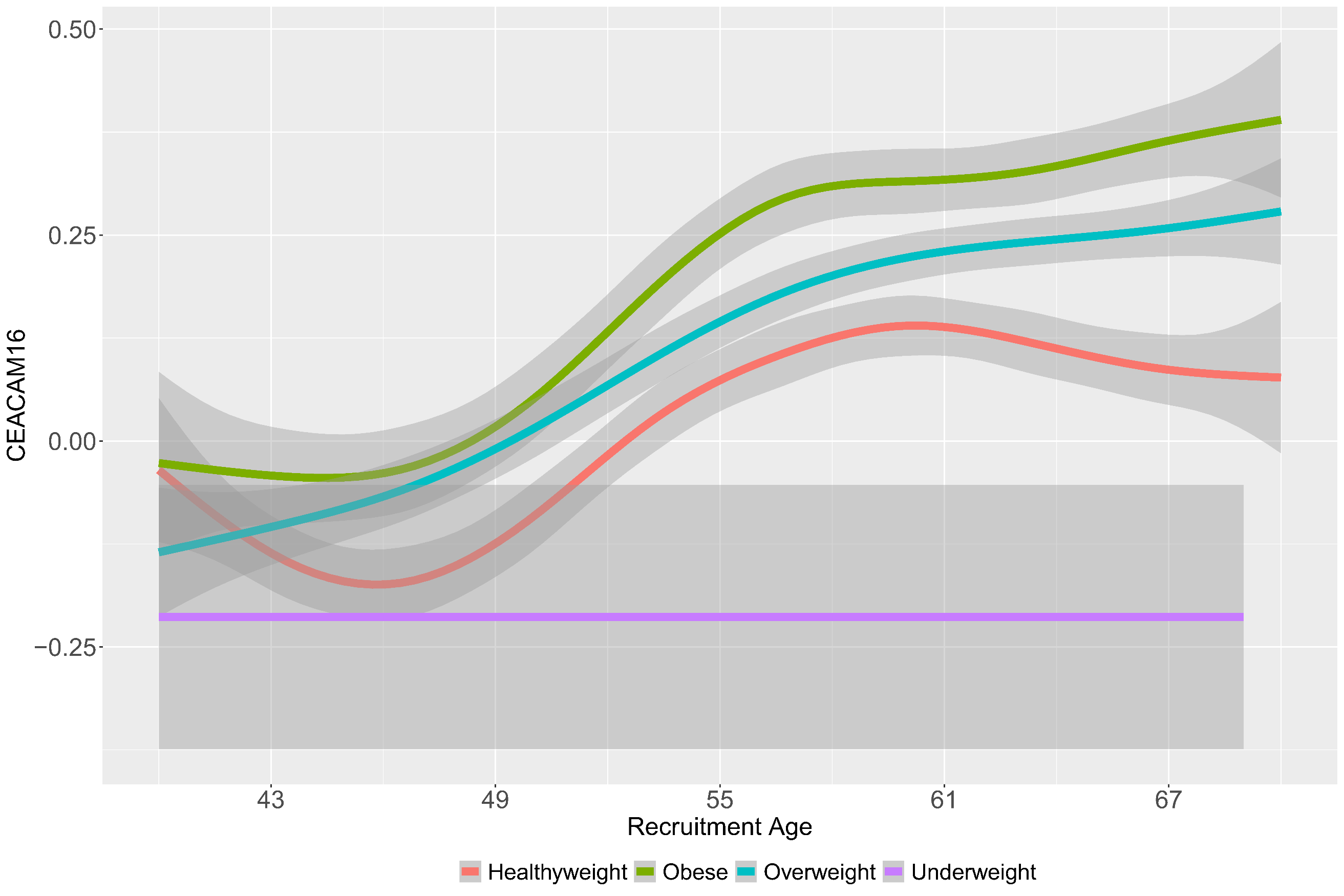

**Supplementary Figure 10**: Age distribution plot of the top aging related sex dimorphic proteins by BMI groups. Left panel depicts females and right panel depicts males.

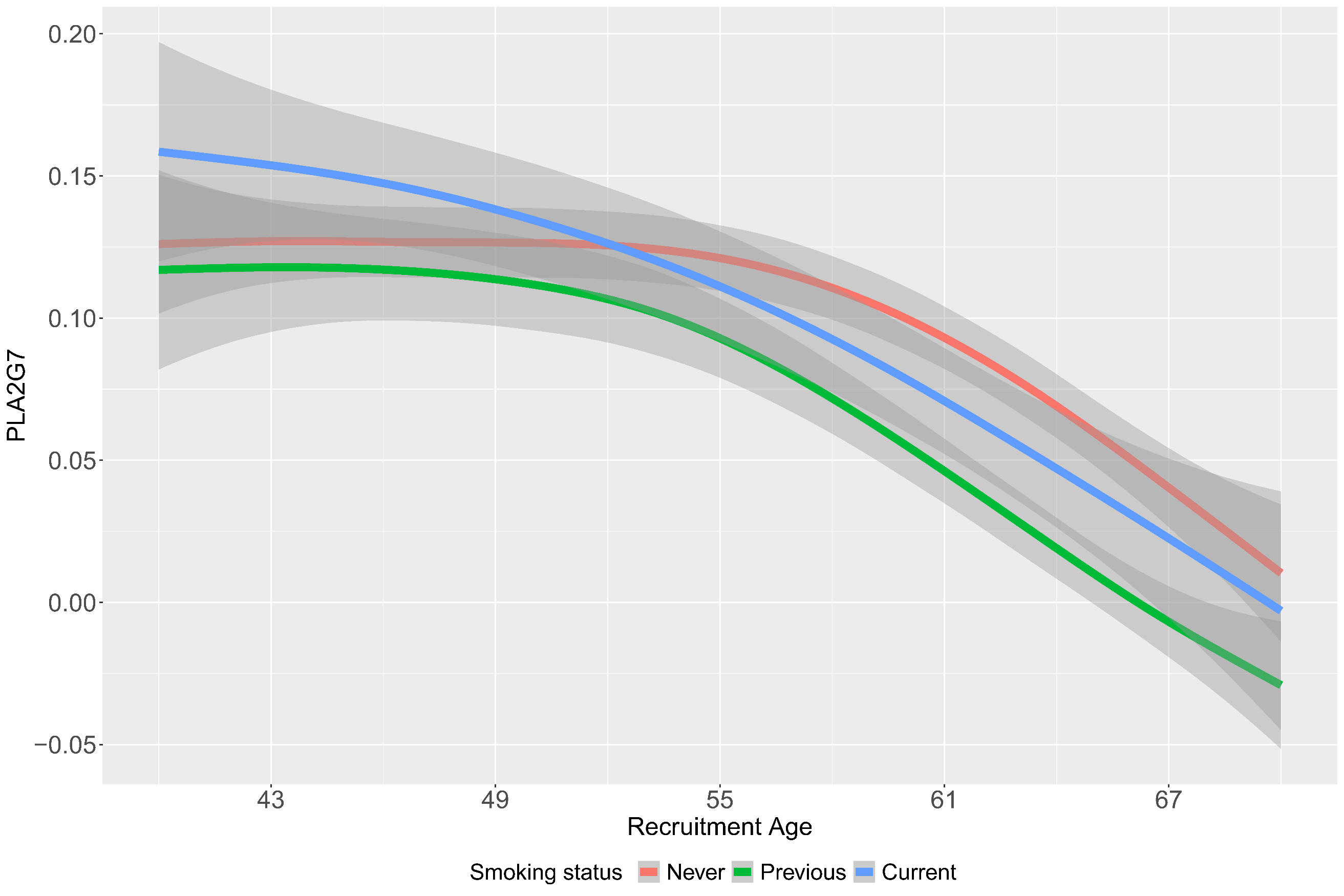

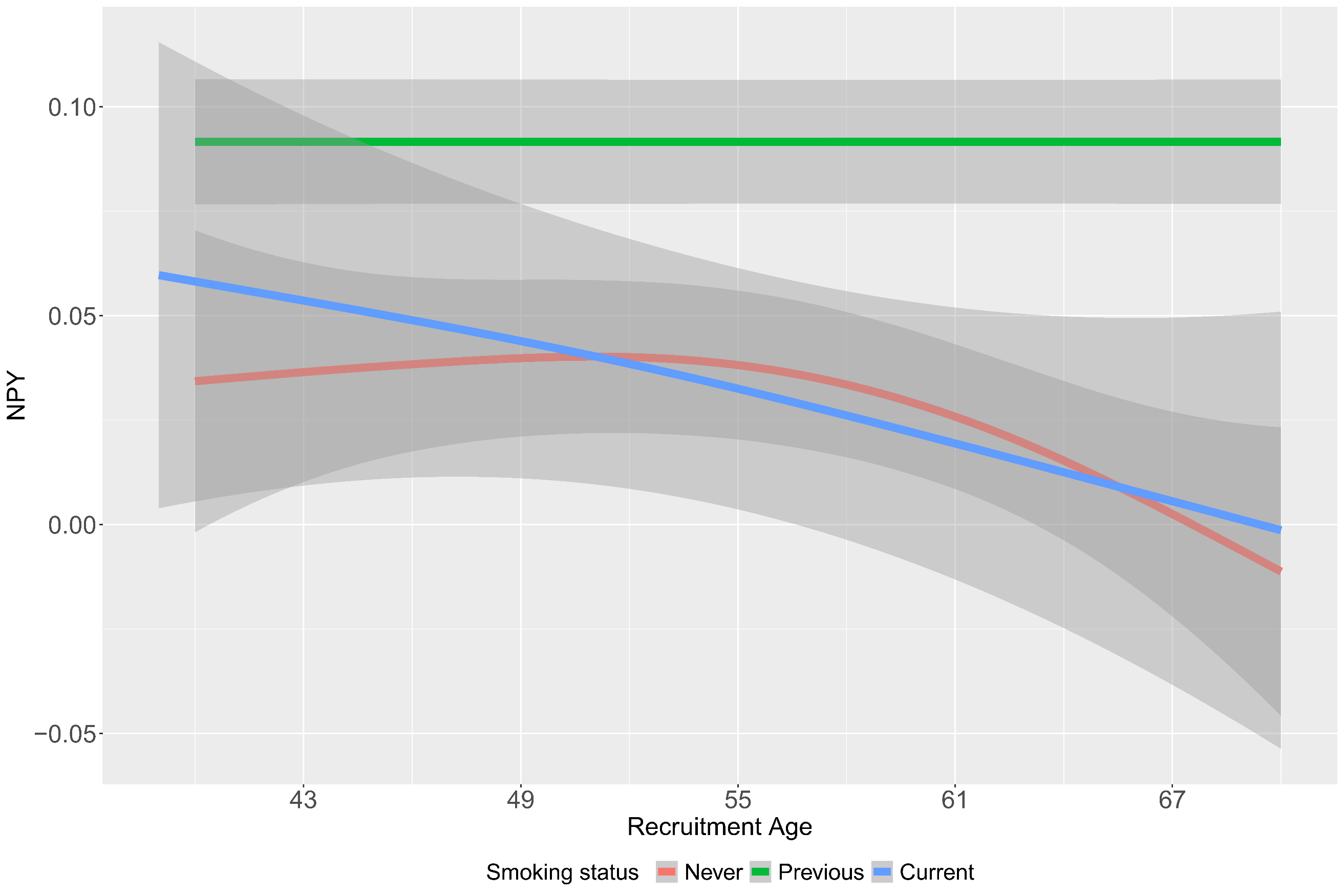

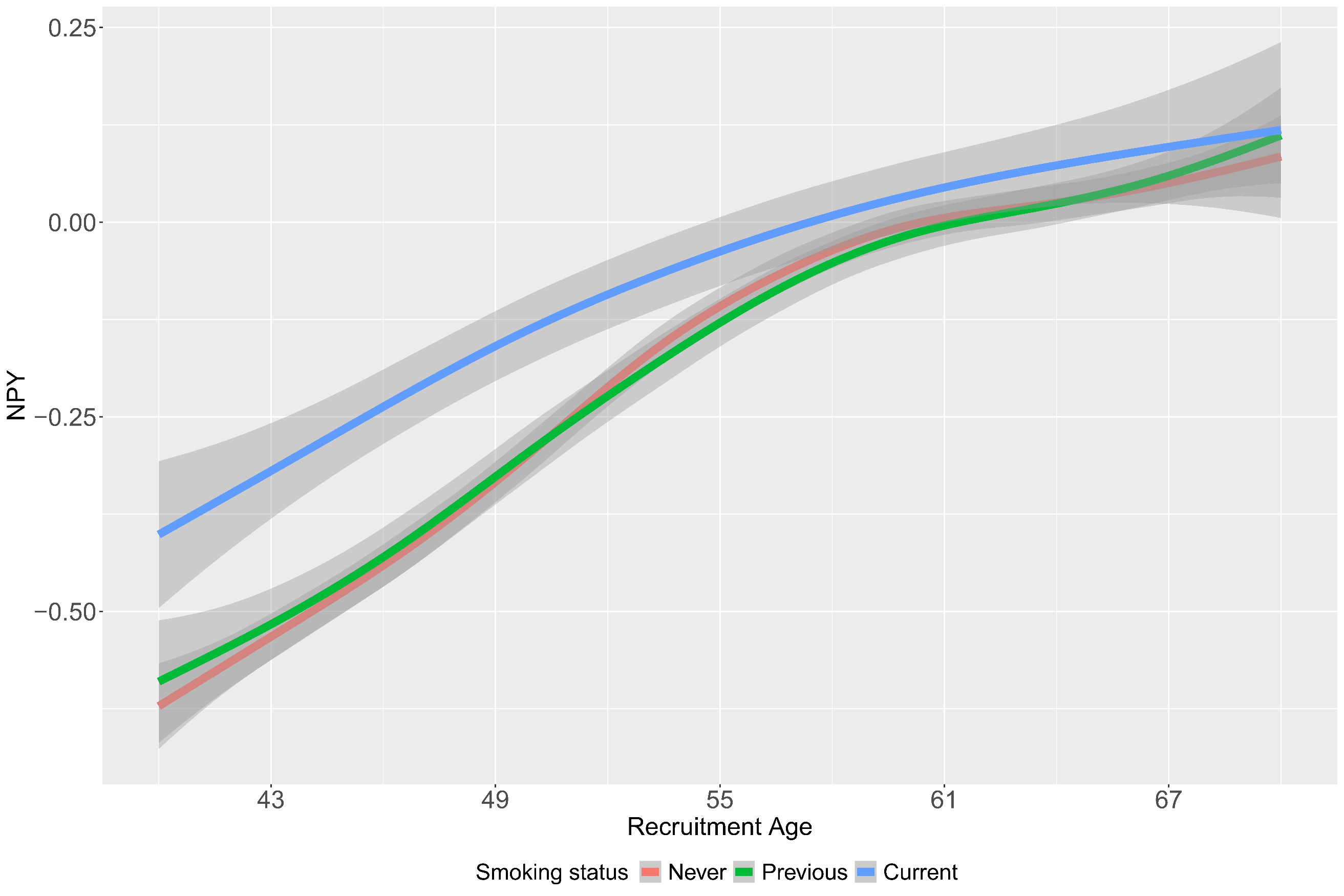

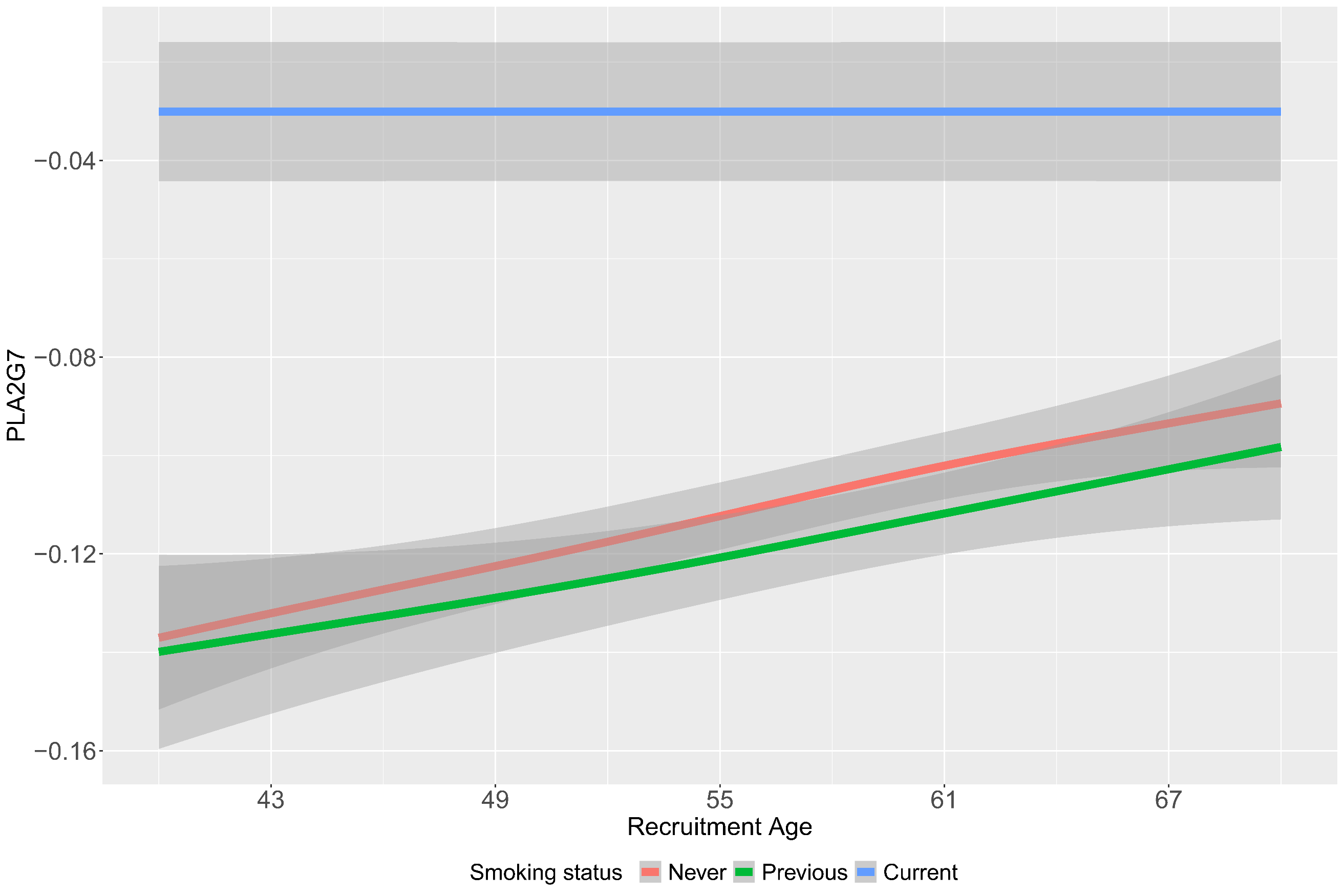

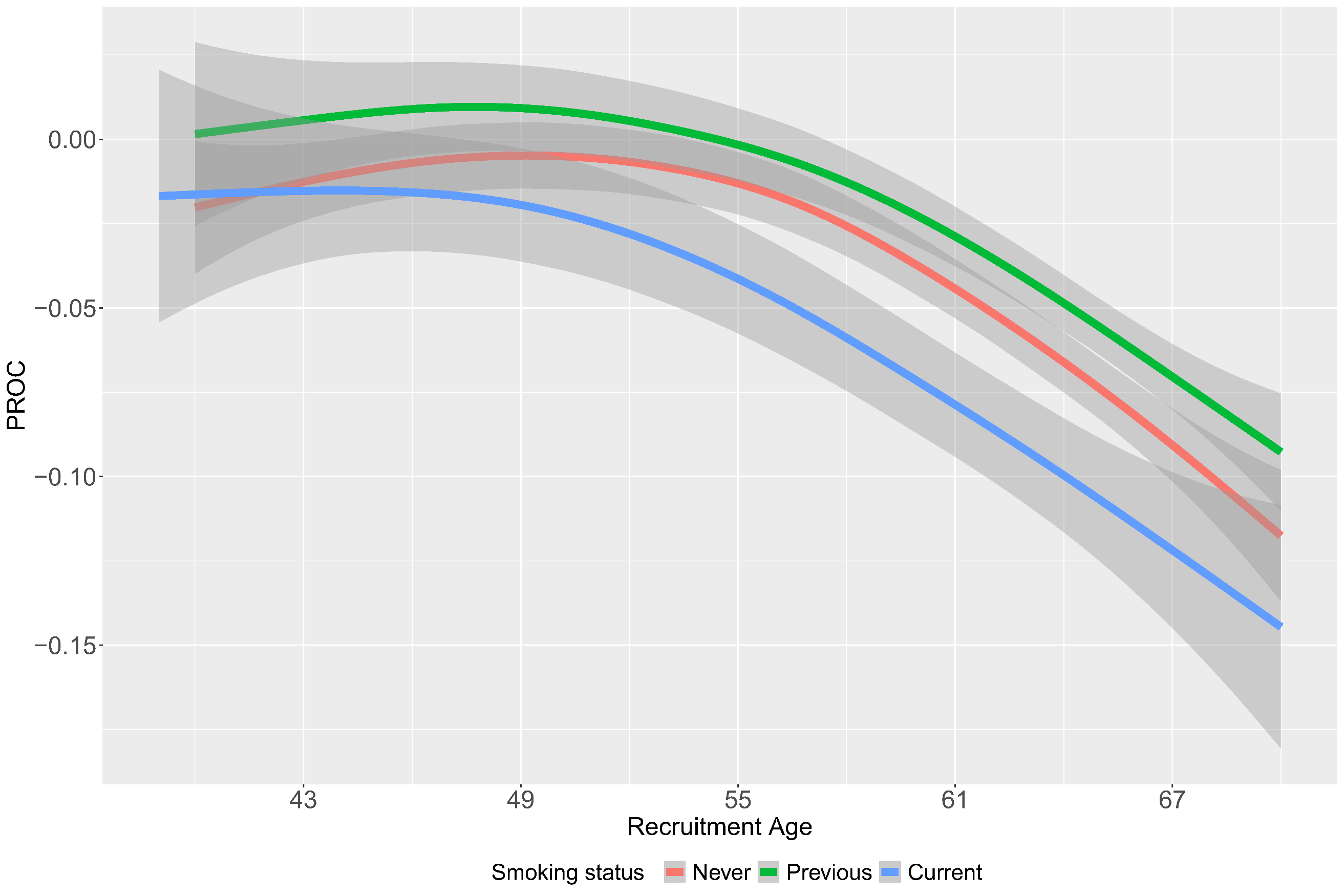

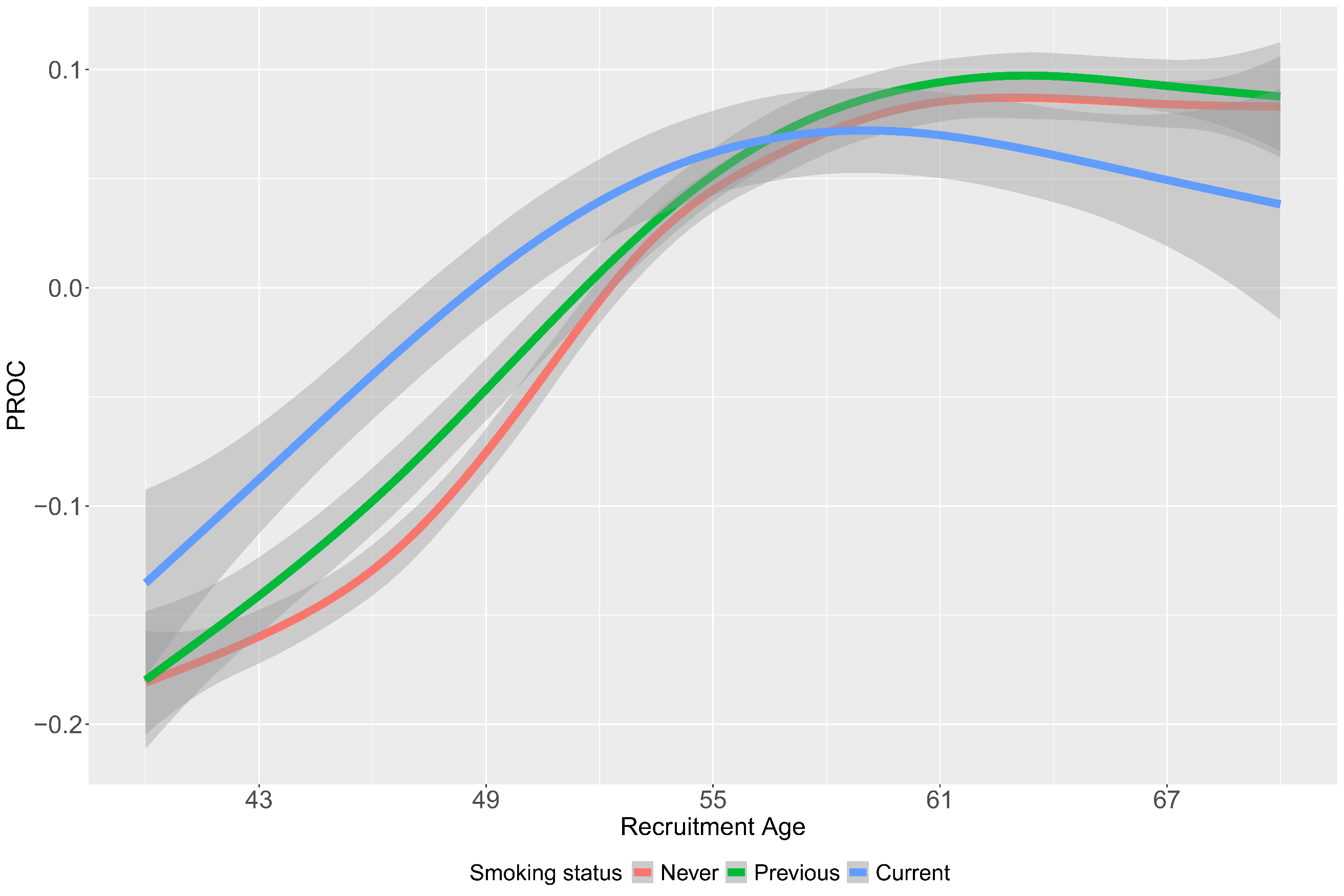

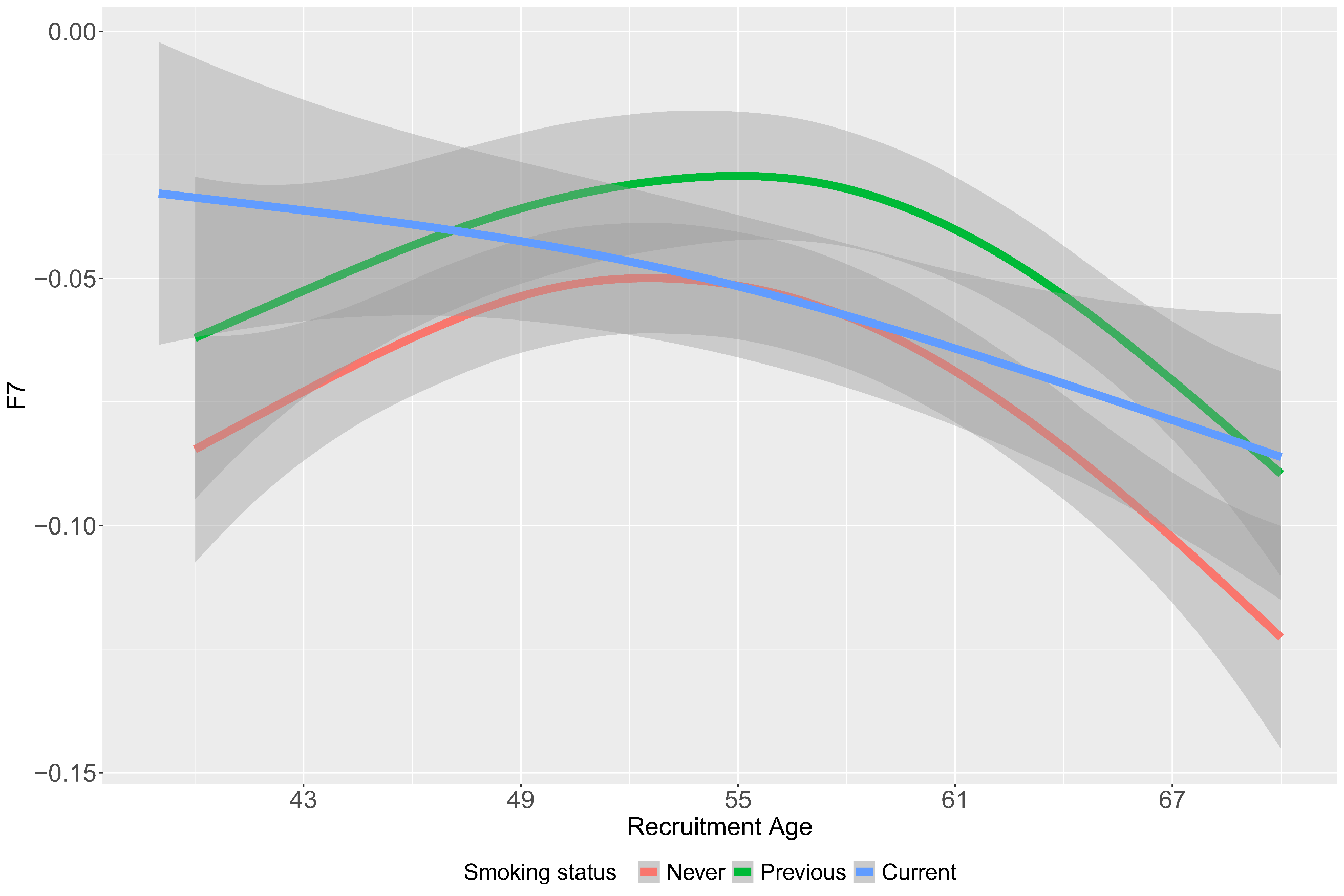

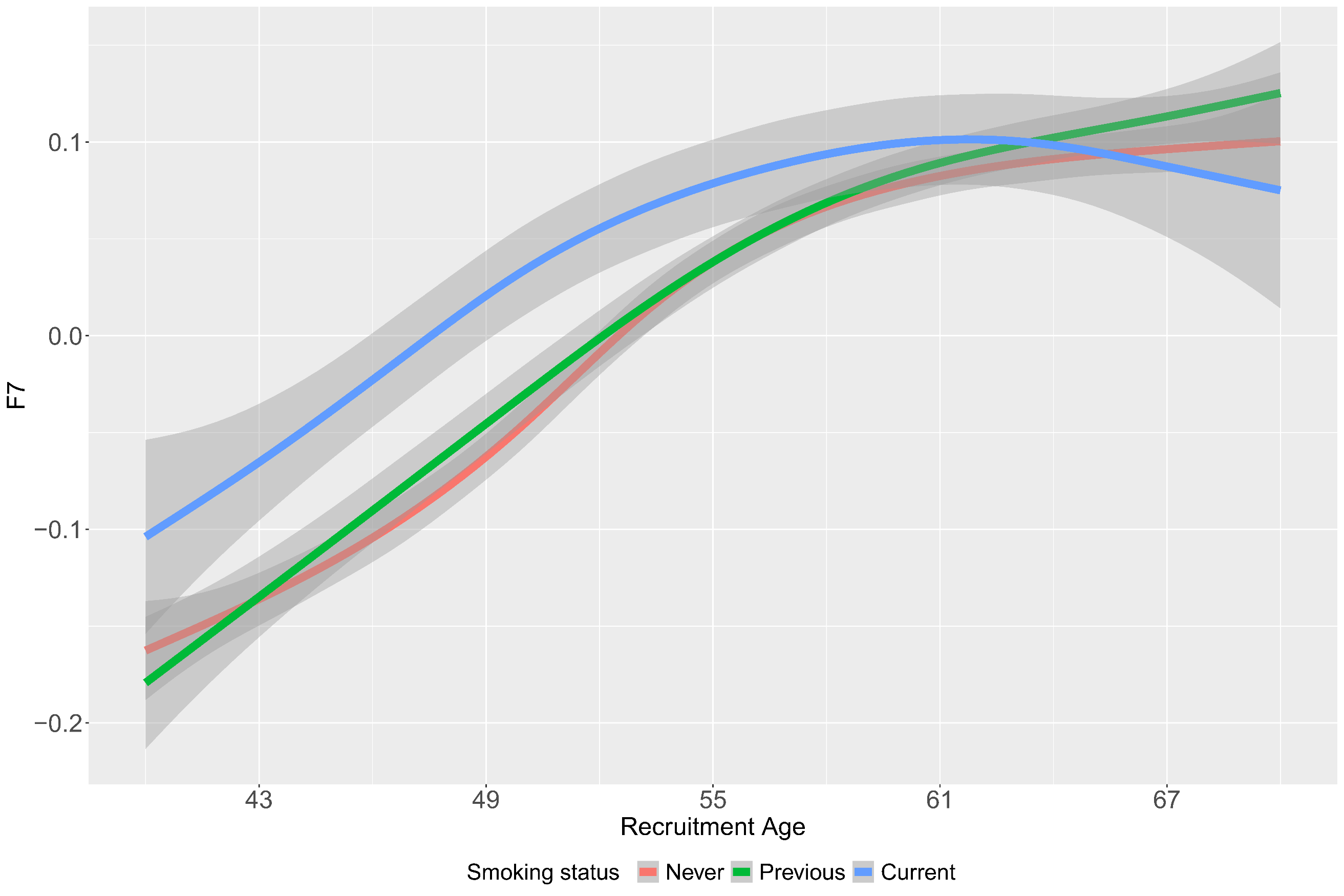

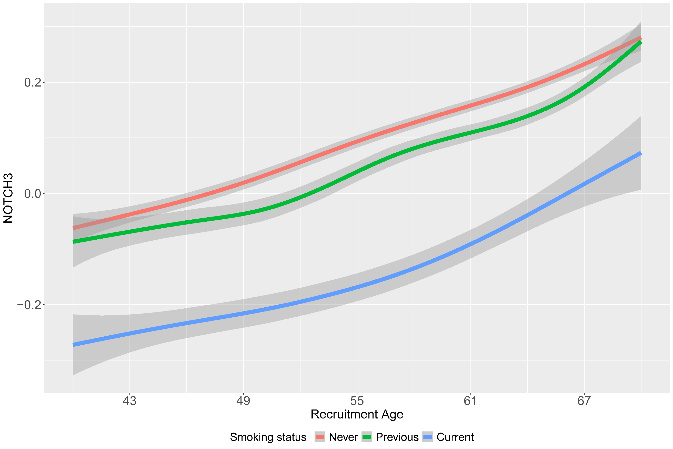

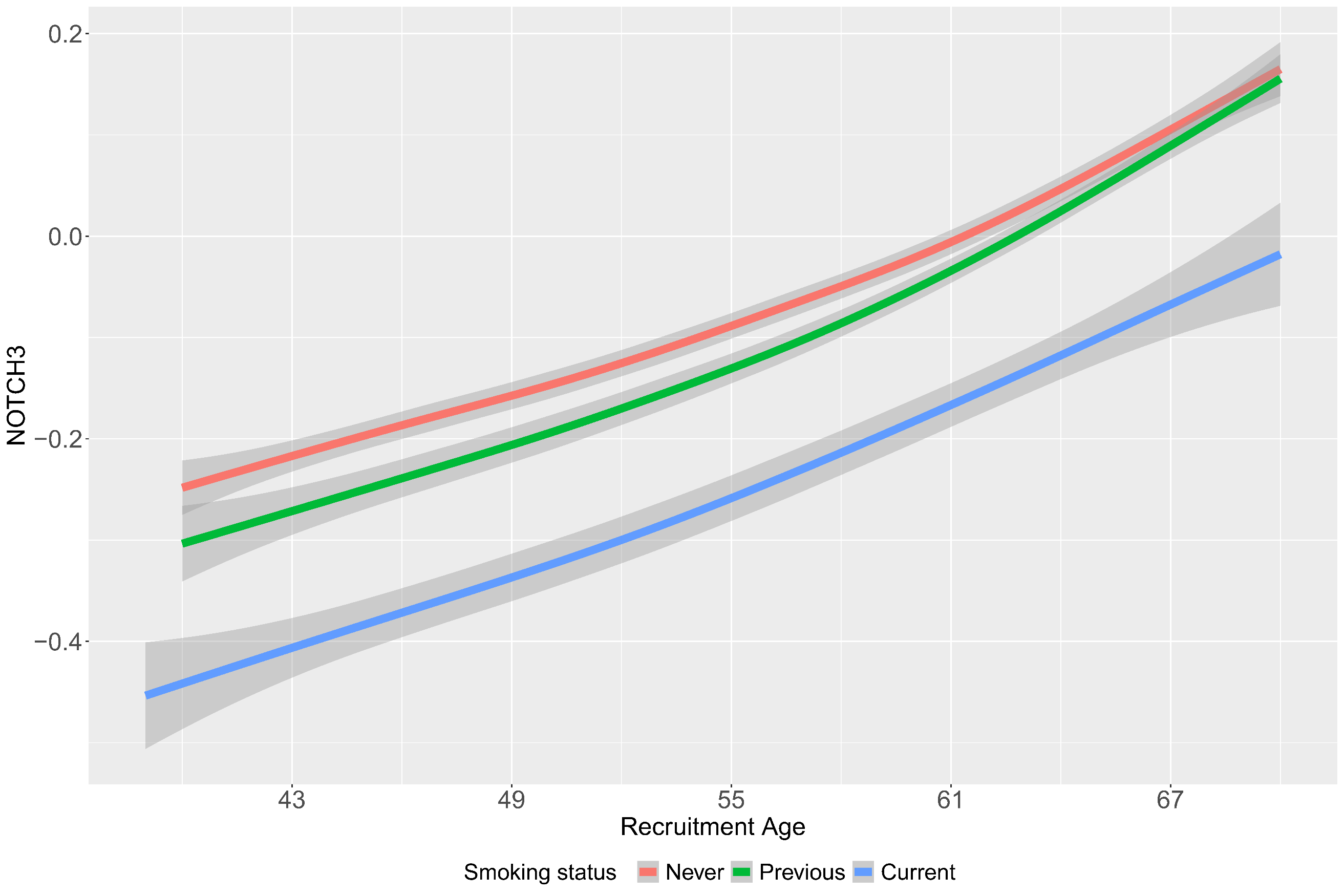

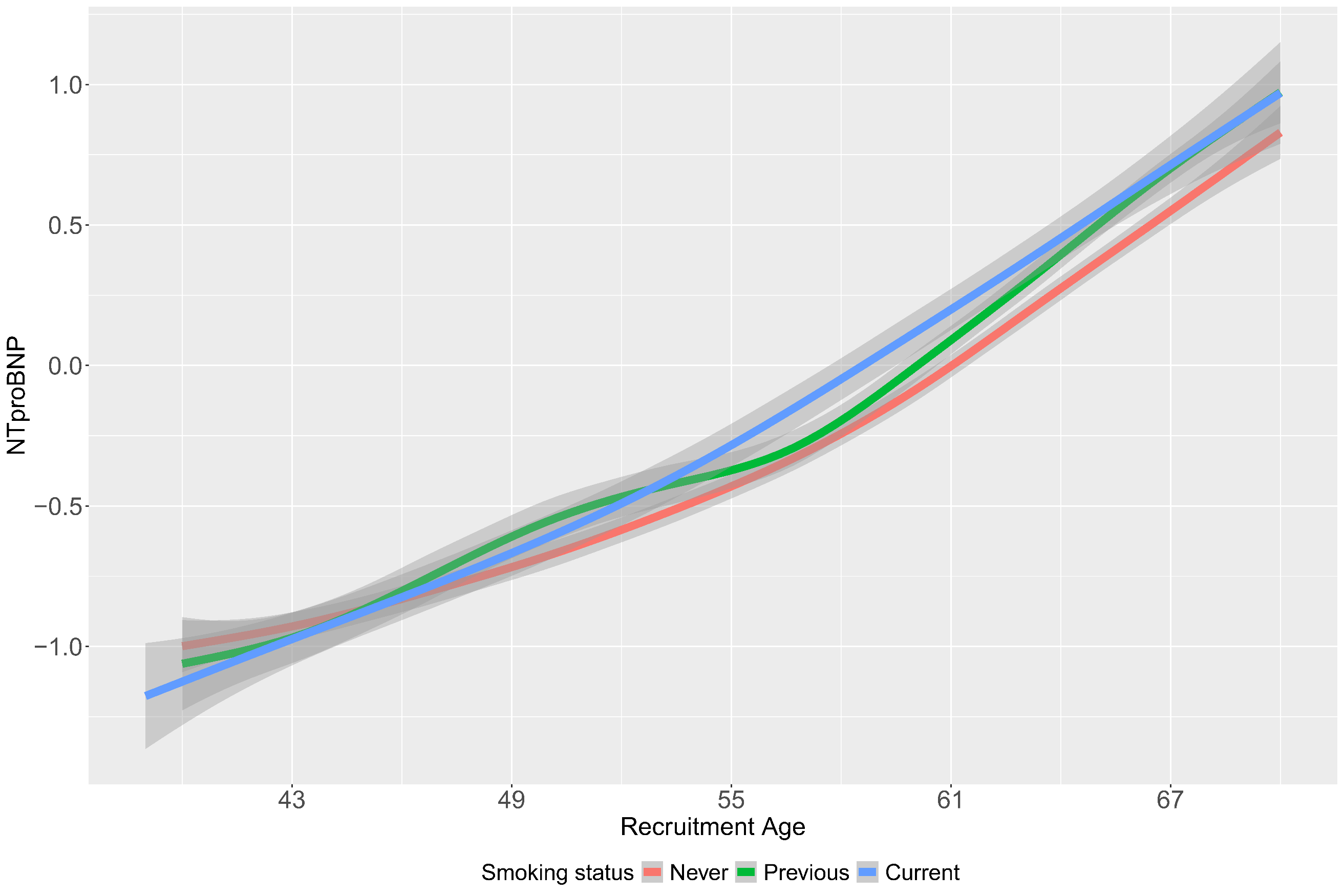

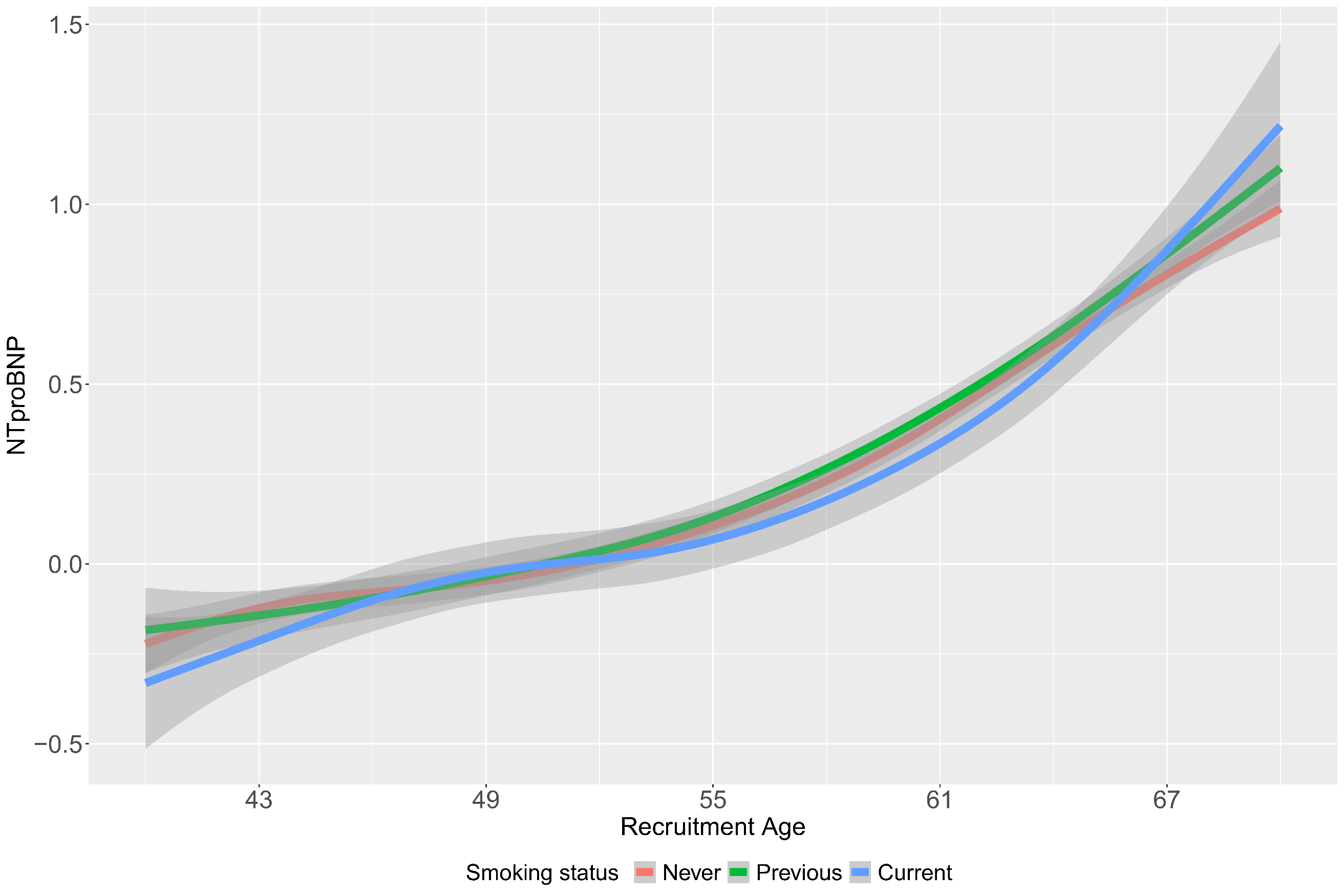

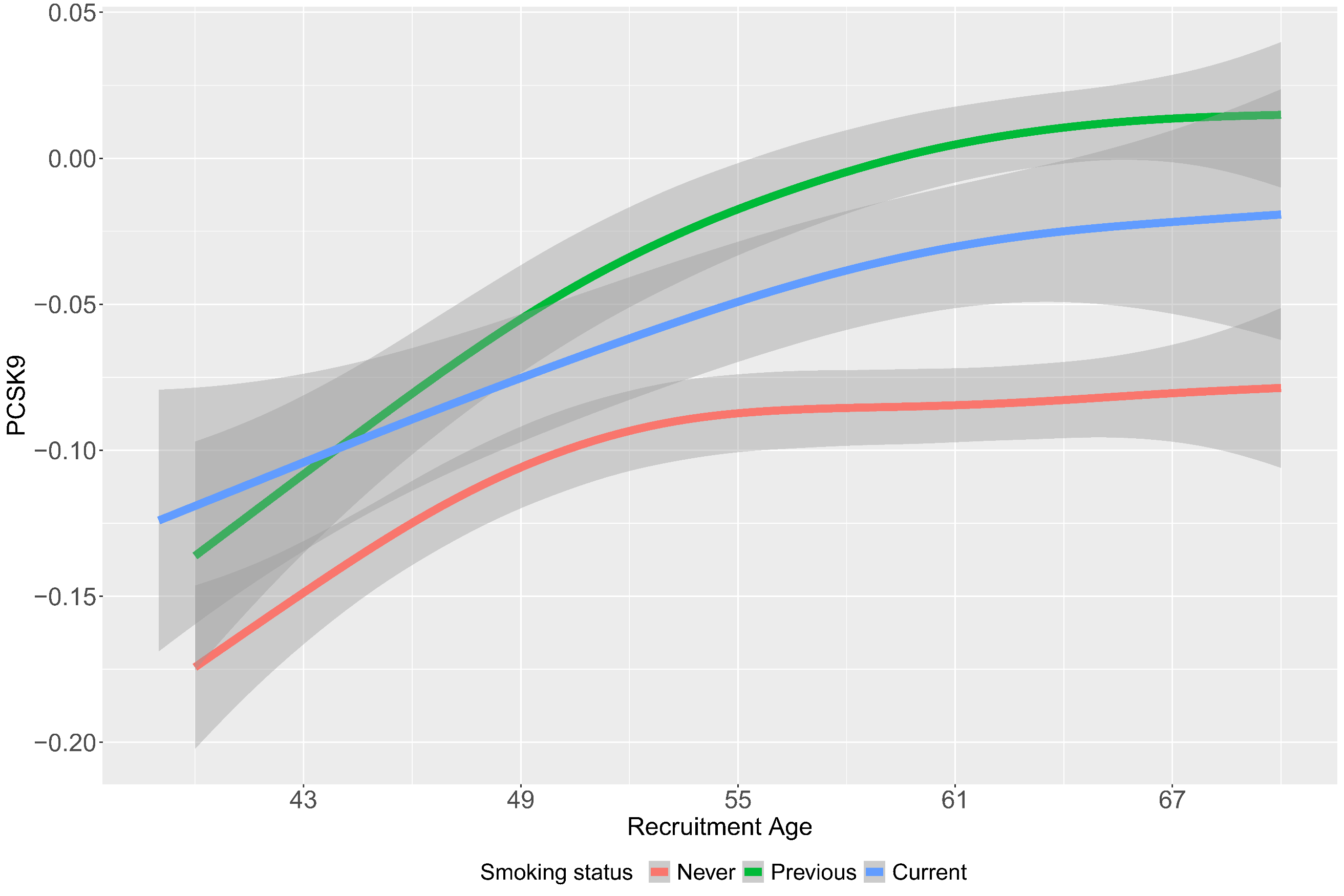

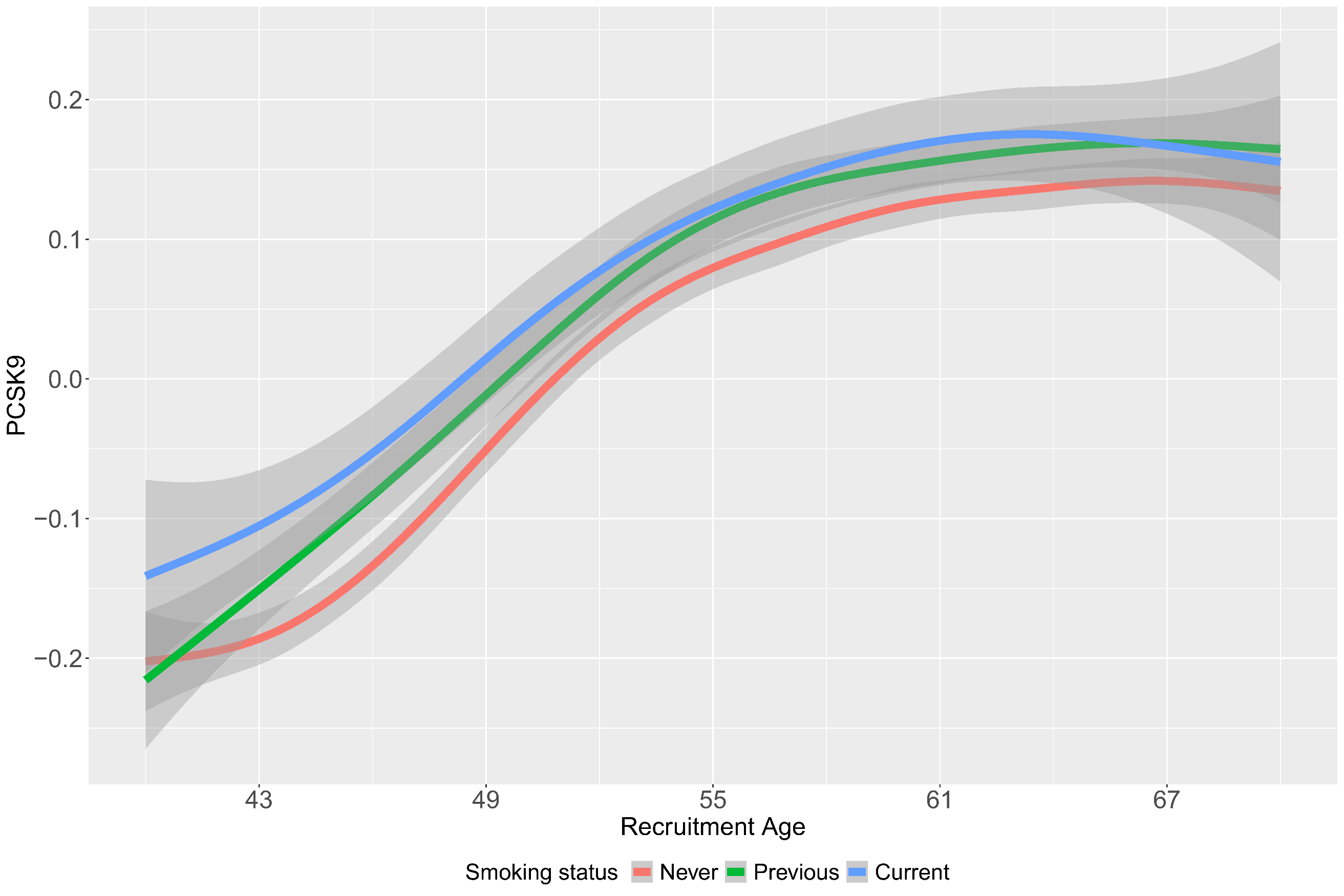

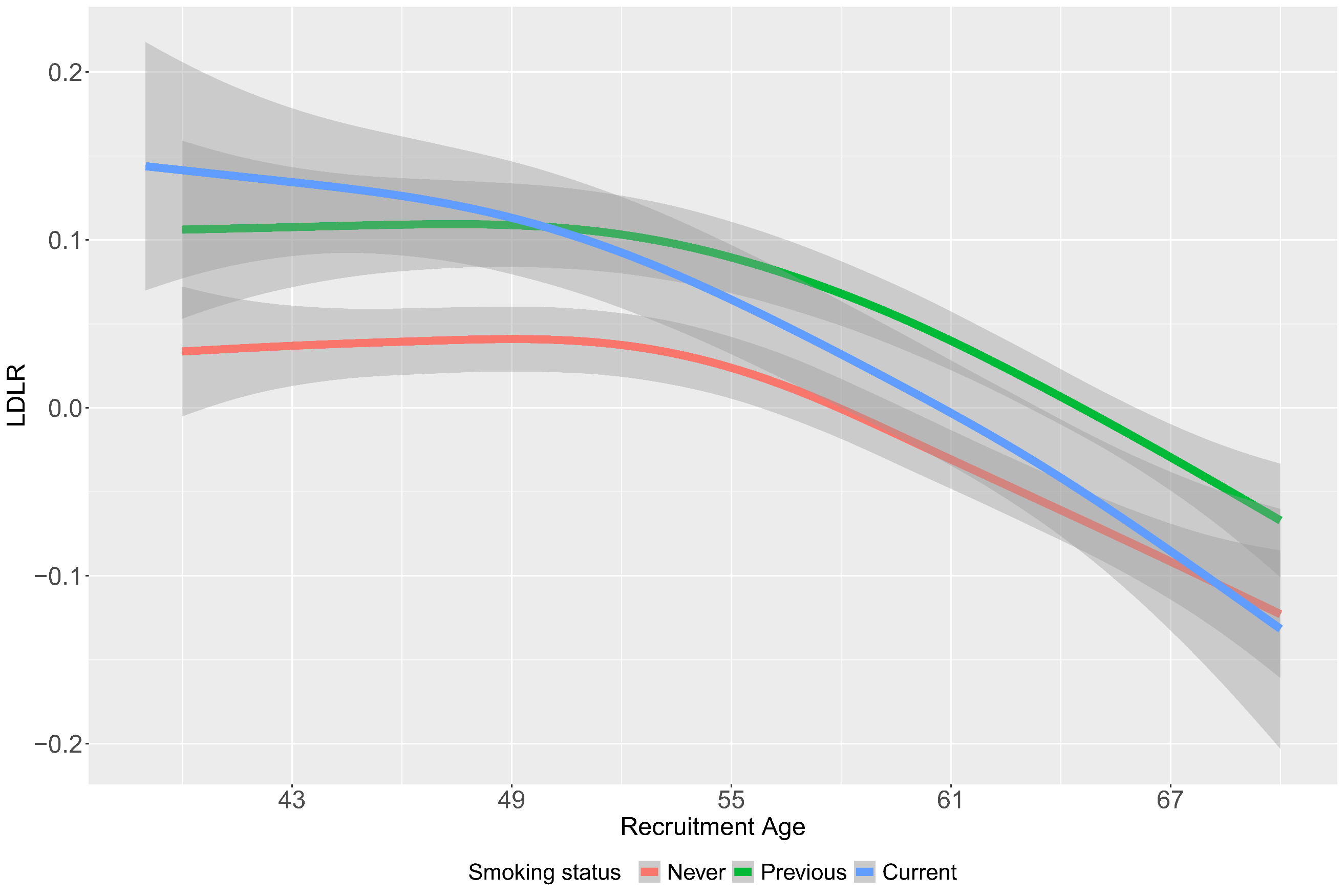

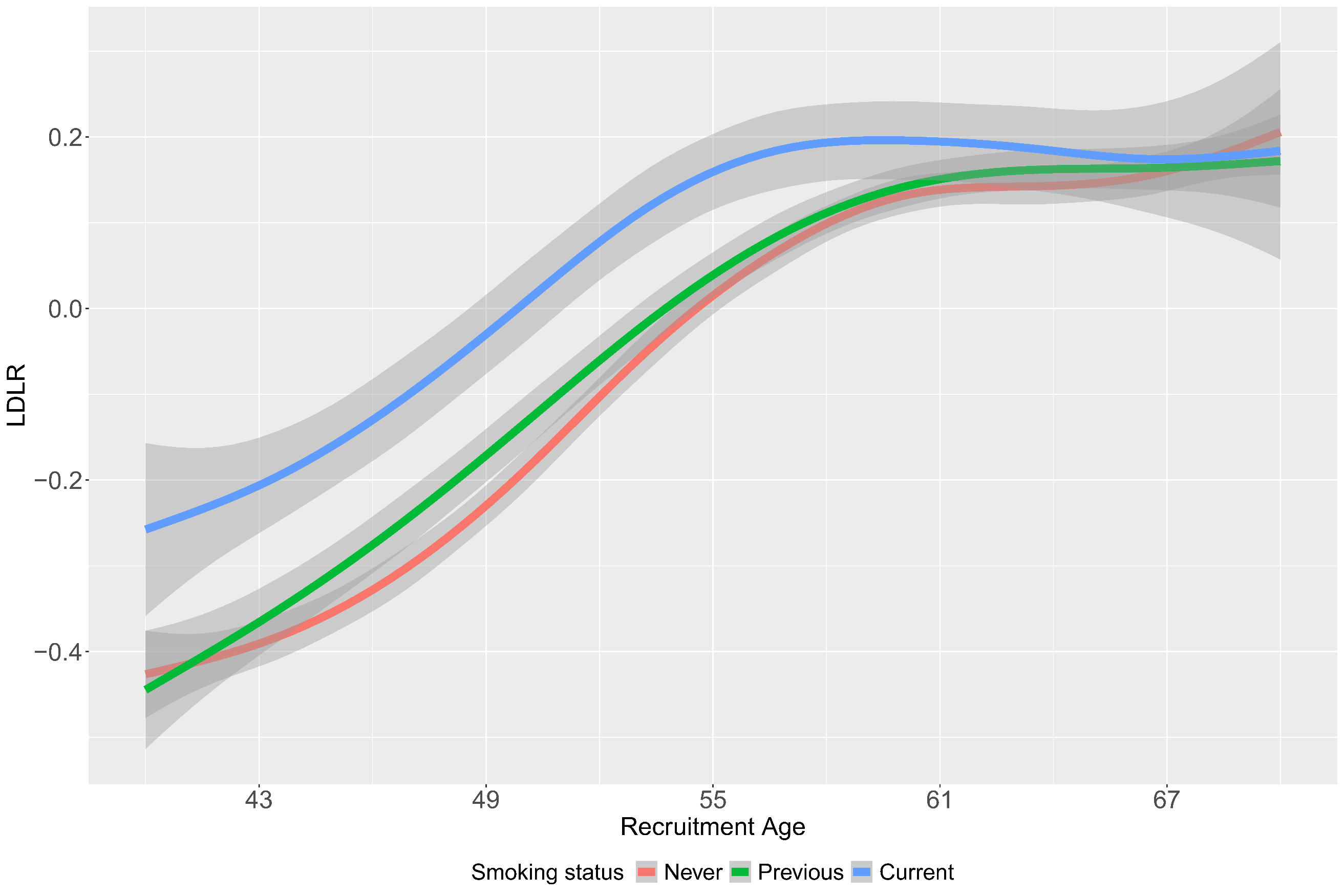

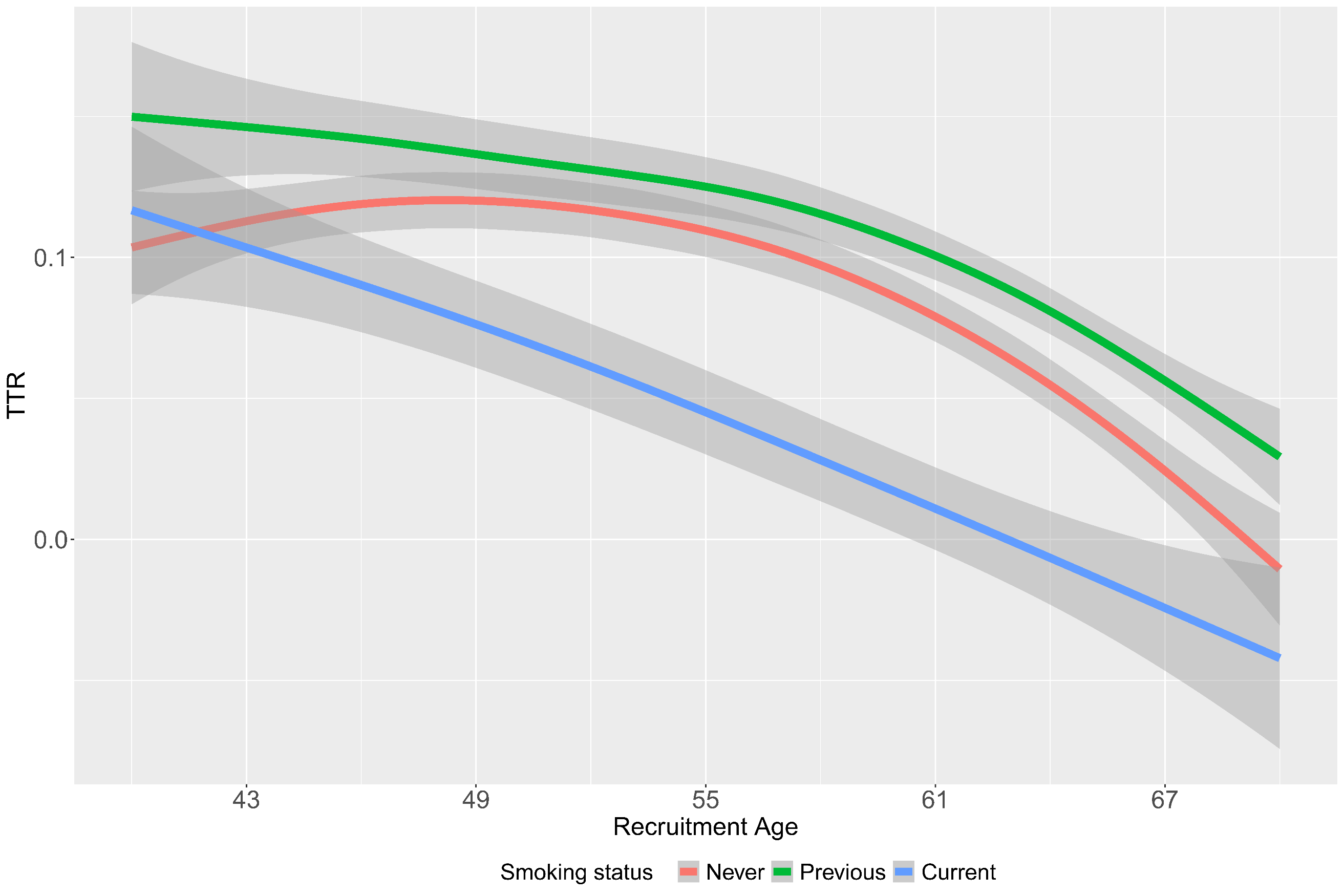

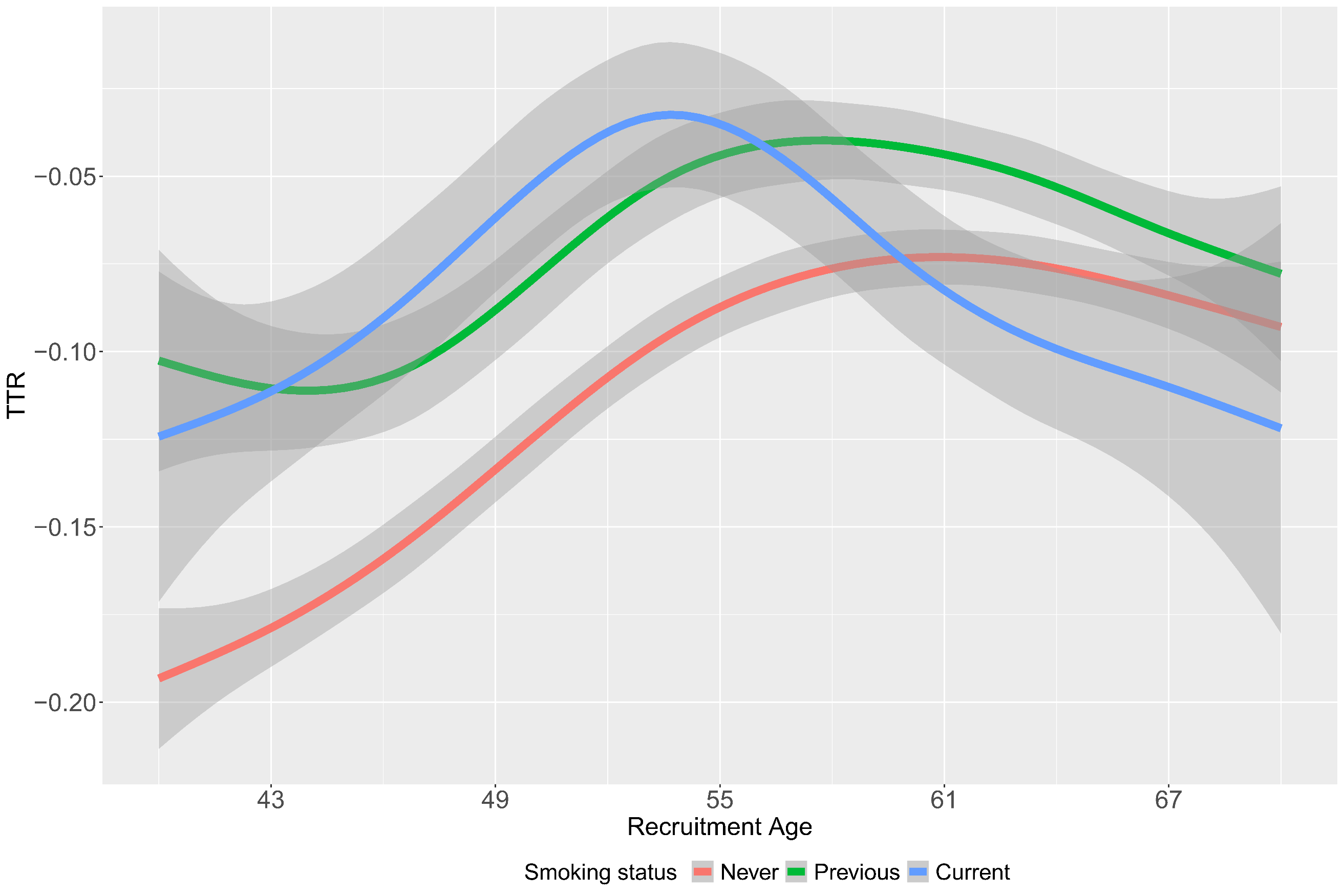

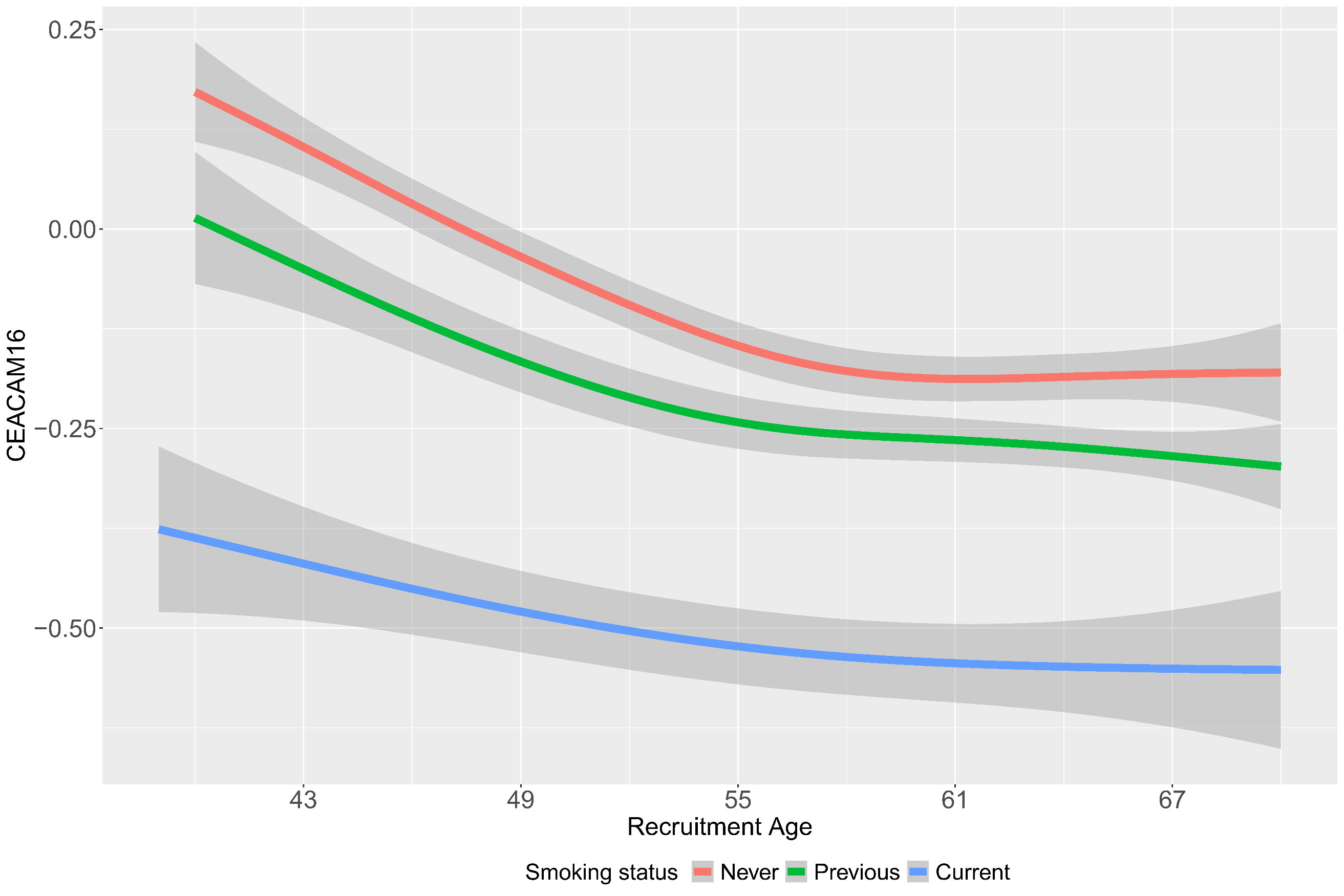

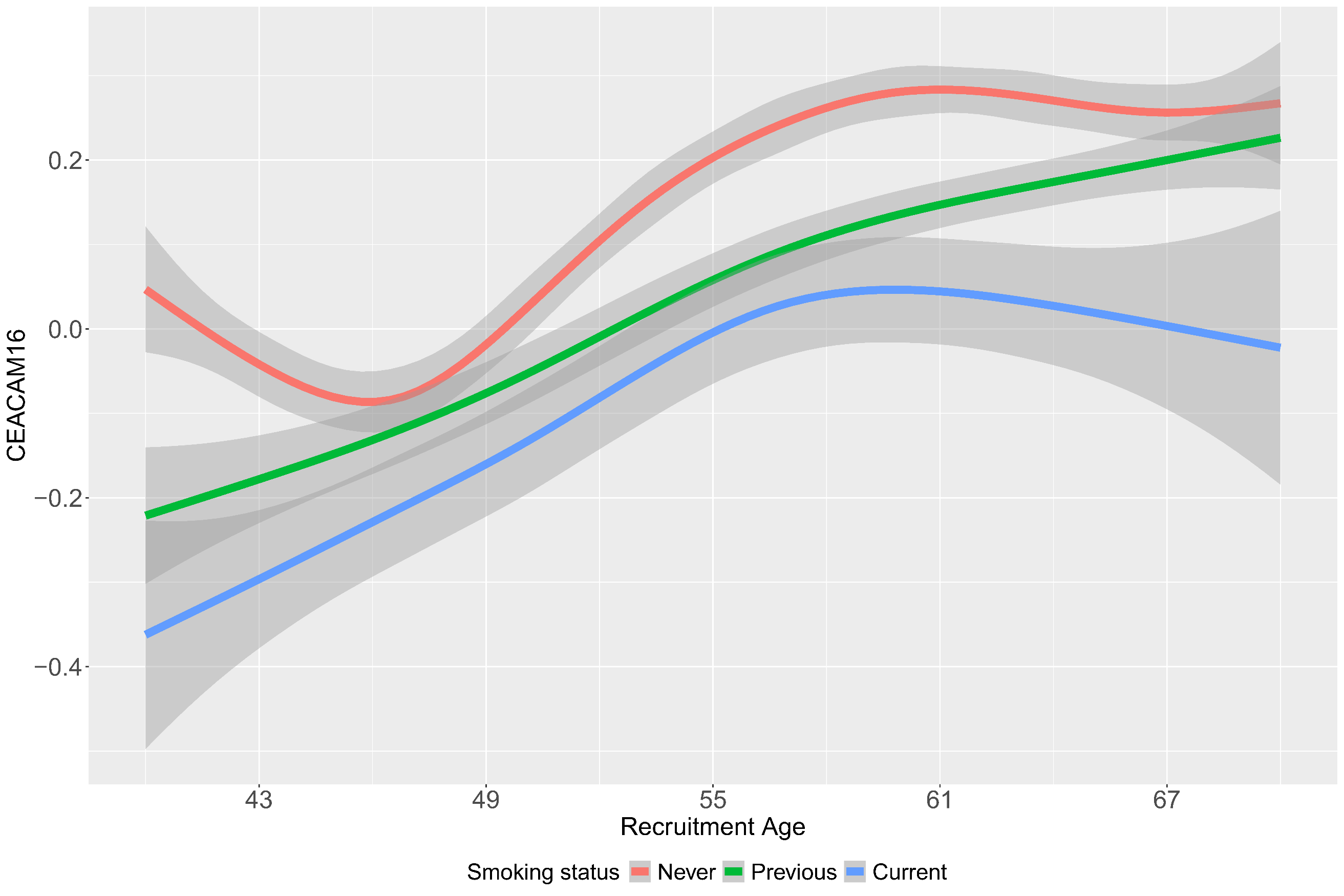

**Supplementary Figure 11**: Age distribution plot of the top aging related sex dimorphic proteins by smoking status. Left panel depicts females and right panel depicts males.

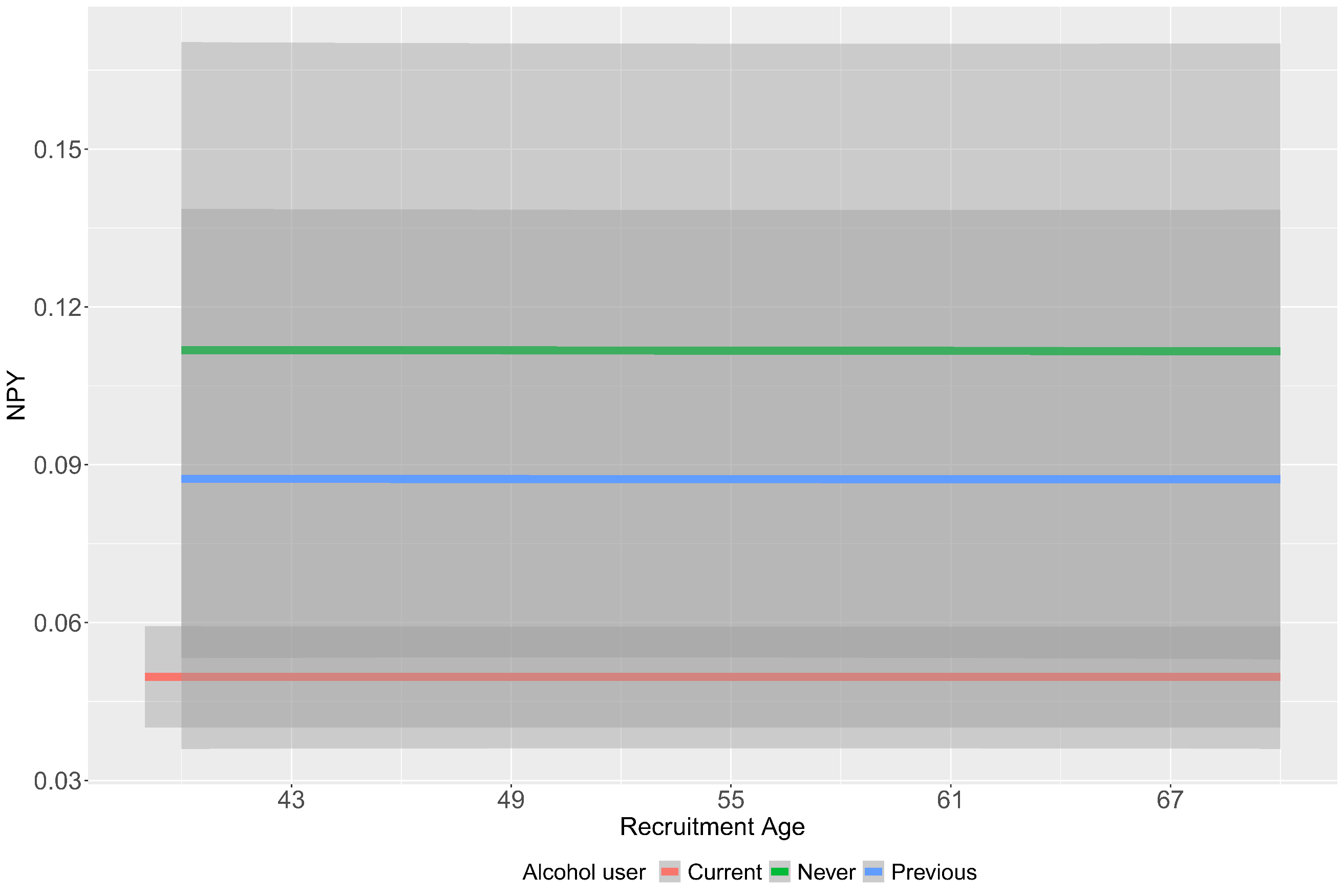

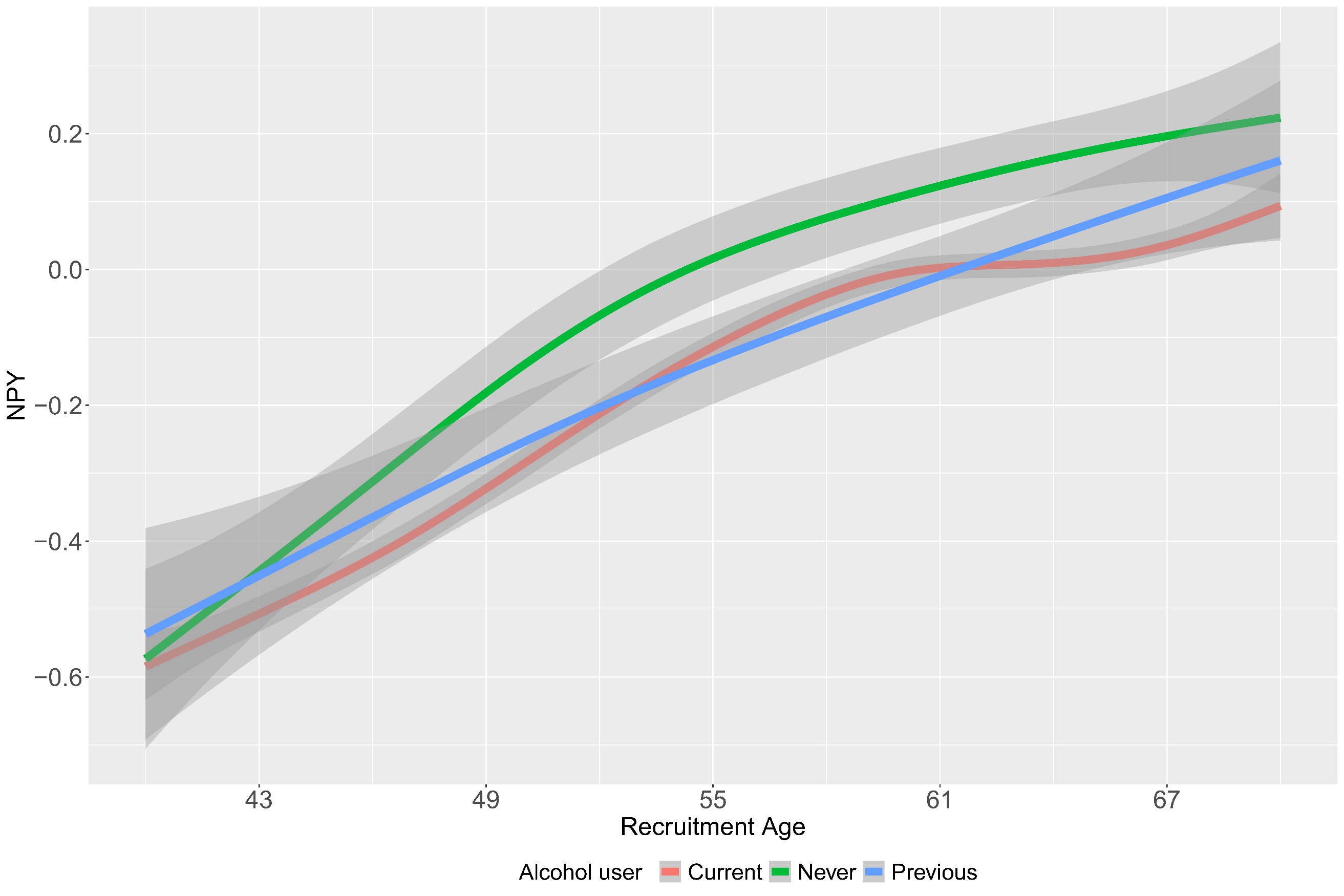

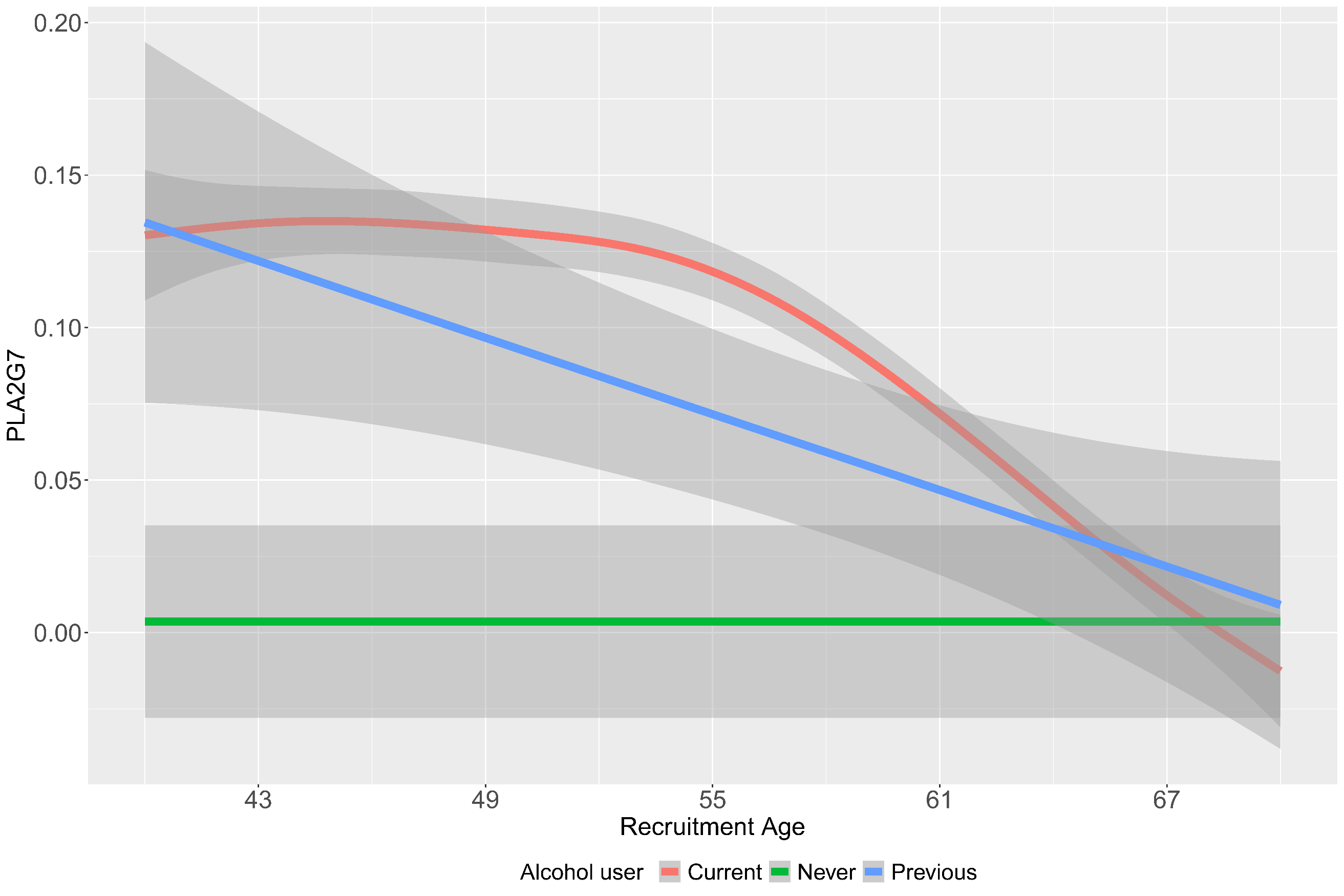

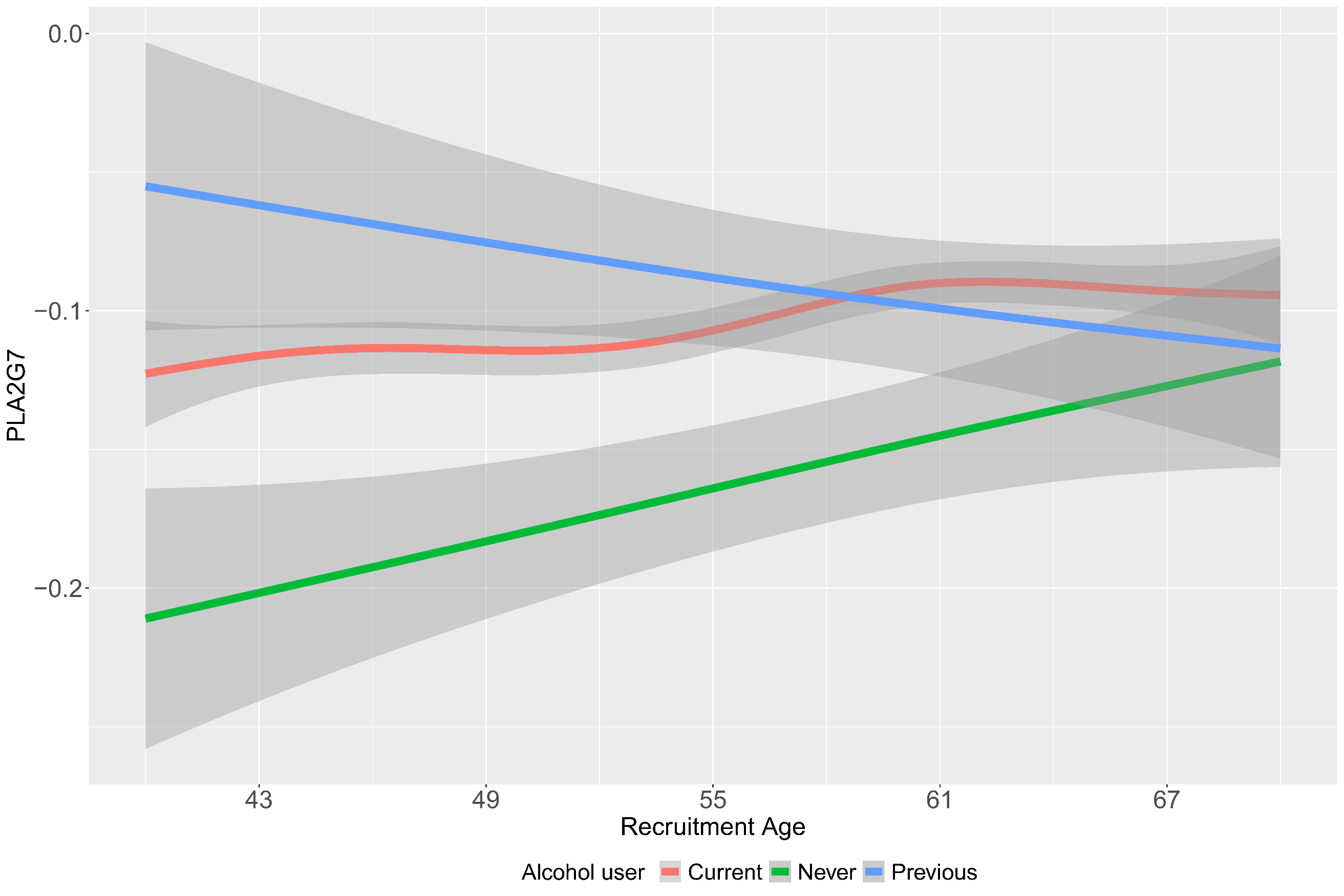

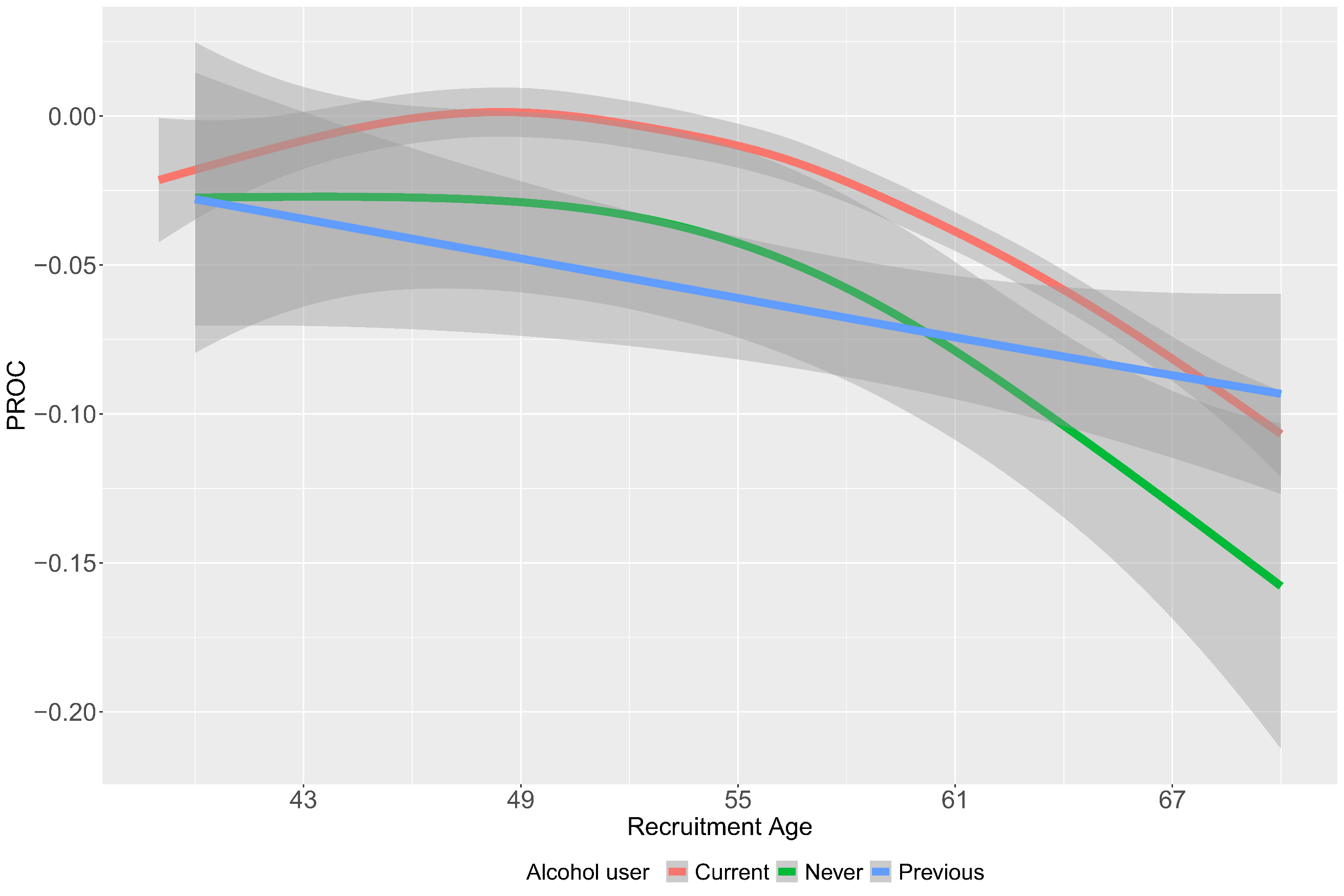

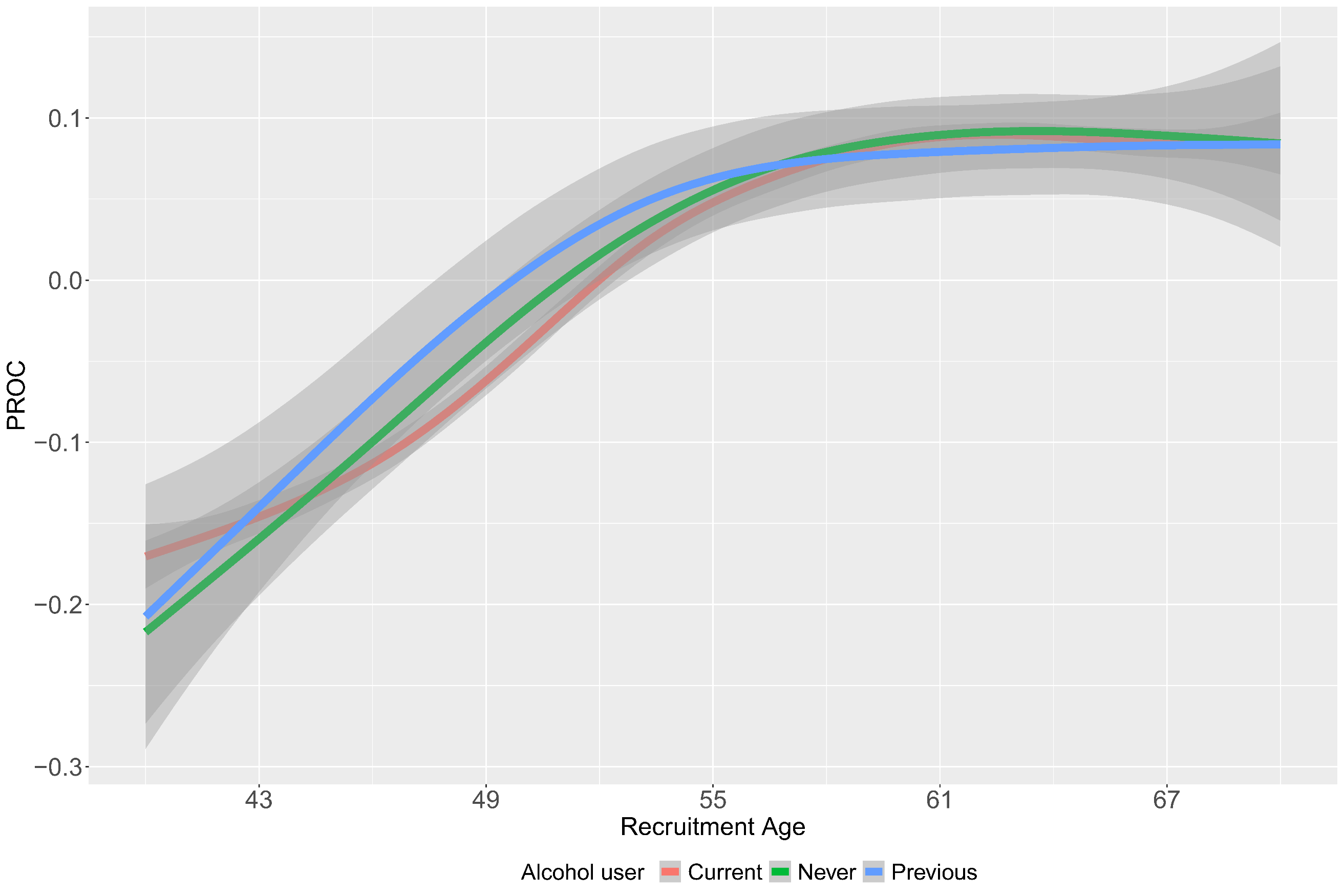

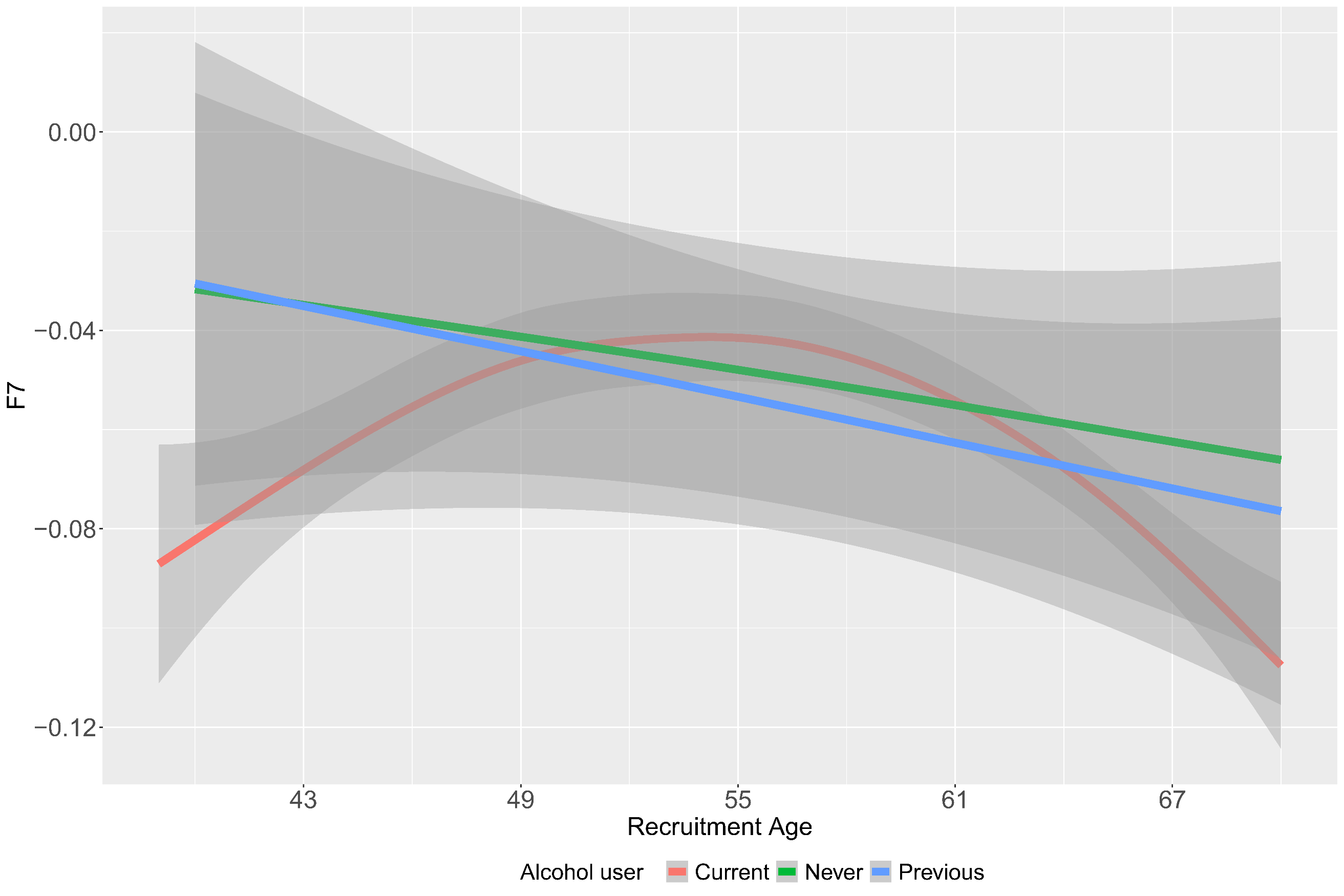

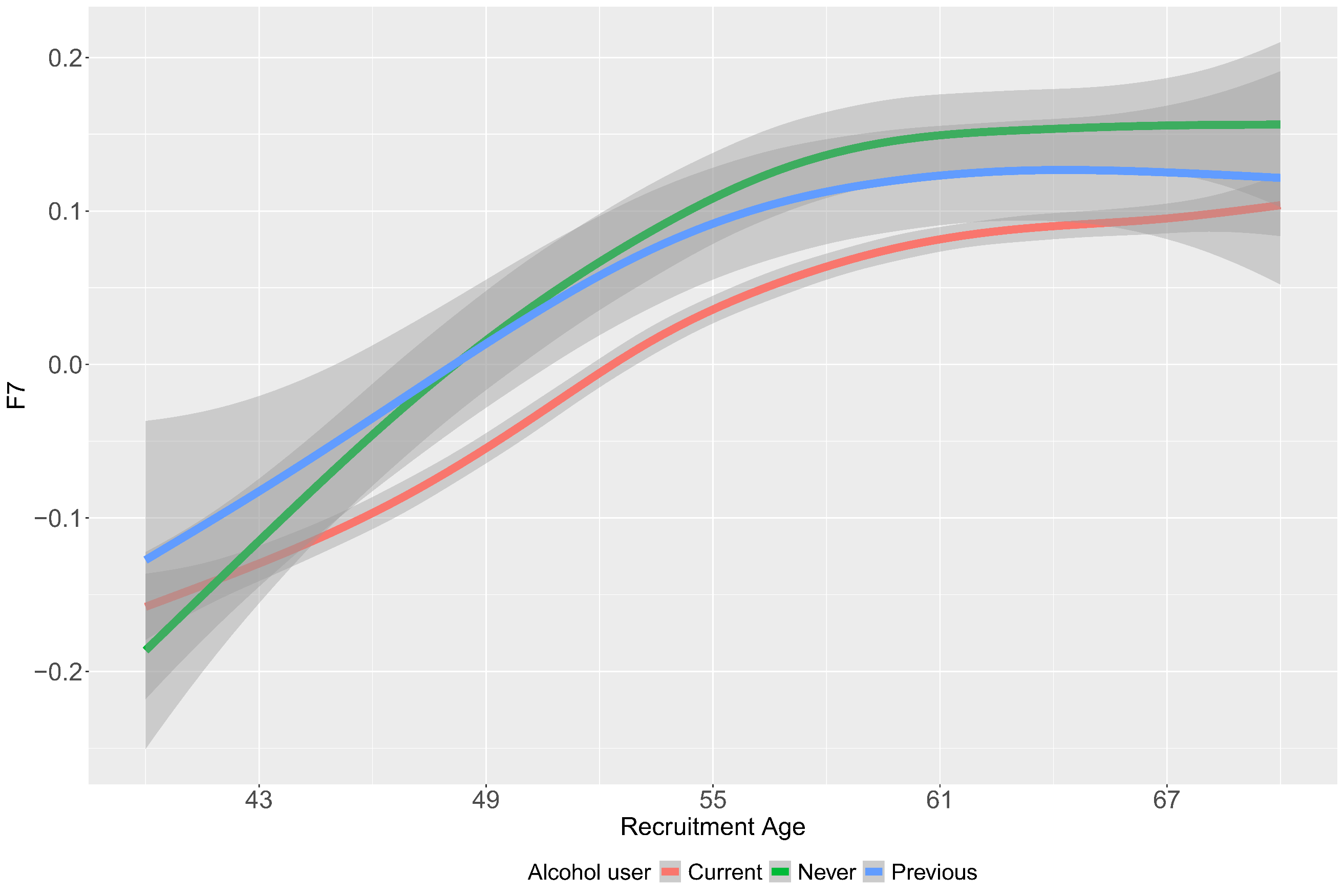

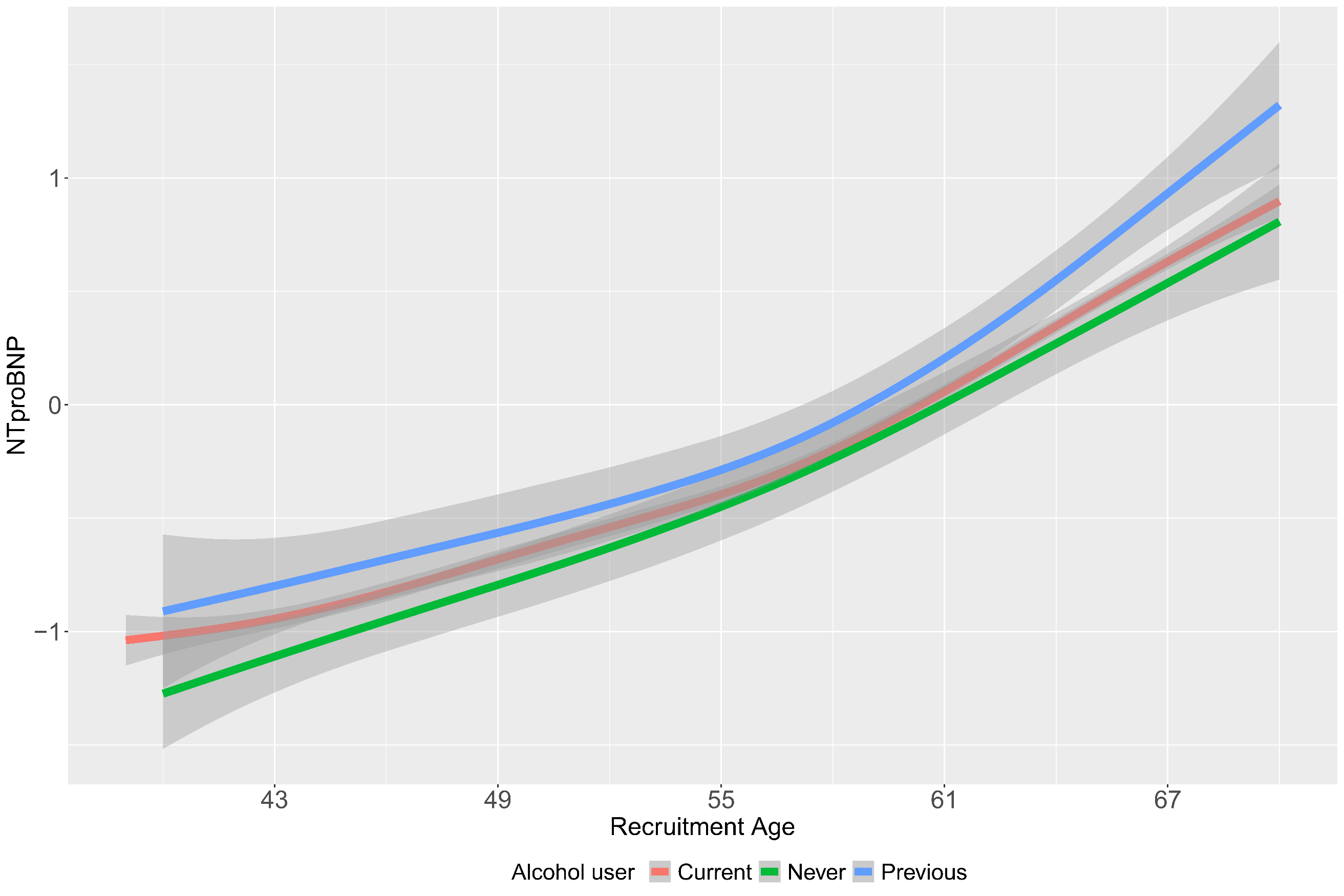

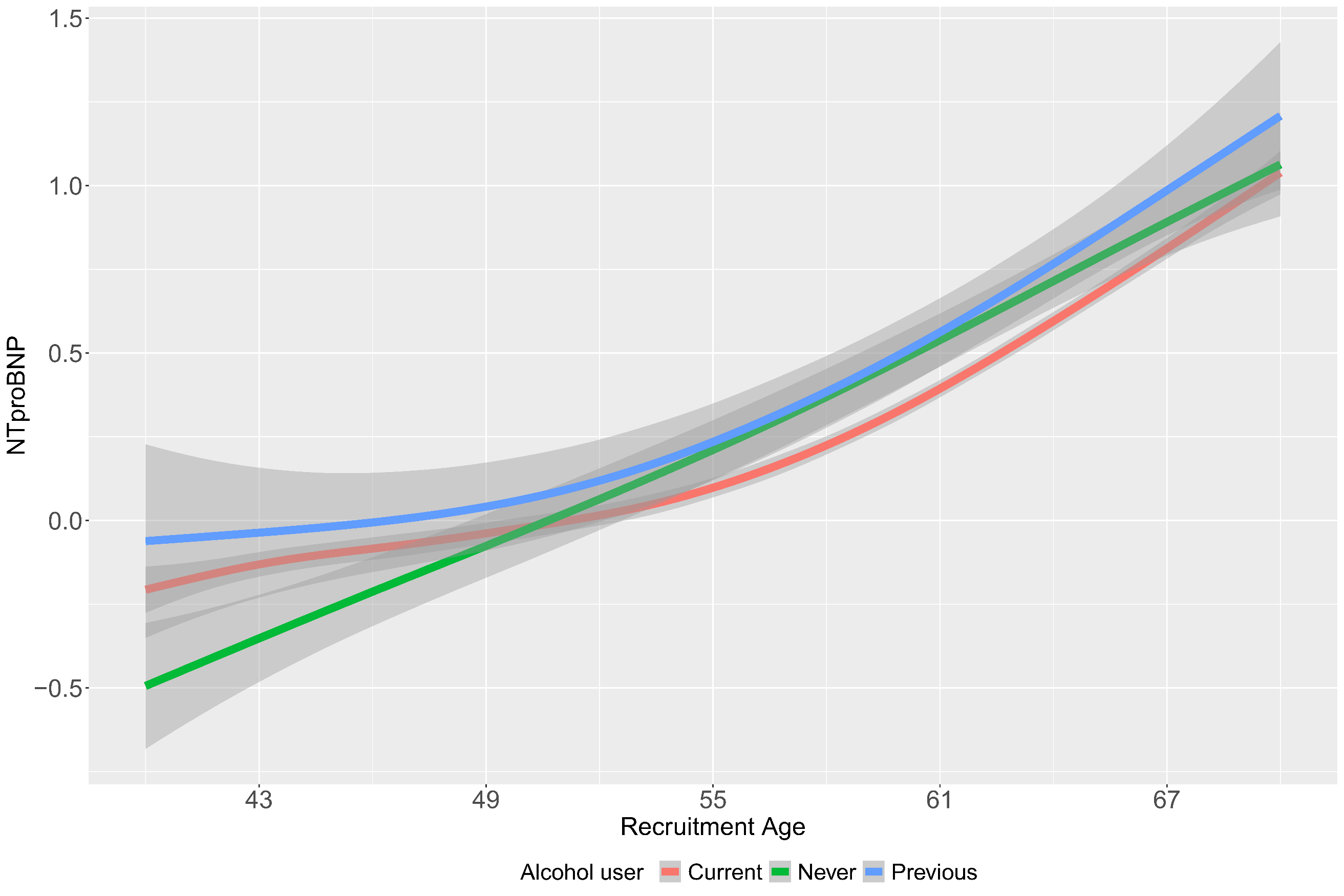

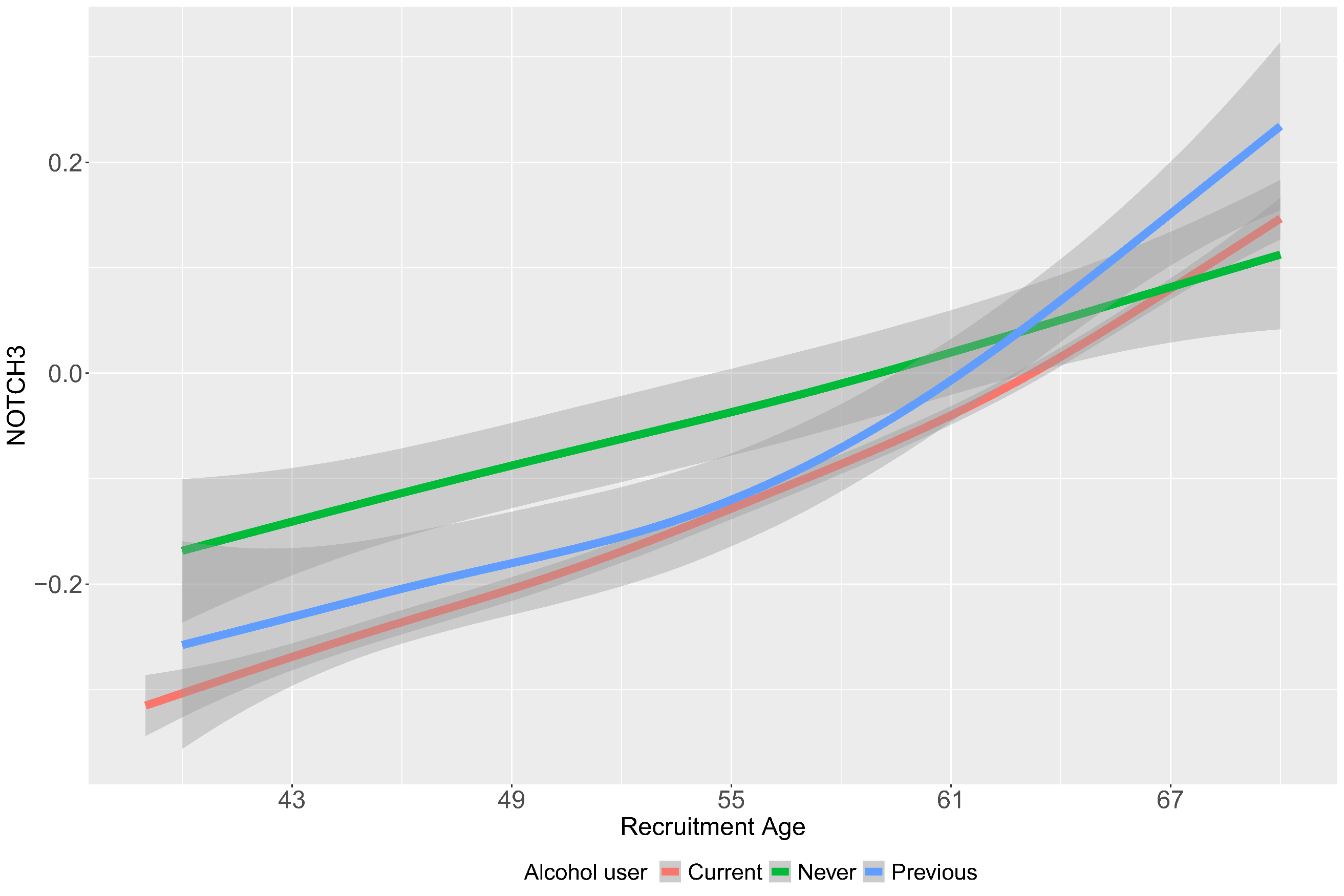

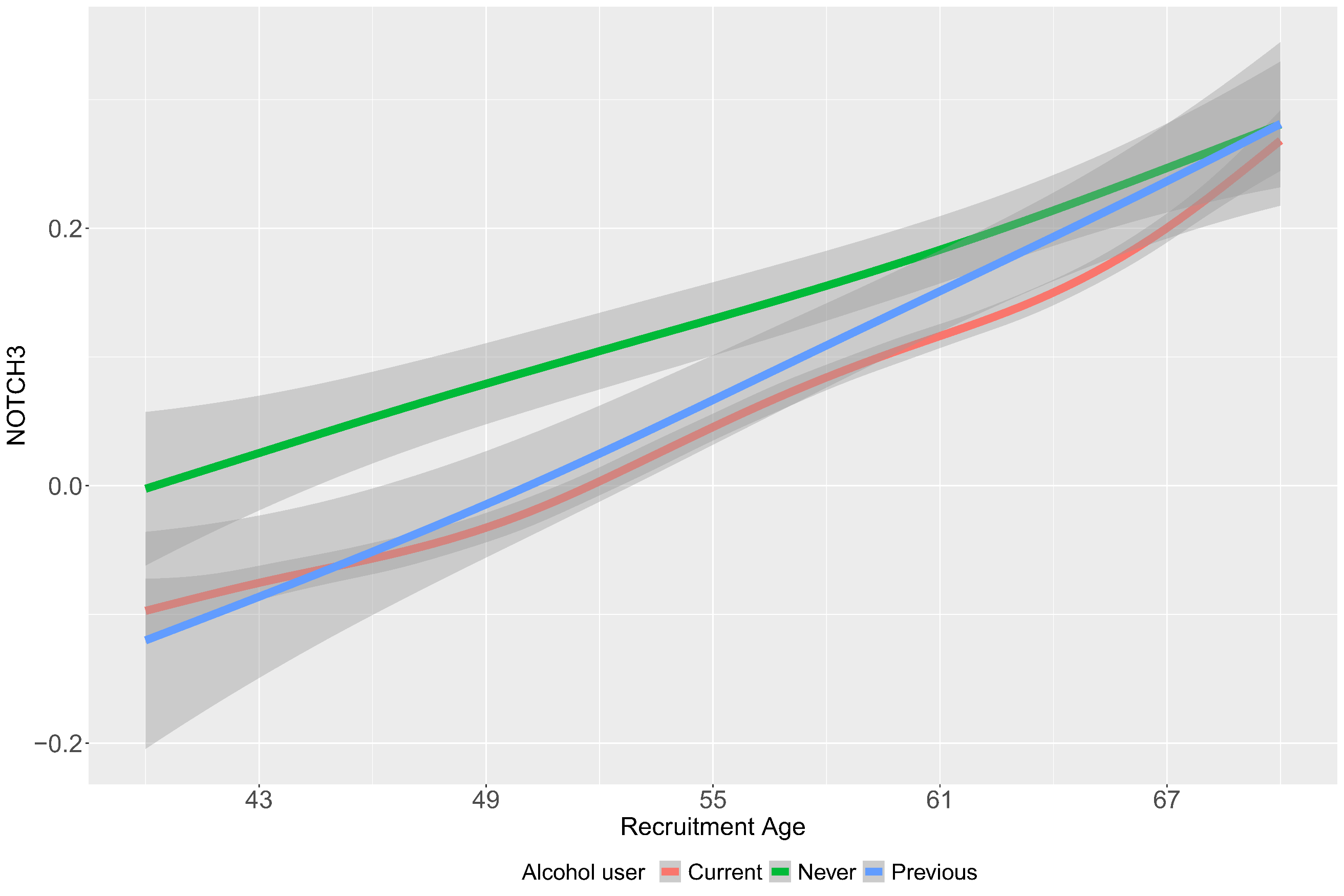

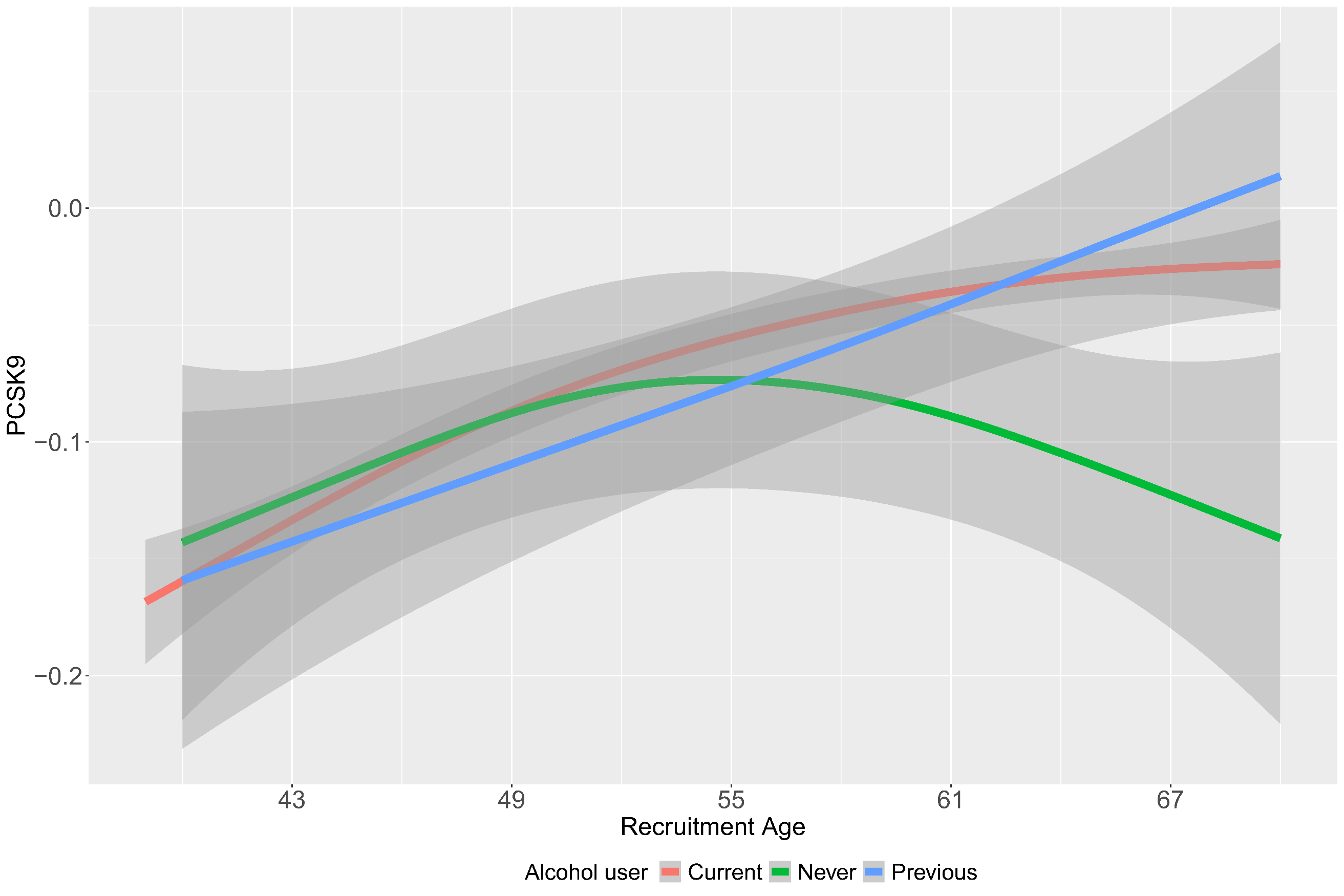

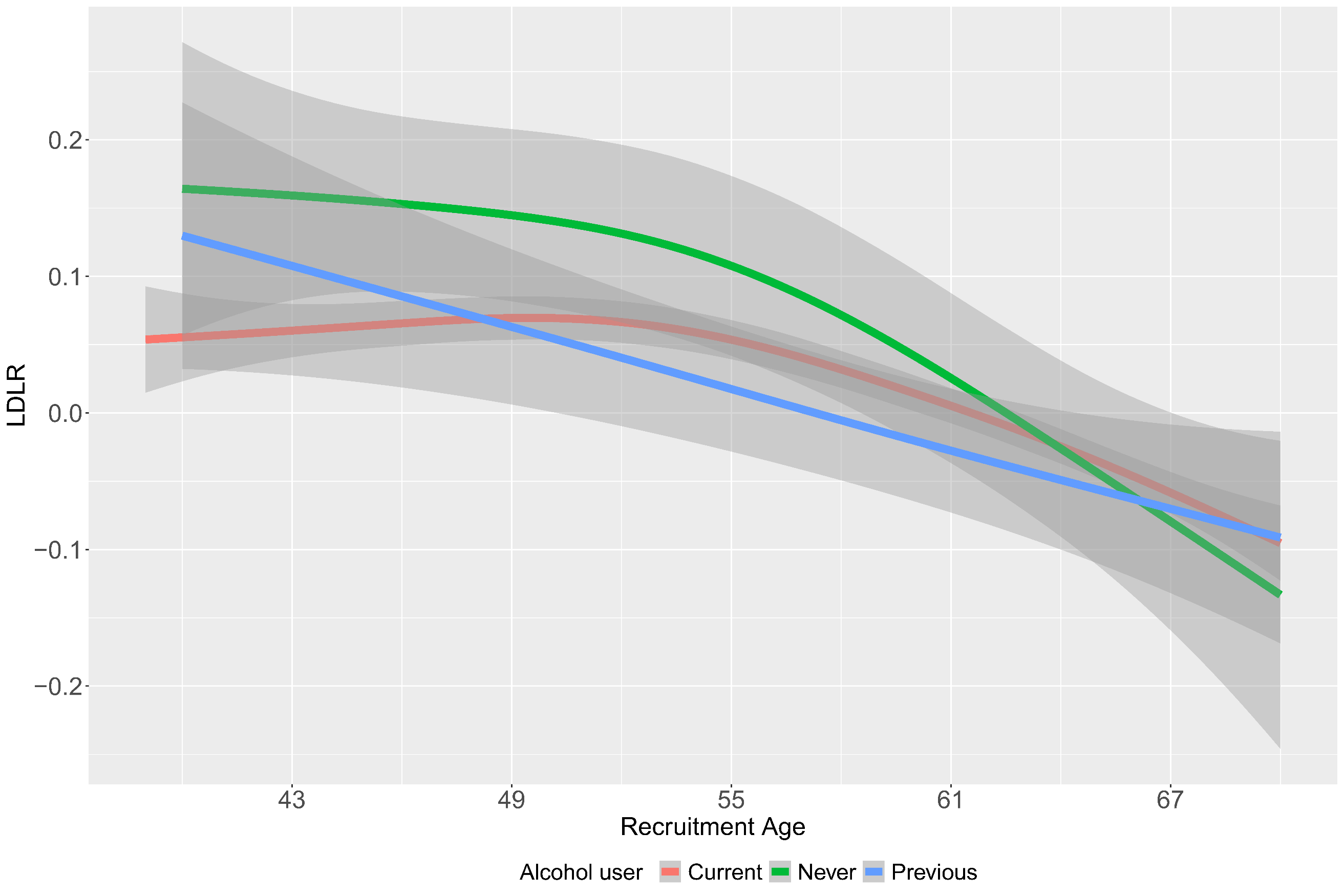

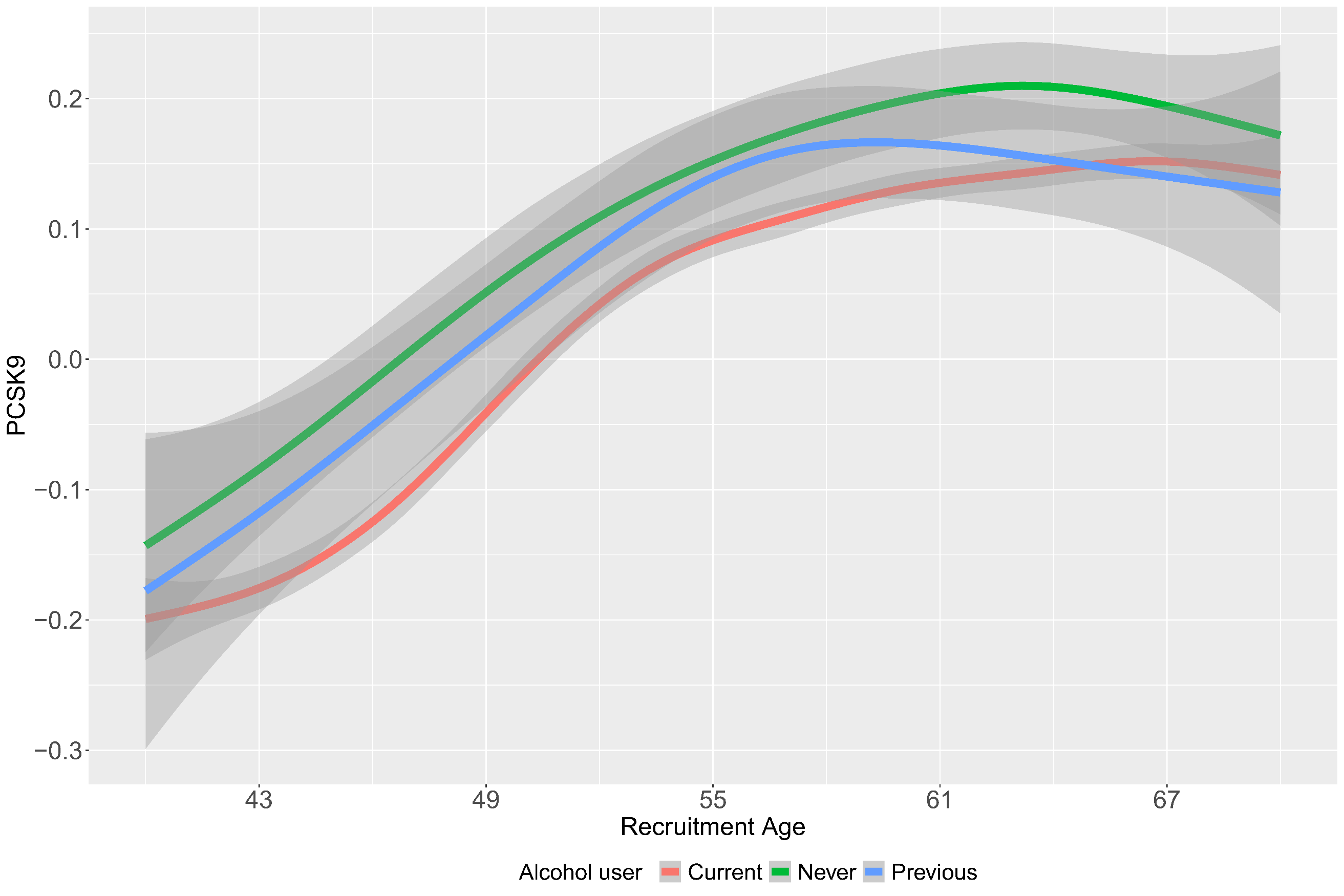

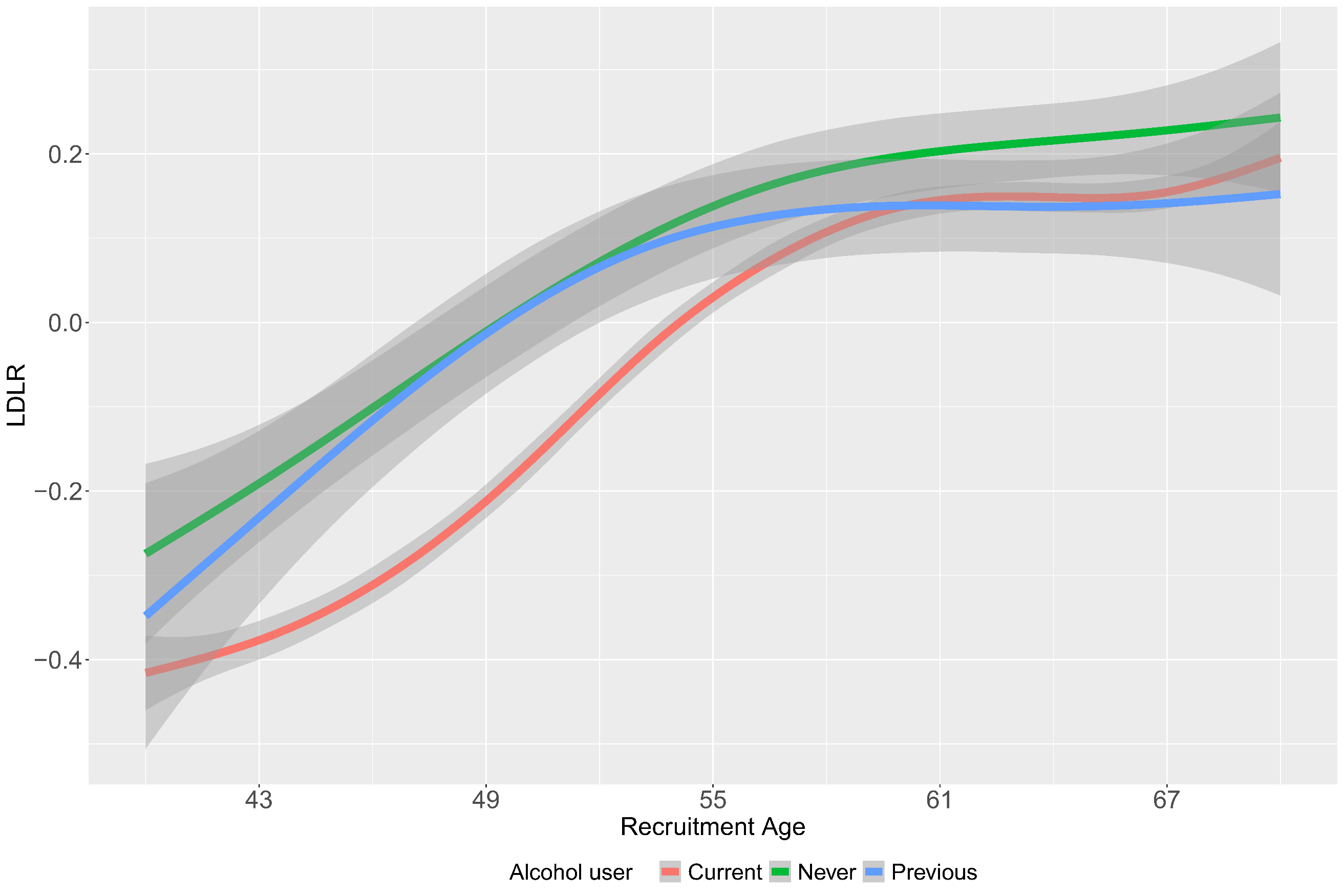

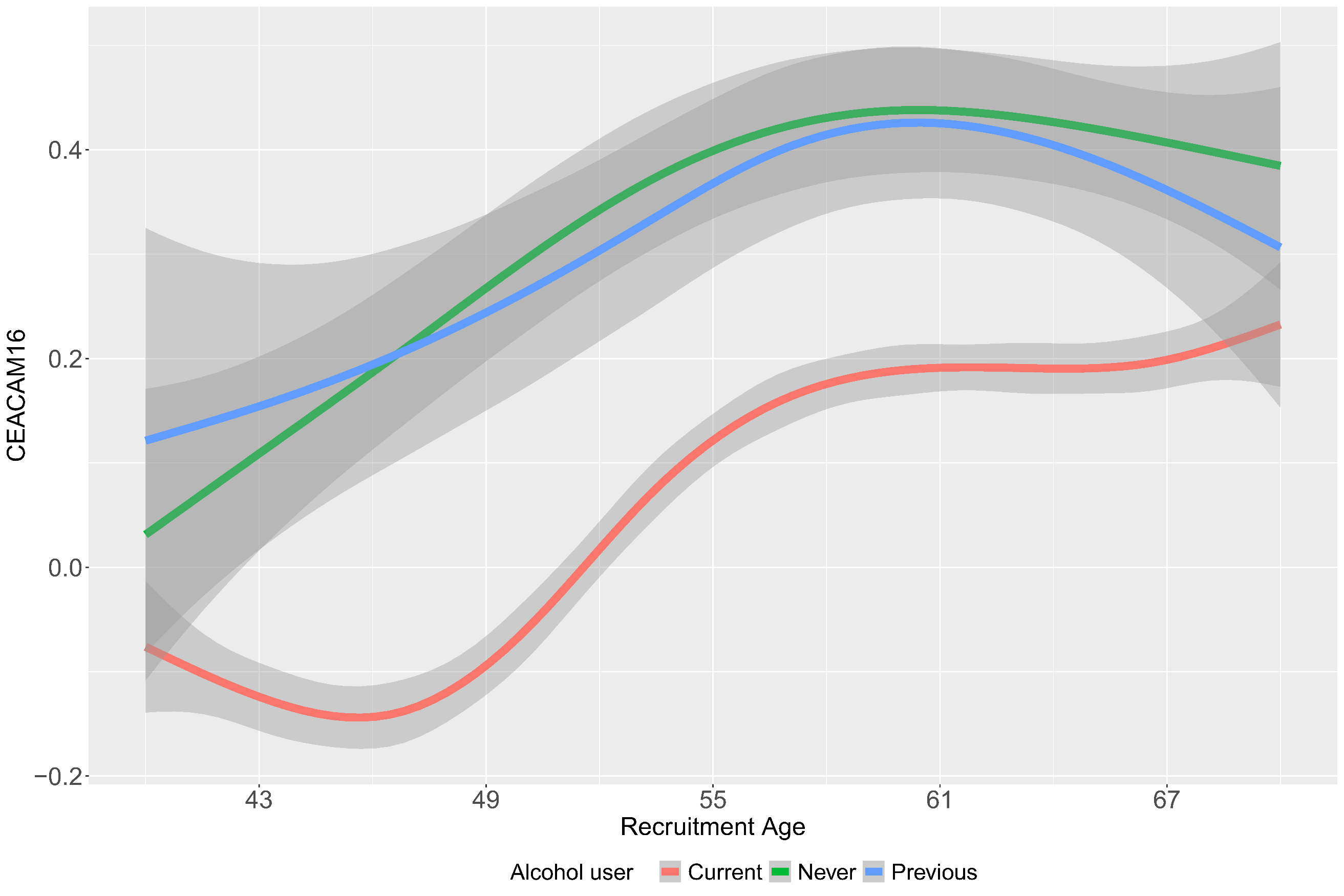

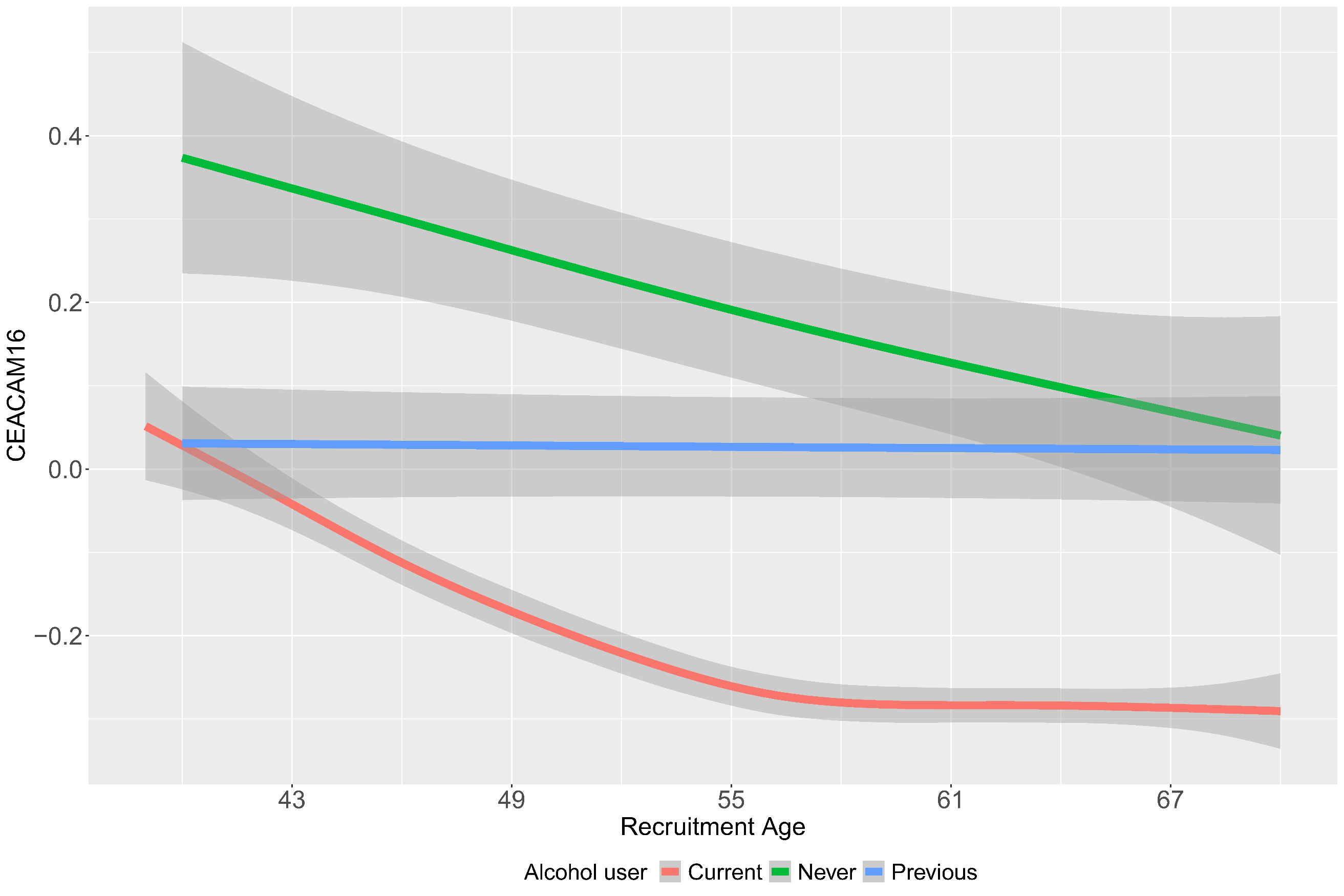

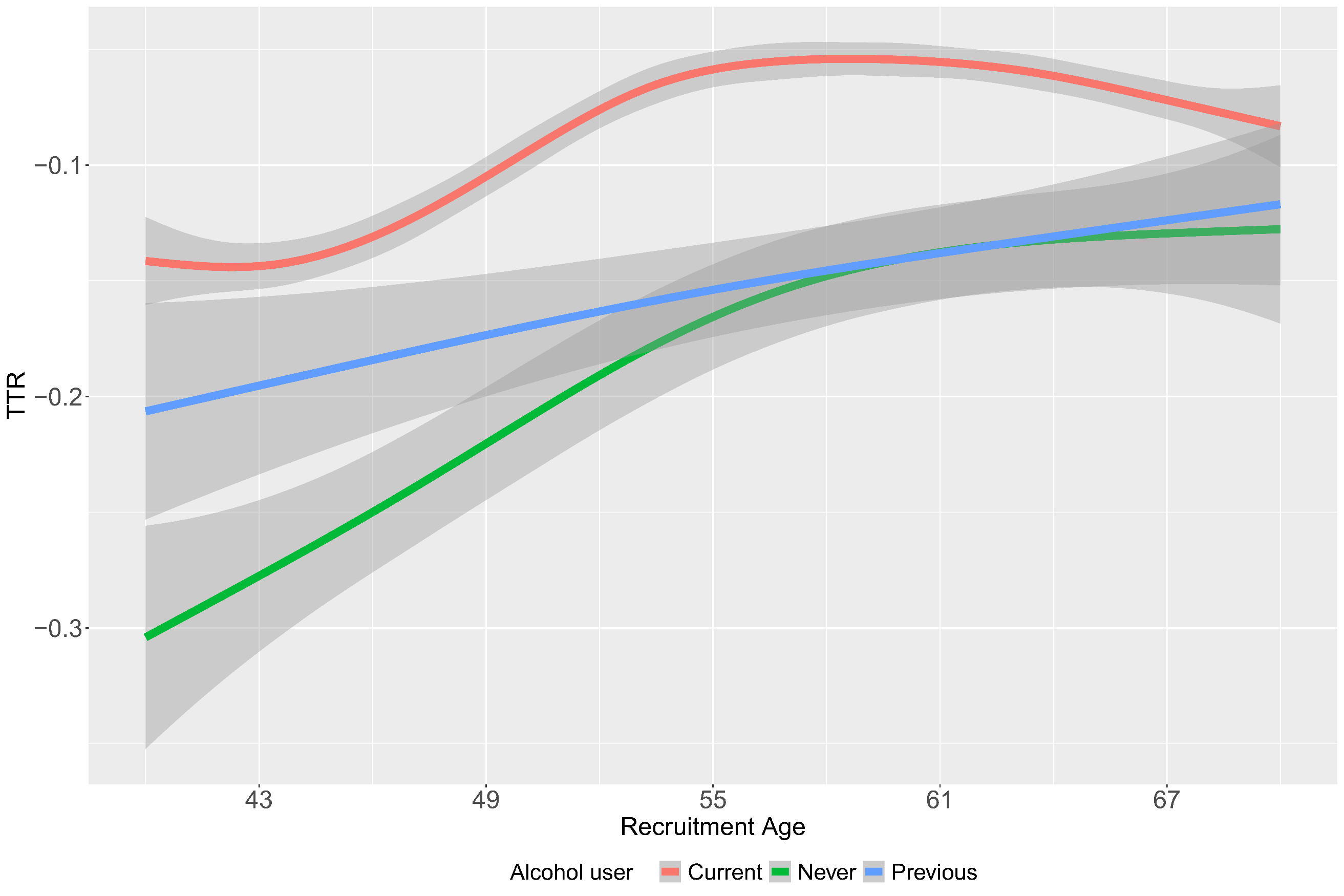

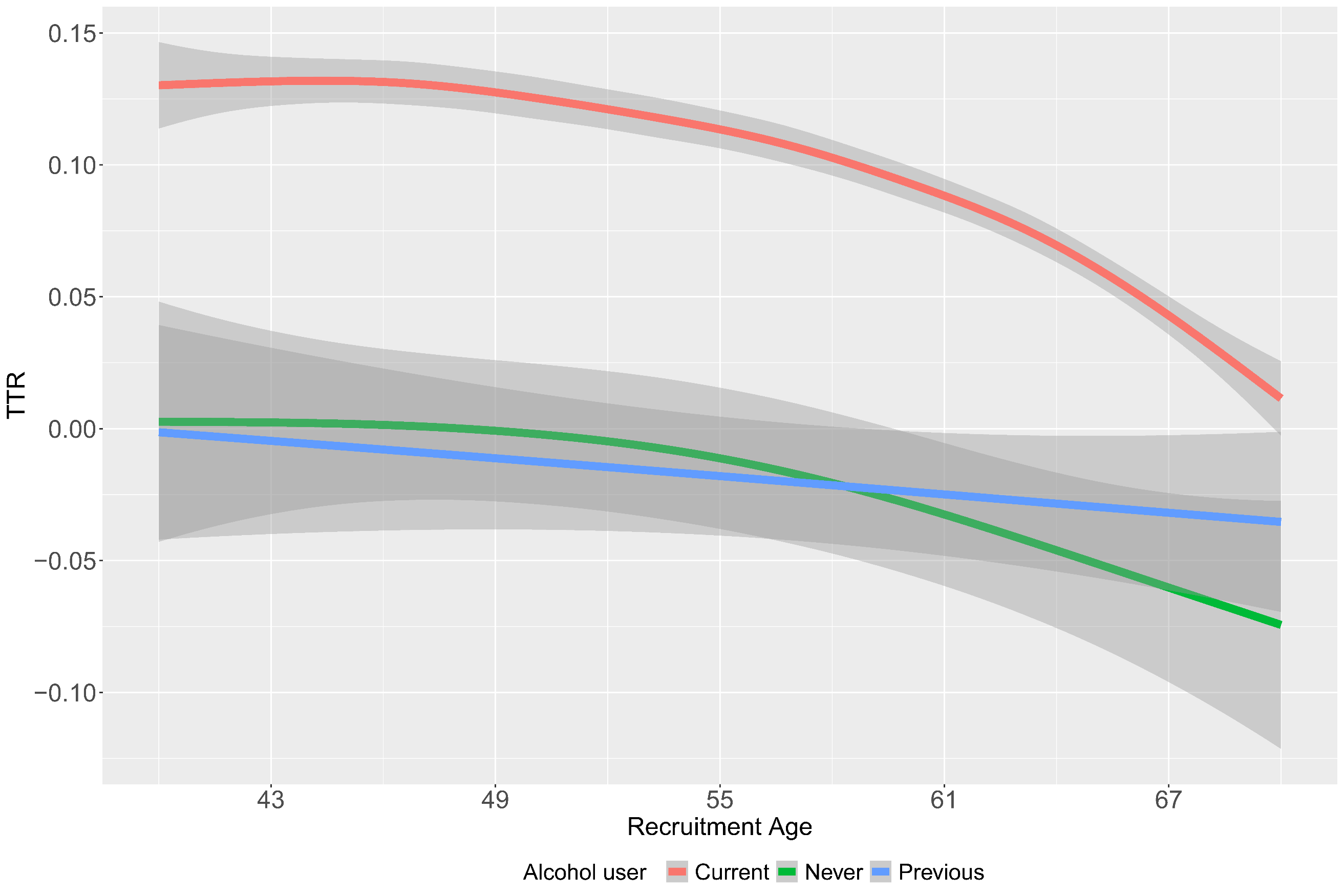

**Supplementary Figure 12**: Age distribution plot of the top aging related sex dimorphic proteins by alcohol use. Left panel depicts females and right panel depicts males.

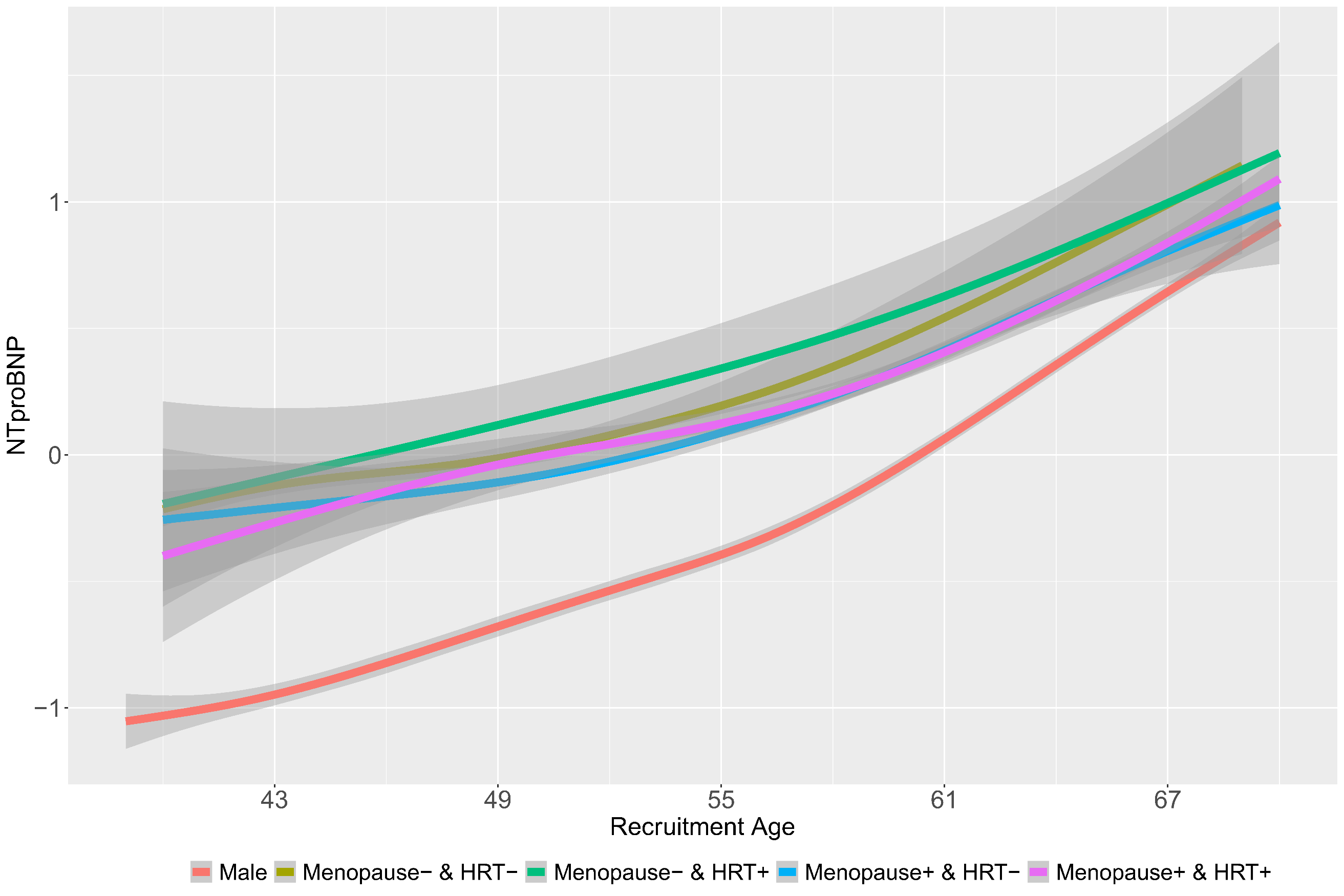

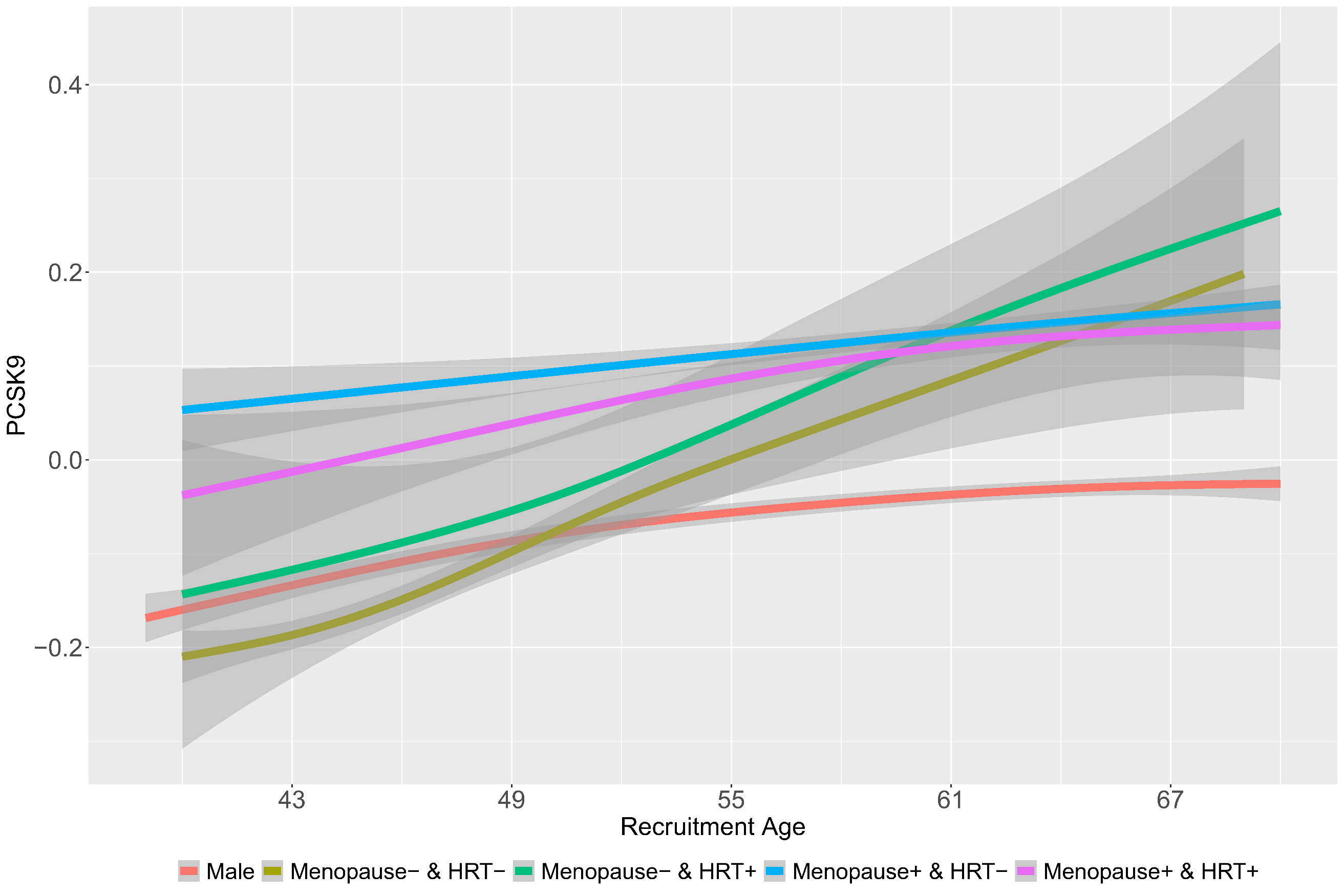

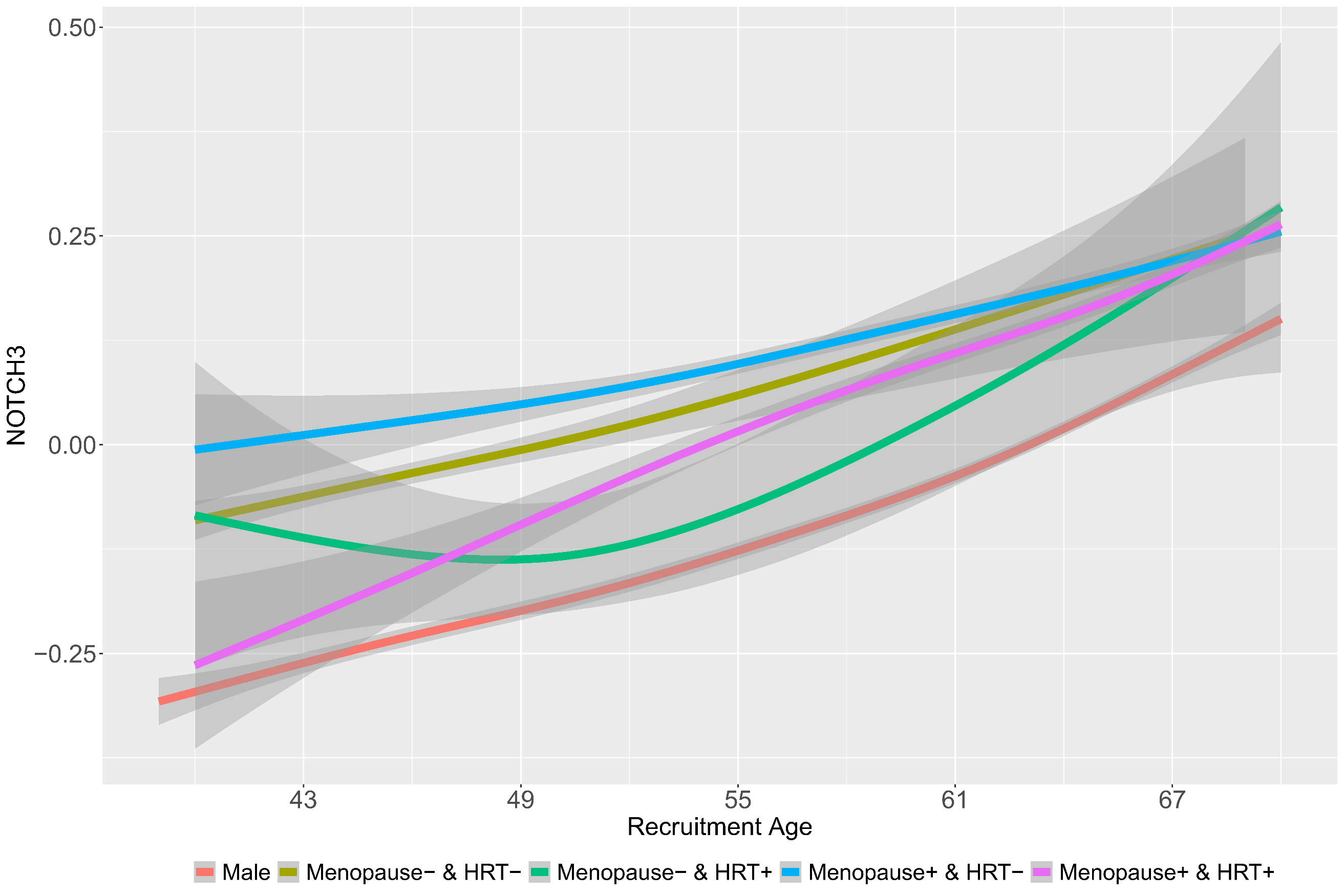

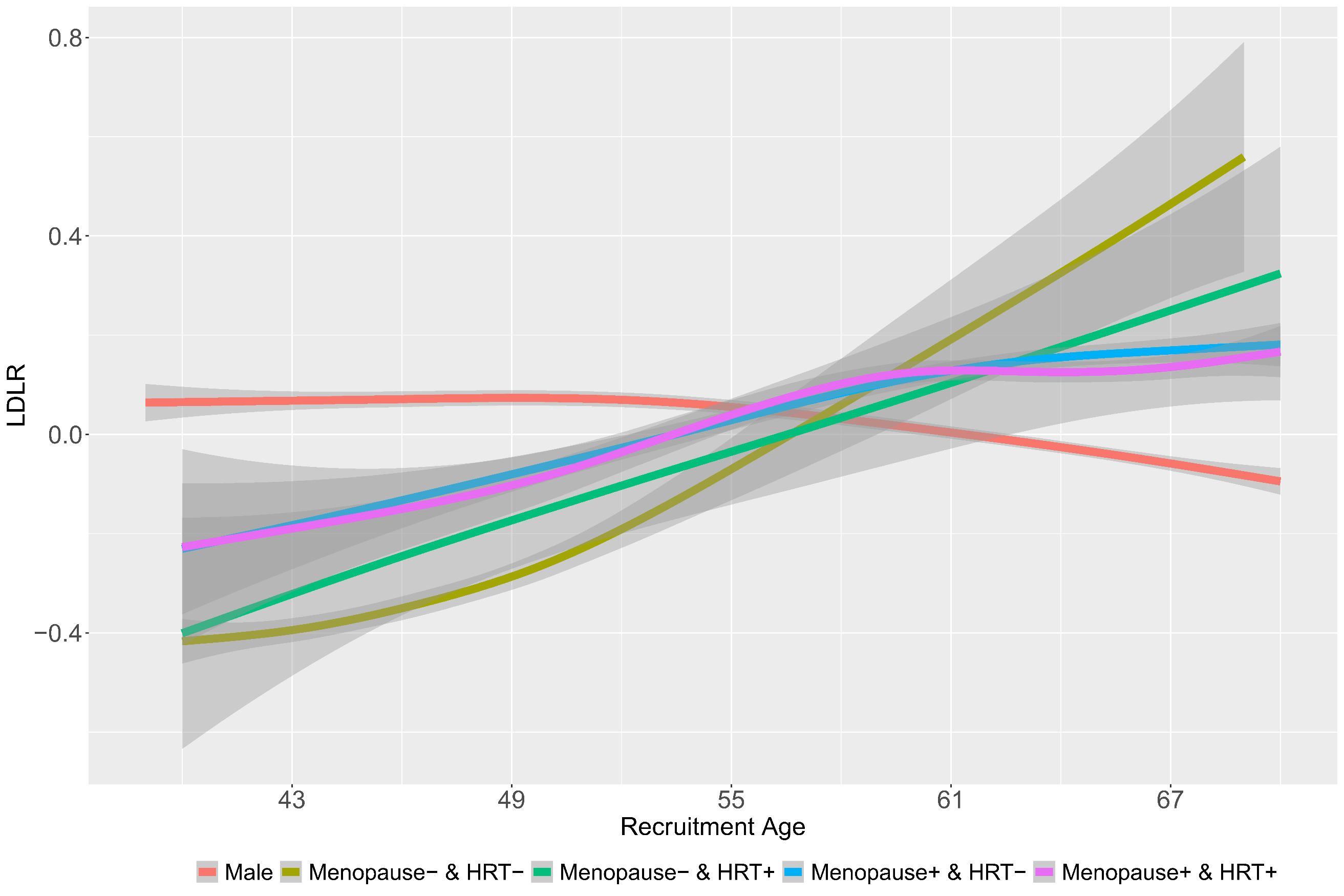

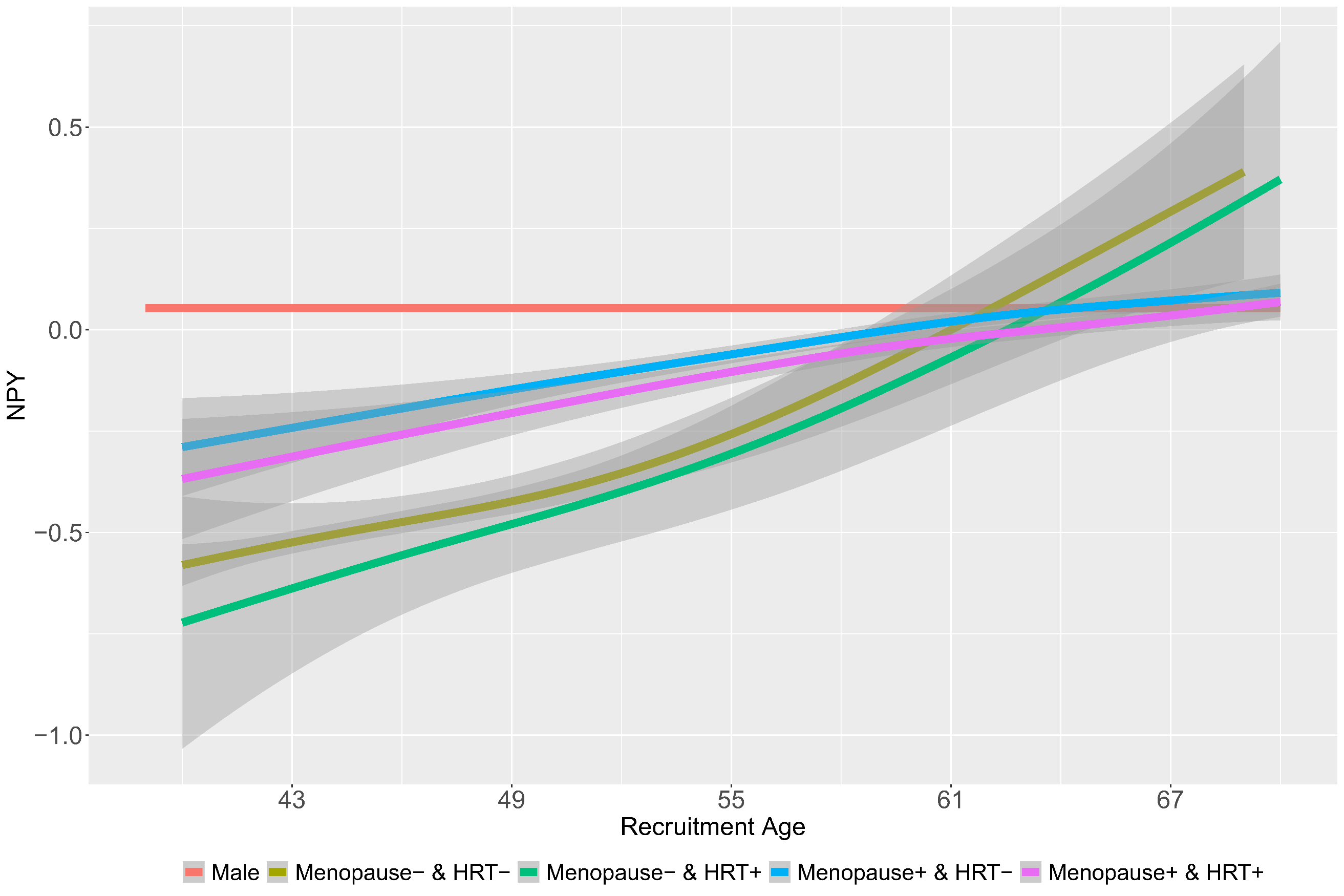

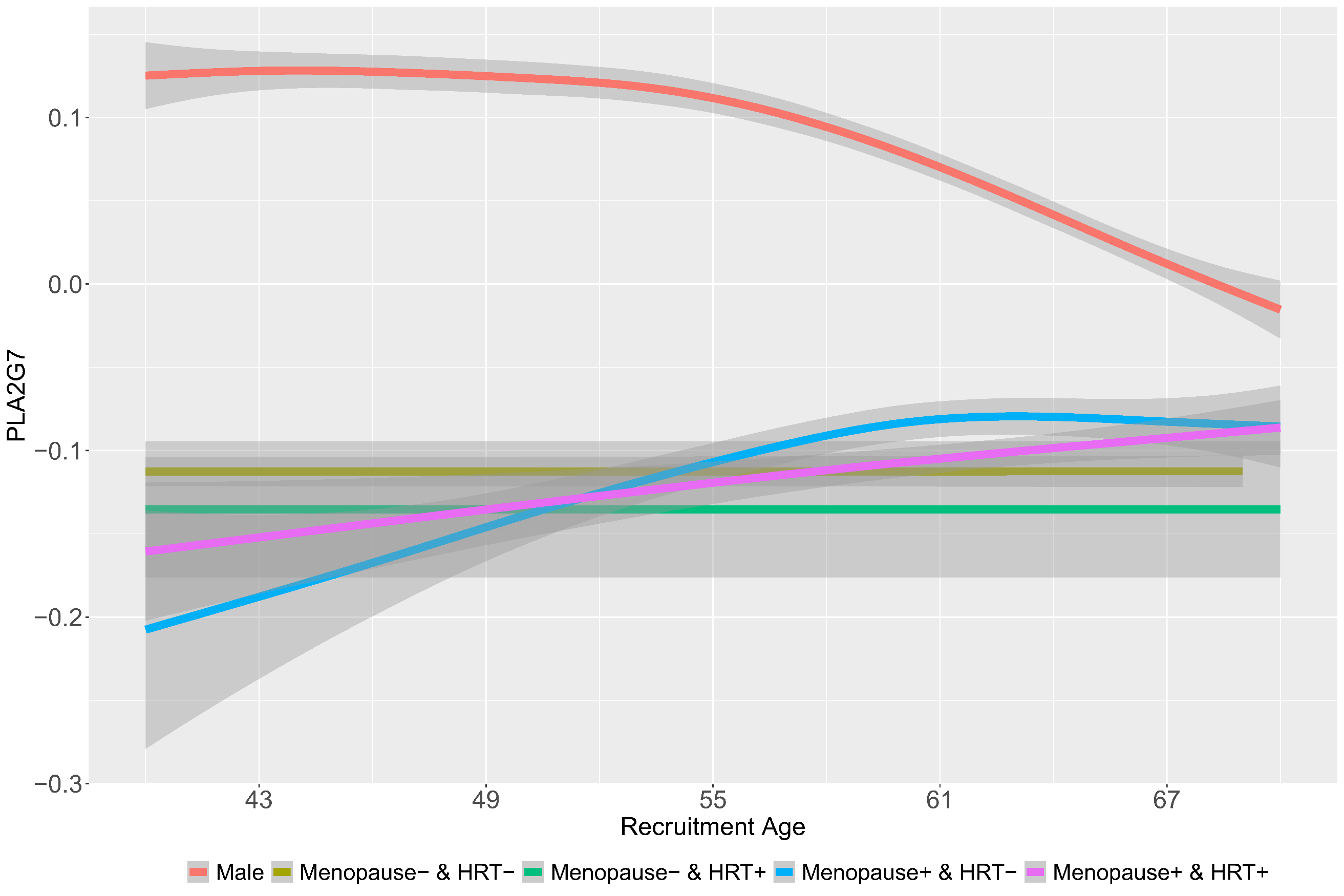

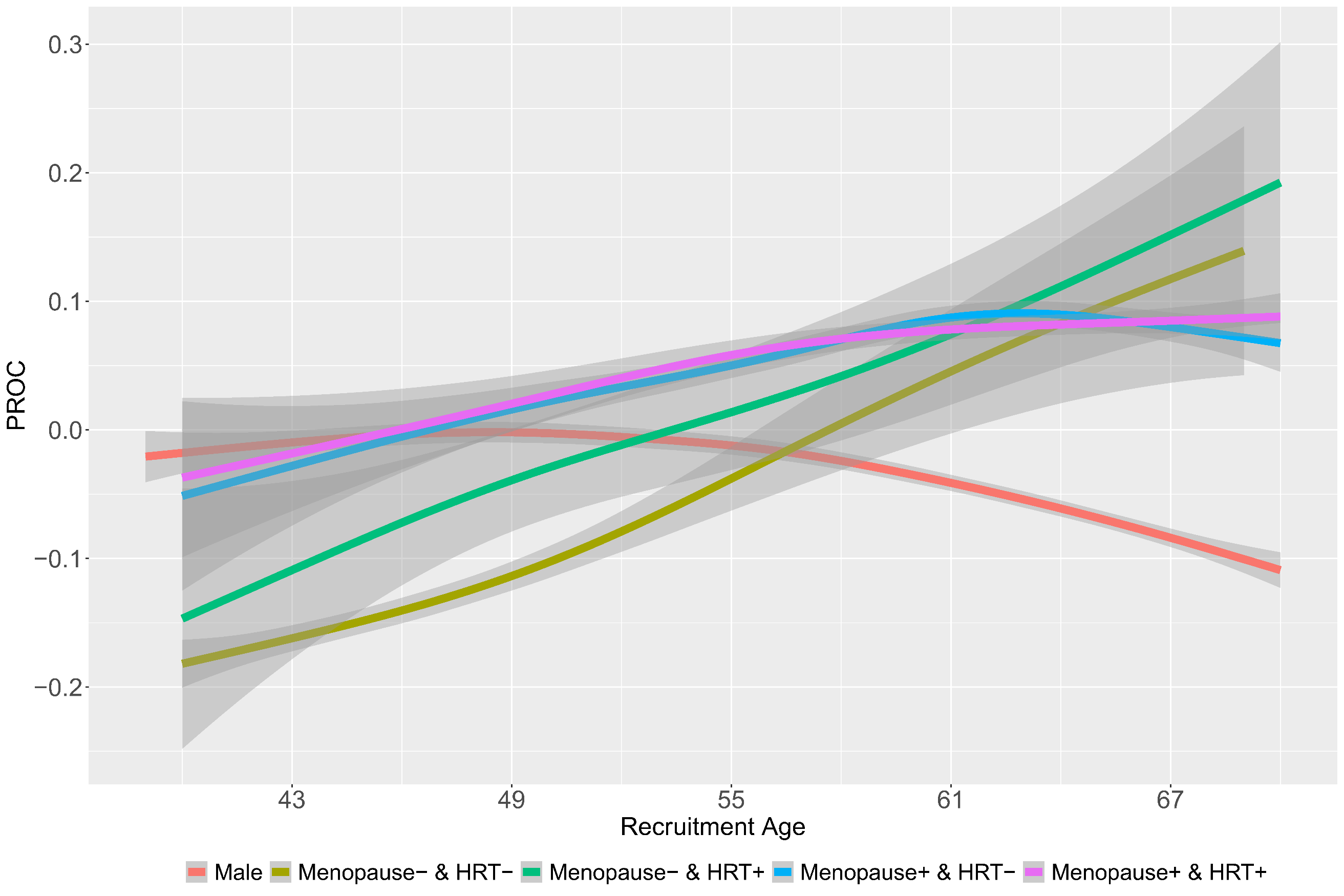

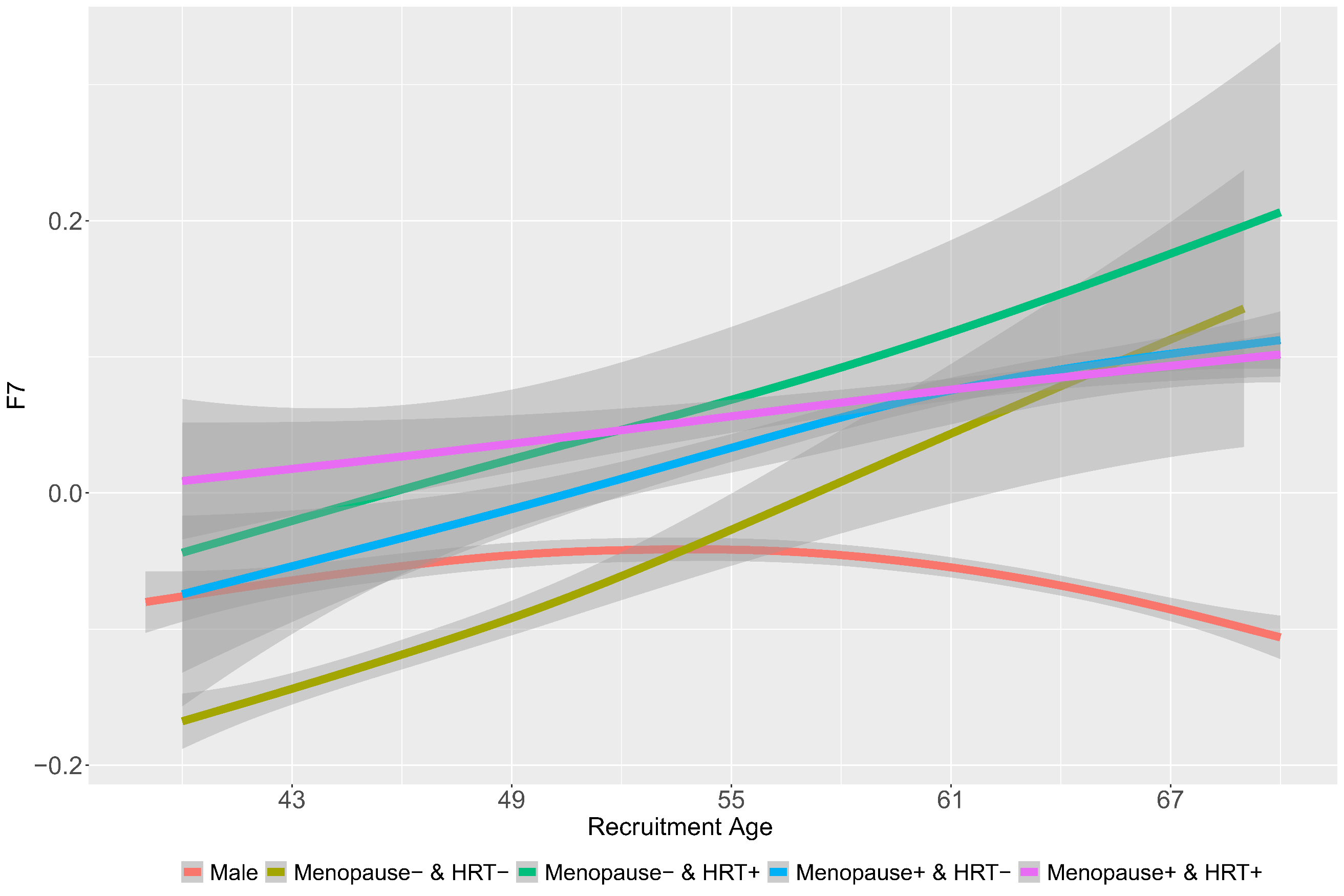

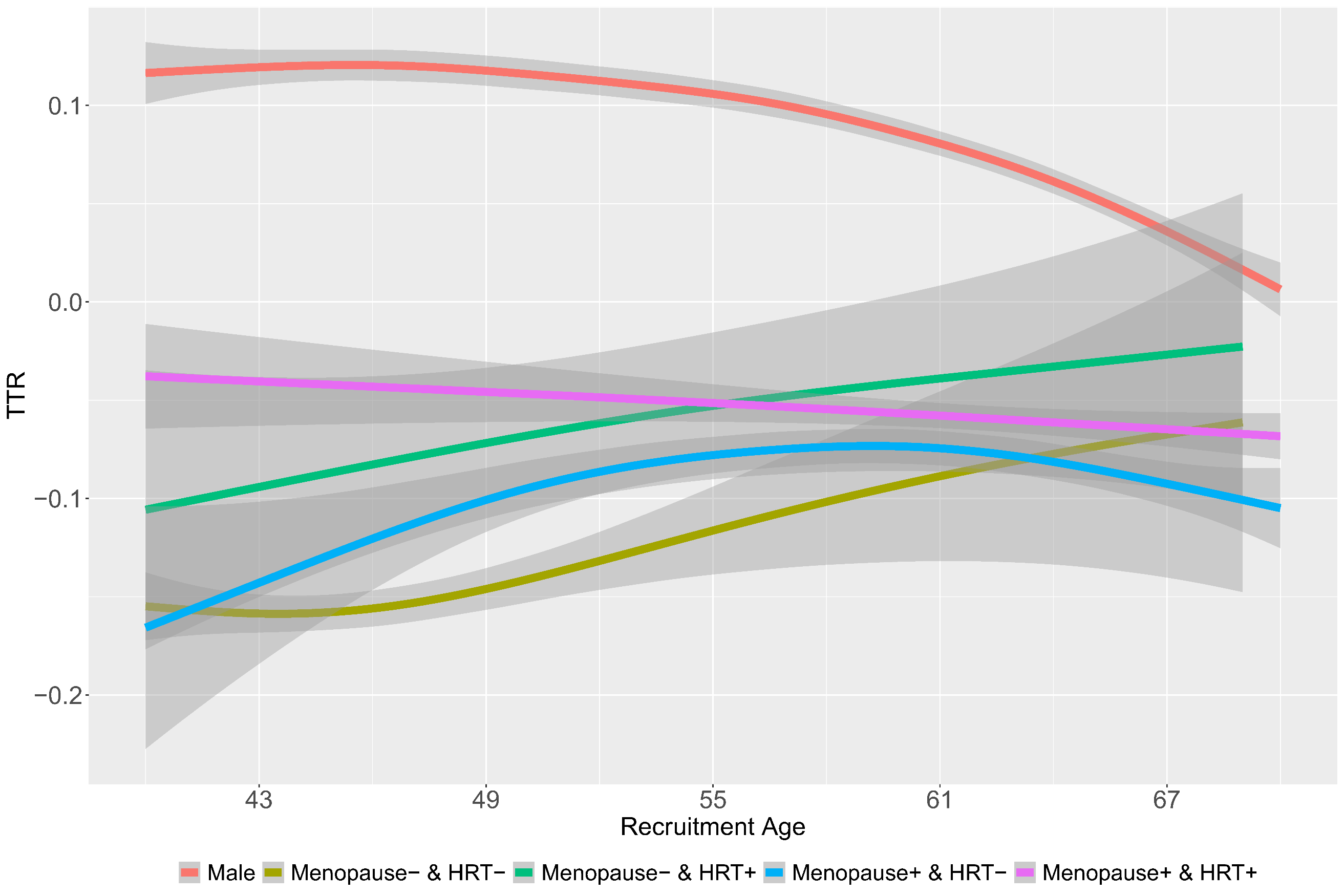

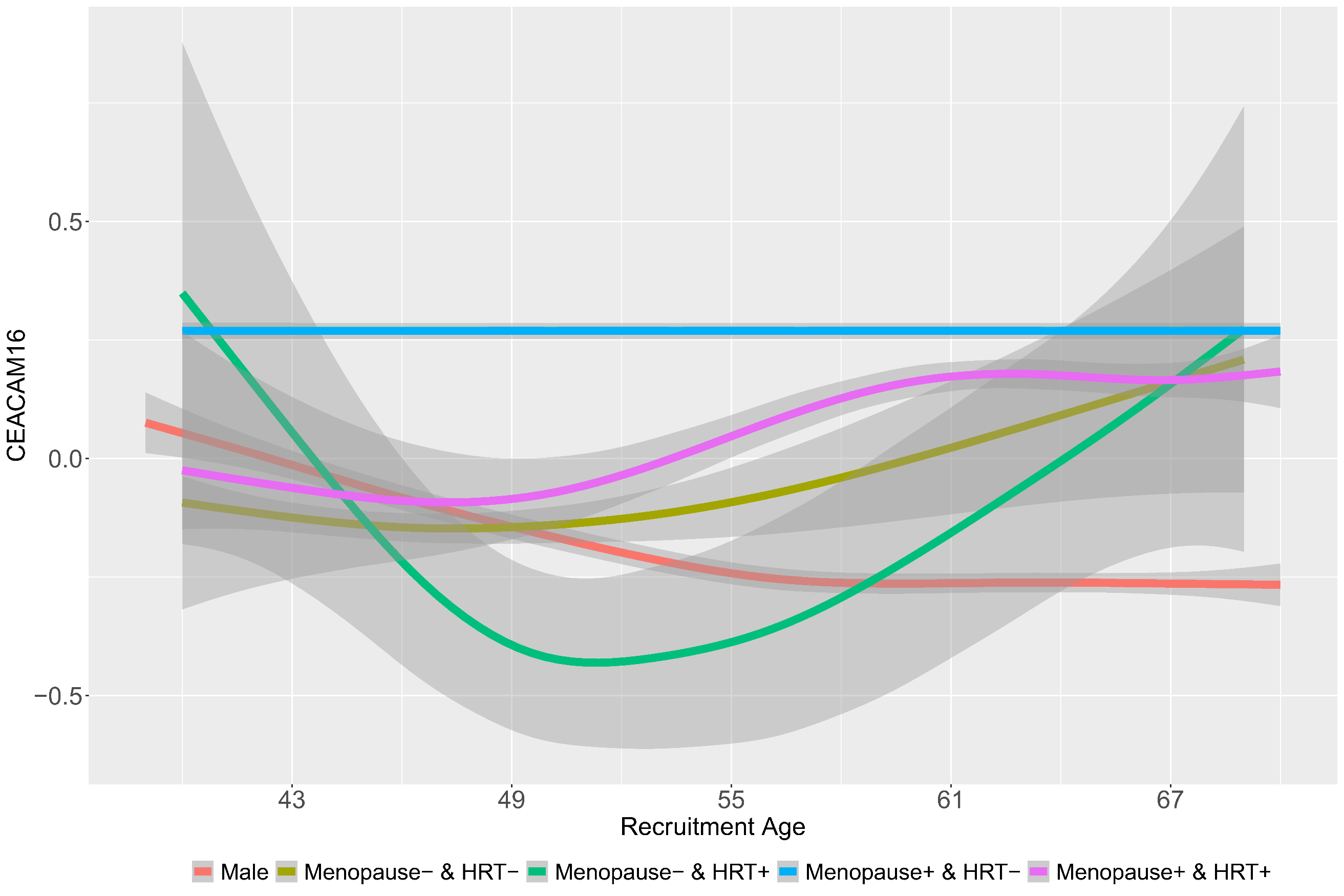

**Supplementary Figure 13**: Age distribution plot of the top aging related sex dimorphic proteins by menopause and hormone replacement therapy (HRT) use.

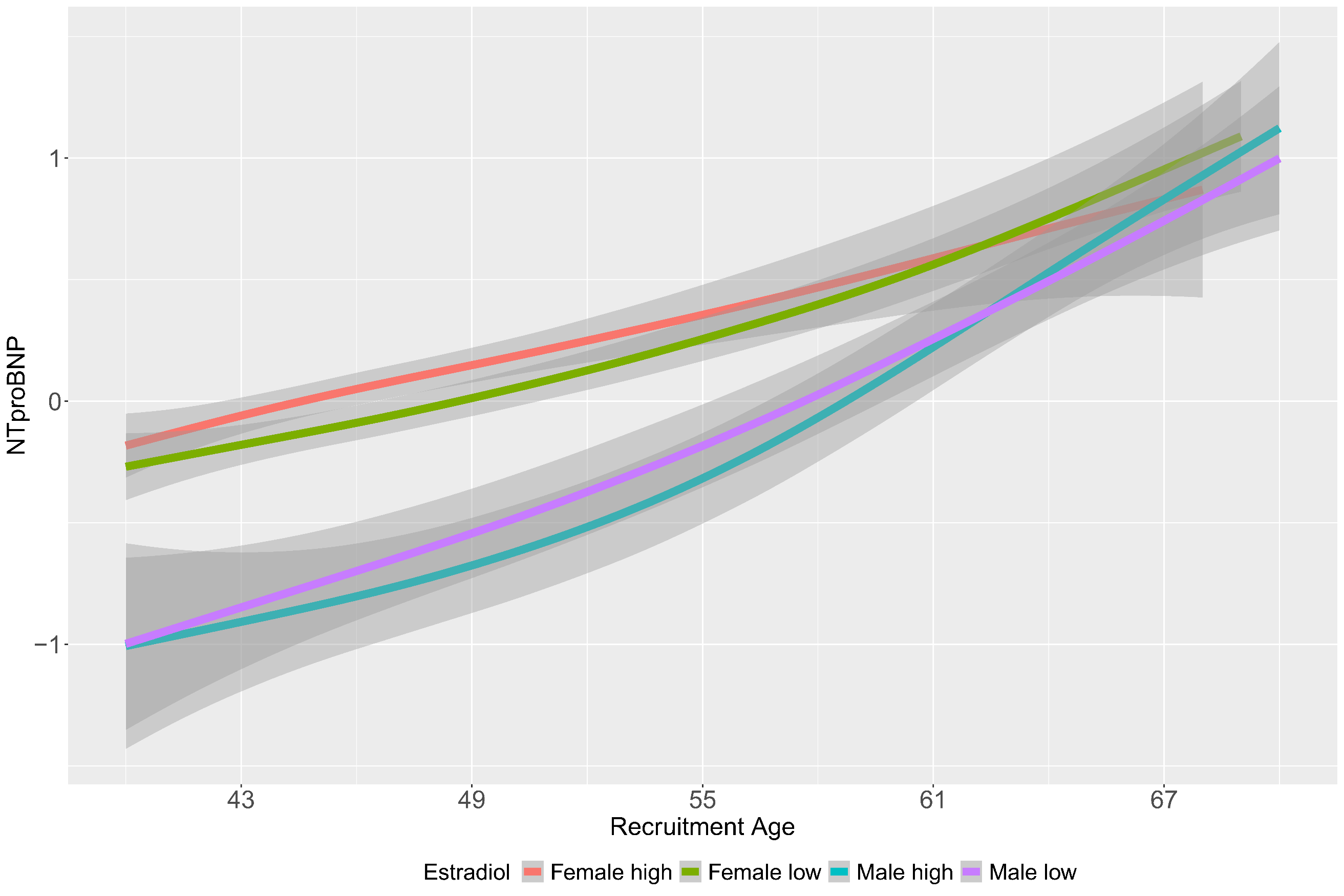

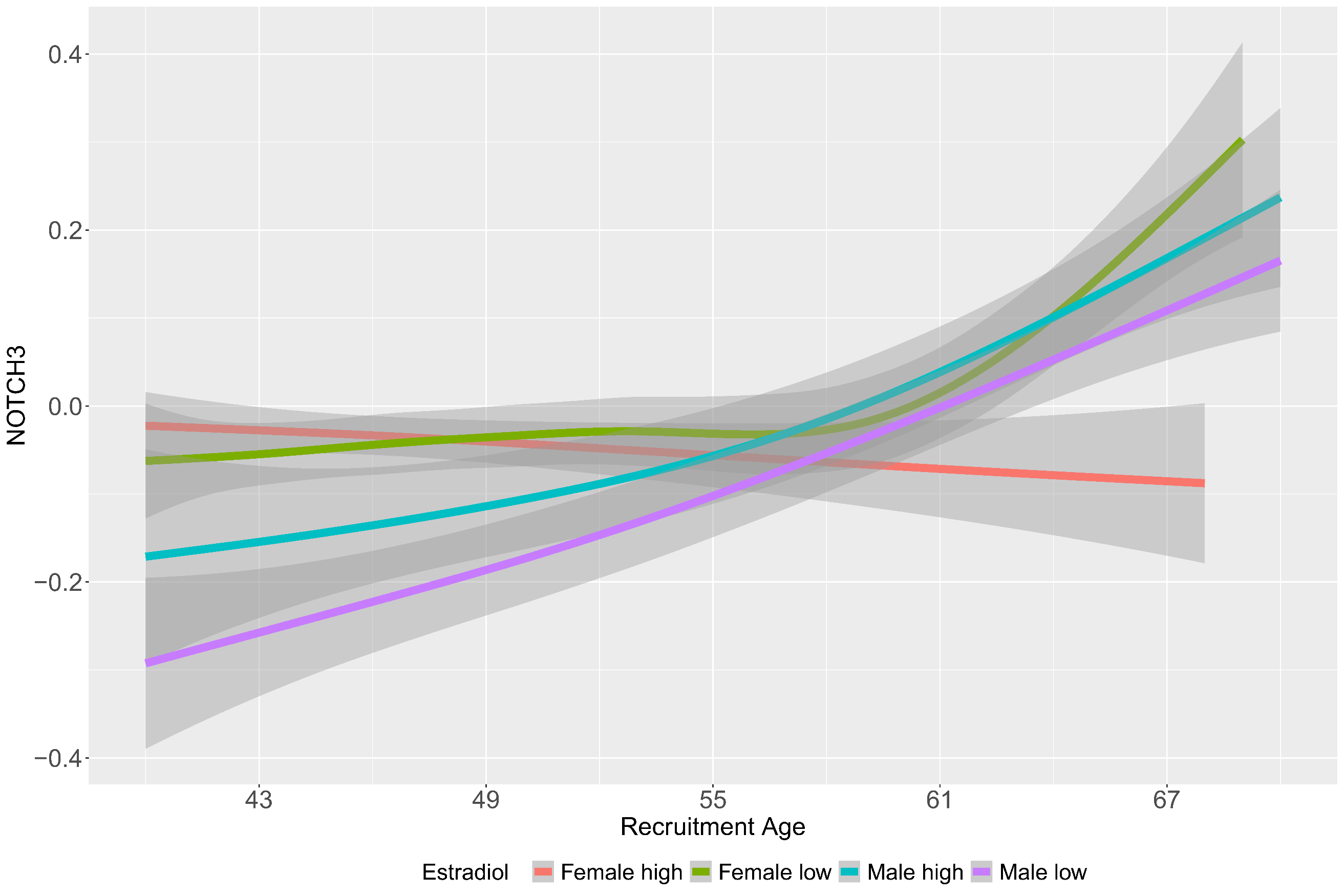

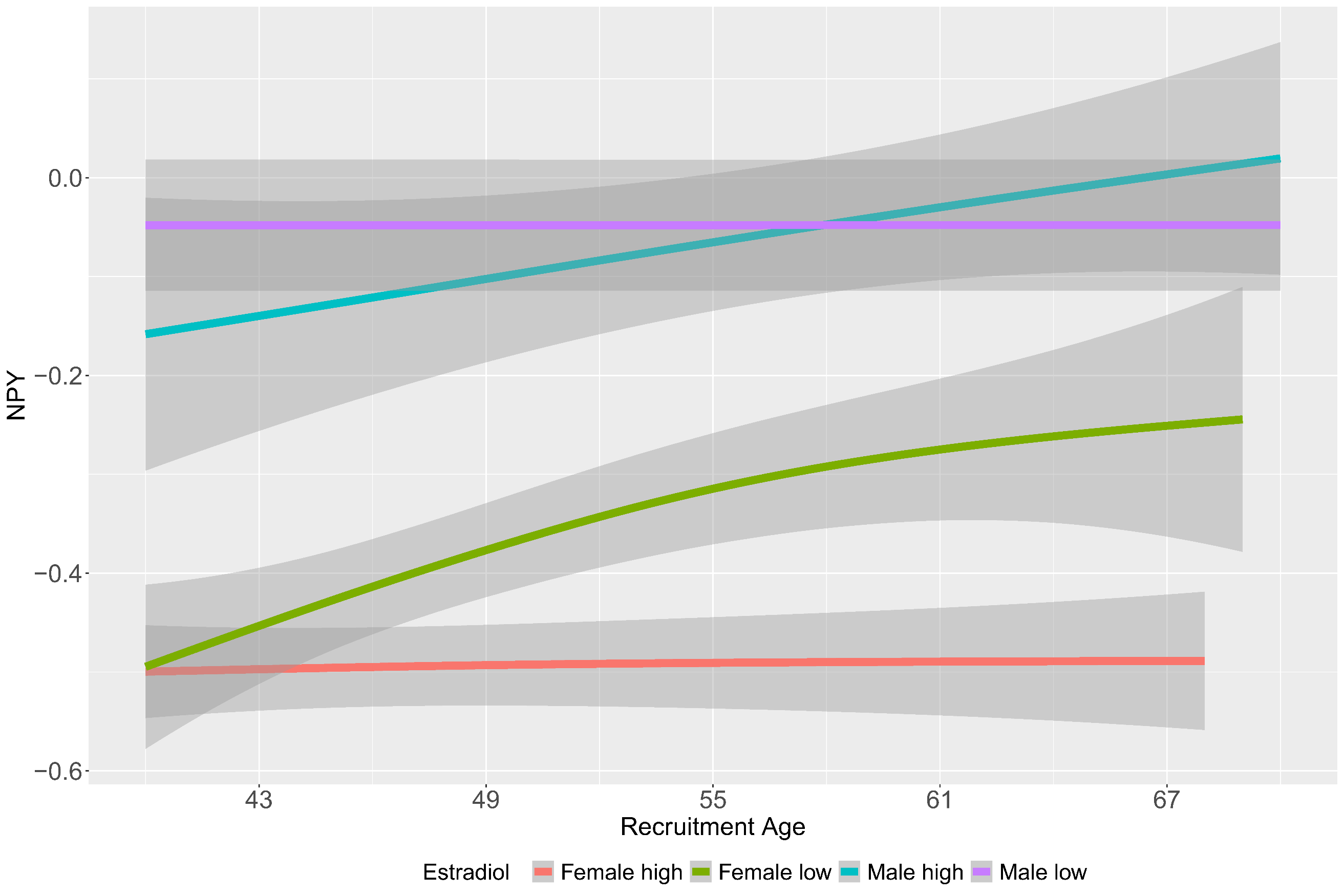

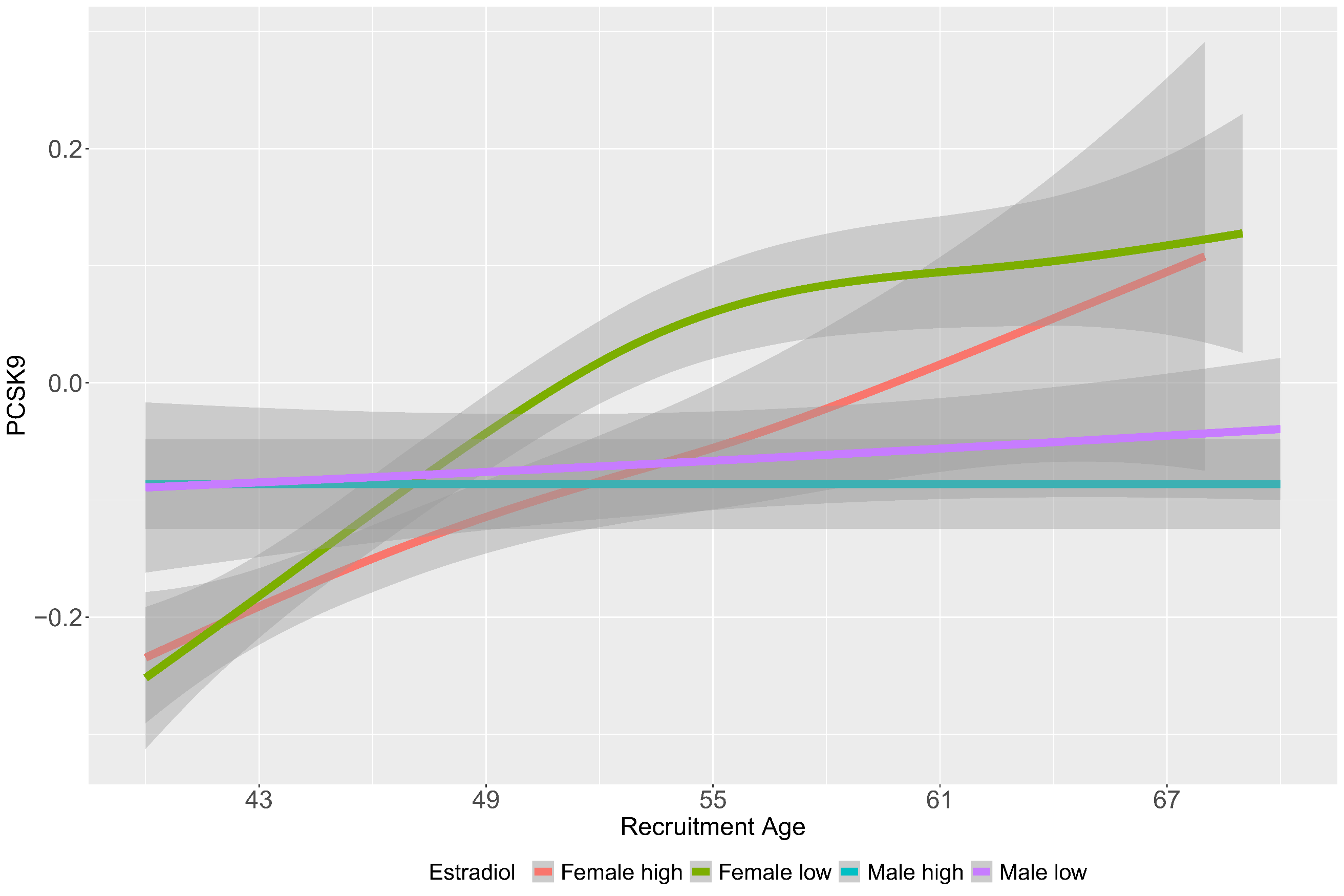

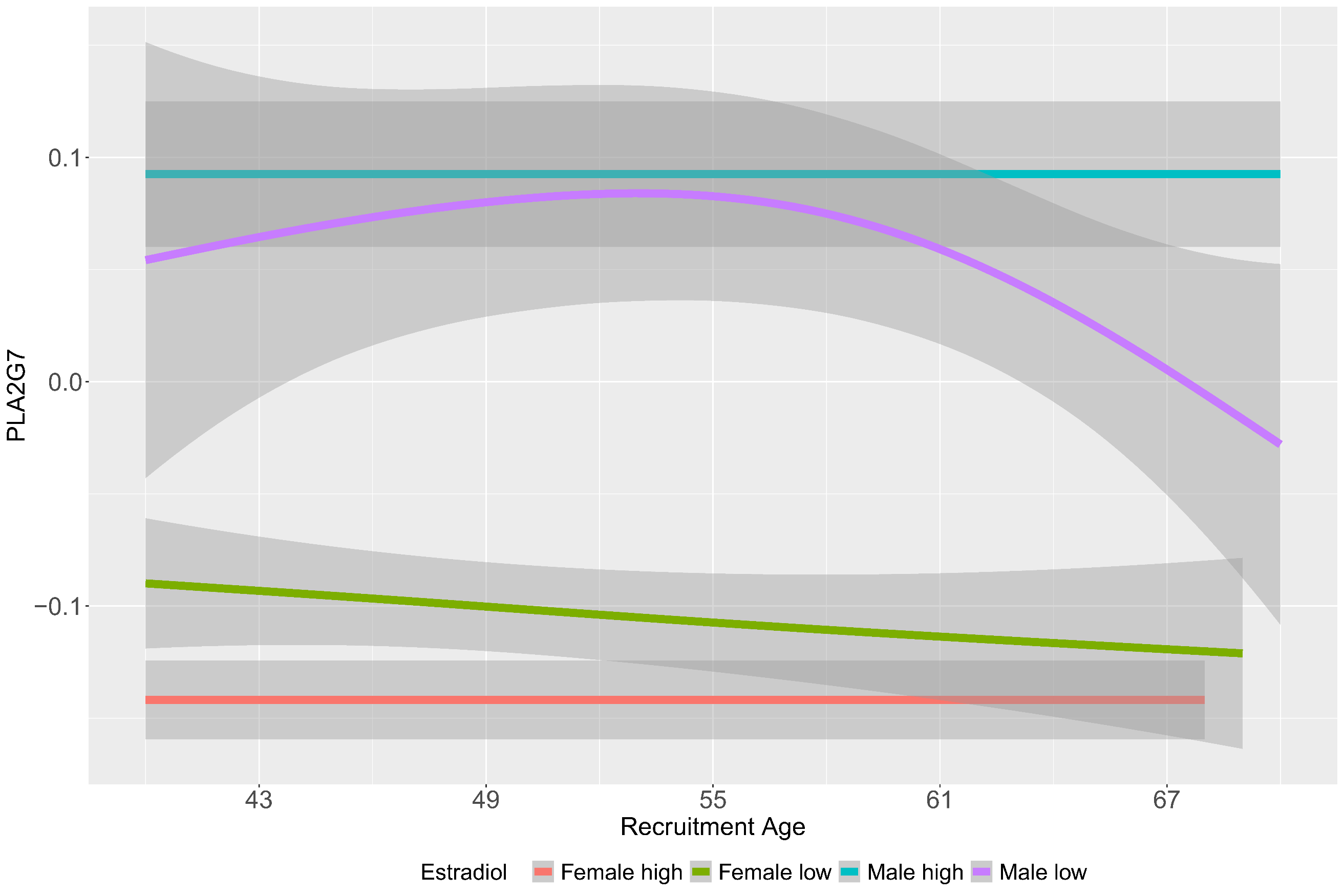

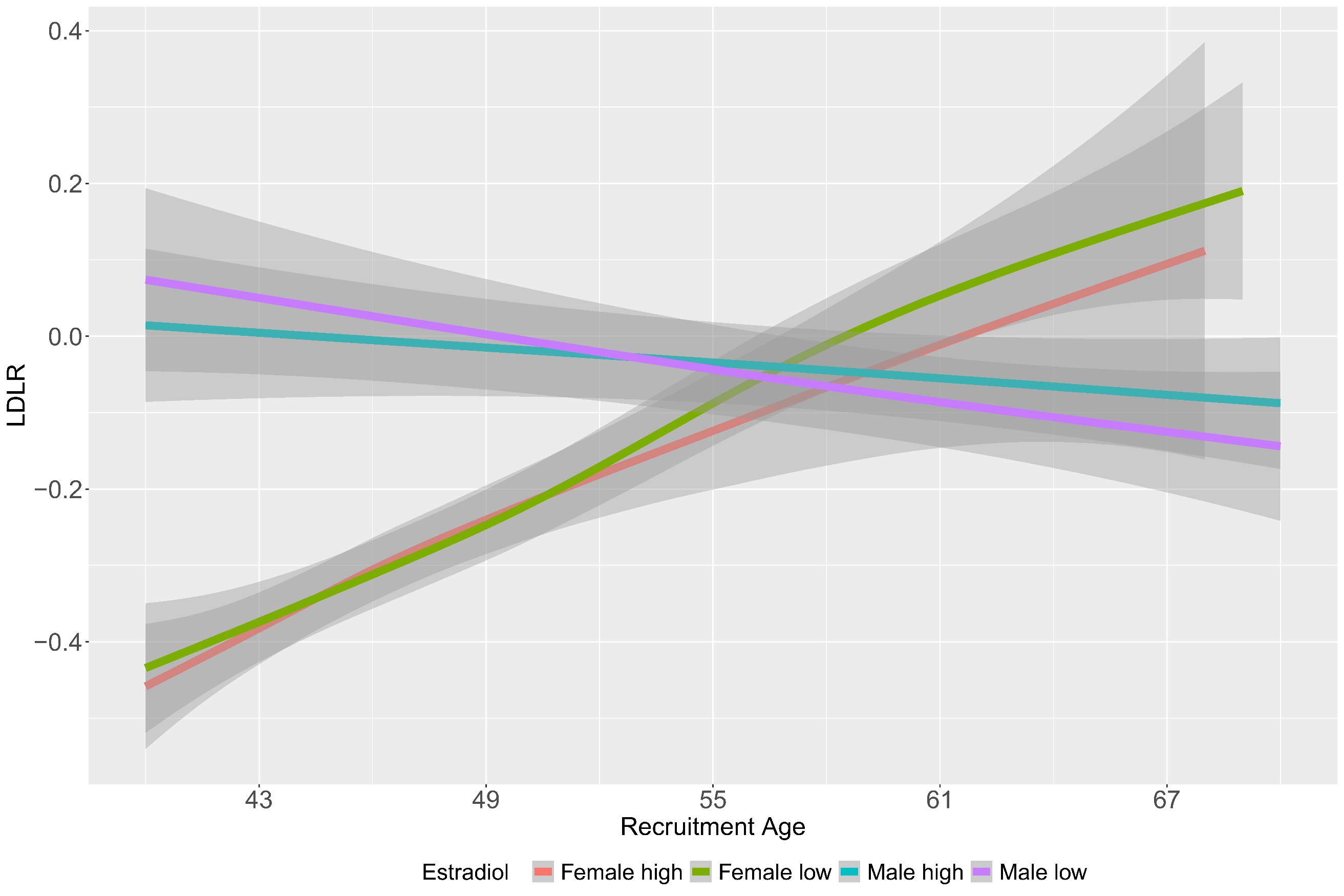

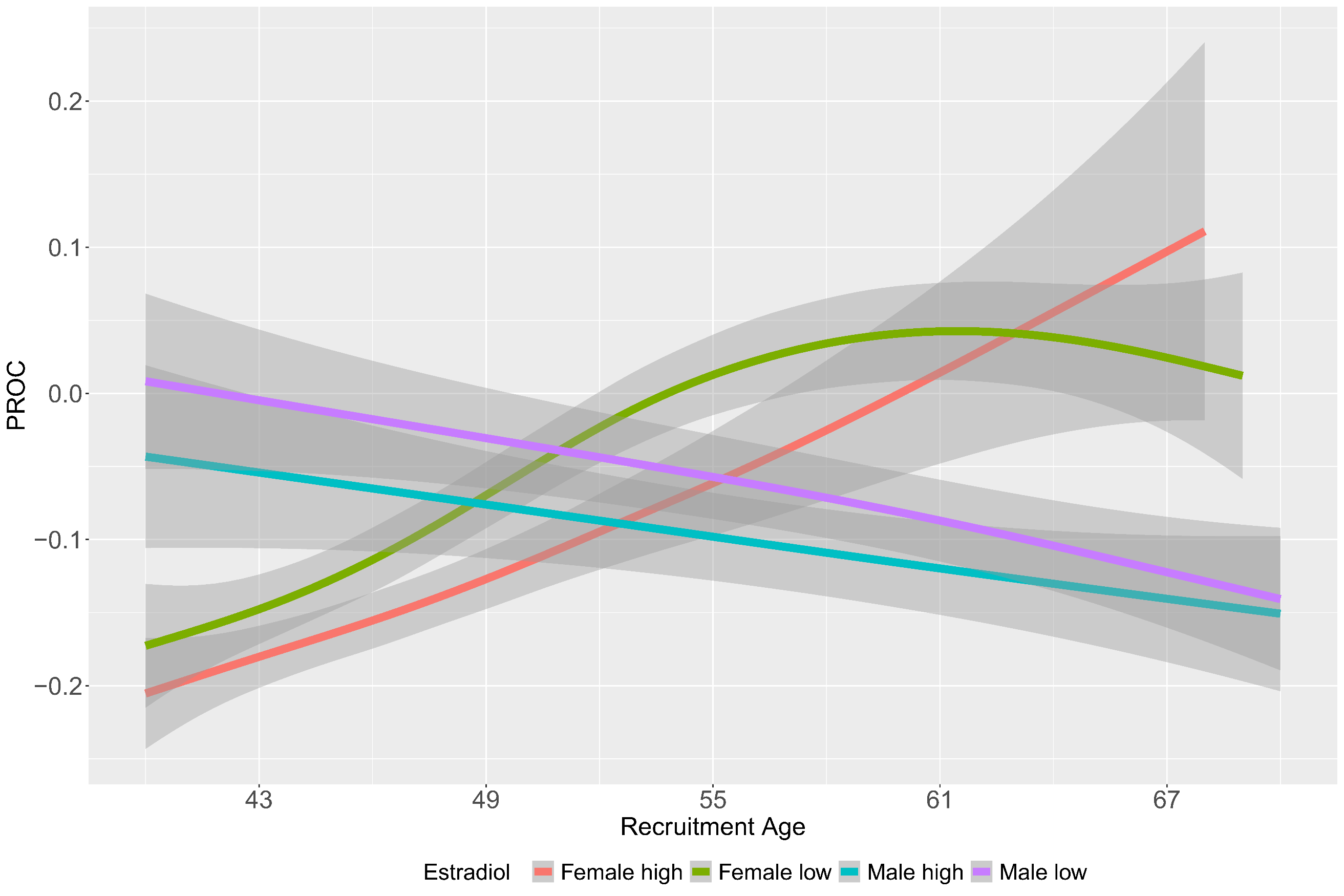

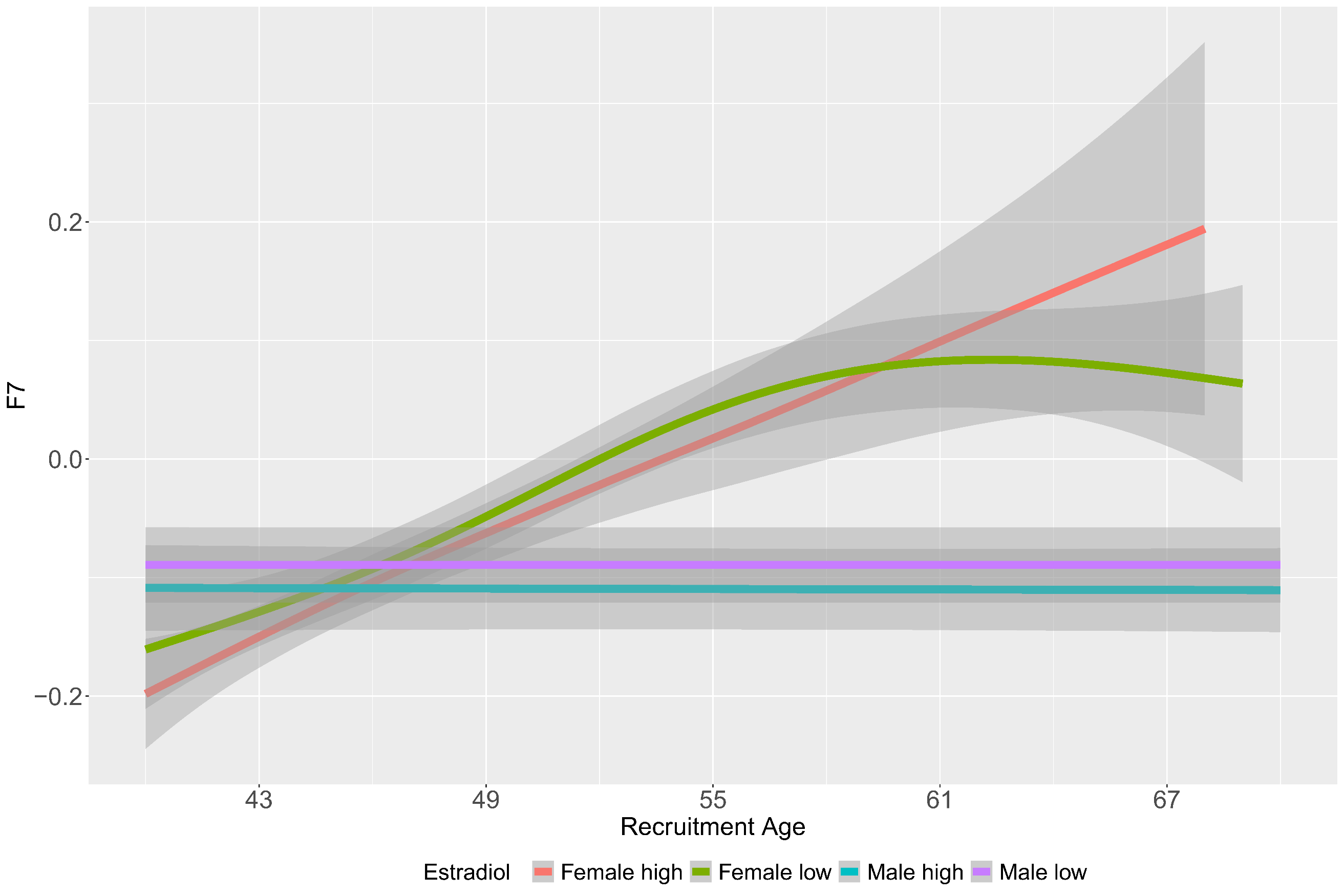

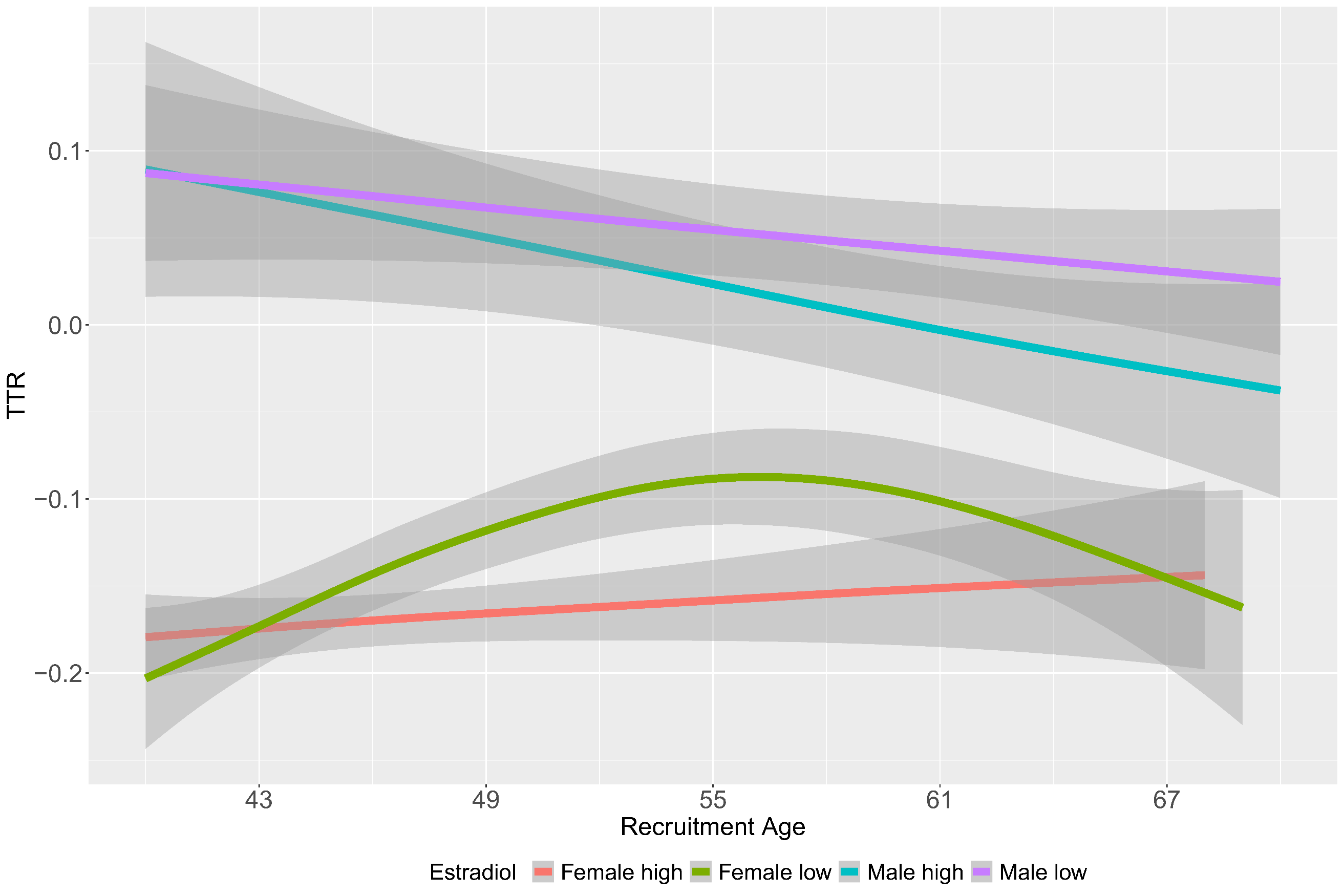

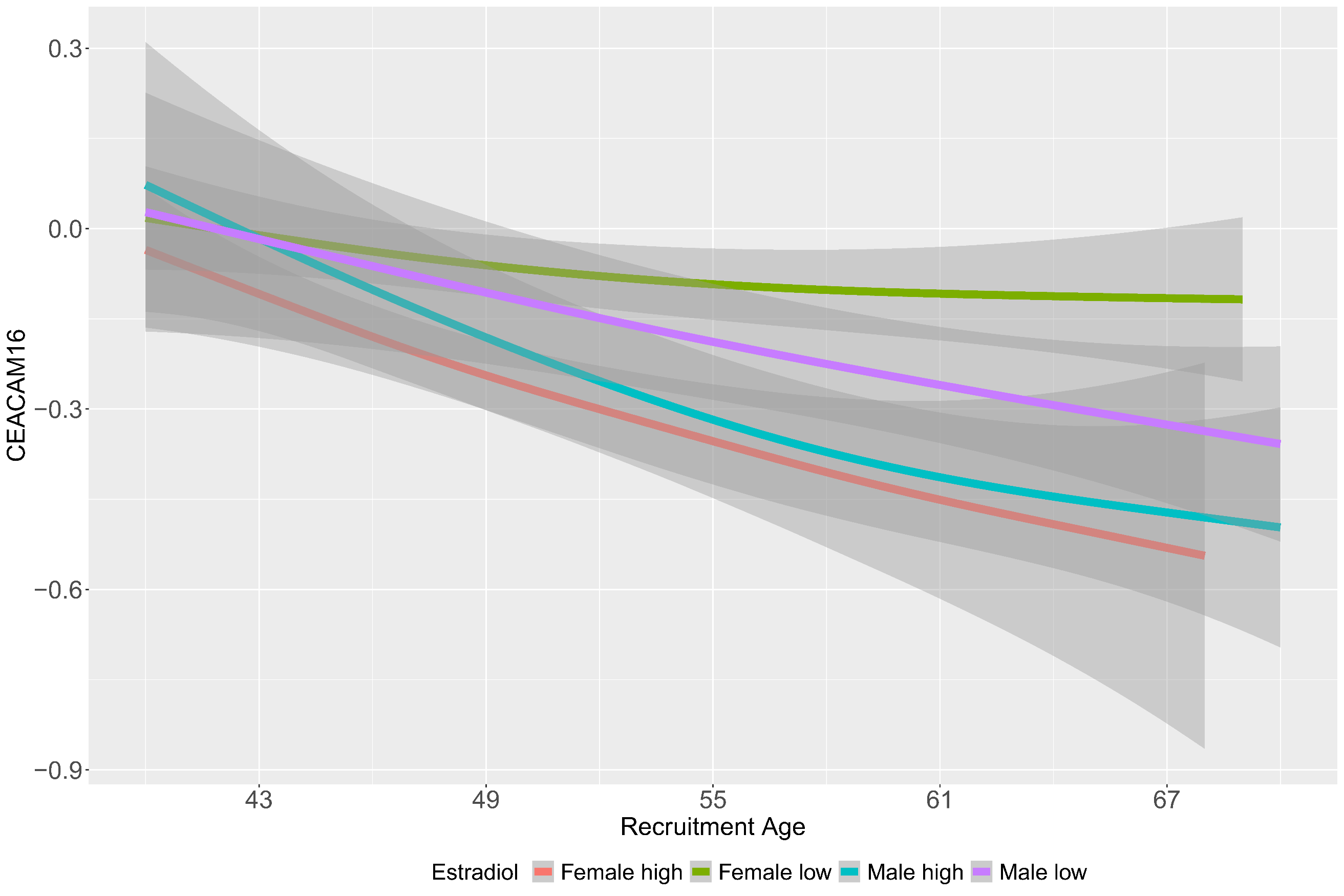

**Supplementary Figure 14**: Age distribution plot of the top aging related sex dimorphic proteins by top and bottom quartiles of estradiol.

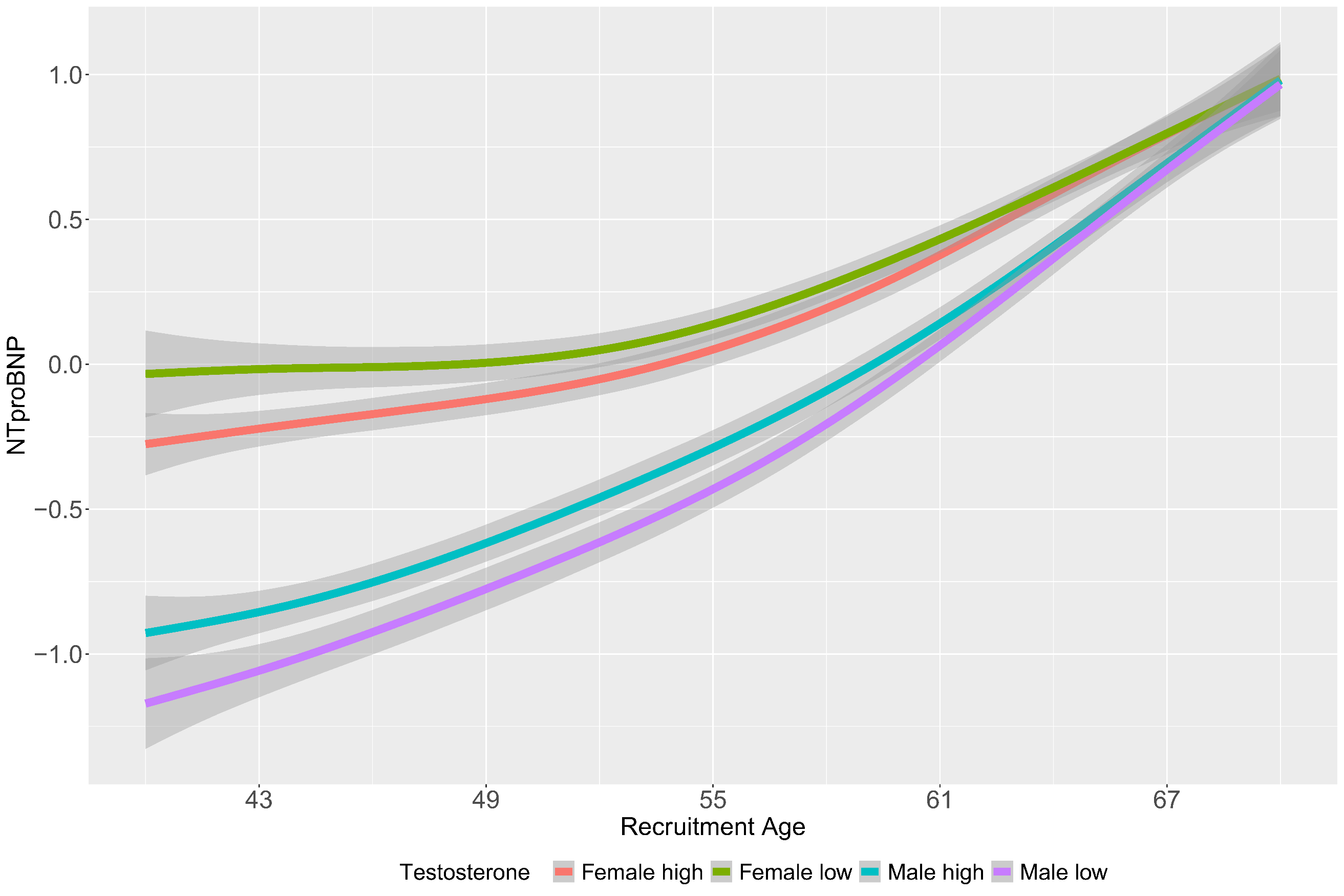

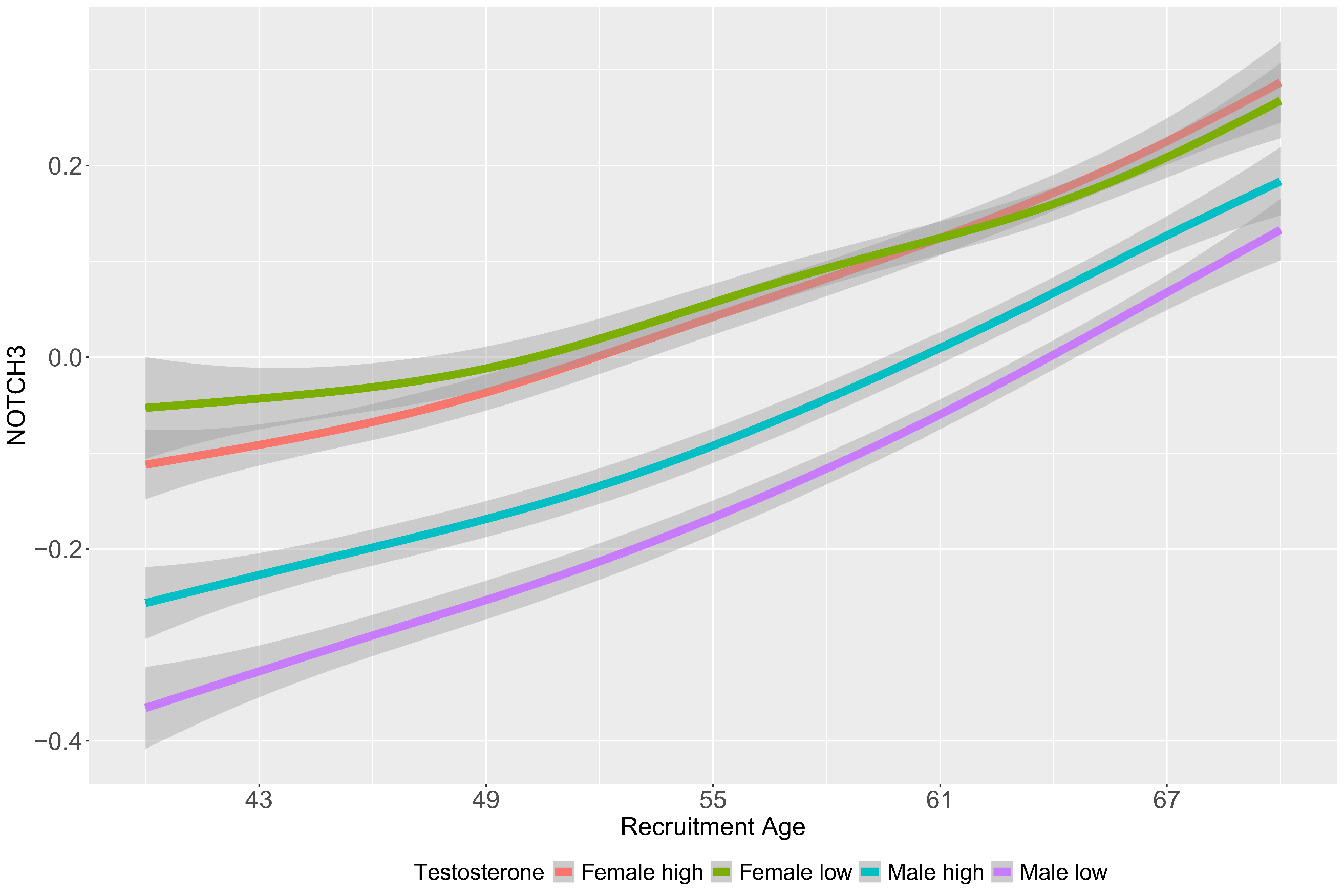

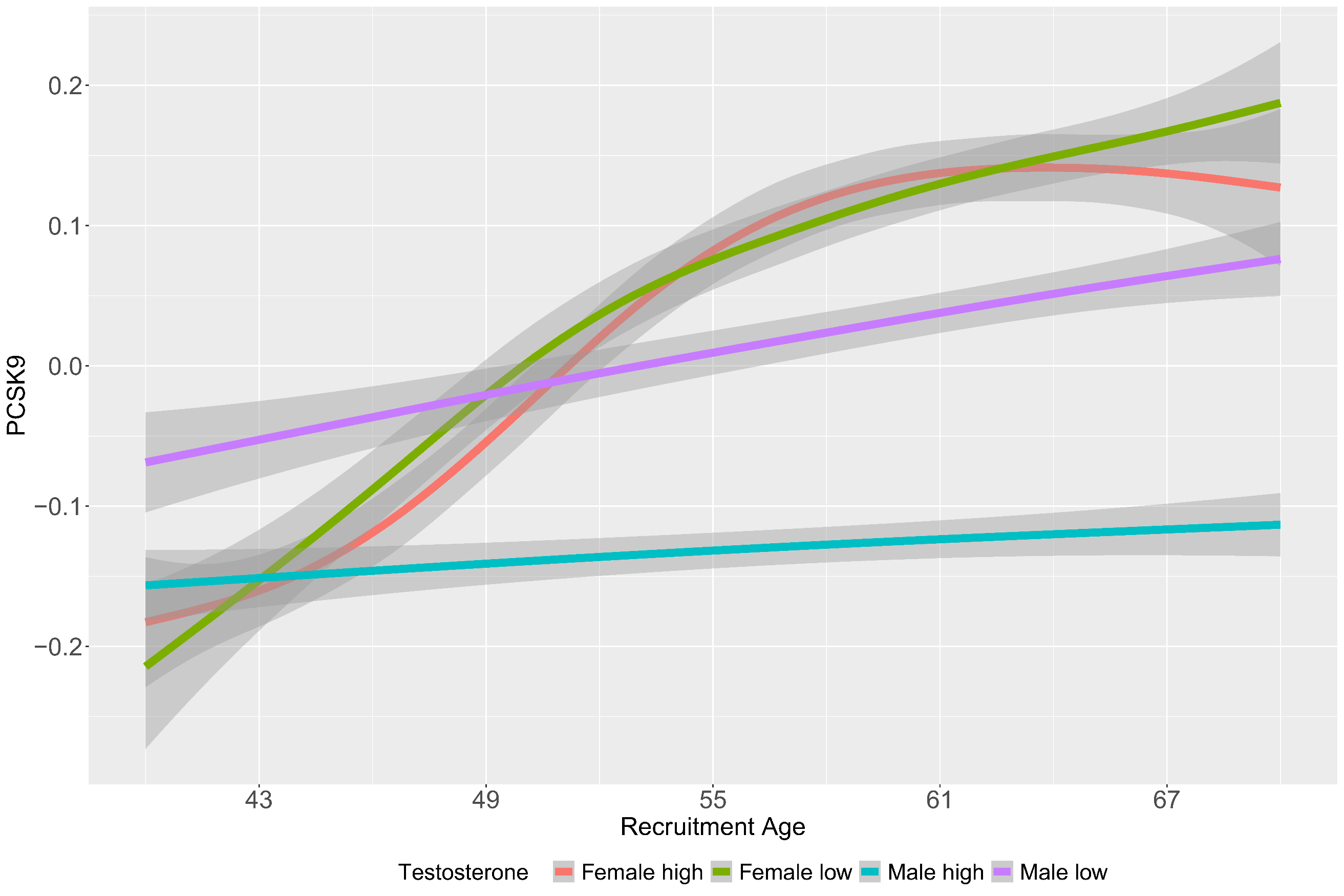

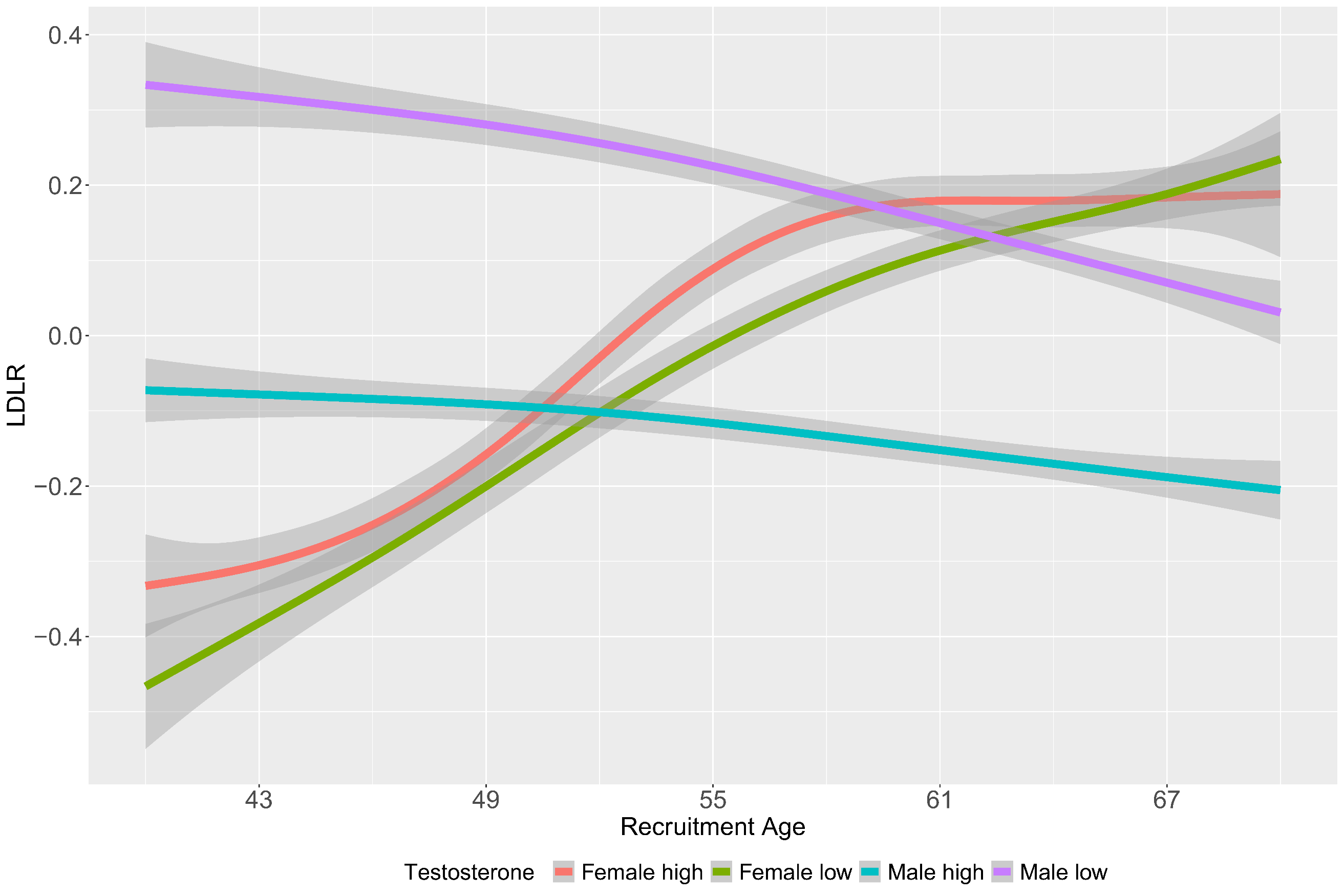

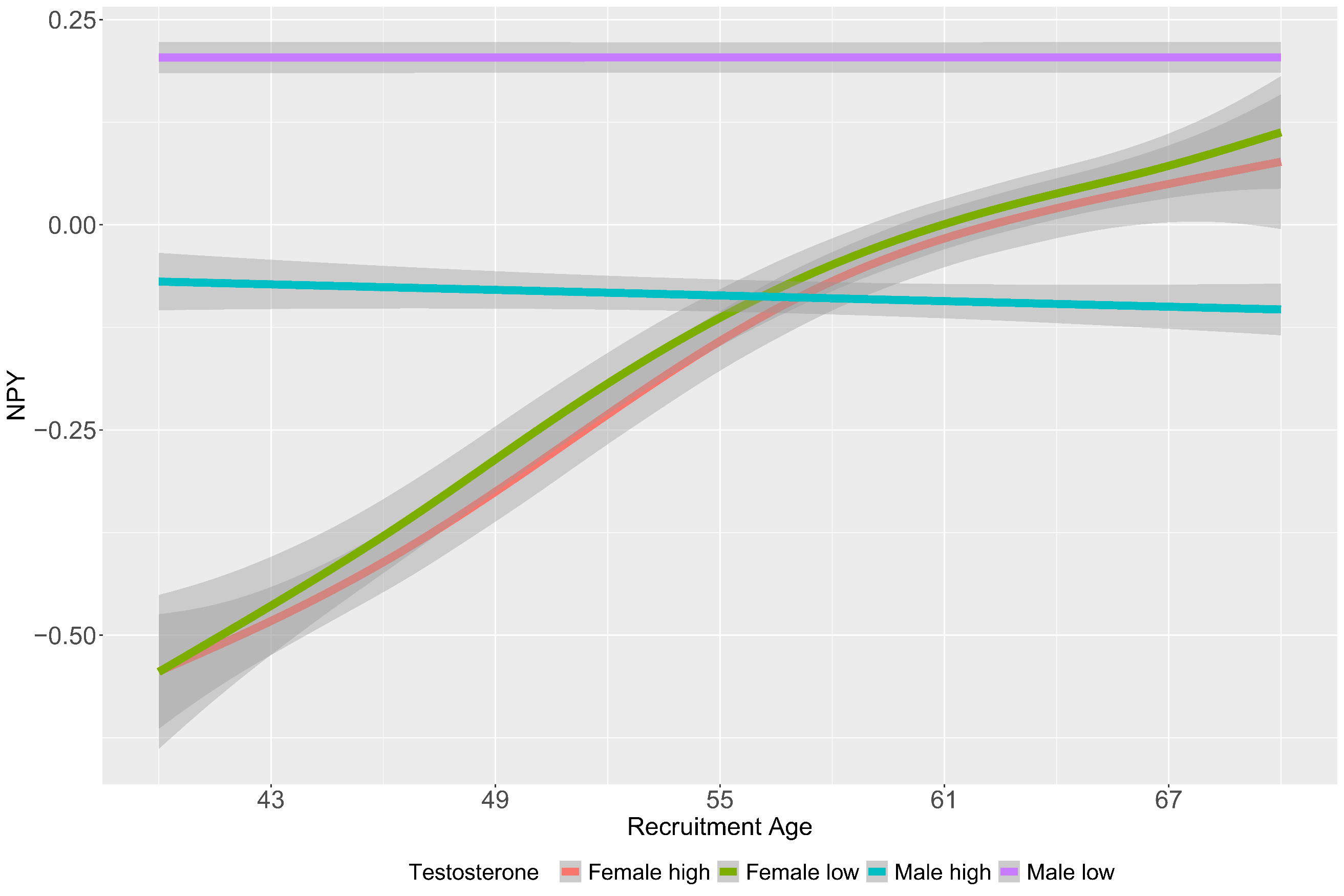

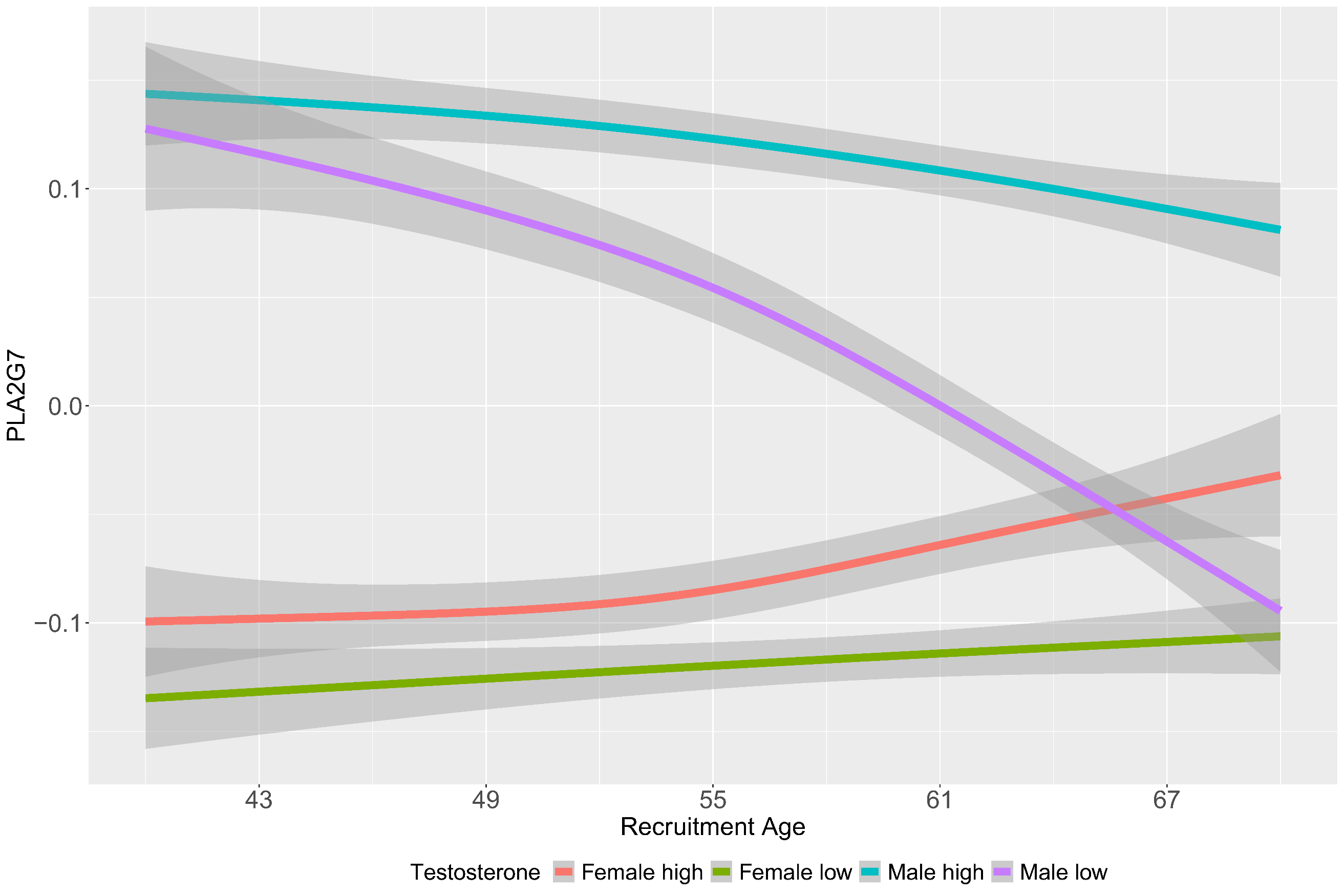

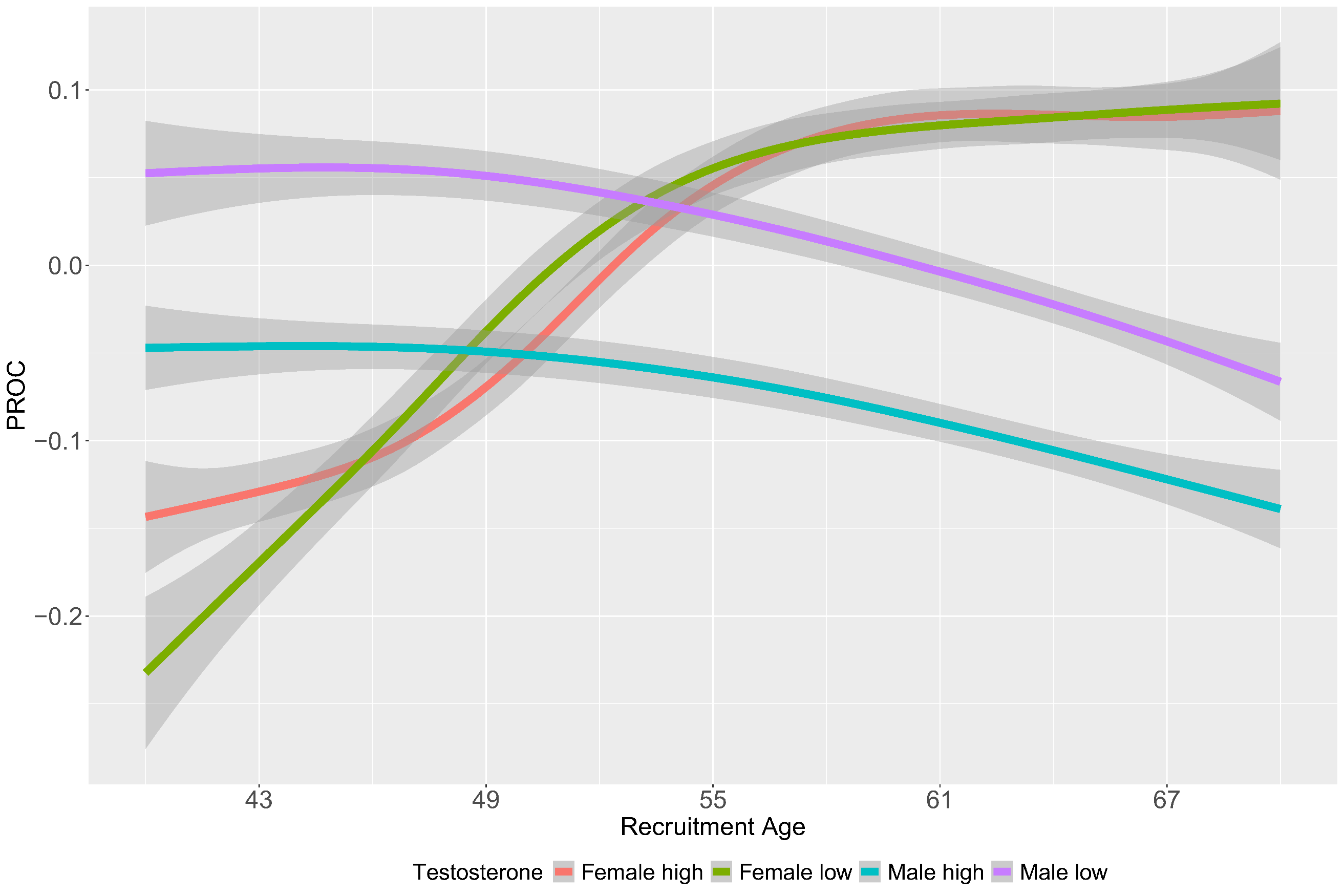

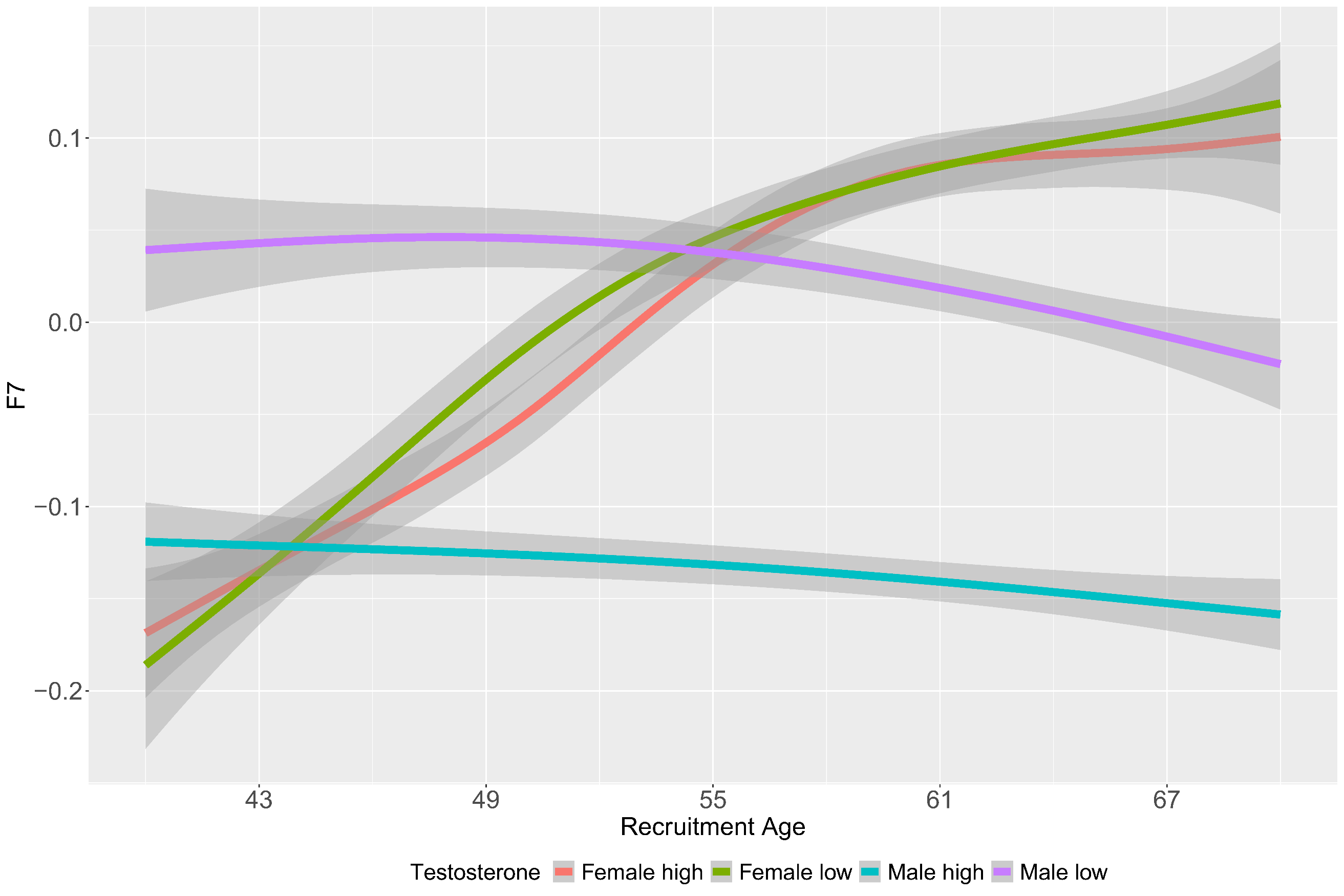

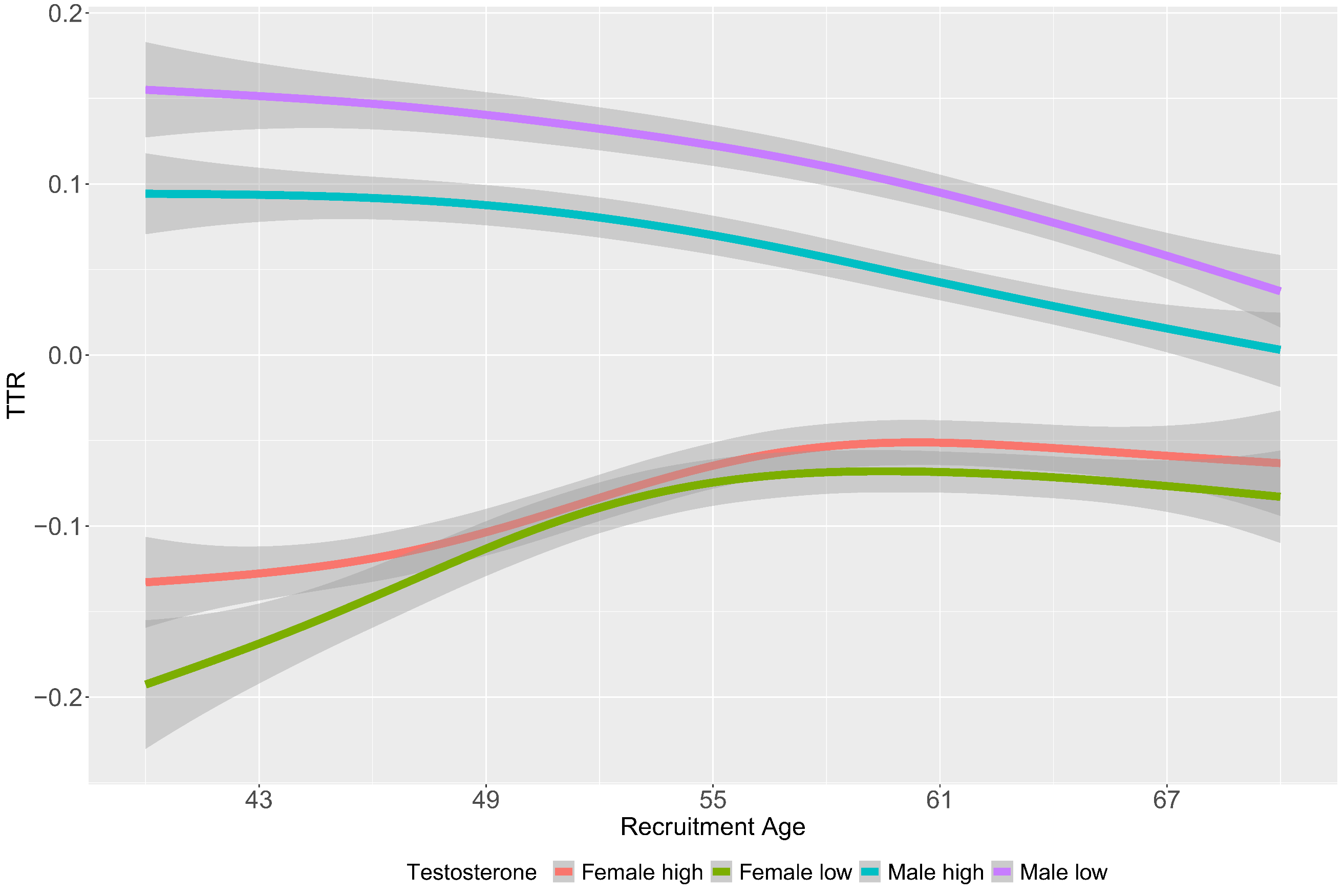

**Supplementary Figure 15**: Age distribution plot of the top aging related sex dimorphic proteins by top and bottom quartiles of testosterone.

**Supplementary Figure 16**: Association of all *cis* & *trans* pqtls of top sex dimorphic proteins with metAgeGap in males (Y-axis) and females (X-axis). Axis depict the Z scores from the association analysis. Each dot depicts a pqtl, where black ones are pqtls genome-wide (GW) significantly associated (p-value < 5.5*10^-08^) with metAgeGap in both males and females, red GW significantly associated with metAgeGap only in females and blue pqtls GW significantly associated with metAgeGap in males only.

**Supplementary Figure 17**: **(A)** Cumulative incidence plot of top, and bottom 10% of the metAgeGap in diseases significant only in males (Cox model 3). X-axis denotes the chronological age and Y-axis denotes cumulative risk. Cumulative incidence and number at risk at each age point is shown in Supplementary Tables 10-13. **(B)** Cumulative incidence plot of top, and bottom 10% of the metAgeGap in vascular dementia.

**Supplementary Figure 18**: Association of metAgeGap to risk of common diseases in males. Model 1 adjusted for chronological age; model 2 was adjusted for recruitment centre, Townsend deprivation index, and ethnicity; model 3 was further adjusted for physical activity, BMI, smoking status, and alcohol frequency.

**Supplementary Figure 19**: Association of metAgeGap to risk of common diseases in females. Model 1 adjusted for chronological age; model 2 was adjusted for recruitment centre, Townsend deprivation index, and ethnicity; model 3 was further adjusted for physical activity, BMI, smoking status, and alcohol frequency.

**Supplementary Figure 20**: Association of metAgeGap to risk of cancers in males. Model 1 adjusted for chronological age; model 2 was adjusted for recruitment centre, Townsend deprivation index, and ethnicity; model 3 was further adjusted for physical activity, BMI, smoking status, and alcohol frequency.

**Supplementary Figure 21**: Association of metAgeGap to risk of cancers in females. Model 1 adjusted for chronological age; model 2 was adjusted for recruitment centre, Townsend deprivation index, and ethnicity; model 3 was further adjusted for physical activity, BMI, smoking status, and alcohol frequency.
